# Sotrovimab versus usual care in patients admitted to hospital with COVID-19: a randomised, controlled, open-label, platform trial (RECOVERY)

**DOI:** 10.1101/2025.01.24.25321081

**Authors:** Peter W Horby, Jonathan R Emberson, Leon Peto, Natalie Staplin, Mark Campbell, Guilherme Pessoa-Amorim, Richard Stewart, Dipansu Ghosh, Graham Cooke, Natalie Blencowe, Jeronimo Moreno-Cuesta, Purav Desai, Paul Hine, Jonathan Underwood, Nicholas Easom, Jaydip Majumdar, Sanjay Bhagani, J Kenneth Baillie, Maya H Buch, Saul N Faust, Thomas Jaki, Katie Jeffery, Edmund Juszczak, Marian Knight, Wei Shen Lim, Alan Montgomery, Aparna Mukherjee, Andrew Mumford, Kathryn Rowan, Guy Thwaites, Marion Mafham, Richard Haynes, Martin J Landray, Group RECOVERY Collaborative

## Abstract

**Background:** Sotrovimab is a neutralising monoclonal antibody that has been proposed as a treatment for patients admitted to hospital with COVID-19.

**Methods:** In this randomised, controlled, open-label platform trial, several possible treatments were compared with usual care in patients hospitalised with COVID-19 pneumonia. In the sotrovimab comparison, eligible and consenting patients were randomly allocated to either usual care alone or usual care plus a single 1g dose of sotrovimab, using web-based unstratified randomisation. Participants were retrospectively categorised according to their baseline serum SARS-CoV-2 nucleocapsid antigen concentration as ‘high-antigen’ or ‘low-antigen’, using the median concentration as a cut-off. The primary outcome was 28-day mortality assessed by intention to treat. Secondary outcomes were time to discharge alive from hospital, and, among those not on invasive ventilation at baseline, progression to invasive ventilation or death. Recruitment closed on 31 March 2024 when funding ended. ISRCTN (50189673) and clinicaltrials.gov (NCT04381936).

**Findings:** From 4 January 2022 to 19 March 2024, 1723 patients were recruited to the sotrovimab comparison. 720 (42%) were classified as high-antigen, 717 (42%) as low-antigen, and 286 (17%) had unknown antigen status. Over 80% of patients were vaccinated, over 80% had anti-spike antibodies at randomisation, and almost all were infected with Omicron variants. In the prespecified primary efficacy population of high-antigen patients, 82/355 (23%) allocated sotrovimab versus 106/365 (29%) allocated usual care died within 28 days (rate ratio 0.75; 95% CI 0.56-0.99; p=0.046). In an analysis of all randomised patients (regardless of antigen status), 177/828 (21%) allocated sotrovimab versus 201/895 (22%) allocated usual care died within 28 days (rate ratio 0.95; 95% CI 0.77-1.16; p=0.60).

**Interpretation:** In patients hospitalised with COVID-19, sotrovimab was associated with reduced mortality in the primary analysis population of patients with a high serum SARS-CoV-2 antigen concentration at baseline, but not in the overall population.

**Funding:** UK Research and Innovation (Medical Research Council) and National Institute of Health Research (Grant ref: MC_PC_19056).

## INTRODUCTION

Treatment with neutralising monoclonal antibodies (mAbs) targeting the SARS-CoV-2 spike protein has been found to substantially reduce the risk of hospitalisation or death in patients with early COVID-19 who are at high risk of complications.^1–3^ nMAbs were also found to reduce the risk of death among hospitalised patients, but this benefit was restricted to those who had not yet developed their own anti-SARS-CoV-2 antibody response (i.e. who are seronegative).^4–6^ The RECOVERY casirivimab-imdevimab comparison, which recruited UK patients from September 2020 to May 2021 is the largest randomised evaluation of mAb therapy in hospitalised patients. In this comparison, 28-day mortality in patients who were seronegative at randomisation was double that of seropositive patients (30% versus 15%), and mAb therapy reduced this to 24% (rate ratio 0.79; 95% confidence interval 0.69-0.91; p=0.0009; number of seronegative patients treated to save one life = 16).^4^ Following this, targeted mAb therapy for seronegative patients hospitalised with COVID-19 was adopted into routine practice in the UK and elsewhere.

A major limitation of mAb therapy has been the frequent emergence of new SARS-CoV-2 variants that are not effectively neutralised by existing antibodies.^7,8^ When the first Omicron variant, BA.1, became globally dominant in December 2021, it contained spike mutations conferring high-level resistance to most mAbs in clinical use.^8^ This included the casirivimab-imdevimab combination, leading to its withdrawal from guidelines. Sotrovimab, a mAb originally developed from an antibody recovered from a patient who had recovered from SARS-CoV-1, targets a relatively conserved spike protein epitope, and in the COMET-ICE trial of patients with early infection conducted in 2020-21 it reduced the risk of hospitalisation or death by 79%.^1^ The neutralisation potency of sotrovimab was modestly reduced against BA.1 (∼3-5-fold) compared to wild-type virus, but it retained more activity than many other mAbs, which made it a promising candidate for continued use in hospitalised patients and prompted its evaluation in RECOVERY.^9,10^ A further reduction in activity against BA.2 led to the withdrawal of FDA Emergency Use Authorization in the U.S. for sotrovimab in April 2022. However, it retained enough *in vitro* activity against viral variants prevalent in 2022-23 to suggest it could retain clinical benefit, either via direct neutralisation or via Fc-dependent effector mechanisms.^11,12^ During November 2023, BA.2.86 and JN.1 SARS-CoV-2 variants became dominant in the UK and elsewhere, which have an additional spike gene mutation that confers high-level resistance to sotrovimab.^13^

The current role of therapeutic neutralising mAbs in hospitalised patients is also complicated by increasing population immunity to SARS-CoV-2, as the previous trials that identified a benefit in seronegative patients were performed before widespread vaccination and natural immunity. By the time the Omicron BA.1 variant emerged, most people hospitalised in the UK with COVID-19 had been vaccinated and many had had previous infection. In this setting, patients would be expected to have detectable anti-SARS-CoV-2 antibodies at admission, but this could reflect immune responses to previous vaccination or infection that had failed to prevent the current illness, rather than adaptive immunity to the current infection. This suggests that alternative biomarkers of infection status may now be required to identify which, if any, hospitalised patients could benefit from mAb treatment.

One possible biomarker is SARS-CoV-2 antigenaemia. Viral nucleocapsid antigen is detectable in the blood of most hospitalised patients, and high concentrations are strongly correlated with more severe disease and worse prognosis.^14–16^ In most hospitalised patients, antigen levels fall rapidly in the first few days of admission as the infection is cleared.^17^ The degree of antigenaemia is inversely correlated with specific antibody responses, but, unlike antibodies, detection of viral antigen almost certainly relates only to the current infection.

Here we report the results of the sotrovimab comparison in RECOVERY, a randomised, open-label platform trial evaluating treatments for patients hospitalised with COVID-19 pneumonia. Recruitment occurred in the UK in a period in which Omicron variants were dominant and most people were vaccinated against SARS-CoV-2. The prespecified primary analysis population was patients who had serum SARS-CoV-2 nucleocapsid antigen concentration at randomisation that was above the median value of all trial participants in this comparison.

## METHODS

### Study design and participants

The Randomised Evaluation of COVID-19 therapy (RECOVERY) trial is an investigator-initiated, individually randomised, controlled, open-label, adaptive platform trial to evaluate the effects of potential treatments in patients hospitalised with COVID-19. Details of the trial design and results for other treatments have been published previously and are available at www.recoverytrial.net/results.^4,18–28^ The trial was conducted at hospital organisations in the United Kingdom and supported by the National Institute for Health and Care Research Clinical Research Network. 107 hospitals in the UK enrolled participants in the sotrovimab comparison (appendix pp5-32). The trial is coordinated by the Nuffield Department of Population Health at the University of Oxford (Oxford, UK), the trial sponsor, and is conducted in accordance with the principles of the International Conference on Harmonisation–Good Clinical Practice guidelines and is approved by the UK Medicines and Healthcare products Regulatory Agency (MHRA) and the Cambridge East Research Ethics Committee (ref: 20/EE/0101). The protocol, statistical analysis plan, and additional information are available in the appendix (pp70-186) and on the study website www.recoverytrial.net.

Patients admitted to hospital were eligible for the study if they had confirmed SARS-CoV-2 infection with a pneumonia syndrome thought to be related to COVID-19, and no medical history that might, in the opinion of the managing physician, put the patient at significant risk if they were to participate in the trial. Patients were excluded if they were aged <12 years or were aged <18 years and weighed <40kg. Pregnant women were eligible. Written informed consent was obtained from all patients, or a legal representative if patients were too unwell or otherwise unable to provide informed consent.

### Randomisation and masking

Eligible and consenting patients were randomly assigned in a 1:1 ratio to either usual standard of care plus sotrovimab or usual standard of care alone, using web-based simple (unstratified) randomisation with allocation concealed until after randomisation (appendix pp42-44). Patients allocated to sotrovimab were to receive 1g in 100ml 0.9% saline or 5% glucose intravenously over 60 minutes as soon as possible after randomisation. This is double the licensed dose for early infection and was selected because of reduced neutralisation activity against Omicron BA.1 compared to wild-type virus.

As a platform trial, and in a factorial design, patients could be simultaneously included in other concurrently evaluated treatment comparisons, each having its allocation determined by an independent 1:1 randomisation: (i) empagliflozin versus usual care, (ii) higher-dose corticosteroids versus usual care, (iii) molnupiravir versus usual care, and (iv) nirmatrelvir-ritonavir versus usual care (appendix pp42-43). Participants and local study staff were not masked to allocated treatment. Other than members of the Data Monitoring Committee, all individuals involved in the trial were masked to aggregated outcome data while recruitment and 28-day follow-up were ongoing.

### Procedures

Baseline data were collected using a web-based case report form that included demographics, level of respiratory support, major comorbidities, suitability of the study treatment for a particular patient, SARS-CoV-2 vaccination status, and study treatment availability at the study site (appendix pp47). A serum sample and nose swab were collected at randomisation and sent to central laboratories for testing. Serum was tested for SARS-CoV-2 nucleocapsid antigen, anti-SARS-CoV-2 spike antibodies, and anti-SARS-CoV-2 nucleocapsid antibodies using Roche Elecsys assays (Roche Diagnostics, Basel, Switzerland). Patients were classified as having high- or low-serum nucleocapsid antigen using the trial population median value, and as positive or negative for anti-spike and anti-nucleocapsid antibodies using manufacturer defined thresholds (testing was retrospective, so results were not available to the patient’s medical team). Nose swabs were tested for SARS-CoV-2 RNA using TaqPath COVID-19 RT-PCR (Thermo Fisher Scientific, Massachusetts, US). Samples with sufficient viral RNA were sequenced using the ONT Midnight protocol (Oxford Nanopore Technologies, Oxford, UK).^29^ Sequence data were used to detect spike protein mutations associated with >5-fold reduction in sotrovimab neutralisation, which were identified from the sotrovimab summary of product characteristics and the Stanford University Coronavirus Antiviral and Resistance Database.^30^ Further details of laboratory analyses and the resistance mutations included are in the appendix (pp33-34, 188-203).

Follow-up nose swabs were collected on day 3 and day 5 (counting the day of randomisation as day 1). These were analysed in the same manner as the baseline swab described above.

An online follow-up form was completed when participants were discharged, had died or at 28 days after randomisation, whichever occurred earliest (appendix pp48-56). Information was recorded on adherence to allocated study treatment, receipt of other COVID-19 treatments, duration of admission, receipt of respiratory or renal support, major safety outcomes, and vital status (including cause of death). In addition, routine healthcare and registry data were obtained, including information on vital status (with date and cause of death), discharge from hospital, receipt of respiratory support, or renal replacement therapy.

### Outcomes

Outcomes were assessed at 28 days after randomisation, with further analyses specified at 6 months (not reported here). The primary outcome was all-cause mortality at 28 days. Secondary outcomes were time to discharge from hospital, and, among patients not on invasive mechanical ventilation at randomisation, invasive mechanical ventilation (including extra-corporal membrane oxygenation) or death. Prespecified subsidiary clinical outcomes were use of invasive or non-invasive ventilation (including high-flow nasal oxygen) among patients not on any ventilation at randomisation, and use of renal dialysis or haemofiltration. Prespecified safety outcomes were cause-specific mortality, major cardiac arrhythmia, thrombotic and major bleeding events, non-SARS-CoV-2 infections, hyper/hypoglycaemia, seizures, acute liver or kidney injury, and infusion reactions to sotrovimab. Virological outcomes were viral RNA copy number in nose swabs taken at day 3 and day 5, and the frequency of detection of resistance mutations. Information on suspected serious adverse reactions was collected in an expedited fashion to comply with regulatory requirements. Details of the methods used to ascertain and derive outcomes are provided in the appendix (pp164).

### Statistical Analysis

For all outcomes, intention-to-treat analyses compared patients randomly allocated sotrovimab with patients randomly allocated usual care. For the primary outcome of 28-day mortality, the hazard ratio from a Cox model with adjustment for age in three categories (<70 years, 70-79 years, and 80 years or older) and ventilation status at randomisation in four categories (no oxygen, simple oxygen only, non-invasive ventilation and invasive mechanical ventilation) was used to estimate the mortality rate ratio. We constructed Kaplan-Meier survival curves to display cumulative mortality over the 28-day period (starting on the day of randomisation and ending 28 days later). We used the same Cox regression method to analyse time to hospital discharge and successful cessation of invasive mechanical ventilation, with patients who died in hospital right-censored on day 29.

Median time to discharge was derived from Kaplan-Meier estimates. For the composite secondary outcome of progression to invasive mechanical ventilation or death within 28 days, and the subsidiary clinical outcomes of receipt of ventilation and use of haemodialysis or haemofiltration, the precise dates were not available and a log-binomial regression model was used to estimate the risk ratio adjusted for age and ventilation status (in the same categories as listed above). Estimates of rate and risk ratios are shown with 95% confidence intervals. SARS-CoV-2 viral RNA levels in nose-swabs were estimated with analysis of covariance (ANCOVA) using the log transformed values after adjustment for each participant’s baseline value, age and level of respiratory support at randomisation. Missing baseline and follow-up values of SARS-CoV-2 viral RNA levels were estimated using multiple imputation, with 20 replicate sets and combination of results across sets using the methods of Rubin.^31^

When the sotrovimab comparison was added to the protocol in December 2021, there was insufficient information to decide if anti-S or anti-N antibody status should define the primary analysis population, or if serum antigen status would be preferable. The statistical analysis plan stated that this would be determined at a future date (but prior to unblinding of the investigator team). Shortly after recruitment closed, but before the investigators were unblinded, high-antigen patients were selected as the primary analysis population because of low numbers of seronegative patients in the trial population and because antigen positivity best predicted mortality (described in the updated statistical analysis plan, appendix pp154-156). It was hypothesised that any beneficial effect of sotrovimab would be larger among high-antigen patients and may be negligible in low-antigen patients. Formal hypothesis-testing of the effect of allocation to sotrovimab on 28-day mortality was to be done firstly in high-antigen participants (the primary analysis population), and was to be done among all randomised participants only if a reduction in mortality in high-antigen patients was seen at 2p<0.05. Formal testing of secondary outcomes was only to be done if a mortality reduction among all participants was seen at 2p<0.05. A prespecified comparison of the effects of allocation to sotrovimab on 28-day mortality in high-antigen versus low-antigen participants was done by performing a test for heterogeneity. Tests for heterogeneity according to other baseline characteristics were also prespecified (age, sex, ethnicity, level of respiratory support, days since symptom onset, use of corticosteroids, anti-SARS-CoV-2 antibody status, and immunosuppression).

Because trial recruitment and event rates during the COVID-19 pandemic were unpredictable, RECOVERY treatment comparisons have not had a predetermined sample size. With high levels of recruitment, the intention would have been to continue until enough primary outcomes had accrued for a 90% power to detect a proportional risk reduction of 20% at 2p=0.01 (approximately 5,500 participants if mortality were 20% without treatment). Following the initial wave of Omicron infection in the UK in early 2022, the number of patients hospitalised with COVID-19 pneumonia reduced substantially in the UK, as did trial recruitment. The trial comparison closed on 31^st^ March 2024 when funding for the trial ended.

The full database is held by the study team which collected the data from study sites and performed the analyses at the Nuffield Department of Population Health, University of Oxford (Oxford, UK). Analyses were performed using SAS version 9.4 and R version 4.0.3. The trial is registered with ISRCTN (50189673) and clinicaltrials.gov (NCT04381936).

### Role of the funding source

Neither the study funders, nor the manufacturers of sotrovimab, had any role in study design, data collection, data analysis, or writing of the report. GSK and Vir Biotechnology supported the study through supply of sotrovimab and reviewed the draft publication for scientific consistency and completeness. The corresponding authors had full access to all the data in the study and had final responsibility for the decision to submit for publication.

## RESULTS

Between 4 January 2022 and 19 March 2024, 1723/1824 (94%) patients enrolled into the RECOVERY trial at sites participating in the sotrovimab comparison were eligible and agreed to be included in sotrovimab comparison, of whom 1448 (84%) were recruited in 2022. 828 were allocated sotrovimab and 895 were allocated usual care without sotrovimab (figure 1). The mean age of study participants was 70.7 years (SD 14.8), 1389 (81%) had received a COVID-19 vaccine, and 414 (24%) were severely immunocompromised in the opinion of the managing clinician (table 1, appendix pp59-60). At randomisation, the median time since symptom onset was 6 days (IQR 3-11 days), 1467 (85%) were receiving oxygen or ventilatory support, and 628 (36%) were receiving remdesivir. Serological results were available for 1439 (84%) of patients, among whom 720 (50%) had a serum concentration above the median (‘high-antigen’), 1179 (82%) were anti-SARS-CoV-2 spike antibody positive, and 454 (32%) were anti-SARS-CoV-2 nucleocapsid antibody positive (table 1).

**Figure 1:**
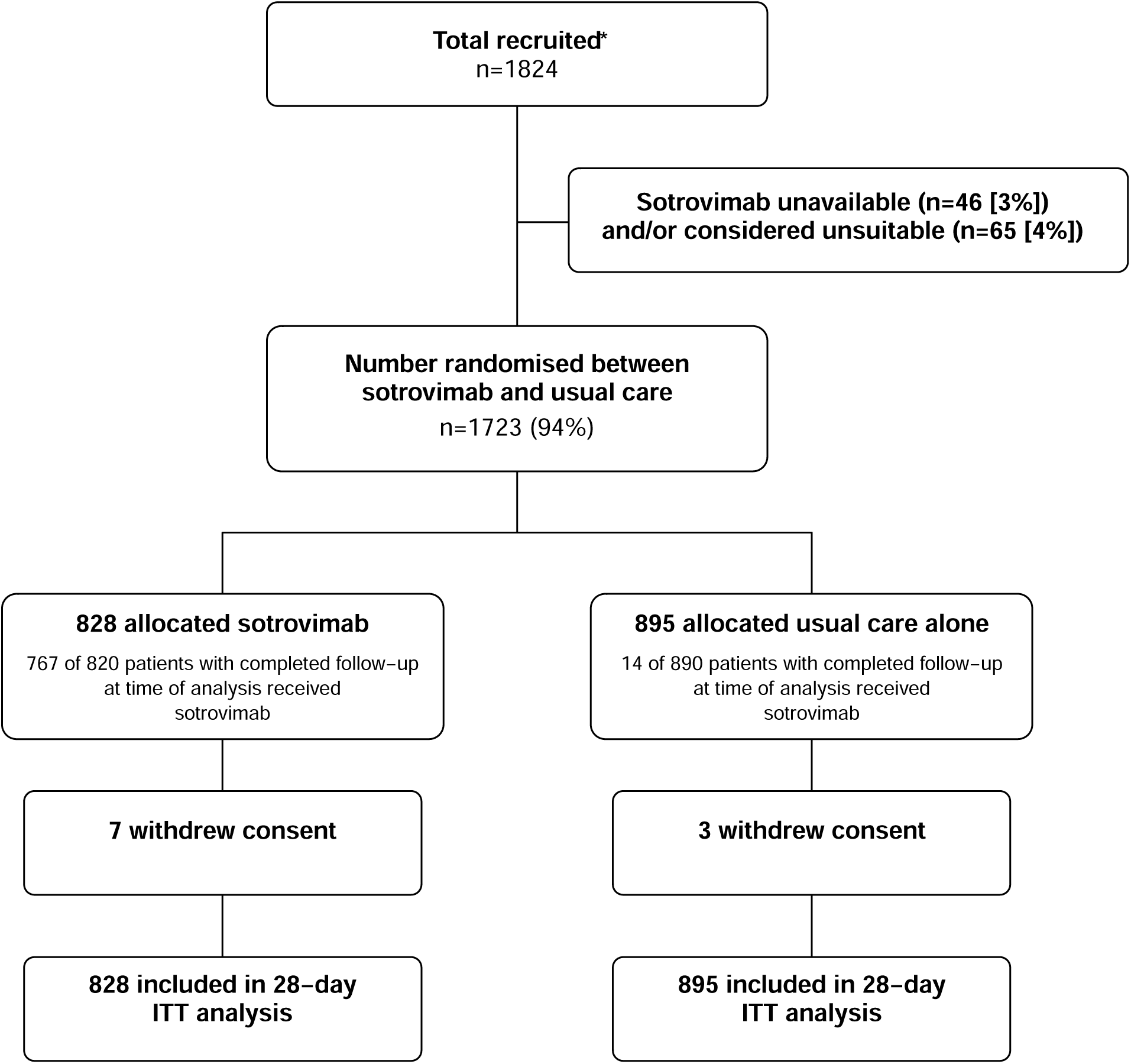
Trial profile. ITT=intention to treat. Drug unavailability and unsuitability are not mutually exclusive. *Number recruited overall at all sites participating in the sotrovimab comparison during period that this comparison was open.

**Table 1:** Baseline characteristics.

|  | High-antigen patients |  | All patients |  |
| --- | --- | --- | --- | --- |
|  | Sotrovimab<br>(n=355) | Usual care<br>(n=365) | Sotrovimab<br>(n=828) | Usual care<br>(n=895) |
| Age, years | 72.5 (13.3) | 72.1 (13.7) | 70.9 (14.2) | 70.4 (15.4) |
| <70 | 123 (35%) | 141 (39%) | 342 (41%) | 369 (41%) |
| ≥70 to <80 | 123 (35%) | 121 (33%) | 251 (30%) | 272 (30%) |
| ≥80 | 109 (31%) | 103 (28%) | 235 (28%) | 254 (28%) |
| Sex |  |  |  |  |
| Male | 218 (61%) | 226 (62%) | 490 (59%) | 543 (61%) |
| Female | 137 (39%) | 139 (38%) | 338 (41%) | 352 (39%) |
| Ethnicity |  |  |  |  |
| White | 301 (85%) | 333 (91%) | 706 (85%) | 779 (87%) |
| Black, Asian, and minority ethnic | 32 (9%) | 16 (4%) | 64 (8%) | 65 (7%) |
| Unknown | 22 (6%) | 16 (4%) | 58 (7%) | 51 (6%) |
| Number of days since symptom onset | 6 (3-11) | 6 (3-12) | 6 (3-11) | 6 (3-11) |
| Number of days since admission to hospital | 2 (1-5) | 2 (1-5) | 2 (1-5) | 2 (1-5) |
| Respiratory support received |  |  |  |  |
| None | 43 (12%) | 54 (15%) | 119 (14%) | 137 (15%) |
| Simple oxygen | 226 (64%) | 213 (58%) | 512 (62%) | 557 (62%) |
| Non-invasive ventilation | 71 (20%) | 87 (24%) | 168 (20%) | 169 (19%) |
| Invasive mechanical ventilation | 15 (4%) | 11 (3%) | 29 (4%) | 32 (4%) |
| Previous diseases |  |  |  |  |
| Diabetes | 107 (30%) | 84 (23%) | 249 (30%) | 219 (24%) |
| Heart disease | 119 (34%) | 113 (31%) | 259 (31%) | 272 (30%) |
| Chronic lung disease | 123 (35%) | 128 (35%) | 327 (39%) | 325 (36%) |
| Tuberculosis | 0 (0%) | 1 (<0.5%) | 2 (<0.5%) | 4 (<0.5%) |
| HIV | 3 (1%) | 1 (<0.5%) | 6 (1%) | 5 (1%) |
| Severe liver disease * | 6 (2%) | 3 (1%) | 19 (2%) | 16 (2%) |
| Severe kidney impairment † | 45 (13%) | 41 (11%) | 84 (10%) | 74 (8%) |
| Any of the above | 242 (68%) | 237 (65%) | 578 (70%) | 602 (67%) |
| Severely immunocompromised ‡ | 112 (32%) | 112 (31%) | 206 (25%) | 208 (23%) |
| Received a COVID-19 vaccine | 296 (83%) | 292 (80%) | 675 (82%) | 714 (80%) |
| Use of other treatments |  |  |  |  |
| Corticosteroids § | 329 (93%) | 334 (92%) | 755 (91%) | 801 (89%) |
| Remdesivir | 144 (41%) | 128 (35%) | 315 (38%) | 313 (35%) |
| Tocilizumab | 66 (19%) | 60 (16%) | 144 (17%) | 137 (15%) |
| Plan to use tocilizumab within the next 24 hours | 28 (8%) | 33 (9%) | 48 (6%) | 67 (7%) |
| Viral load in baseline nose swab |  |  |  |  |
| Median level (log viral copies per ml) | 6.1 (4.6-7.0) | 6.1 (5.0-7.2) | 5.6 (3.7-6.7) | 5.6 (3.9-6.8) |
| Antigen status |  |  |  |  |
| High | 355 (100%) | 365 (100%) | 355 (43%) | 365 (41%) |
| Low | 0 (0%) | 0 (0%) | 339 (41%) | 378 (42%) |
| Unknown | 0 (0%) | 0 (0%) | 134 (16%) | 152 (17%) |
| Serostatus (anti N) |  |  |  |  |
| Positive | 62 (17%) | 76 (21%) | 214 (26%) | 240 (27%) |
| Negative | 293 (83%) | 289 (79%) | 481 (58%) | 504 (56%) |
| Unknown | 0 (0%) | 0 (0%) | 133 (16%) | 151 (17%) |
| Serostatus (anti S) |  |  |  |  |
| Positive | 252 (71%) | 262 (72%) | 569 (69%) | 610 (68%) |
| Negative | 103 (29%) | 103 (28%) | 126 (15%) | 133 (15%) |
| Unknown | 0 (0%) | 0 (0%) | 133 (16%) | 152 (17%) |

Data are mean (SD), n (%), or median (IQR). 4 pregnant women were randomised. \* Defined as requiring ongoing specialist care. † Defined as estimated glomerular filtration rate <30 mL/min per 1.73 m<sup>2</sup>. ‡ In the opinion of the managing clinician. § Including all those randomised into the comparison of high vs low dose steroids.

The follow-up form was completed for 1710 (99%) patients, and among them 767/820 (94%) allocated sotrovimab received the treatment, compared to 14/890 (2%) allocated usual care (figure 1). Use of other treatments for COVID-19 was similar among patients allocated sotrovimab and those allocated usual care (appendix p61). Primary and secondary outcome data are known for more than 99% of randomly assigned patients.

In patients who had high-antigen at baseline, allocation to sotrovimab was associated with a reduction in the primary outcome of 28-day mortality compared with usual care alone: 82/355 (23%) patients in the sotrovimab group died versus 106/365 (29%) patients in the usual care group (rate ratio 0.75; 95% CI 0.56-0.99; p=0.046; table 2, figure 2a, figure 3). Among all patients randomised (including those with high, low, or unknown baseline antigen status), there was no significant difference in the primary outcome of 28-day mortality between the two randomised groups: 177/828 (21%) patients in the sotrovimab group died versus 201/895 (22%) patients in the usual care group (rate ratio 0.95; 95% CI 0.77-1.16; p=0.60; figure 2b, figure 3, appendix p62). There was no evidence that the proportional effects on mortality differed among any pre-specified subgroups among high-antigen patients, or among all patients (figure 4, appendix pp67-69).

**Figure 2:**
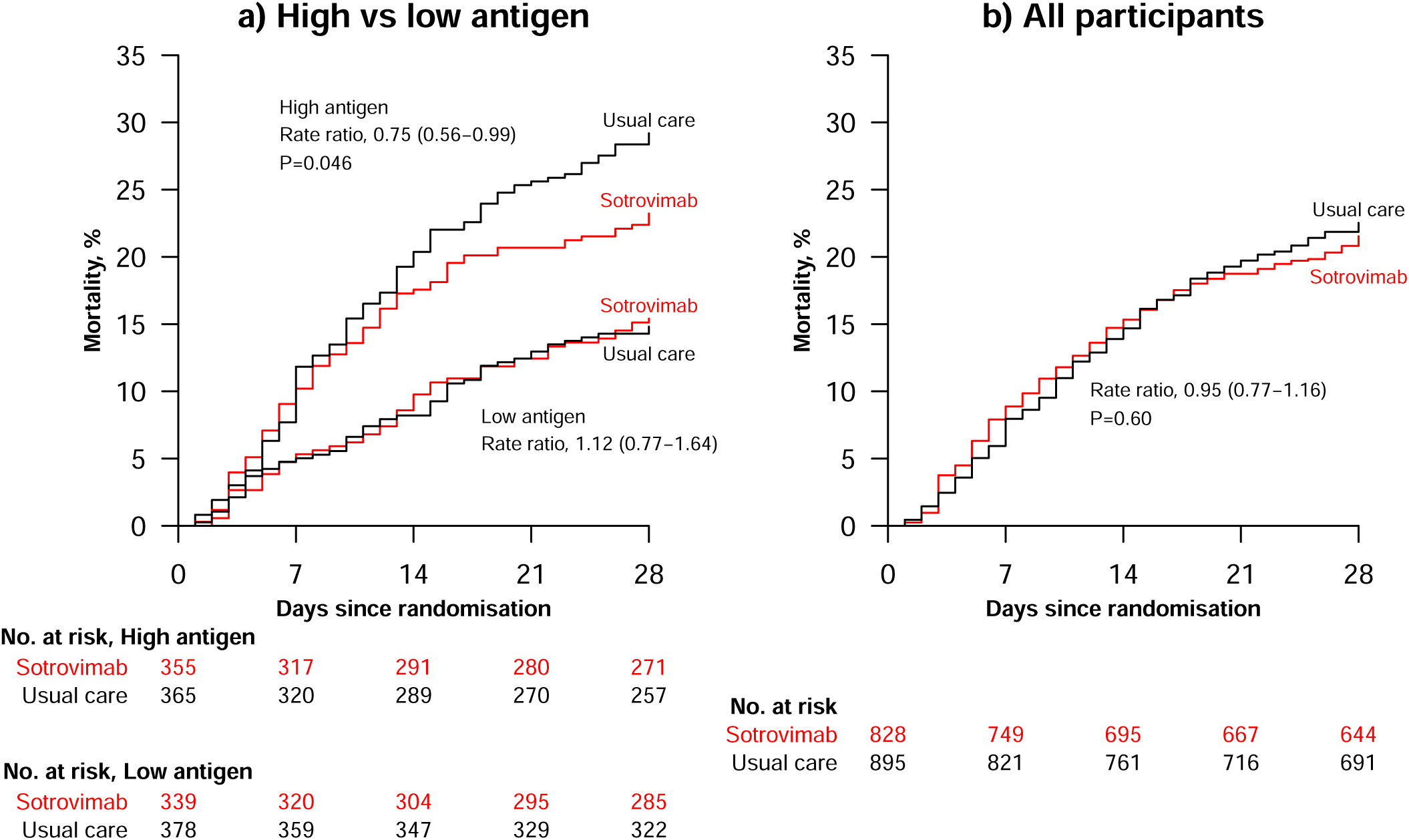
Effect of allocation to sotrovimab on 28-day mortality in (a) high-antigen versus low-antigen patients, and (b) all patients.

**Figure 3:**
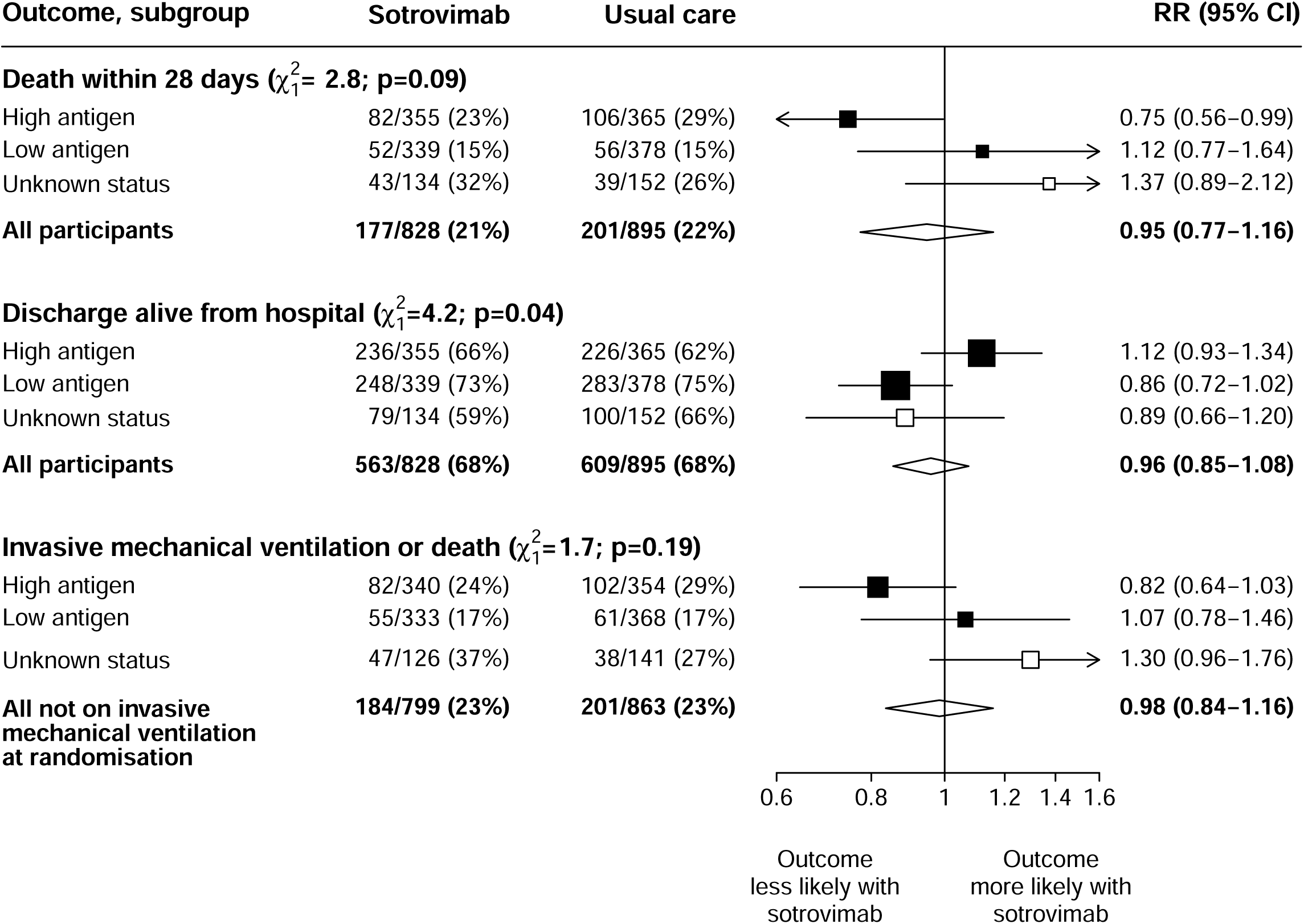
Primary and secondary outcomes, overall and by baseline antigen status. Subgroup-specific RR estimates are represented by squares (with areas of the squares proportional to the amount of statistical information) and the lines through them correspond to the 95% CIs. Open squares represent participants with unknown status, solid squares represent participants with known status. The tests for heterogeneity compare the log RRs in high-antigen versus low-antigen patients (i.e. excluding those with unknown antigen status). All participants are included in the overall summary diamonds. RR=risk ratio for the composite outcome of invasive mechanical ventilation or death, and rate ratio for the other outcomes.

**Figure 4:**
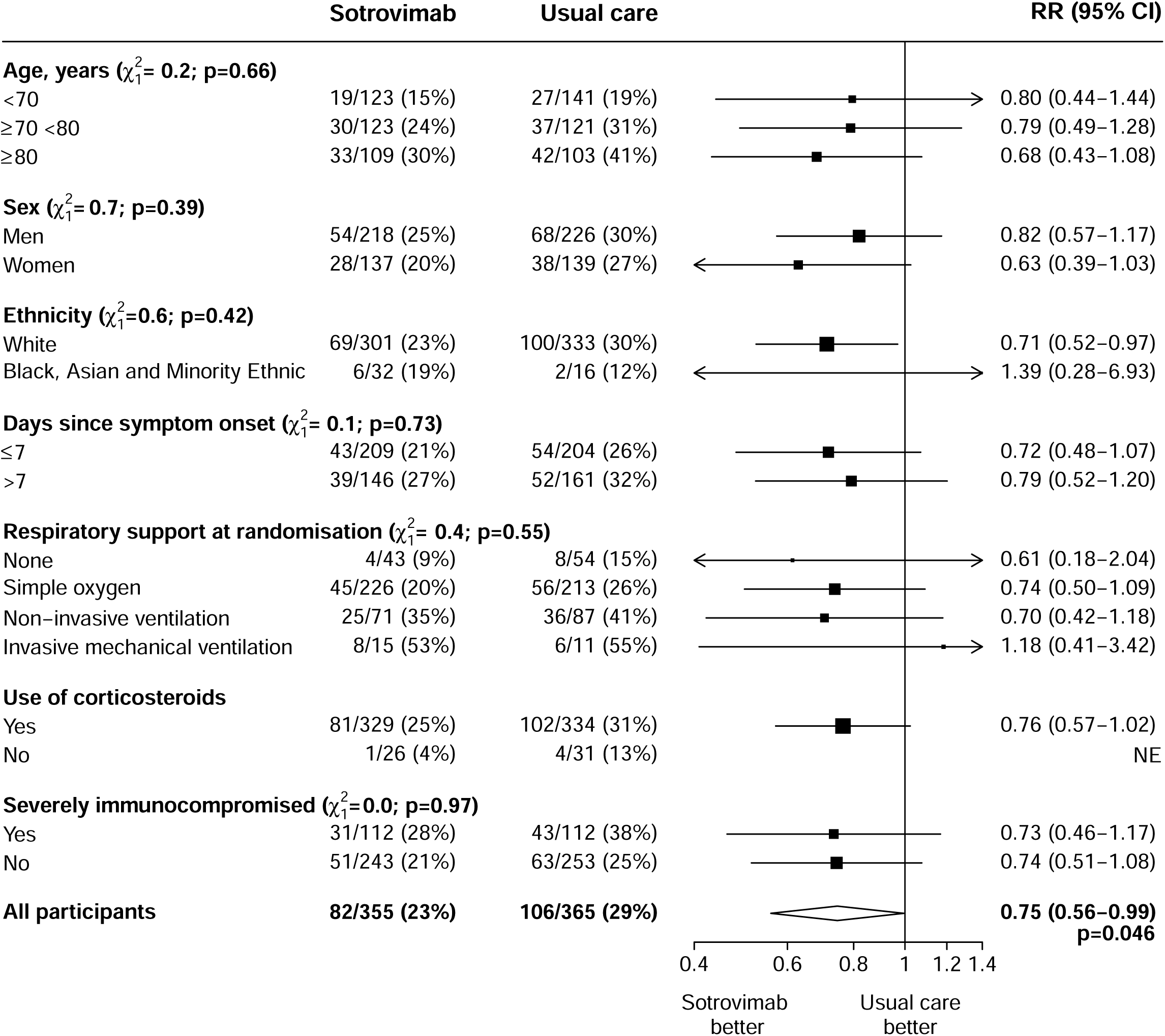
Effect of allocation to sotrovimab on 28-day mortality by baseline characteristics in high-antigen participants. Subgroup−specific rate ratio estimates are represented by squares (with areas of the squares proportional to the amount of statistical information) and the lines through them correspond to the 95% CIs. The ethnicity, days since onset and use of corticosteroids subgroups exclude those with missing data, but these patients are included in the overall summary diamond.

**Table 2:** Effect of allocation to sotrovimab on key study outcomes in patients with among high-antigen levels.

|  | Sotrovimab<br>(n=355) | Usual care<br>(n=365) | RR (95% CI) or<br>mean difference | p-value |
| --- | --- | --- | --- | --- |
| **Primary outcome** |  |  |  |  |
| 28-day mortality | 82 (23%) | 106 (29%) | 0.75 (0.56-0.99) | 0.046 |
| **Secondary outcomes** |  |  |  |  |
| Median (IQR) time to being discharged alive, days | 13 (7 to >28) | 16 (7 to >28) |  |  |
| Discharged from hospital within 28 days | 236 (66%) | 226 (62%) | 1.12 (0.93-1.34) |  |
| Receipt of invasive mechanical ventilation or death* | 82/340 (24%) | 102/354 (29%) | 0.82 (0.64-1.03) |  |
| Invasive mechanical ventilation | 14/340 (4%) | 11/354 (3%) | 1.71 (0.81-3.61) |  |
| Death | 74/340 (22%) | 100/354 (28%) | 0.74 (0.58-0.95) |  |
| **Subsidiary clinical outcomes** |  |  |  |  |
| Use of ventilation† | 41/269 (15%) | 41/267 (15%) | 0.97 (0.66-1.44) |  |
| Non-invasive ventilation | 40/269 (15%) | 41/267 (15%) | 0.95 (0.64-1.41) |  |
| Invasive mechanical ventilation | 6/269 (2%) | 3/267 (1%) | 1.82 (0.47-7.11) |  |
| Successful cessation of invasive mechanical ventilation ‡ | 5/15 (33%) | 3/11 (27%) | 1.07 (0.25-4.65) |  |
| Use of haemodialysis or haemofiltration § | 12/347 (3%) | 6/356 (2%) | 1.97 (0.77-5.06) |  |
| **Virological outcomes** |  |  |  |  |
| Baseline-adjusted viral load (log copies/ml) on day 3 | 4.89 (0.10) | 4.94 (0.10) | -0.05 (-0.32, 0.23) |  |
| Baseline-adjusted viral load (log copies/ml) on day 5 | 4.26 (0.11) | 4.35 (0.10) | -0.09 (-0.38, 0.20) |  |

Data are n (%) or n/N (%), unless otherwise indicated. RR=rate ratio for the outcomes of 28-day mortality and hospital discharge, risk ratio for other clinical outcomes, and mean difference for virological outcomes. CI=confidence interval. Estimates of the RR or mean difference and their 95% CIs are adjusted for age in three categories (<70 years, 70-79 years, and 80 years or older) and ventilation status at randomisation in four categories (none, simple oxygen, non-invasive ventilation, and invasive mechanical ventilation). P-values are not shown for the secondary, subsidiary or virological outcomes because the hierarchical testing strategy pre-specified in the statistical analysis plan stated that such tests would only be performed if null hypothesis for the primary outcome of 28-day mortality was rejected in both the antigen positive subgroup *and* in the whole population \* Excluding patients receiving invasive mechanical ventilation at randomisation. † Excluding patients receiving invasive or non-invasive ventilation at randomisation. ‡ Excluding patients not receiving invasive mechanical ventilation at randomisation. § Excluding patients receiving renal replacement therapy at randomisation.

Among high-antigen patients, discharge alive within 28 days did not differ between those allocated sotrovimab compared to usual care (66% versus 58%; rate ratio 1.12, 95% CI 0.93-1.34; median time to being discharged alive 13 days versus 16 days) (table 2, figure 3). There was also no difference in this outcome among the overall study population (68% versus 68%; rate ratio 0.96, 95% CI 0.85-1.08; median time to being discharged alive 11 days versus 11 days) (figure 3, appendix p62).

Among high-antigen patients not on invasive ventilation at baseline, allocation to sotrovimab was not associated with a lower risk of progressing to the composite secondary outcome of invasive ventilation or death (24% versus 29%, risk ratio 0.82, 95% CI 0.64-1.03) (table 2, figure 3). There was also no difference in this outcome among the overall study population (23% versus 23%, risk ratio 0.98, 95% CI 0.84 to 1.16) (figure 3, appendix p62).

We found no evidence of any difference between groups in the prespecified subsidiary outcomes among high-antigen patients, or among all patients, including in use of ventilation in those not on ventilation at baseline, successful cessation of ventilation, or use of renal replacement therapy (table 2, appendix p62).

1479/1723 (86%) of patients had at least one nose swab available for analysis. Allocation to sotrovimab was not associated with a lower baseline-adjusted viral RNA copy number in nose swabs taken on day 3 or day 5 (table 2). 1119 (65%) patients had at least one successfully sequenced sample, and of those with at least one high quality sample (≥90% genome coverage), 1021/1026 (>99%) were identified as Omicron variants (primarily BA.1, BA.2, BA.5, and XBB). 1655/1723 (96%) patients were recruited before November 2023, and of these 14/1026 (1%) with a sequenced sample had a sotrovimab resistance mutation detected at baseline, and 3/692 (<0.5%) with sequenced baseline and follow-up samples had a new sotrovimab resistance mutation arising after trial entry, two of whom had received sotrovimab (relevant mutations are listed in appendix p34). Among the 68/1723 (4%) patients recruited after 1 November 2023, 14/35 (40%) with a sequenced sample were infected with BA.2.86 variants, which are known to contain the K356T spike mutation associated with high-level sotrovimab resistance.

Infusion reactions were reported for 12/781 (2%) patients receiving sotrovimab. Of these, nine were mild (no intervention required), two moderate (antihistamines or steroids required) and one severe (adrenaline required). Two serious adverse reactions to sotrovimab were reported, both of which were infusion reactions included above, one of which was in a patient with suspected anaphylaxis that resolved with treatment. We found no difference between groups in other safety outcomes, including cause-specific mortality, new cardiac arrhythmia, thrombosis, bleeding, non-coronavirus infections, hypo- or hyper-glycaemia, seizures, acute kidney injury or liver injury (appendix pp63-64).

## DISCUSSION

In this randomised trial including over 1700 patients with COVID-19 pneumonia, sotrovimab was associated with a reduction in 28-day mortality in those with a high serum nucleocapsid antigen concentration, although there was substantial uncertainty about the size of this apparent benefit (RR 0.75; 95% CI 0.56-0.99; p=0.046). An analysis of all patients, regardless of antigen concentration, did not show evidence of any benefit of treatment on 28-day mortality. By contrast with our previous study of monoclonal antibody treatment in this setting, the current study was performed during a period of Omicron infection and widespread vaccination and natural immunity.^4^

The number of patients hospitalised with COVID-19 pneumonia fell dramatically after vaccination was introduced and Omicron became dominant, so this comparison could not provide results as definitive as those of the earlier RECOVERY casirivimab-imdevimab comparison that recruited nearly 10,000 patients. However, the pattern of results from the two RECOVERY mAb comparisons are similar, despite using different markers of infection status to categorise patients. In both, a subset of patients with immune responses that were not yet adequate to clear infection were at higher risk of death than patients with more robust immune responses, and in that higher risk subset mAb therapy reduced the risk of death. During the period this comparison was recruiting, SARS-CoV-2 infection in hospitalised patients was often an incidental finding or associated with non-respiratory illness, and the benefits of antiviral therapy in these patients may be limited. In contrast, RECOVERY only included those with pneumonia thought to be related to COVID-19 and in 81% of participants this had developed despite previous COVID-19 vaccination. In keeping with this, over 80% of those with known serostatus had anti-spike antibodies, although two-thirds were anti-nucleocapsid antibody negative, indicating that this was likely their first SARS-CoV-2 infection.^32^ The risk of death from COVID-19 was high. 28-day mortality was 22% in those allocated usual care, similar to the risk among RECOVERY patients recruited in the pre-Omicron era. Since the emergence of Omicron, immunocompromised patients have made up a higher proportion of those hospitalised and dying from COVID-19 pneumonia, and in keeping with this one-quarter of the RECOVERY patients were considered severely immunocompromised.^33^

The benefit of mAb therapy in SARS-CoV-2 antibody negative hospitalised patients was established in previous trials, but this approach to targeting therapy was necessarily short-lived in the context of increasing population immunity.^4–6^ In contrast, targeting therapy on the basis of antigenaemia remains possible for future hospitalised patients, and is practical using existing commercial assays (the one used in RECOVERY takes 20 minutes on a widely available automated clinical laboratory platform). The ACTIV-3/TICO platform trial is the only previous trial of mAb therapy reporting outcomes by baseline blood antigen status, and this evaluated four mAb therapies, although three of these were stopped early for futility.^6,14,34^ In the single comparison not stopped early, 1417 hospitalised patients were randomised to receive tixagevimab-cilgavimab or placebo. Among patients with blood antigen above the median value, 90-day mortality was 43/340 (13%) in those allocated mAb versus 51/342 (15%) in those allocated placebo (hazard ratio 0.84; 95%CI 0.56-1.26; p=0.39); although inconclusive the point estimate is consistent with this RECOVERY result that is based on twice as many events.

Neutralising mAbs emerged as powerful therapeutic tools during the pandemic, which highlighted their potential uses but also their limitations, particularly the loss of activity against emergent viral variants. Despite retaining potentially valuable neutralising activity against Omicron variants prevalent in 2022-23, high-level sotrovimab resistance was identified in Omicron lineages that became globally dominant in early 2024, including BA.2.86 and JN.1, and it is no longer likely to have useful activity against currently circulating variants that have retained sotrovimab resistance mutations.^35^ The loss of all anti-SARS-CoV-2 mAbs that were in clinical use has led to new approaches to mAb therapy, including attempts to target more highly conserved viral epitopes, new antibody fragments or formulations that may have better potency or tissue penetration, and antibody cocktails or poly-specific antibodies that may be more robust to viral evolution.^36^ The results of this comparison suggest that if new mAb therapies can be developed that effectively neutralise current and future SARS-CoV-2 variants then they could continue to benefit hospitalised patients.

Most patients in the RECOVERY sotrovimab comparison were recruited in 2022, and, other than lineage-defining Omicron mutations, there were few important sotrovimab resistance mutations identified in either baseline or follow-up samples. Because of concerns about possible reduced sotrovimab activity against BA.1, a 1g dose was used in RECOVERY rather than the 500mg dose tested previously, and this was well tolerated with no new safety concerns. The lack of any measurable effect of sotrovimab on nasal SARS-CoV-2 carriage by day 5 may be related to the early sampling timepoints used, as even in seronegative patients treated with a well-matched mAb, a reduction in carriage of viral RNA is mainly apparent from day 7 onwards.^5^ In contrast to changes in viral RNA carriage, a large reduction in culturable SARS-CoV-2 can be seen as early as 24 hours after mAb therapy, but virological testing in RECOVERY did not extend to culture.^37^

Strengths of this trial include that it was randomised, had broad eligibility criteria, and a large sample size, being the second largest trial of mAb therapy performed in patients hospitalised with COVID-19. It includes baseline characterisation of markers of SARS-CoV-2 immune status and infection, and more than 99% of patients were followed up for the primary and secondary outcomes. The study has some limitations: the use of serum antigen to define the primary analysis population was prespecified, but this is a novel therapeutic biomarker and there is little existing evidence to support the threshold used to classify patients. The distribution of serum antigen in our population was unimodal with no natural cut-point, so other thresholds could have been selected. In a larger trial it may have been possible to retrospectively identify an optimal antigen threshold, but this kind of sensitivity analysis would not be robust in our study given the limited number of outcome events. Use of other antiviral treatments was relatively common, with remdesivir received by one-third of patients, and it is possible that sotrovimab would have had a greater effect in the absence of other antivirals. The RECOVERY trial is open label, so participants and local hospital staff were aware of the assigned treatment. This could potentially have affected clinical management or the recording of some trial outcomes, although we found no evidence that management differed by treatment allocation (appendix p61), and the primary and secondary outcomes are unambiguous and were ascertained without bias through linkage to routine health records. Although virological outcomes were included, this did not include viral culture, and no information on radiological or physiological outcomes was collected. The RECOVERY trial only studied patients who had been hospitalised with COVID-19 and, therefore, is not able to provide any evidence on the safety and efficacy of treatments in other patient groups, such as those with early infection.

In summary, the results of this randomised trial suggest that some hospitalised COVID-19 patients at high risk of death could continue to benefit from mAb therapy, and that antigen testing could help to identify these patients. Although no currently available mAbs have satisfactory activity against current SARS-Cov-2 variants, these results should inform future mAb evaluation and treatment strategies.

## Data Availability

The protocol, consent form, statistical analysis plan, definition & derivation of clinical characteristics & outcomes, training materials, regulatory documents, and other relevant study materials are available online at www.recoverytrial.net. As described in the protocol, the Trial Steering Committee will facilitate the use of the study data and approval will not be unreasonably withheld. Deidentified participant data will be made available to bona fide researchers registered with an appropriate institution within 3 months of publication. However, the Steering Committee will need to be satisfied that any proposed publication is of high quality, honours the commitments made to the study participants in the consent documentation and ethical approvals, and is compliant with relevant legal and regulatory requirements (e.g. relating to data protection and privacy). The Steering Committee will have the right to review and comment on any draft manuscripts prior to publication. Data will be made available in line with the policy and procedures described at: https://www.ndph.ox.ac.uk/data-access. Those wishing to request access should complete the form at https://www.ndph.ox.ac.uk/files/about/data_access_enquiry_form_13_6_2019.docx and e-mailed to

## Contributors

This manuscript was initially drafted by LP, RH, PWH and MJL, further developed by the Writing Committee, and approved by all members of the trial steering committee. NS, JRE, PWH, MJL, RH and LP had access to the study data. NS and JRE accessed and verified the data. JRE did the statistical analysis. PWH and MJL vouch for the data and analyses, and for the fidelity of this report to the study protocol and data analysis plan, and had final responsibility for the decision to submit for publication. PWH, NS, JRE, JKB, MB, SNF, TJ, EJ, KJ, MK, WSL, AMo, AMuk, AMum, KR, GT, MM, RH, and MJL designed the trial and study protocol. MM, MC, G P-A, LP, RS, DG, GC, FH, JM-C, PD, PH, JU, NE, JM, SB, the Data Linkage team at the RECOVERY Coordinating Centre, and the Health Records and Local Clinical Centre staff listed in the appendix collected the data. All authors contributed to data interpretation and critical review and revision of the manuscript.

## Data Monitoring Committee

Peter Sandercock, Janet Darbyshire, David DeMets, Robert Fowler, David Lalloo, Mohammed Munavvar, Janet Wittes.

## Declaration of interests

The authors have no conflict of interest or financial relationships relevant to the submitted work to disclose. No form of payment was given to anyone to produce the manuscript. All authors have completed and submitted the ICMJE Form for Disclosure of Potential Conflicts of Interest. The Nuffield Department of Population Health at the University of Oxford has a staff policy of not accepting honoraria or consultancy fees directly or indirectly from industry (see https://www.ndph.ox.ac.uk/files/about/ndph-independence-of-research-policy-jun-20.pdfhttps://www.ctsu.ox.ac.uk/about/ctsu_honoraria_25june14-1.pdf).

## Acknowledgements

Above all, we would like to thank the patients who participated in this trial. We would also like to thank the many doctors, nurses, pharmacists, other allied health professionals, and research administrators at NHS hospital organisations across the whole of the UK, supported by staff at the National Institute of Health and Care Research (NIHR) Clinical Research Network, NHS DigiTrials, UK Health Security Agency, Department of Health & Social Care, the Intensive Care National Audit & Research Centre, Public Health Scotland, National Records Service of Scotland, the Secure Anonymised Information Linkage (SAIL) at University of Swansea, and the NHS in England, Scotland, Wales and Northern Ireland.

The RECOVERY trial was supported by grants to the University of Oxford from UK Research and Innovation (UKRI) and NIHR (MC_PC_19056), and by core funding provided by the NIHR Oxford Biomedical Research Centre, the Wellcome Trust, the Bill and Melinda Gates Foundation, the Foreign, Commonwealth and Development Office, Health Data Research UK, the Medical Research Council, the NIHR Health Protection Unit in Emerging and Zoonotic Infections, and NIHR Clinical Trials Unit Support Funding. TJ is supported by grants from the UK Medical Research Council (MC_UU_0002/14 and MC_UU_00040/03). WSL is supported by core funding provided by NIHR Nottingham Biomedical Research Centre. JU is supported by a UK Medical Research Council grant (MR/T023791/1).

The views expressed in this publication are those of the authors and not necessarily those of the NHS, the NIHR, or the UK Department of Health and Social Care.

## Notes

### Competing Interest Statement

The authors have declared no competing interest.

### Clinical Trial

ISRCTN50189673 and NCT04381936

### Clinical Protocols

https://www.recoverytrial.net/

### Author Declarations

Approval was given by the Cambridge East Research Ethics Committee (UK)

