## Supplementary Material for "Sotrovimab versus usual care in patients admitted to hospital with COVID-19: a randomised, controlled, open-label, platform trial (RECOVERY)"

#### SUPPLEMENTARY APPENDIX

**RECOVERY Collaborative Group**

Webtable 1: Baseline characteristics of patients considered unsuitable for randomisation to sotrovimab compared with those randomised to sotrovimab versus usual care 59
Webtable 2: Baseline characteristics by patient baseline antigen status and randomised allocation 60
Webtable 3: Treatments given (antigen positive and all patients) by randomised allocation 61
Webtable 4: Effect of allocation to sotrovimab on key study outcomes in all patients 62
Webtable 5: Effect on cause-specific 28-day mortality 63
Webtable 6: Effect on non-coronavirus infection, new cardiac arrhythmia, thrombotic events, clinically significant bleeds, metabolic complications and other outcomes 64
**Supplementary Figures** .................................................................................. 65
Webfigure 1: Distribution of serum antigen values in trial participants 66
Webfigure 2: Effect of allocation to sotrovimab on 28−day mortality in ALL PARTICIPANTS by baseline characteristics 67
Webfigure 3: Primary and secondary outcomes, overall and by Roche anti−S status 68
Webfigure 4: Primary and secondary outcomes, overall and by Roche anti−N status 69
**Appendices** .................................................................................. 70
Appendix 1: RECOVERY Trial Protocol V27.0 71
Appendix 2: RECOVERY Trial Statistical Analysis Plan V5 114
Appendix 3: Definition and Derivation of Baseline Characteristics and Outcomes 164
Appendix 4: Validation of serum nucleocapsid antigen assay 188

### Details of the RECOVERY Collaborative Group

#### Writing Committee

Peter W Horby[a,b,c]*, Jonathan R Emberson[d]*, Leon Peto[d,e]*, Natalie Staplin[d], Mark Campbell[d,e], Guilherme Pessoa-Amorim[d], Richard Stewart[f], Dipansu Ghosh[g], Graham Cooke[h], Natalie Blencowe[i], Jeronimo Moreno-Cuesta[j], Purav Desai[k], Paul Hine[l], Jonathan Underwood[m], Nicholas Easom[n,o], Jaydip Majumdar[p], Sanjay Bhagani[q], J Kenneth Baillie[r], Maya H Buch[s], Saul N Faust[t], Thomas Jaki[u,v], Katie Jeffery[e,w], Edmund Juszczak[x], Marian Knight[y], Wei Shen Lim[z], Alan Montgomery[x], Aparna Mukherjee,[za] Andrew Mumford[zb], Kathryn Rowan[zc], Guy Thwaites[a,zd], Marion Mafham[d†], Richard Haynes[d,e†], Martin J Landray[d,ze†]

[a] Centre for Tropical Medicine and Global Health, Nuffield Department of Medicine, University of Oxford, Oxford, United Kingdom

[b] International Severe Acute Respiratory and emerging Infections Consortium (ISARIC), University of Oxford, Oxford, United Kingdom

[c] Pandemic Sciences Centre, University of Oxford, Oxford, United Kingdom

[d] Nuffield Department of Population Health, University of Oxford, Oxford, United Kingdom

[e] Oxford University Hospitals NHS Foundation Trust, Oxford, United Kingdom

[f] Milton Keynes University Hospital, Milton Keynes, United Kingdom

[g] Leeds Teaching Hospitals NHS Foundation Trust, Leeds, United Kingdom

[h] Imperial College Healthcare NHS Trust, London, United Kingdom

[i] University Hospitals Bristol and Weston NHS Foundation Trust and University of Bristol, Bristol, United Kingdom

[j] North Middlesex University Hospital NHS Trust

[k] Calderdale and Huddersfield NHS Foundation Trust, Huddersfield, United Kingdom

[l] Liverpool University Hospitals NHS Foundation Trust, Liverpool, United Kingdom

[m] Cardiff & Vale University LHB, Cardiff, United Kingdom

[n] Hull York Medical School, Hull, United Kingdom

[o] Hull University Teaching Hospitals NHS Trust, Hull, United Kingdom

[p] Mid Cheshire Hospitals NHS Foundation Trust, United Kingdom

[q] Royal Free London NHS Foundation Trust, London, United Kingdom

[r] Roslin Institute, University of Edinburgh, Edinburgh, United Kingdom

[s] Centre for Musculoskeletal Research, University of Manchester, Manchester, United Kingdom

[t] NIHR Southampton Clinical Research Facility and Biomedical Research Centre, University Hospital Southampton NHS Foundation Trust and University of Southampton, Southampton, United Kingdom

[u] Faculty for Informatics and Data Science, University of Regensburg, Germany

[v] MRC Biostatistics Unit, University of Cambridge, UK

[w] Radcliffe Deparment of Medicine, University of Oxford, United Kingdom

[x] School of Medicine, University of Nottingham, Nottingham, United Kingdom

[y] National Perinatal Epidemiology Unit, University of Oxford, United Kingdom

[z] Respiratory Medicine Department, Nottingham University Hospitals NHS Trust, Nottingham, United Kingdom

[za] Indian Council of Medical Research, New Delhi, India

[zb] School of Cellular and Molecular Medicine, University of Bristol, Bristol, United Kingdom

[zc] Intensive Care National Audit & Research Centre, London, United Kingdom

[zd] Oxford University Clinical Research Unit, Ho Chi Minh City, Viet Nam

[ze] NIHR Oxford Biomedical Research Centre, Oxford University Hospitals NHS Foundation Trust, Oxford, United Kingdom

*,[†] equal contribution

**Steering Committee**

*Co-Chief Investigators* PW Horby, MJ Landray, *Members* JK Baillie, M Buch, L Chappell (until August 2021), J Day (until May 2022), SN Faust, R Haynes, T Jaki, K Jeffery, E Juszczak, M Knight (from August 2021), WS Lim, M Mafham, A Montgomery, A Mukherjee, A Mumford, K Rowan, G Thwaites.

**International Committee**

*Chair* DV Dung *Regional Lead Investigators* G Thwaites, J Day *Independent members* NN Quang, Prof. Binh, E Burhan, B Alisjahbana, J Koirala, S Basnet *Other members* E Kestelyn, B Basnyat, P Gyanwali, RL Hamers, P Horby

**Data Monitoring Committee**

P Sandercock (chair), J Darbyshire, D DeMets, R Fowler, D Lalloo, M Munavvar (from January 2021), I Roberts (until December 2020), A Warris (from March 2021), J Wittes *Non-voting statisticians* J Emberson, N Staplin.

**RECOVERY Trial Central Coordinating Office**

*Co-Chief Investigators* P Horby, MJ Landray; *Clinical Trial Unit Lead* R Haynes; *Trial management* A Cradduck-Bamford (coordinator), J Barton, A Basoglu, R Brown, W Brudlo, E Denis, L Fletcher, S Howard, S Musini, K Taylor; *Programming and validation* G Cui, B Goodenough, A King, M Lay, D Murray, W Stevens, K Wallendszus, R Welsh; *Data linkage* C Crichton, J Davies, R Goldacre, C Harper, F Knight, M Mafham, M Nunn, H Salih, J Welch; *Clinical support* M Campbell, G Pessoa-Amorim, L Peto, M Zayed; *Quality assurance* J Wiles; *Statistics* J Emberson, E Juszczak, E Spata, N Staplin; *Communications* G Bagley, S Cameron, S Chamberlain, B Farrell, H Freeman, A Kennedy, A Whitehouse, S Wilkinson, C Wood; *Evidence synthesis (Vascular Overviews Group)* C Reith (coordinator), K Bird, K Davies, H Halls, L Holland, A Roddick, R Truell, K Wilson; *Administrative support* L Howie, M Lunn, P Rodgers

**RECOVERY Trial Regional Coordinating Centres**

Vietnam: *Oxford University Clinical Research Unit* G Thwaites, C Thwaites, E Kestelyn, NTH Thuong, TB Huyen, NT Thao, NB Tran, NTT Nguyen, NTP Dung, NTH Quyen, VTK Thi, NTH Trang, R van Doorn, HX Nam, PTT Hien, NY Nhi, NH Khanh

Nepal: *Nepal Health Research Council* P Gyanwali, M Dhimal, S Pant *Oxford University Clinical Research Unit* B Basnyat, A Karkey, S Rijal, S Shrestha

Indonesia: *Oxford University Clinical Research Unit Indonesia* RL Hamers, K Baird, K Puspatriani, M Rahardjani, A Rimainar, F Wulandari *Faculty of Medicine Universitas Indonesia* EJ Nelwan

India: *Indian Council for Medical Research* A Mukherjee, JJ Cherian, G Kumar

South Africa: *Wits Health Consortium* J Nel, H Rees

Ghana: *Kumasi Center for Collaborative Research in Tropic Medicine* J Amuasi, O Maiga, J Bonney, E Matey, A Afum-Adjei Awuah, H Hasford, A Amoako Adusei

Gambia: *MRC Unit The Gambia at LSHTM* B Nadjm, A Rocca, U D'Alessandro, E Usuf, M Bittaye, C Roberts, A Jagne

**SARS-CoV-2 antibody and antigen testing laboratory**

*Emerging Pathogen Serology group, Vaccine Development and Preparedness Centre, UK Health Security Agency, Porton Down:* A Otter (lead), B Hicks, T Coleman, C Rowe, J Hewson, G Hemingway, H Selman, A Lloyd, S Francis, H White, H Lewis, M Royds, E Cross, S Millward, S D'Arcangelo, S Jones, T Jones, V Summer, N Berry, K Ebisine, L Gatcombe, D Blick, C Wilson

**SARS-CoV-2 PCR and sequencing laboratory**

*Oxford University Hospitals NHS Foundation Trust, Oxford:* L Dunn (lead), J Bastable, B Constantinides, D Crook, A Justice, S Oakley, J Swan

**UK National Institute for Health Research Clinical Research Network**

*Coordinating Centre* A Barnard, J Beety, C Birch, M Brend, E Chambers, L Chappell, S Crawshaw, C Drake, H Duckles-Leech, J Graham, T Harman, H Harper, S Lock, K Lomme, N McMillan, I Nickson, U Ohia, E OKell, V Poustie**,** S Sam, P Sharratt, J Sheffield, H Slade, W Van't Hoff, S Walker, J Williamson; *Urgent Public Health Clinical Links* A De Soyza, P Dimitri, SN Faust, N Lemoine, J Minton; *East Midlands* K Gilmour, K Pearson *Eastern* C Armah, D Campbell, H Cate, A Priest, E Thomas, R Usher; *North East & North Cumbria* G Johnson, M Logan, S Pratt, A Price, K Shirley, E Walton, P Williams, F Yelnoorkar; *Kent, Surrey & Sussex* J Hanson, H Membrey, L Gill, A Oliver; *North West London* S Das, S Murphy, M Sutu; *Greater Manchester* J Collins, H Monaghan, A Unsworth, S Beddows; *North West Coast* K Barker-Williams, S Dowling, K Gibbons, K Pine; *North Thames* A Asghar, P Aubrey, D Beaumont-Jewell, K Donaldson, T Skinner; *South London* J Luo, N Mguni, N Muzengi, R Pleass, E Wayman; *South West Peninsula* A Coe, J Hicks, M Hough, C Levett, A Potter, J Taylor; *Thames Valley and South Midlands* M Dolman, L Gerdes, C Hall, T Lockett, D Porter *Wessex* J Bartholomew, L Dowden, C Rook, J Walters; *West of England* E Denton, H Tinkler; *Yorkshire & Humber* A Alexander, H Campbell, K Chapman, A Hall, A Rodgers; *West Midlands* P Boyle, M Brookes, C Callens, H Duffy, C Green, K Hampshire, S Harrison, J Kirk, M Naz, L Porter, P Ryan, J Shenton, J Warmington; *Devolved nations* M Amezaga, P Dicks, J Goodwin, H Hodgson, S Jackson, M Odam, D Williamson.

**Paediatric working group**

SN Faust (coordinator), A Bamford, S Bandi, J Bernatoniene, K Cathie, P Dmitri, S Drysdale, M Emonts, J Evans, A Finn, P Fleming, J Furness, C Gale, R Haynes, CE Jones, E Juszczak, D Jyotish, D Kelly, C Murray, N Pathan, L Pollock, A Ramanan, A Riordan, C Roehr, M Wan, E Whittaker.

**Obstetric working group**

M Knight (coordinator), K Hodson, S Pavord, C Williamson.

**Clinical support**

*Clinical Trial Service Unit Out of Hours clinical support* L Bowman, F Chen, R Clarke, M Goonasekara, R Haynes, W Herrington, P Judge, M Mafham, K Mayne, S Ng, D Preiss, C Reith, E Sammons, D Zhu.

**Health records**

*NHS DigiTrials, Southport* H Pinches, P Bowker, V Byrne-Watts, G Chapman, G Coleman, J Gray, A Rees, MJ Landray, M Mafham, N Mather, T Denwood; *Intensive Care National Audit & Research Centre, London* D Harrison; *National Records of Scotland* G Turner; *Public Health Scotland* J Bruce; *SAIL Databank, University of Swansea* C Arkley, S Rees.

**Local Clinical Centre RECOVERY trial staff**
(listed in descending order of the number of patients randomised per site)

**Milton Keynes University Hospital NHS Foundation Trust** R Stewart (PI), J Alin, L Anguvaa, J Bae, G Bega, S Bowman, A Chakraborty, E Clare, S Fox, S Franklin, S George, L How, M Kennedy, J Mead, L Mew, D Mital, L Moran, E Mwaura, M Nathvani, A Rose, D Scaletta, S Shah, L Siamia, O Spring, S Sutherland, F Teasdale, S Velankar, L Wren, F Wright.

**Leeds Teaching Hospitals NHS Trust** D Ghosh (PI), S Ahmed, A Ashworth, N Balatoni, M Baum, L Bonney, J Calderwood, E Carter, S Charlton, J Clarke, C Coupland, M Crow, C Favager, J Glossop, J Hemingway, S Hemphill, K Holliday, A Humphries, S James, K Johnson, A Jones, M Kacar, K Khokhar, P Lewthwaite, A Marcyniuk, G Martin, F McGill, J Minton, D Mistry, J Murira, Z Mustufvi, S O'Riordan, K Robinson, G Saalmink, R Saman (Associate PI), D Singh, B Staniforth, S Straw, A Westwood.

**Imperial College Healthcare NHS Trust** G Cooke (PI), L Young (Co-PI), O Adedeji, E Adewunmi, Z Al-Saadi, R Ashworth, J Barnacle, N Bohnacker, A Cann, F Cheng, J Clark, S Cooray, S Darnell, A Daunt (Associate PI), V Dave, A D'Mello, L Evison, S Fernandez Lopez, F Fitzgerald, C Gale, M Gibani, S Hamilton, S Hunter, A Jimenez Gil, S Johal, B Jones, A Kountourgioti, J Labao, V Latham, N Madeja, S Mashate, C Matthews, H McLachlan, A Mehar, J Millard (Associate PI), M Molina, A Perry, H Rafferty, S Rey, S Ryder, R Shah, E Sidebotham, R Thomas, D Thornton, J Tuff, E Whittaker, C Wignall, P Wilding, C Wong, T Yates (Associate PI), C Yu.

**University Hospitals Bristol and Weston NHS Foundation Trust** G Hamilton (PI), N Blencowe (Co-PI), E Stratton (Co-PI), M Abraham, D Adams, B Al-Ramadhani, B Amit, A Archer (Associate PI), G Asher (Associate PI), G Aziz, A Balcombe, K Bateman, M Baxter, L Beacham, K Beard, K Belfield (Associate PI), N Bell, M Beresford, J Bernatoniene, A Bhat, D Bhojwani, S Biggs, C Blair, J Blazeby, K Bobruk, S Brooks, N Brown, L Buckley, P Butler, A Cannon, C Caws, E Chakkarapani, K Chatar, D Chatterton, B Chivima, E Clark, C Clemente de la Torre, K Cobain, H Cooke, D Cotterill, E Courtney, S Cowman, K Coy, H Crosby, K Curtis, P Davis, O Drewett, K Druryk, R Duncan, H Dymond, K Edgerley, M Ekoi, M Elokl, B Evans, T Farmery, N Fineman, A Finn, L Gamble, F Garty, B Gibbison, L Gourbault, D Grant, K Gregory, M Griffin, R Groome, L Gurung, V Haile, M Hamdollah-Zadeh, A Hannington, R Harrison, J Heywood, A Hindmarsh, N Holling, C Horrobin, R Houlihan, J Hrycaiczuk, H Hudson, K Hurley, J Iqbal, R Jarvis, B Jeffs, A Jones, R E Jones, E King-Oakley, E Kirkham, L Kirkpatrick, R Kumar, M Kurdy, A Lagnado, S Lang, L Leandro, H Legge, F Loro, A Low, H Martin, J Mayer, T Mayo, L McCullagh, G McMahon, L Millett, K Millington, J Mok, J Moon, L

Morgan, S Mulligan, L Murray, T Nandwani, C O'Donovan, E Payne, C Penman, M Pezard-Snell, J Pickard, M Pitchford, C Plumptre, D Putensen, A Ramanan, J Ramirez, S Ratcliffe, N Redman, E Robbins, V Roberts, J Robinson, M Roderick, S Scattergood, A Schadenberg, E Schofield, R Sheppeard, C Shioi, J Shurlock, D Simpson, P Singhal, A Skorko, B Smart, N Smith, R Squires (Associate PI), V Stefania, C Stewart, M Stuttard, P Sugden, S Sundar, C Swanson-Low, T Swart, E Swift, A Tate, M Thake, K Thompson, M Trevelyan, K Turner, S Turner, A Tyer, S Vergnano, R Vincent, R Ward, A White, S Wilkinson, J Williams, S Williams, J Willis, H Winter, Z Woodward, L Woollen, R Wright, A Younes Ibrahim.

**North Middlesex University Hospital NHS Trust** J Moreno-Cuesta (PI), A Dhariwal (Co-PI), S Jain (Co-PI), T Amin, D Bakthavatsalam, P Brooks, M Farah, A Govind, A Haldeos, K Leigh-Ellis, T Light, S Rokadiya, C Vansomeren, R Vincent, L Walker.

**Calderdale and Huddersfield NHS Foundation Trust** P Desai (PI), A Abbott, K Abouelela, U Akudi, D Appleyard, L-A Bayo, D Bromley, N Chambers, M Collins, S Dale, L Gledhill, J Goddard, J Greig, K Hallas, K Hanson, K Holroyd, M Home, D Kelly, A Maharajh, L Matapure, S Mellor, E Merwaha (Associate PI), M Prior-Ong, K Rajalingam, H Riley, M Robinson, C Rourke, K Sandhu, K Schwarz, N Scriven, L Shaw, L Terrett, M Thompson (Associate PI), G Turner, M Usher, A Wilson, T Wood.

**Liverpool University Hospitals NHS Foundation Trust** P Hine (PI), P Albert (Co-PI), S Todd (Co-PI), I Welters (Co-PI), D Wootton (Co-PI), M Ahmed, R Ahmed, A Al Balushi, M Anderson, R Anderson, Z Ashfak, A Atomode, R Ball, P Banks, D Barr, J Bassett, A Bennett, H Bond, A Bracken, T Brankin-Frisby (Associate PI), G Bretland, P Brinksman, M Brodsky, J Brown, H Burhan, C Burston, J Byrne, F Carlin, S Casey, L Chambers, D Coey, T Cross, J Cruise, J Currie, S D'Souza, L Dobbie, R Downey, A Du Thinh, G Duncan, I Duru, J Early, T Evans, K Fenlon, J Fernandez Roman, I Fordham, H Frankland, S Glynn, J Goodall, S Gould, A Gureviciute, J Hackett, K Haigh, M Hamilton, L Hampson, A Hanson, M Harrison, L Hawker, P Hazenberg, D Heath, S Hicks, S Hope, M Howard, K Hunter, T Ingram, A Islim, K Janes, B Johnston, S Karmali, S Kavanagh, L Keogan, W Khan, S King, K Krasauskas, J Lewis, M Lofthouse, P Lopez, C Lowe, Z Mahmood, F Malein, K Martin, A Mediana, L Melling, Z Mellor, P Merron, B Metcalfe, M Middleton, K Monsell, A Morgan, H Murphy, N Nicholas, A Nuttall, L Oliver, R Osanlou, J Parsons, L Pauls, L Pilling, R Price-Eland, C Prince, S Pringle, E Richardson, L Rigby, M Riley, A Rowe, E Rybka, M Samuel, D Scanlon, J Sedano, D Shaw, F Shiham, C Smith (Associate PI), S Stevenson, A Stockdale, J Tempany, P Thu-Ta, C Toohey, I Turner-bone, S Victoria, A Waite, E Wasson, A Watkin, R Watson, V Waugh, R Westhead, L Wilding, K Williams, A Wood, A Yeoh, D Zeinali.

**Cardiff & Vale University LHB** M Wise (PI), J Underwood (Co-PI), A Balan, B Basker, S Bird, Z Boult, V Britten, L Broad, H Cendl, M Chakraborty, J Cole, E Davies, M Edgar, M Edwards, N Elashhar, S Elliott, J Evans, J Evans, M Evans, E Evans, C Fegan, M Forester, J Forton, S Frayling, S Gage, F Greaves, S Harrhy, M Haynes, A Heavens, H Hill, Z Hilton, L Jones, A Kelly, L Knibbs, N Krishnapalli, D Lau, J May, E McGough, A McQueen, J Milner, M Morgan, F Muhammad, R Norman, K Nyland, C Oliver, K Paradowski, M Patal, B Phillips, M Potdar, K Rahilly, D Rigler, C Robinson, S Scourfield, A Semmens, M Starr, J Stevens, S Struik, E Thomas, R Thomas-Turner, G Thueux, G Williams, J Williams, M Williams, S Zaher.

**Hull University Teaching Hospitals NHS Trust** N Easom (PI), K Adams, K Ahmed, D Allsup, G Barlow, R Barton, J Batac, D Clark, L Cullen, K Drury, A Frygier, S Gribben, P Gunasekera, A Harvey, M Hayes, M Ivan, S Khan, M Kolodziej, P Lillie, V Lowthorpe, V Mathew, S Mongolu, A Morrow, I Muazzam, D Norton, K Nu, R Patmore, T Perinpanathan, C Philbey, B Pickwell-Smith, D Purchase, S Rastogi, A Richards, L Rollins, A Samson, T Sathyapalan, Y See (Associate PI), L Sherris, K Sivakumar, D Smith, T Taynton (Associate PI), G Walker, A Wolstencroft, H Yates.

**Mid Cheshire Hospitals NHS Foundation Trust** J Majumdar (PI), T Adeyemo, K Best, G Bridgwood, R Broadhurst, C Brockelsby, T Brockley, J Brown, R Bujazia, A Burton, S Clarke, J Cremona, C Dixon, S Dowson, H Drogan, F Duncan, M Emms, H Farooq, D Fullerton, N G, C Gabriel, S Hammersley, R Hum, T Jones, S Kay, E Kelly, M Kidd, D Lees, R Lowsby, D Maren, D Maseda, E Matovu, K McIntyre, H Moulton, K Nourein, K Pagett, A Ritchings, S Smith, J Taylor, K Thomas, K Turbitt, M Williams, S Yasmin.

**Royal Free London NHS Foundation Trust** T Mahungu (PI), H Tahir (Co-PI), A Abdul, R Abdul-Kadir, H Aboelela, M Al-Khalil, N Allan, I Alshaer, M Anderson, M Araujo, G Badhan, A Bakhai, S Bhagani, B Bobie, A Brraka, B Caplin, A Carroll, A Carroll, H Century, E Cheung, D Cohen (Associate PI), O Coker, D Collier, V Conteh, N Cooper, J Crause, N Davies, R Davies, V Deelchand, M Dosani, L Ehiorobo, C Ellis, G Ferenando, J Franklin, P Gardiner, F Geele, J Gosai, N Handzewniak, E Hanison, S Hanson, N Holdhof, H Hughes, C Jack, C Jarvis, V Jennings, H Koo, V Krishnamurthy, A Kurani (Associate PI), Z Ladan, L Lamb, A Lang, V Le, S Lee, S Lo, A Luintel, A Maharajh, H Mahdi, T Majekdunmi, D Matila, S Melander, F Mellor, A Molloy, R Moores, J Morales, G Moray, A Nandani, S Nasa, S O'Farrell, A Oomatia, A Osadcow, J Osei-Bobie, G Pakou, P Patel, C Patterson (Associate PI), E Pyart, E Quek, S Rabinowicz, T Rampling, R Rankhelawon, A Rodger, A Scobie, S Sharma, C Singh, S Sithiravel, T Sobande, P Talbot, P Taribagil, S Veerasamy, G Wallis, J Whiteley, E Witele, A Wong, E Woodford, N Yaqoob.

**Sheffield Teaching Hospitals NHS Foundation Trust** P Collini (PI), A Ali, A Alli, S Anderson, A Ang, I Aslam, A Basran, J Belcher, G Benoy, S Bird, M Boothroyd, H Bowler, A Card, K Cawthorne, L Chapman, K Chin, A Clarke, S Clenton, V Clubb, L Cockayne, D Cohen, J Cole, H Colton, A Condliffe, R Condliffe, A Creaser-Myers, M Cribb, S Curran, S Darby, T Darton, D de Fonseka, T de Silva, A Dunn, C Durojaiye, P Easton, S Eggleston, A Ellwood, A Emery, E Ferriman, H Foot, A Ford, R Foster, Z Gabriel, V Goodall, J Greig, N Guthrine, J Hall, H Hanratty, S Hardman, K Harrington, J Hau, E Headon, C Holden, B Holroyd-Hind, K Housely, A Howell, L Hunt, E Hurditch, J Hurdman, Z Hussein, F Ilyas, J Jackman, Y Jackson, C Jarman, F Kibutu, B Kilner, T Kitching, S Lassa, R Lawson, C Lee, R Lenagh, L Lewis, N Lewis, T Locke, H Luke, A Lye, I Macharia, L Mair, G Margabanthu, P May, A Mbuyisha, J McNeill, S Megson, J Meirill, J Meiring, J Middle, J Middleton, L Milner, P Morris, D Mosby, A Nazir, H Newell, T Newman, L Nwafor, V Palissery, V Parkash, L Passby, R Payne, I Pernicova, P Phillips, M Plowright, A Raithatha, G Rana, P Ravencroft, S Renshaw, A Rothman, M Roycroft, D Sammut, S Sammut, R Sellars, S Sherwin, J Sidebottom, P Simpson, L Smart, M Sterrenburg, B Stone, M Surtees, A Telfer, B Thamu, R Thompson, Y Thompson, N Tinker, H Trower, K Turner, M Ul Haq, N Vethanayagam, P Wade, J Wadsley, S Walker, J G R Watson, J Wesonga, R West, R Whitham, T Williams, J Willson, A Wilson, I Wilson, T Zak, E Zincone, STH CRF COVID Nursing team.

**University Hospitals Birmingham NHS Foundation Trust** C Green (PI), T Whitehouse (Co-PI), I Ahmed, N Anderson, C Armstrong, A Bamford, H Bancroft, M Bates, M Bellamy, T Bellamy, C Bergin, K Bhandal, E Butler, M Carmody, N Cianci (Associate PI), K Clay (Associate PI), L Cooper, J Daglish, J Dasgin, A Desai, S Dhani, D Dosanjh, E Forster, J Gresty, E Grobovaite, N Haider, B Hopkins, D Hull, Y Hussain, A Kailey, M Lacson, M Lovell, D Lynch, C McGhee, C McNeill, F Moore, A Nilsson, J Nunnick, W Osborne (Associate PI), S Page, D Parekh, C Prest, K Price, V Price, M Sangombe, H Smith, I Storey, L Thrasyvoulou, K Tsakiridou, D Walsh, S Welch, H Willis, L Wood, J Woodford, G Wooldridge, C Zullo.

**Epsom and St Helier University Hospitals NHS Trust** S Winn (PI), J Evans (Co-PI), F Abbas, S Acheampong, S Ahamed Sadiq, A Aldana, B Al-Hakim, A Alipustain, P Allen, G Azzopardi, E Balogh, Z Baxter, C Bond, C Cheeld, R Chicano, I Chukwulobelu, N Colbeck, R Dogra, E Doherty, A Elradi, J Emberton, L Evans, R Ganapathy, N Hamilton, J Handford, M Haque, J Hayes, R Hayre, L Ilves, C Jagadish, S Jain, K Jian, A Johnson, L Johnson, B Kim, J Kotecha, A Kundu, D Langer, E Lester, A Lunia, Y Mashhoudi, K Mathias, T Medveczky, F

Mellor, G Mic, S Moore, M Morgan, O Mushtaq, S Nafees, A Oluwole-Ojo, V Palagiri Sai, M Phanish, H Rahimi, S Ramanna, A Ramnarine, J Ratoff, M Ridha, S Rozewicz, T Samuel, G Saxena, S Selvendran, B Shah, S Shahnazari, R Shail, A Sharif, S Singh, S Somalanka, R Suckling, P Swift, L Tarrant, V Tyagi, N Vilimiene, P Virdee, R Wake, L Walsh.

**Portsmouth Hospitals NHS Trust** T Brown (PI), J Andrews, R Baker, M Baker-Moffatt, A Bamgboye, D Barnes, S Baryschpolec, V Bataduwaarachchi, S Begum, L Bell, H Blackman, J Borbone, N Borman, P Braga Sardo, L Brimfield, M Broadway, F Brogan, R Bungue-Tuble, K Burrows, A Chauhan, E Cowan, M Czekaj, Z Daly, A Darbyshire, A Das, M Davey, M David, J Denham, R Duhoky, C Edwards, S Elliott, H Evison, L Fox, Z Garner, I Gedge, B Giles, S Glaysher, S Gosling, A Gribbin, J Hale, Y Harrington-Davies, L Hawes, J Hayward, A Hicks, A Holmes, S Howe, H Htet, B Jones, C Lameirinhas, E Lavington, B Longhurst, L Lopes, M Mamman, J Marshall, S McCready, L Milne, C Minnis, M Moon, J Mouland, L Murray, J Ouyang, C Pereira Dias Alves, M Rason, L Richmond, D Rodgers, S Rose, M Rowley, M Rutgers, T Scorrer, L Shayler, R Sievers, K Siggens, S Smith, C Stemp, N Szarazova, E Taylor, R Thornton, A Tiller, C Turner, L Vinall, E Walmsley, M Wands, J Warren, L Watkins, M White, L Wiffen (Associate PI), J Winter, C Wong, K Wren, Y Yang.

**South Tees Hospitals NHS Foundation Trust** D Chadwick (PI), A Aboagye-Odei, S Armstrong, D Athorne, A Awadelkareem, M Branch, J Brolly, S Brown, J Cheaveau, H Chen, Y Chua, N Cunningham, M Dafalla, S Davies, J Dodds, S Dorgan, D Dunn, P Dunn, M Elsayed, E Hammond, P Harper, H Harwood, K Hebbron, F Hunt, A Kala Bhushan, P Lambert, C Lawrence, D Leaning, T Linn, T Manders, B McCarron, N Miller-Biot, C Milne, W Mohammad, M Mollet, J Mulcahy, A Murad, S Ooi, M Owston, J Potts, C Proctor, S Puliyakkadi, B Puvaneswaran, S Rao, R Raw (Associate PI), M Seelarbokus, P Singh, V Srirathan, L Swithenbank, A Szekeres, L Thompson, H Wardy, L Wiblin, J Widdrington, J Williams, P Winder, C Wroe.

**University College London Hospitals NHS Foundation Trust** H Esmail (PI), R Heyderman (Co-PI), V Johnston (Co-PI), D Moore (Co-PI), A Andrews, F Beynon, P Bodalia, I Bokobza, X Chan, Z Chaudhry, C Chung, J Cohen, D Crilly, S Eisen, N Fard, J Gahir, L Germain, J Glanville, R Gupta, N Lack, A Last, A Luintel, M Merida Morillas, B Nadjm, N Platt, T Rampling, H Rickman, S Rokadiya, S Roy, A Samson, R Shortman, I Skorupinska, M Skorupinska, J Spillane, C Thakker, L Wellings.

**NHS Fife** D Dhasmana (PI), F Adam, K Aniruddhan, J Boyd, N Bulteel, P Cochrane, I Fairbairn, S Finch, K Gray, L Hogg, S Iwanikiw, M Macmahon, P Marks, A McGregor, A Morrow, J Penman, J Pickles, J Ramsay, A Scullion, H Sheridan, M Simpson, D Sloan, C Stewart, J Tait, A Timmins, M Topping.

**West Suffolk NHS Foundation Trust** M Moody (PI), S Barkha, H Cockerill, K Durrant, J Godden, J Kellett, T Murray, A Saraswatula, A Williams, L Wood.

**Medway NHS Foundation Trust** R Sarkar (PI), K Abernethy (Associate PI), C Adams, L Adams, A Addo, F Aliyuda, S Archer, A Arya, E Attubato, F Babatunde, M Bachour, P Balasingam, A Bhandari, F Brokke, R Chauhan, V Chawla, R Chineka, A Davis, N Edmond, M Elbeshy, C Ezenduka, S Ferron, C Gnanalingam, D Gotham, M Hollands, M Iqbal, A Jamal, B Josiah, S Kidney, M Kim, K Koukou, T Kyere-Diabour, L Leach, A Liao, A Maheswaran, M Mansour, N Miah, J Morilla, L Naglik, K Naicker, Z Nurgat, S Rai, I Redknap, Z Rehman, A Ryan, Y Samuel, A Shaibu, P Soor, R Squires, W Stagg, W Ul Hassan, P Vankayalapati, E Vyras, A Williams, J Wood, N Zuhra (Associate PI).

**Cambridge University Hospitals NHS Foundation Trust** M Knolle (PI), E Gkrania-Klotsas (Co-PI), O Abani, R Bousfield (Associate PI), T Dymond, L Eadie, K Gajewska-Knapik, J

Galloway, R Gore, K Leonard, T Mikolasch, H-P Mok, N Pathan, P Polgarova, J Sahota, K Sharrocks, R Swain.

**Stockport NHS Foundation Trust** R Stanciu (PI), M Afridi, M Al Dakhola, M Alafifi, S Bennett, L Brown, C Cooper, A Davison, D Eleanor, J Farthing, A Ferrera, S Gandhi, L Gomez, Z Hassan, P Haywood, C Heal, H Jackson, J Johnston, A Lloyd, S McCaughey, R Mills, R Owen, A Pemberton, F Rahim, H Robinson, N Sadiq, R Samlal, V Subramanian, D Suresh, D Tudor, H Wieringa, I Wright, B Xia.

**Nottingham University Hospitals NHS Trust** W S Lim (PI), M Ali, L Anderson, A Andrews, S Ashraf, D Ashton, G Babington, G Bartlett, D Batra, L Bendall, N Benetti, T Brear, A Buck, G Bugg, J Butler, R Cammack, J Cantliff, L Clark, E Connor, P Davies, M Dent, C Dobson, A Fatemi, M Fatemi, L Fleming, J Grundy, J Hallas, L Hodgen, S Hodgkinson, S Hodgson, L Howard, C Hutchinson, B Jackson, J Kaur, E Keddie-Gray, E Kendall, C Khurana, M Langley, L Lawless, L Looby, M Meredith, L Morris, H Navarra, R Nicol, J Oliver, C Peters, B Petrova, R Purdy, Z Rose, L Ryan, J Sampson, G Squires, J Squires, R Taylor, A Thomas, J Thornton, K Topham, O Vincent, S Warburton, S Wardle, H Waterfall, S Wei, T Wildsmith, L Wilson.

**Dorset County Hospital NHS Foundation Trust** J Chambers (PI), F Best, J Birch, L Bough, S Caddy, A Cave, J Chambers, J Colton, J Graves, S Horton, J Rees, R Thomas, W Verling, P Williams, S Williams, B Winter-Goodwin, S Wiseman, D Wixted.

**East Suffolk and North Essex NHS Foundation Trust** J Douse (PI), M Ramali (Co-PI), K Ahmed, S Alam, A Arumaithurai, B Atraskiewicz, J Bailey, I Balluz, D Beeby, S Bell, J Bloomfield, S Blows, N Broughton, C Buckman, M Burton, C Calver, J Campbell, P Carroll, C Chabo, R Chalmers, K Cheung, M Chowdhury, G Christoforou, K Cooke, N Deole, T H Dinh (Associate PI), C Driscoll, J Dulay, S Finbow, I Floodgate, R Francis, C Galloway, E Galloway, M Garfield, A Ghosh, G Gray, P Greenfield, A Gribble, M Gunawardena, M Hadjiandreou, H Hewer, M Hossain, R Howard-Griffin, K Howlett, C Huah, N Innes, V Inpadhas, A Islam, L James, Z Jiao, K Johannessen, J Kathirgamachelvam, M Khan Tharin, V Kushakovsky, S Lee, R Lewis, R Lloyd, LH Lui, L Mabelin, P Mallett, D Morris, S Nallapareddy, S Nishat, H C Ooi (Associate PI), R Osagie, H Patel, AK Phyo, M Pretorius, B Purewal, F Ramali, H Rawlins, P Ridley, V Rivers, J Rosier, E Rushforth, S Sethi, A Sharma, S Sharma, A Sheik, J Shoote, M Shuvo, R Skelly, R Smith, R Smith, R Sreenivasan, P Tovey, A Turner, K Turner, K Vithian, I Weichert, W Win.

**Wye Valley NHS Trust** I DuRand (PI), J Al-Fori, A Almahroos, J Annett, A Barclay, J Bartlett, J Birch, N Bray, A Carrasco, M Cohn, E Collins, K Crowley, A Davies, M Forkan, A Fratila, H Gashau, S Gayle, K Hammerton, A Hassan, A Hedges, R Homewood, Z Khan, S Maryosh, S Meyrick, L Moseley, B Mwale, L Myslivecek, A Phillips, J Porteous, M Qayum, P Ryan, A Salam, C Seagrave, F Suliman, A Talbot -Smith, S Turner, S Vaughan, E Wales, H Walker, J Woolley.

**Blackpool Teaching Hospitals NHS Foundation Trust** G Hardy (PI), A Abdullah (Associate PI), N Ahmed, O Assaf, A Barnett, A Beasley, L Benham, D Bennett, P Bradley, Z Bradshaw, M Brunton, T Capstick, M Caswell, V Cunliffe, J Cupitt, R Downes, L Elawamy, J Howard, C Jeffs, N John, N Latt, J Mason, R McDonald, A Mulla, E Mutema, J Navin, A Parker, A Potter, S Preston, N Slawson, E Stoddard, S Traynor, V Vasudevan, E Ward, S Warden, A Wignall, J Wilson, A Zmierczak.

**Royal Cornwall Hospitals NHS Trust** D Browne (PI), Z Berry, H Chenoweth, A Collinson, F Hammonds, C James, L Jones, E Laity, K Morgan, C Murphy, T Nisbett, R Sargent, L Trethowan, K Watkins, L Welch.

**Wirral University Teaching Hospital NHS Foundation Trust** A Wight (PI), F Adeoye, L Bailey, E Barker, S Bokhari, S Brownlee, A Bull, J Corless, C Denmade, N Ellard, A Farrell, A Hufton, R Jacob, A Jones, K Jones, H Kerss, J McEntee, N Morris, R Myagerimath, T Newcombe, M Parsonage, H Peake, D Pearson, R Penfold, S Rath, A Reddington, R Saunders, A Sharp, B Spencer, A Suliman, S Sutton, H Tan, D Tarpey, L Thompson, T Thornton, E Twohey, D Wagstaff, Z Wahbi, G Wardere, S Williams.

**Royal Berkshire NHS Foundation Trust** M Frise (PI), R Arimoto, J Armistead, A Aslam, A Barrett, S Bartley, P Bhuie, K Bostock, A Burman, C Camm, R Carson, H Coles, J D'Costa, A Donohoe, E Duffield, F Emond, S Everden, E Gabbitas, E Garden, N Gould, S Gurung Rai, S Hadfield, A Hayat, S Haysom, J Hilton, N Jacques, L Keating, C Knowles, H Lawrence, K Lennon, A McGown, B Mitchell, L Mokogwu, T Okeke, G Parsons, S Rai, L Sathyanarayanan, F Selby, M Thakker, S Vettikumaran, A Walden.

**South Tyneside and Sunderland NHS Foundation Trust** H Grover (PI) (Associate PI), A MacNair (Co-PI), C Brown, A Burns, C Caroline, M Chopra, R Davidson, M Dickson, J Doughty, N Elkaram, I Emmerson, L Fairlie, L Fuller, M Hashimm, J Henderson, K Hinshaw, J Holden, R Hovvels, S Laybourne, K Martin, M McKee, J McKenna, J Moore, N Mullen, P Murphy, L Palmer, G Parish, M Rangar, M Richardson, A Rostron, A Smith, L Smith, L Terry, A Trotter, F Wakinshaw, E Walton, M Walton.

**University Hospitals Of Morecambe Bay NHS Foundation Trust** S Bari (PI), S Achieng, M Al-Hayali, M Al-Jibury, K Allison, V Anu, C Bartlett, L Bartlett, M S Bhuiyan, S Bhuiyan, L Bishop, M Bukhari, K Burns, J Craig, A Davies, A Dent, J Dodd, A Dow, M El-Naggar, A Fielding, M Glover, M Gorst, J Han, C Hay, A Higham, O Igwe, I Irabor, S Kaprapina, J Keating, H Khalid, T Khan, R Kubaisi, L Lobosco, M Lwin, F Mahmood, P Mallinder, B Marwan, L McDougall, S Moorby, H Morris, K Ngwenya, O Onuoha, Z Patel, S Peters, D Power, J Ritchie, P Sarmiento, A Shams, S Sharma, K Simpson, H Spickett, C Stokes, M Tai, H Thatcher, C Till, A Varghese, H Williams, M Williams, T Win, F Wood, S Woodmansey.

**NHS Lothian: Western General Hospital** O Koch (PI), A Abu-Arafeh, E Allen, L Bagshaw, C Balmforth, R Barnes, A Barnett-Vanes, R Baruah, S Begg, S Blackley, M Braithwaite, G Clark, S Clifford, D Dockrell, M Evans, V Fancois, C Ferguson, S Ferguson, N Freeman, E Gaughan, E Godden, S Hainey, R Harrison, B Hastings, S Htwe, A Ju Wen Kwek, M Ke, O Lloyd, C Mackintosh, A MacRaild, W Mahmood, E Mahony, J McCrae, S Morris, C Mutch, S Nelson, K Nunn, D O Shea, I Page, M Perry, J Rhodes, N Rodgers, J Schafers (Associate PI), A Shepherd, G Soothill, S Stock, R Sutherland, A Tasiou, A Tufail, D Waters, R Weerakoon, T Wilkinson, R Woodfield (Associate PI), J Wubetu.

**Royal Devon and Exeter NHS Foundation Trust** R Sheridan (PI), M Masoli (Co-PI), H Bakere, A Bowring, T Burden, A Corr, P Czylok, L Dobson, A Forrest, E Goodwin, H Gower, A Hall, S Heddon, G Joseph, L Knowles, H Mabb, A Mackey, V Mariano, E Matkins, E McEvoy, L Mckie, P Mitchelmore, L Morgan, T Nightingale, R Oram, C Oreilly, N Osborne, H Palfrey, S Patten, J Pearce (Associate PI), I Seaton, A Smallridge, P Smith, M Steward (Associate PI), D Sykes, J Tipper, S Todd, C Webb, S Whiteley, S Wilkins, N Withers, K Zaki, L Zitter.

**Countess Of Chester Hospital NHS Foundation Trust** S Scott (PI), A Abdelaziz, A Abdulakeem, M Abouibrahim, A Adeyanju, A Aditya, M Ahmed, S Ahmed, S H Ahmed, A Ajibode, L Alomari, F Ashworth, A Asrar, E Austin, A Awan, P Bamford, A Banerjee, A Barclay, L Barker, K Barker-Williams, T Barnes, W Barnsley, H Batty, F Bawa, G Beech, J Bellamu, K Bennion, I Benton, J Bhanich Supapol, S Billingham, K Blythe, T Bowker, S Brearey, S Brigham, V Brooker, A Brown, C Burchett, M Burgess, R Cade, F Cameron, R Cannan, M Cardwell, K Cawley, N Chavasse, L Cheater, Z Cheng, K Clark, R Clarke, R Clifford, E Cole, C Cook, C Cotton, S Cremona, D Curran, H Dallow, J Dangerfield, A Davidson, A Davies, A Davies, K Davis, M Devonport, Y Doi, E Doke, C Dragos, H Duffy, L Eagles, M Edwards, L

Ellerton, R Ellis, H Elmasry, E Entwistle, A Fallow, M Faulkner, E Ferrelly, K Flewitt, C Foden, D Font, L Fuller, R Gale, L Gamble, M Gardiner, K Gillespie, L Goodfellow, M Grant, S Griffiths, Y Griffiths, J Grounds, V Guratsky, L Hartley, P Hayle, C Haylett, B Hemmings, N Henry, P Hettiarachchi, L Hill, H Hodgkins, S Holt, S Hunt, M Ibraheim, M Irshad, M Iyer, Z Jamal, R James, A Jayachandran, R Jayaram, H Jeffrey, A Johari, O Johnson, C Jones, C Jones, E Jones, R Jones, A Kazeem, N Kearsley, K Kouranloo, A Kumar, S Leason, N Lightfoot, B Lim, D Llanera, R Llewellyn, S Loh, E London, C Lonsdale, T Maheswaran, G Markham, E Martin, P Maskell, A Mather, A Mathew, I Mazen, M McBuigan, J McBurney, M McCarthy, R McEwen, M McGuigan, E Meeks, G Metcalf-Cuenca, L Michalca-Mason, S Middleton, L Mihalca-Mason, D Mock Font, A Moore, L Morris, R Mufti, K Murray, I Nagra, A Nallasivan, M Nayyar, F Naz, R Nelson, R Nicholas, C Oakley, E Okpo (Associate PI), F Olonipile, C O'Neill, C Ord, J Osman, E Owens, S Pajak, S Pal, J Parker, H Parry, S Pearson, K Penney, E Perritt, D Phillips, C Pickering, A Ponnuswamy, S Powell, V Prescott, J Prince, E Pye, S Rahman, P Rath, S Read, J Reed, A Reid (Associate PI), J Roberts, N Robin, R Robinson, M Saladi, D Scholes, A Scott, S Scott, H Shabbir, M Shah, G Sheppard, F Shiham, T Spencer, J Spriggs, C Steele, Z Syrimi, L Theocharidou, M Thevendra, C Thorne, A Timmis, S Tomlin, T Trussell, O Vass, B Vlies, A Walters, L Walton, K Webb, T Webster, S Wheaver, D Whitmore, A Williams, C Wyatt, W Yap, J Ye, C Yeung, E Young, L Zammit, E Zin.

**Great Western Hospitals NHS Foundation Trust** A Kerry (PI), A Aldesouki, A Azeem, V Barlow, A Beale, T Benn, S Bhandari, A Brooks, C Browne, J Butler (Associate PI), J Callaghan, B Chandrasekaran, N Clark, L Davies, R Davies, T Elias, J Evans, D Finch, S Flockhart, P Foley, E Fowler, E Fraile, G Gowda, J Gregory, C Hunt, A Ipe, A Jaffery, M Juniper, S Khan, I Laing-Faiers, H Langton, G Laura, C Lewis-Clarke, J Lodge, C Mackinlay, P Mappa, A Maxwell, L McCafferty, W Mears, E Mousley, T Novak, C Novis, L Pannell, S Peglar, A Pereira, I Ponte Bettencourt dos Reis, E Price, A Quayle, Q Qurratulain, M Ryder, S Small, H Smith, C Strait, E Stratton, M Tinkler, J Ugoji, A van der Meer, L von Oven, A Waldron, R Waller, M Watters, S White, L Whittam, T Wiliams, Z Xia, K Yein, V Zinyemba, G Zubikarai.

**Salisbury NHS Foundation Trust** M Sinha (PI), K Ames, A Anthony, J Barr, A Barton, L Bell, G Chaplin, M Cook, S Diment, P Donnison, B Eapen, S Evans, R Fennelly, S Gray, H Harcourt, A Hawkins, I Jenkins, M Johns, T Jones, L Joy, V King, I Leadbitter, W Matimba-Mupaya, L Mattocks, R Mehta, H Morgan, A Rand, S Salisbury, K Seymour, S Strong-Sheldrake, C Thompson, F Trim, E Underhill.

**The Newcastle Upon Tyne Hospitals NHS Foundation Trust** E Hunter (PI), R Agbeko, A Bailey, K Baker, A Barr, J Brolly, E Cameron, Q Campbell Hewson, R Capstick, J Cheaveau, I Clement, A De Soyza, C Duncan, M Emonts, A Fenn, S Francis, L Gardner, B Ghavami Kia, J Glover Bengtsson, A Greenhalgh, A Hanrath, H Hanson, C Hays, K Houghton, D Jerry, G Jones, S Kelly, A Kimber, N Lane, J Macfarlane, P McAlinden, I McCullagh, S McDonald, O Mohammed, P Nwajiugo, R Obukofe, J Parker, B Patel, A Patience, B Payne, R Percival, D Price, Z Razvi, N Rice, S Robson, A Sanchez Gonzalez, B Shillitoe, A Stanton, E Stephenson, N Trewick, S Tucker, G Waring, R Welton, S West, E Williams, E Wong, F Yelnoorkar.

**University Hospitals Of Derby and Burton NHS Foundation Trust** T Bewick (PI), P Daniel (Co-PI), U Nanda (Co-PI), A Anada, P Basvi, G Bell, H Cox, C Downes, K English, A Fletcher, J Hampson, S Hathaway-Lees, M Hayman, N Jackson, A Matthews, C McDonald, S Ojha, L Prince, J Radford, K Riches, G Robinson, A Sathyanarayanan, C Smith, E Starkey, J Usher, L Wilcox, L Wright, M Zawadzka.

**University Hospitals Of North Midlands NHS Trust** T Kemp (PI), J Alexander, W Al-Shamkhani, L Bailey, N Bandla, O Bani-Saad, A Bland, N Bodasing, S Brammer, A Cadwgan, M Chaudhary, L Cheng, M Chikungwa, S Church, F Clark, N Coleman, M Davies, L Diwakar, A Farmer, F Farook, M Ganaie, U Garcia, M Gellamucho, K Glover, M Haris, J Humphries, I

Hussain, J Ibrahim, N Idrees (Associate PI), S Khan, L Korcierz, S Krueper, J Lee, S Lord, J Marshall, I Massey, H McCreedy, A McGowan, G Muddegowda, I Mustapha, N Mustfa, K Nettleton, Z Noori, W Osman, H Parker, N Patel, I Ponce, A Quinn, M Ram, A Remegoso, E Sadler, L Scott, T Scott, E Sernicola, N Sheikh, R Shorrocks, R Swift, C Thompson, J Tomlinson, K Tomlinson, R Varquez, L Verueco, L Walker, J Weeratunga, C White, J White, P Wu.

**University Hospitals Plymouth NHS Trust** D Lewis (PI), D Affleck, O Anichtchik, K Bennett, M Chopra, J Corcoran, M Cramp, H Davies, J Day, M Dobranszky Oroian, E Freeman, C Gordon, L Jose, L Madziva, R Mansour, G Marsh, E May, H McDill, P Moodley (Associate PI), C Morton, M Mwadeyi, H Newman, H Notman, C Orr, A Patrick, L Pritchard, G Selby, J Shawe, H Tan, N Thiri Phoo (Associate PI), K Whitehorn.

**Buckinghamshire Healthcare NHS Trust** N Wong (PI), J Abrams, A Alkhudhayri, N Aung, A Baldwin, O Bannister, J Barker, H Beddall, H Blamey, E Chan, J Chaplin, B Chisnall, C Cleaver, M Corredera, S Crotty, H Cui, B Davies, P Dey, L Downs, S Gettings, B Hammans, S Jackman, P Jenkins, M Kononen, S Kudsk-Iversen, A Kudzinskas, M Laurenson, R Mancinelli, J Mandeville, K Manso, B Marks, S McLure, O Michalec, E Morgan-Smith, A Ngumo, H Noe (Associate PI), R Oxlade, A Parekh (Associate PI), V Pradhan, M Rahman, C Robertson, R Rule, S Shah, H Smith, J Tebbutt, N Vella, M Veres, A Watson, R West, L Western, M Zammit-Mangion, M Zia.

**Sherwood Forest Hospitals NHS Foundation Trust** M Roberts (PI), L Allsop, K Amsha, K Bennett, T Brear, P Buckley, G Cox, N Downer, L Dunn, M Flynn, M Gill, C Goodwin, C Heeley, D Hodgson, L Holloway, M Holmes, R Holmes, J Hutchinson, R Johnson, S Kalsoom, J Kirk, E Langthorne, W Lovegrove, G Mabeza, A Molyneux, C Moulds, D Nash, Z Noor, J Rajeswary, T Sewell, S Shelton, H Shirt, K Slack, S Smith, N Thorpe, S Turner, B Valeria, I Wynter, M Yanney, I Ynter.

**Bolton NHS Foundation Trust** M Balasubramaniam (PI), C Acton, S Ahmad, R Ahmed, A Ajmi, A Al-Asadi, S Altaf, A Amin, A Bajandouh, R Carey, Z Carrington, J Chadwick, S Cocks, C Dawe, S Farzana, O Froud, A Gibson, A Green, P Hill, A Hindle, R Holmes, G Hughes, R Hull, M Ijaz, R Kalayi, M Khan, A Koirata, S Latham, G Lipscomb, K Lipscomb, A McCorkindale, M McNulty, O Meakin, N Meghani, N Natarajan, D Nethercott, P Nicholas, T Pandya, A Parkinson, V Priyash, L Pugh, J Rafique, J Robertson, M Saleh, W Schneblen, B Sharma, O Sharma, D Shaw, Z Shehata, J Shurmer (Associate PI), R Sime, S Singh, R Smith, C Subudhi, R Tallent, E Tanton, K Teasdale, D Tewkesbury, P Thet, S Thornton, J Timerick, C Underwood, N Wang, M Watts, I Webster, B Wilson.

**King's College Hospital NHS Foundation Trust** J Galloway (PI), M McPhail (Co-PI), J Adeyemi, J Aeron-Thomas, M Aissa, E Alveyn, A Buazon , T Buttle, S Candido, E Canonizado, A Cavazza , M Cesay, K Clark, M Cockerell , A Comerford, M Depante, G Dimitriadis, C Donaldson, K El-Bouzidi, S Finch, C Finney, K Garrero, G Godwin, S Gogoi, N Griffiths, A Gupta, L Hall, J Hannah, Y Hu, E Jerome (Associate PI), N Jones, N Kametas, M Kumar, L Linkson, N Long, M Magtalas, E Makanju, H Martin, D Nagra, H Noble, K O’Reilly, V Patel, T Pirani, A Posada, N Powell, J Prince, D Rao, S Ratcliff, S Saha, B Sari, N Sikondari, J Smith, B Solis, A Te, C Tey, M Tran, R Uddin, A Varouxaki, J Vidler (Associate PI), J Ward, M West, C Williamson, M Yates, A Zamalloa.

**Maidstone and Tunbridge Wells NHS Trust** A Ross-Parker (PI), A Abbott, L Adams, S Anandappa, B Babiker, M Barbosa, G Chamberlain, K Cox, D Datta, M Davey, N Earl, F Gallam, R Gowda, S Goyal, A Gupta, E Harlock, C Hart, E Hughes, S Husaini, E Hutchinson, A Keough, A Khan, S Kumar, R Nemane, L Orekoya, I Pamphlett, T Patel (Associate PI), C Pegg, P Rajagopalan, A Richards, S Sathianandan, R Seaman, S Siddavaram, O Solademi, M Szeto, P Tsang, V Uppal, A Waller, H Winwood.

**NHS Greater Glasgow and Clyde: Glasgow Royal Infirmary** K Puxty (PI), O Adeagbo, C Aiken, J Alexander, L Bailey, H Bayes, A Begg, M Broom, A Brown, S Carmichael, S Cathcart, F Christie (Associate PI), C Clayton, R Colbert, R Cowan, H Cranston, I Crawford, S Currie, J Cuthill, A Dougherty, K Duffy, A Duncan, C Dunne, T French, A Goodfellow, T Grandison, S Griffiths, P Grose, Z Harzeli, J Hawkins, N Hickey, M Hughes, J Ireland, A Jamison, D Jenkins, J Johnstone, R Kearns, D Kernaghan, K Lake, L Latif, L Littlejohn, J Luveta, L Martin, M McCloskey, C McCue, C McGovern, M McIntyre, A Munro, E Murphy, S Nelson, C Ogilvie, L Paton, H Peddie, G Piper, L Pollock, A Puxty, T Quasim, D Rimmer, K Scott, G Semple, M Shaw, S Speirs, F Steffensen, J Tait, S Thornton, L Turner.

**University Hospital Southampton NHS Foundation Trust** S Fletcher (PI), M Avery, M Bader, A Baltmr, S Bartolmeu-Pires, S Beeby, J Bigg, C Bolger, D Brooks, H Burke, K Cathie, S Chabane, B Clancy, M Coleman, A Collier, A Cook, M Dibas, R Digpal, S N Faust, A Fazleen, M Felongo, S Fletcher, J Forbes, T Francis-Bacon, A Freeman, J Halliday, M Hardwick, E Holliday, M Johnson, A Jones, C E Jones, T Jones, A Koutalopoula, J Law, E Marouzet, C Mc Cague, S Michael, M Nelson, J Nolasco, A Pambouka, S Patel, M Petrova, T Phillips, L Presland, A Procter, S Rahmany, N Rayner, J Rojkova, R Samuel, L Sanga, T Sass, M Shaji, C Silva Moniz, A Tanner, T Thomas, K Tluchowska, A Torokwa, S Triggs, N Tucker, C Watkins, B Welham, S Wellstead, H Wheeler.

**Belfast HSC Trust** D Downey (PI), A Blythe, S Carr, A Cassells, D Comer, D Dawson, K Hayes, R Ingham, L Keith, J Kidney, E Killen, J Leggett, D Linden, N Magee, R Marshall, D McClintock, M McFarland, L McGarvey, C McGettigan, S McGinnity, S McMahon, A Nugent, M Paul, A Redfern-Walsh, J Richardson, L Speirs, M Spence, R Stone, D Tweed, A Usher-Rea, B Wells.

**London North West University Healthcare NHS Trust** A Whittington (PI), B Akinbiyi, R Ali, J Barnacle, J Barrett, Z Cahilog, F Cawa, S Chhabra, S Chita, M Colin, A Dagens, E Davenport, E Dhillon, S Filson, J Goodall, R Gravell, A Gupta-Wright, S Gurram, H Houston, G Hulston (Associate PI), S Isralls, N Khan, C Kukadiya, J Low, K Man, B Nayar, O Ojo, P Papineni, V Parris, M Patel, S Quaid, H Rafferty, A Sturdy (Associate PI), B Tyagi, L Vaccari, N van der Stelt, K Vutipongsatorn, G Wallis, E Watson.

**Northumbria Healthcare NHS Foundation Trust** B Yates (PI), C Ashbrook-Raby, A Aujayeb, L Barton, J Bell, S Bourke, H Campbell, D Charlton, K Connelly, D Cooper, A Dawson, L Dismore, S Dodds, C Edwards, S Fearby, V Ferguson, A Green (Associate PI), N Green, H Grover, E Hall, I Hamoodi, S Haney, P Heslop, P Jones, H Lewis, J Luke, L Mackay, C McBrearty, G McCafferty, I McEleavy, H Mckie, N McLarty, U McNelis, A Melville, J Miller, A Morgan, S Parker, L Patterson, H Peggie, S Pick, H Rank (Associate PI), D Ripley, S Robinson, E Rosby, J Rushmer, H Shah, T Smith, V Smith, D Snell, J Steer, E Sykes, A Syndercombe, C Tanney, L Taylor, J Ward, R Warren, M Weatherhead, R Whittle, L Winder.

**St George's University Hospitals NHS Foundation Trust** T Bicanic (PI), T Harrison (Co-PI), Y Aceampong, A Adebiyi, M Ali, D Baramova, M Caudwell, J De Sousa, P Diwan, S Drysdale, L Hamzah, O Harrison, J Hayat, A Janmohamed, H Ju, A Khalil, A Lisboa, E Marler, M Mencias, J Millard, C Page, J Pang, K Patel, A Perry, A Rana, V Raspa, P Ribeiro, N Said, T Samakomva, A Seward, O Skelton, J Sousa, K Spears, A Sturdy, V Tavoukjian, J Texeira, S Tinashe, O Toffoletti, C Ward.

**University Hospitals Of Leicester NHS Trust** C Brightling (PI), N Brunskill (Co-PI), M Wiselka (Co-PI), S Adenwalla, P Andreou, H Aung, P Bakoulas, S Bandi, S Batham, T Beaver, K Bhandal, M Bourne, L Boyles, M Cannon, A Charalambou, C Cheung, A Christou, R Cotter, S Debbie, S Diver, A Dunphy, O Elneima, J Fawke, J Finch, N Fowkes, C Gardiner-Hill, G Genato, M Graham-Brown, J Hailstone, C Haines, B Hargadon, H Holdsworth, W Ibrahim, L

Ingram, J Jesus Silva, M Joshi, S Kapoor, K Kaul, J Kim, A Kuverji, K Kyriaki, A Lea, T Lee, B Leonard, A Lewszuk, L Lock, K Macconaill, R Major, Y Makkeyah, H McAuley, P McCourt, D Mullasseril Kutten, A Palfreeman, D Pan (Associate PI), E Parker, K Patel, M Patterson, R Phillips, L Plummer, I Qureshi, M Raval, R Russell, H Selvaskandan, S Southin, C Torrance, K Tsilimpari, D Vail, V Wenn, C Wiesender, D Yewatkar, A Yousuf.

**Bedford Hospital NHS Trust** T Chapman (PI), I Nadeem (Co-PI), H Ahmed, A Amjad, N Amjad, A Anthony-Pillai, I Armata, R Arora, A Ashour, D Bagmane, V Bastion, R Bhanot, D Callum, K Chan, P Chrysostomou, C Currie, F De Santana Miranda, B Djeugam, C Donaldson, P Eskander, S Farnworth, F Farrukh, N Fatimah, C Fernando, R Field, K Gelly, L Grosu, A Haddad, M Hannun, T Hashimoto, K Hawes, E Hawley-Jones, M Hayathu, M Hikmat, B Jallow, C Jenkins, U Keke, U Khatana, I Koopmans, T Large, P Lemessy, H Lindsay-Clarke, E Lister, R Lorusso, G Lubimbi, W-O Makinde, A Mian, F Miranda, J Morris, N Nasheed, N Nathaniel, M Negmeldin, K Ng, C Ong, K Pandya, S Peerbhoy, M Penacerrada, J Prieto, Q Quratulain, S Rahama, M Revels, L Salih, J Sarella, Z Shaida, V Simbi, N Simon, B Small, M Smith, N Suresh, W Tan, E Thomas, S Trussell, U Vaghela, A Vaidya, J Valentine, O Walker, F Wang, J Wischhusen, R Wulandari, Y Yanagisawa, L Ylquimiche Melly.

**East Sussex Healthcare NHS Trust** A Marshall (PI), A Abousamra, S Ahmed (Associate PI), S Blankley, H Brooke-Ball, T Christopherson, M Clark, T de Freitas, E de Sausmarez, D Dharmasena, I Doss, A Eggink, A Ekunola, S Ghazal, D Hemsley, J Highgate, A Iakovou, J Jeater, T Kalmus Eliasz, O Kankam, J Khoo, A Lowe, T Mak, S Merritt, Y Mohammed, T Morley, A Newby, S Panthakalam, S Qutab (Associate PI), A Rajagopal, N Roberts, M Simon, K Subba, S Tieger, A Trimmings, R Venn, F Willson, T Win, M Yakubi, A Zubir.

**Hampshire Hospitals NHS Foundation Trust** A Goldsmith (PI), R Partridge (Co-PI), Y Abed El Khaleq, M Adamczyk, M Alvarez Corral, A Arias, E Bevan, J Chan, M Chapman, J Conyngham, E Cox, E Defever, A Edwards, C Fitton, D Griffin, A Heath, M Jordan, B King, E Levell, X Liu, J Martin, N Muchenje, M Mupudzi, A Nejad, A Prabhu, S Senra, F Stourton, R Thomas, B White (Associate PI), G Whitlingum, L Winckworth, C Wrey Brown, S Zagalo.

**NHS Lanarkshire: University Hospital Hairmyres** M Patel (PI), T Baird, D Bell, R Boyle, F Burton, L Clark, K Douglas, L Glass, R Hamill, L Jamieson, E Lee, L Lennon, S Marshall, J McKeane, T McLennan, C Meney, R Tejwani, B Welsh.

**The Royal Bournemouth and Christchurch Hospitals NHS Foundation Trust** M Schuster Bruce (PI), D Baldwin, N Barratt, Z Clark, M Dale, D Griffiths, E Gunter, A Hogan, S Horler, T Joyce, M Keltos, S Kennard, N Lakeman, R Lee (Associate PI), L Mallon, R Miln, M Mirela, N Moore, S Nix, S Orr, S Pitts, L Purandare, L Rogers, J Samways, L Sankaran (Associate PI), E Stride, H Tiller, L Vamplew, L Wallis, A Wilce.

**Whittington Health NHS Trust** C Parmar (PI), S Ahmed, W Ang, S Ashcroft, H Bateman, L Booker, N Brown, D Burrage, M Christy, P Dlouhy, J Flor, R Fromson, G Fung, K Gilbert, F Green, M Horsford, L Howard, N Ivin, I Jenkin, C Jetha, J Kibaru, M Kousteni, N Kulkarni, A Kyei-Mensah, A Lillis, I Lim, S Lock, C Macfadyen, C Mason, P McCormack, J Merritt, S Mindel, H Montgomery, S Myers, F Nur, L Parker, B Pattenden, A Pender, A Prately, N Prevatt, A Reid, S Rudrakumar, J Sabale, S Sharma, P Sharratt, K Simpson, J Southwell, S Taylor, L Veys, J Webster, N Wolff, D Worley, J Yuan, A Zuriaga-Alvaro.

**Kettering General Hospital NHS Foundation Trust** N Siddique (PI), M Abedalla, A Abeer Ul Amna, K Adcock, S Adenwalla, O Adesemoye, R Ahmed Ali, A Ali, S Aransiola, M Ashraf, M Ashraq, S Ashton, H Asogan, A Aung, H Aung, A Baral, F Bawani, S Beyatli, H Britton, S Chowdhury, M Curtis, J Dales, R Deylami, A Elliott, S Gali, A Gkioni, L Hollos, N Hollos, M Hussam El-Din, A Ibrahim, Y Jameel, G Keyte, J Khatri, S Kiran, R Kusangaya, S Little, K Lwin, E Lyka, A Madu, G Margabanthu, M Mustafa, H Naeem, D Ncomanzi, G Nikonovich

(Associate PI), A Nisha James, C Oo, Y Owoseni, N Pandian, D Patel, D Patel, I Petras, M Raceala, D Ramdin, G Ramnarain, S Saunders, A Shaikh, M Shakeel, S Sharma, S Stapley, S Sudershan, P Swift, N Veli, A Virk, T Ward, A Wazir, S White, J Wood, N Zakir.

**Mid Yorkshire Hospitals NHS Trust** B Sloan (PI), J Ashcroft, R Beckitt, P Blaxill, S Bond, S Boot, S Buckley, G Castle, E Clayton, N de Vere, A Dwarakanath, J Ellam, D Gomersall, S Gordon, C Hettiarachchi, C Hutsby, R Kousar´┐¢, K Lindley, A Major, A Metcalfe, S Oddy, A Poole, U Prasad, J Quinn, K Rajalingam, A Rose, L Slater, B Taylor, S Taylor, M Thirumaran.

**NHS Lanarkshire: University Hospital Monklands** M Patel (PI), C McGoldrick (Co-PI), P Anstey, T Baird, C Beith, A Blunsum, D Cairney, A Crothers, S Dundas, F Farquhar, G Fleming, D Garner, L Glass, P Goyal, P Grant, R Hamill, S Howard, T Jones, E Lee, J Lees, L Lennon, C Macrae, S Marshall, A McAlpine, M McFadden, R McGovern, C McInnes, C McKeag, A McKie, M McLaughlin, R Millington, J Neil, N Quail, M Ralston, J Reid, J Robb, M Rodger, S Rundell, T Sammut, M Smith, A Stark, C Sykes, M Taylor, B Welsh, J West, R Williams.

**North Bristol NHS Trust** N Maskell (PI), H Adamali, R Adhikary (Associate PI), M Alvarez, D Arnold, M Attwood, P Bailey, N Bale, G Bardsley, S Barratt, S Bevins, R Bhatnagar, A Bibby, L Bradshaw, A Brown, C Burden, H Cheshire, S Clarke, A Clive, R Cousins, S Dawson, A Dipper, J Dodd, F Easton, K Farmer, L Fox, L Gethin, P Halford, F Hamilton, J Hardy, D Higbee, S Holloway, A Jayebalan, A Jenkins, L Jennings, S Jones, C Kilby, H Lee, M Martin, H McDill, H McNally, S Merritt, A Milne, K Minou, E Morab, A Morley, G Nava, N Novas Duarte, S Patole, A Pereira, E Perry, S Rafiq, M Rigby, N Rippon, V Sandrey, C Sellar, M Slowinska, K Smith, L Solomon, L Stadon, M Tout, J Townley, R Wach, D Warbrick, C Watkins, H Welch, C Woods.

**North Cumbria Integrated Care NHS Foundation Trust** C Graham (PI), A Abdelaziz, P Adair, O Ali, W Armstrong, J Atkinson, G Bell, C Brewer, E Carver, M Clapham, R Cleeton, S Cook, H Craig, J Craig, M Elsebaei, R Graham, J Gregory, S Hanif, R Harper, M Hodson, M Holliday, P Jaques, M Lane, J Masters, E Mawson, A McSkeane, P Mead, R Mutch, G Mynott, A Nelson, M Phipps, U Poultney, K Poulton, S Pritchard, S Shah, J Shawcross, G Simmons, C Smit, S Trous, P Tzavaras, V Vasadi, A Wilson, T Wilson, M Wood, D Zehnder.

**Cwm Taf Morgannwg University LHB** C Lynch (PI), Z Auer, E Davies, B Deacon, S Eccles, A Gaurav, B Gibson, M Hibbert, C Lai, L Margarit, D Nair, S Owen, C Pothecary, J Pugh, L Roche, S Sathe, J Singh, J Smeaton, D Tetla, K-A Wilson, C Woodford.

**Shrewsbury and Telford Hospital NHS Trust** J Moon (PI), R Baldwin-Jones, N Biswas, A Bowes, H Button, E Cale, M Carnahan, E Crawford, E Damm (Associate PI), S Deshpande, D Donaldson, C Fenton, R Heinink, S Hester, Y Hussain, K Ibison, M Ibrahim, S Islam (Associate PI), J Jones, S Jose, A Makan, H Millward, J Moorcroft, N Motherwell, H Moudgil, C Mowatt, J Nixon, S Pajak, S Passey (Associate PI), S Pickstock, L Price, M Rees, E Roddy, N Rowe, N Schunke, K Srinivasan, A Stephens, J Stickley, M Tadros, H Tivenan, G Wood.

**South Eastern HSC Trust** F McElwaine (PI), V Adell, D Alderdice, H Allsop, J Baker, A Campbell, J Courtney, R Eadie, A Eccles, J Elder, J Foreman, G Gamble, P Gillen, S Graham, S Hagan, L Hammond, D Hart, K Henry, R Hewitt, K Jones, S Kelly, A Kerr, D Kinnear, C Loughlin, J MacIntyre, C Madden-McKee, K McCollum, J McFlynn, J McKeever, K McMillen, L Moore, C Mulligan, C O'Gorman, F O'kane, J Patterson, S Regan, S Rowan, A Smith, T Trinick, B Valecka, P Yew, G Young.

**The Royal Wolverhampton NHS Trust** S Gopal (PI), R Barlow, S Bhasin, A Bland, C Chacko, C Cheong, D Churchill, K Davies, S Ganguly, M Green, N Harris, W Hawkins, S Kaur, A

Kumar, A Macduff, S Metherell, S Milgate, R Pearse, E Qureshi (Associate PI), L Radford, J Rogers, I Sayers, A Smallwood, K Vassell, D Warrender, L Wild.

**Western Sussex Hospitals NHS Foundation Trust** L Hodgson (PI), M Margarson (Co-PI), L Albon, A Alcala, K Amin, M Bailey, Y Baird, S Beckley, N Botting, A Butler, V Cannons, C Chandler, A Colino-Acevedo, N Falcone, A Fletcher, S Floyd, L Folkes, H Fox, S Funnell, N Gent, J Gibson, J Gilbert, E Glenday, R Gomez-Marcos, C Gonzalez, M Gorniok, C Goumalatsou, D Green, D Heasman, K Hedges, G Hobden, A Hunter, M Iftikhar, P Jane, R Khan, S Kimber, K King, M King, C Lau, M Linney, L Lipskis, J Margalef, T Martindale, A Mathew, P McGlone, E Meadows, D Melville, S Moore, S Murphy, Q Nguyen, R Njafuh, L Norman, N Numbere, D O Rinn, J Park, M Perera, E Petrovics, D Ravaccia, D Raynard, D Reynish, J Richardson, C Ridley, A Roskilly, T Shafi, T Shaw, S Sinha, J Smith, T Standley, F Stourton, S Sweetman, D Szabó, P Tate, Y Thirlwall, P Thorburn, S Troedson, T Tsawayo, E Vamvakiti, R Venn, N White, J Wileman, E Yates.

**Brighton and Sussex University Hospitals NHS Trust** M Llewelyn (PI), R Ahmed, F Baldwin, E Barbon, H Brown, G Bassett, L Bennett, A Bexley, P Bhat, C Bresges, M Bridgett, H Brown, M Campbell, Z Cipinova, A Crepet, E Cross (Associate PI), S Dhillon, A Elkins, L Evans, S Filipa, M Flowerdew, J Gaylard, R Govindan, D Hansen, A Hassan, Z He, C Hunter, O Jones, S Jujjavarapu, C Laycock, E Mekonnen, J Messenger, D Mullan, J Myerson, J Newman, J Nowak, M Pavitt, C Richardson, R Robinson, V Sellick, D Skinner, M Smith, S Sobowiec Kouman, K Stewart, K Trivedi, D Yusef.

**Chesterfield Royal Hospital NHS Foundation Trust** N Spittle (PI), S Beavis, S Beghini, A Blowers, L Blundell, J Bradder, S Broadhead, K Campbell, C Cart, J Cooke, J Cort, J Cresswell, A Crowder, K Dale, S Davies, A Foo, A Forsyth, C Foster, J Gardner, R Gascoyne, P Hickey, M Hollowday, T Horne, M Kelly-Baxter, L Lawson, J Lee, R Lewis (Associate PI), L Lowe, E Mackay, A Mendelski, N Minskip, L Mitchell, E Moakes, L Morgan, K Moxham, I Ogunjembola, A Padmakumar, L Peters, J Pobjoy (Associate PI), K Pritchard, J Salmon, C Sampson, K Sharp, A Smith, R Smith, L Stevenson, S Vittoria, N Weatherly, E Welch, A Whileman, S Williams, E Wolodimeroff, S Wright.

**Manchester University NHS Foundation Trust** T Felton (PI), S Carley (Co-PI), R Lord (Co-PI), A Ustianowski (Co-PI), M Abbas, A Abdul Rasheed, T Abraham, S Aggarwal, A Ahmed, A Ahmed, S Akili, P Alexander, A Allanson, B Al-Sheklly, D Arora, M Avery, C Avram, A Aya, J Banda, H Banks, M Baptist, M Barrera, E Barrow, R Bazaz, R Behrouzi, M Bennett, V Benson, A Bentley, A Bhadi, A Biju, A Bikov, K Birchall, S Blane, S Bokhari, P Bradley, J Bradley-Potts, J Bright, R Brown, S Burgess, M Butt, G Calisti, C Carey, N Chaudhuri, S Chilcott, C Chmiel, A Chrisopoulou, E Church, R Clark, J Clayton-Smith, R Conway, E Cook, S Crasta, G Cummings-Fosong, S Currie, H Dalgleish, C Davies, K Dean, A Desai, R Dhillon, J Digby, D Dolan, G Donohoe, A Duggan, B Duran, H Durrington, C Eades, R Eatough, S Elyoussfi, F Essa, G Evans, A Fairclough, D Faluyi, S Ferguson, J Fielding, S Fiouni, J Flaherty, G Fogarty, S Fowler, A Fox, C Fox, B George, V George, S Giannopoulou, R Gillott, A Gipson, S Glasgow, T Gorsuch, G Grana, G Gray, A Grayson, G Grey, B Griffin, J Guerin, P Hackney, B Hameed, I Hamid, S Hammond, S Handrean, A Harvey, J Henry, S Hey, L Higgins, L Holt, A Horsley, L Howard, S Hughes, A Hulme, P Hulme, A Hussain, M Hyslop, J Ingham, O Ismail, A Jafar, R Jama, S Jamal, L James, F Jennings, A John, M John, E Johnstone, D Kanabar, N Karunaratne, Z Kausar, J Kayappurathu, R Kelly, A Khan, W Khan, J King, S Knight, E Kolakaluri, C Kosmidis, E Kothandaraman, S Krizak, K Kuriakose, N Kyi, F Lalloo, G Lawrence, G Lindergard, C A Logue (Associate PI), L Macfarlane, A Madden, A Mahaveer, L Manderson, G Margaritopoulos, P Marsden, J Mathews, A Mathioudakis, E McCarthy, J McDermott, B McGrath, P McMaster, H McMullen, C Mendonca, A Metryka, D Micallef, A Mishra, H Mistry, S Mitra, S Moss, A Muazzam, D Mudawi, C Murray, M Naguib, S Naveed, P Ninan, M Nirmalan, R Norton, N Odell, R Osborne, G Padden, A Palacios, A Panes, C Pantin, B Parker, L Peacock, A Peasley, N Phillips, M PI, F Pomery, J Potts, N Power, M

Pursell, A Ramchandani, A Rasheed, S Ratcliffe, M Reilly, C Reynard, E Rice, M Rice, P Riley, P Rivera Ortega, J Rogers, T Rogers, R Santosh, T Scoones, A Scott, K Sellers, N Sen, T Shanahan (Associate PI), A Shawcross, S Shibly, C Shilladay, A Simpson, S Sivanadarajah, N Skehan, C Smith, J Smith, L Smith, J Soren, W Spiller, K Stewart, J Stratton, A Stubbs, A Sukumaran, K Swist-Szulik, B Tallon, C Taylor, R Tereszkowski-Kaminski, S Thomas, S Thorpe, M Tohfa, R Tousis, T Turgut, A Uriel, M Varghese, G Varnier, I Venables, S Vinay, R Wang, L Ward, C Warner, E Watson, D Watterson, L Wentworth, C Whitehead, D Wilcock, J Williams, E Willis, L Woodhead, S Worton, B Xavier.

**NHS Forth Valley: Forth Valley Royal Hospital** J Selwyn (PI), A Baggott, K Bohmova, G Clark, J Donnachie, E Henderson, S Huda, G Jayasekera, I Macpherson, M Maycock, E McCann, L McGenily, N McInnes, J McMinn, D Morrison, A Pearson, L Prentice, C Rafique, D Salutous, M Spears, M Stewart, R Thompson, A Todd, P Turner.

**NHS Greater Glasgow and Clyde: Queen Elizabeth University Hospital** M Sim (PI), K Blyth (Co-PI), L Abel, A Arnott, H Bannister, N Baxter, S Beattie, C Doherty, J Ferguson, K Ferguson, S Finan, L Gilmour, R Hague, S Henderson, P Holland, L Jawaheer, L Kelly, S Kennedy-Hay, A Kidd, H King, M Ledingham, K Longbottom, F Lowe, M Lowe, G McCreath, R McDougall, M McFadden, M McGettrick, N McGlinchey, L McKay, B McLaren, F Mcmeeken, C McParland, T McSorley, J McTaggart, J Millar, B Milligan, S Moore, L O'Donohoe, I Orlikowska, N Parker, K Paterson, T Pettigrew, A Phillips, L Pollock, M Ralston, K Robertson, J Rollo, S Silva, H Stubbs, M Tate, L Walker, F Watson, M Wilson, S Wishart.

**NHS Lanarkshire: University Hospital Wishaw** M Patel (PI), N Ali, K Black, R Boyle, S Clements, J Fleming, A Fyfe, L Glass, R Hamill, L Hamilton, C Hughes, E Jarvie, L Lennon, A Lynas, C Macdonald, S Marshall, M Maycock, C McEwan, E McGarry, K Moar, N Moody (Associate PI), S Naidoo, Z Puyrigaud, A Smith, C Stewart, B Welsh, J West, P Wu.

**Swansea Bay University Local Health Board** B Healy (PI), M Baker, S Bareford, I Blyth, A Bone, E Brinkworth, R Chudleigh, A Cook, C Davies, Y Ellis, D Evans, E Evans, S Georges, S Green, R Harford, A Holborow, C Johnston, P Jones, M Krishnan, N Leopold, F Morris, A Mughal, C Murphy, L O'Connell, T Rees, S Richards, M Ryan, G Saleeb, R Stacey, C Thomas, J Travers, P Vallabhaneni, J Watts, M Williams.

**Tameside and Glossop Integrated Care NHS Foundation Trust** B Ryan (PI), J Abbas, A Abraheem, C Afnan, B Ahmed, O Ahmed, R Alhameed, N Amar, M Amsal, M Anim-Somuah, A Armitage, P Arora, N Aziz, C Bann, M Beecroft, P Buckley, T Bull, K Burke, A Butt, N Carter, P Chalakova, S Chandra, M Clark, M Coulding, T Dhorajiwala, S Durogbola, D Enenche, J Fallon, A Fazal, J Foster, I Foulds, V Galvis, N Garlick, H Ghanayem, R Gray, S Gulati, R Hafiz-Ur-Rehman, A Halim, M Hameed, M Hamie, T Heslop, A Hewetson, B Ho, G Hodgetts, V Horsham, W Hughes, W Hulse, A Humphries, M Hussain, N Johal, T Joshi, E Jude, M Kelly, A Kendall-Smith, M Khan, S Kilroy, A Kirk, R Law, C Loan, J Majumdar, A Masud, J McCormick, J McIntosh, V Melnic, O Mercer, T Mirza, J Mukhtar, R Murphy, F Nikita, A Nisar, B Obale, M Pattrick, P Potla, S Pudi, K Qureshi, A Rachid, M Rafique, R Rana, E Rees-Jones, H Rehman, S Ridgway, R Roberts, J Roddy, C Rolls, S Saini, M Sammut, H Savill, M Saxton, M Soliman, S Sommerfield, C Stebbing, J Thomas, A Thornton, V Turner, A Tyzack, A Umate, J Vere, L Whiteley, M Zainab.

**The Hillingdon Hospitals NHS Foundation Trust** S Kon (PI), T Bate, A Chan, W Chia, A Danga, J Ganapathi, N Hadjisavvas, B Haselden, M Holden, M Ibrahim, E Kam, J Korolewicz, M Kovac, A Lam, H Lamont, G Landers, P Law, N Mahabir, M Majumder, N Malhan, T Nishiyama, P Palanivelu, J Potter, S Ramraj, A Seckington, S Vandeyoon, W Varney, D Wahab, J Winterton, C Woollard.

**Betsi Cadwaladr LHB: Ysbyty Gwynedd** C Subbe (PI), N Abdallah, C Bishop, N Boyer, N Boyle, C Butterworth, T Collingwood, S Evans, T Grenier, J Healy, M Joishy, E Knights, A Kutera, C Lewis, M Payne, S Rawashdeh, G Rieck, W Scrase, C Speare, E Stenson, A Thomas, C Thorpe, A Van Loggerenberg.

**Guy's and St Thomas' NHS Foundation Trust** H Winslow (PI), M Abeywickrema, L Aguilar Jimenez, M Ahmad, N Amutio Martin, S Aslam, L Aslett, T Bawa, K Bisnauthsing, L Brace, L Bremner, K Brooks, B Browne, K Burns, J Butler, M Carter, B Castles , K Chan, L Chappell, A Chiapparino, P Cinardo, J Cohen, D Cooke, J Cordle, T Crowley, C D'aloia, P Dargan, A Davies, T Doudouliaki, D Finucane, M Flanagan, O Fox, D Hake, B Hamilton, D Hydes, B Jackson, T Jayatilleke, L Jimenez, L Jose, A Kasiappan Balasubramanian, J Kenny, H Kerslake, E Lee, G Lee, A Lewin, C Marsh, N Martin, L Martinez, M Mathew, A Mazzella, L McCabe, B Merrick, S Mieres, N Mlambo, F Morselli, L Nel, M Ng, Y Ng, S Nizamis, M O Toole, M Opena, M Ostermann, A Packham, D Pamela, T Rajeswaran, A Raynsford, A Rose, J Ross, K Rutkowski, K Satchithananthasivam, T Serafimova, C Singh, L Snell, N Spence, D Symington, H Tarft, H Thompson, S Tohill, E Wayman, C Williamson, K Wilson, A Xhikola, C Yearwood Martin, G Zindoga.

**Luton and Dunstable University Hospital NHS Foundation Trust** M Nisar (PI), S Tariq (Co-PI), N Ahmed (Associate PI), A Alabi, S Ali, S Allen, M Alzetani, C Ambrose, K Aneke, T Angel, Z Aung, A Aziz, R Banerjee, T Baqai, A Batla, M Bergstrom, S Bhakta, N Bibi, K Bull, T Chapman, O Choudhury, S Dipro, L Dirmantaite, M Edmondson, E Elfar, M Elgamal, H El-Sbahi, D Fishman, C Fornolles, T Forshall, A Francioni, S Gent, A Ibrahim, A Ingram, R James, D Joshi, K Kabiru Dawa, F Khan, M Khan, S Lee, C Lingam, C Luximon, N Mahdi, A Malicka, N Marcus, M Masood, R Mejri, A Moharram, C Moss, S Mottershaw, G Naik, A Ng, L Nicholls, V Parmar, I Perera, F Prasanth Raj, V Puisa, V Quick, B Ramabhadran, A Reddy, A Reka, N Riaz, B Rudran, S Sabaretnam, H Sagoo, D Salim, S Sarma, K Savlani, P Shah, D Shaw, S Siddique, S Soo, P Sothirajah, I Southern, M Tate, C Travill, V Uppal, D Vayapooree, B Wali, W Wakeford.

**North Tees and Hartlepool NHS Foundation Trust** B Prudon (PI), M Abouzaid, C Adams, A Al Aaraj (Associate PI), O Alhabsha, M Ali, E Aliberti, D Ashley, D Barker, H Bashir, B Campbell, A Chilvers, E Chinonso, V Collins, E Connell, K Conroy, E Cox, J Deane, J Dunleavy, I Fenner, C Gan, I Garg, C Gibb, S Gowans, W Hartrey, F Hernandez, J Jacob, V Jagannathan, V Jeebun, S Jones, M Khan, Y Koe, D Leitch, L Magnaye, T Mane, T Mazhani, N McDonnell, M Nafei, B Nelson, L O'Rourke, L Poole, E Poyner, S Purvis, J Quigley, A Ramshaw, H Reynolds, L Robinson, I Ross, R Salmon, L Shepherd, E Siddle, S Sinclair, M Smith, R Srinivasan, K Stewart, R Taylor, G Wallace, S Wang, L Watson, M Weetman, B Wetherill, S Wild, K Win.

**The Rotherham NHS Foundation Trust** A Hormis (PI), C Brown, D Collier, J Field, C Graham, J Ingham, V Maynard, J McCormick, S Oakley, S Poku, S Sampath, R Walker, L Zeidan.

**Walsall Healthcare NHS Trust** K Shalan (PI), M Akram, S Al-Hity, M Ali, A Alina, B Allman, J Bearpark, S Bibi, A Bland, A Bowes, L Boyd, W Campbell, Z Chandler, J Collins, L Dwarakanath, A Farg, A Foot, S Gaffarena, A Garg, A Gondal, T Gupta, A Hassan, M Islam, T Jemima, B Jones, P Joseph, O Khan, J Khatri, R Krishnamurthy, H Mahmoud, R Marsh, R Mason, M Matonhodze, S Misra, A Mohammad, O Mostafa, J Muhammad, A Naqvi, S Nortcliffe, S Odelberg, M Phipps, K Punia, H Qureshi, G Rajmohan, P Ranga, A Raymond-White, N Richardson, L Rogers, A Sheikh, P Sinha, A Srirajamadhuveeti, N Sunni, R Turel, E Virgilio, H Willis, F Wyn-Griffiths.

**Yeovil District Hospital NHS Foundation Trust** A Broadley (PI), S Abdul, N Beer, S Board, C Buckley, A Daxter, I Doig, L Every, A Getachew, B Giri, i Hamed-Adekale, E Hindley, L

Howard, R Jonnalagadda, A Kubisz-Pudelko, A Lewis, K Mansi, R Mason, A Melinte, B Mulhearn, Z N Oo, S Pallipparambil Antony, J Reid, A Shah, R Smith, A Uddin, Z Ul-Haq, D Wood.

**Betsi Cadwaladr LHB: Wrexham Maelor Hospital** D Southern (PI), S Ahmer, R Bachar, G Bennett, L Brohan, D Counsell, R Cowell, S Davies, D Dhiru, J Harris, E Heselden, M Howells, R Hughes, S Kelly, J Kilbane, A Lloyd, H Maraj, A Mashta, G Mayers, R Petersen, H Reddy, L Richards, S Robertson, S Sandow, O Smith, S Smuts, G Spencer, G Szabo, S Tomlins.

**Frimley Health NHS Foundation Trust** M Meda (PI), J Democratis (Co-PI), Y Abusamra, J Al-fori, N Barnes, N Brooks, E Bryden, L Chapman, R Conway, J da Rocha, R Dolman, A Edwards, T Foster, F Fowe, S Gee, H Gunasekara, R Ho, C Hodge, S Holland, C Hussain, S Jaiswal, A Jayadev, A Jones, N Kader, M Kain, L Kavanagh, S Lee, Y Madani, H Mahmood, A Maqsood, L McAllister, S Menzies, M Molloholli, T Nisar, J Norcliffe, S Power, A Raguro, H Rayner, F Regan, G Roberts, L Rowe-Leete, A Shaw, V Singler, C Smith, T Sonoiki, D Stewart, H Taylor, O Touma, M van de Venne, T Weerasinghe.

**Gloucestershire Hospitals NHS Foundation Trust** S Message (PI), F Ahmed, O Barker, O Bintcliffe, P Brown, R Bulbulia, S Bullock, A Chavasse, K Collins, J Collinson, T Cope, A Creamer, C Davies, W Doherty, E Eldridge, I Evans, H Farr, M Fredlund, L Gale, J Glass, S Harrington, A Hill, H Iftikhar (Associate PI), M James, S Jones, C Lim, J Mellersh, H Munby, D Nakiboneka-Ssenabulya, J Noble, M O'Flaherty, J Ord, J Patel, R Peek, T Pickett, N Robinson, C Sharp, A Simpson, M Slade, H Steer, C Thompson, A Tyler, H Uru, H Uzu, N Vallotton, J Waldron, D Ward, R Woolf.

**Homerton University Hospital NHS Foundation Trust** S Capocci (PI), A Claxton (Co-PI), Y Akinfenwa, N Aladangady, M Avari, A Begum, H Bouattia, A Boyd, R Brady, L Cabrero, L Canclini, A Chiapparino, R Corser, A Drexel, T Fong, R Frowd, C Gorman, C Holbrook, M Jagpal, S Jain, M James, J Kaur, R Leary, J Miah, R Mullett, C Nimmo (Associate PI), I Noakes, C Quah, P Reynolds, T Tanqueray, I Teeluck, L Terry, E Timlick, K Woods.

**James Paget University Hospitals NHS Foundation Trust** J Patrick (PI), B Burton (Co-PI), A Ayers, R Brooks, H Bye, J Chapman, V Choudhary, S Cotgrove, G Darylile, L Felton, D Griffiths, C Hacon, H Hall, W Harrison, P Hassell, A Hearn, F Iqbal Sait, K Mackintosh, J North, S Parslow-Williams, J Sanghera, H Sutherland, M Whelband (Associate PI), C Whitehouse, E Wilhelmsen, J Wong, J Woods.

**Poole Hospital NHS Foundation Trust** H Reschreiter (PI), S Bokhandi, J Camsooksai, C Colvin, J Dube, S Grigsby, C Humphrey, S Jenkins, E Langridge, S Patch, M Tighe, L Vinayakarao, B Wadams, M Woolcock.

**Royal Papworth Hospital NHS Foundation Trust** R Rintoul (PI), H Barker, F Bottrill, H Brzezicki, G Couch, K Dorey, R Druyeh, M Earwaker, C East, S Fielding, D Finnerty, K Fitzjohn, A Fofana, C Freeman, C Galloway, L Garner, A Gladwell, D Grogono, U Hill, V Hughes, R Hussey, N Jones, T Keady, J Kelliher, C Kosztolanyi, J Krzowski, J Mackie, P Madhivathanan, S Mepham, A Michael, B Moshy, H Munday, M Nizami, J Pack, K Paques, H Parfrey, G Polwarth, J Quijano-Campos, R Staples, S Victor, A Vuylsteke, S Webb, K Woodall, S Woods, J Zamikula.

**Royal Surrey County Hospital NHS Foundation Trust** K McCullough (PI), H Abu, C Beazley, H Blackman, P Bradley, D Burda, B Creagh-Brown, J de Vos, S Donlon, C Everden, J Fisher, H Gale, D Greene, O Hanci, L Harden, E Harrod, N Jeffreys, E Jones, J Jones, R Jordache, C Marriott, I Mayanagao, R Mehra, N Michalak, O Mohamed, S Mtuwa, K Odedra, C Piercy, V Pristopan, A Salberg, M Sanju, E Smith, S Stone, E Tarr, J Verula.

**Royal United Hospitals Bath NHS Foundation Trust** J Suntharalingam (PI), R Anstey, J Avis, C Bressington, C Broughton, A Budds, S Burnard, H Burton, R Carver, D Catibog, T Clark, T Cooke, T Costa, C Demetriou, G Dixon (Associate PI), H Duncan, F Easton, J Evans, J Fiquet, J Ford, J Goodlife, O Griffiths, R Hamlin, J Harper, T Hartley, S Jones, I Kerslake, A Kirby, J Macaro, R MacKenzie Ross, C Marchand, V Masani, Z Maseko, R Mason, H McDill, C McKerr, S Mitchard, J Noble, J Nolan, A Palmer, C Parrish, J Pullen, L Ramos, M Rich, J Rossdale (Associate PI), A Seatter, S Sturney, G Towersey, J Tyler, J Walters, K White, T Williams, S Winearl, S Zulfikar.

**Sandwell and West Birmingham Hospitals NHS Trust** S Clare (PI), M Ahmed, Y Beuvink, K Blachford, S Clamp, J Colley, P De, D Devonport, L Fares (Associate PI), M Fenton, B Gammon, M Gohel, R Grenfell, A Hayes, L Henry, S Hussain, S Joseph, F Kinney, T Knight, R Kumar, W Leong, T Lim, B Mahay, M Nelson, Y Nupa, A Orme, W Osborne, Z Pilsworth, S Potter, S Prew, N Rajaiah, A Rajasekaran, H Senya, N Shah, N Shamim, S Sivakumar, L Smith, S Spray, P Thozthumparambil, N Trudgill, A Turner, L Wagstaff, S Willetts, H Willis, M Yan.

**Barking, Havering and Redbridge University Hospitals NHS Trust** M Phull (PI), A Umaipalan (Co-PI), N Adams, M Awaly, C Baldwin, A Basumatary, S Bates, H Chandler, A Davison, G De-La-Cedra, P Dugh, K Dunne, K Fielder, A George, P Greaves, W Halder, J Hastings, H Hlaing, A Holman, S Huang, K Hunt, V Karunanithi, V Katsande, M Khalid, R Khan, P Knopp, A Loverdou, A McGregor, A Misbahuddin, A Mohamed, H Neils, D Nicholls, N O'Brien, J Olatujoye, L Parker, T Pogreban, L Rosaroso, S Sagrir, E Salciute, P Saravanamuthu, M Sharma, G Siame, H Smith, R Suthar, E Visentin, H Ward, E Yacoba.

**Doncaster and Bassetlaw Teaching Hospitals NHS Foundation Trust** C H Wong (PI), S Allen, Y Aung, R Chadwick, R Codling, V Cooke, M Cox (Associate PI), F Dunning, M Fairweather, G Herdman, M Highcock, J Kay, N Khoja, G Kirkman, C Knapp (Associate PI), M Kyi, M Luscombe, C Marsden, V Maxwell, A McGeachan, D Murray, E Mustafa, A Nasimudeen, A Natarajan, P Prentice, D Pryor, F Shafique, L Thompson (Associate PI), M Wallis, D Walstow, L Warren, N Wilkinson.

**East Kent Hospitals University NHS Foundation Trust** G Boehmer (PI), A Alegria (Co-PI), R Kapoor (Co-PI), N Richardson (Co-PI), K Adegoke, L Allen, S Anantapatnaikuni, D Baker, E Beranova, H Blackgrove, T Boumrah, P Christian, T Cosier, N Crisp, T Curtis, J Davis, J Deery, A Elgohary, T Elsefi, A Gillian, C Hargreaves, T Hazelton, G Hector, R Hulbert, A Ionita, A Knight, C Linares, S Liu, D Loader, K Lodhia, S Mandal, E Matisa, J McAndrew, K Mears, S Millington (Associate PI), M Montasser, A Moon, C Oboh, P Offord, S Parashar (Associate PI), M Patel, C Pelham, C Price, J Quindoyos, A Rajasri, J Rand, S Rogers, S Saminathan, N Schumacher, A Skaria, R Solly, D Starnes, D Stephensen, S Stirrup, L Tague, S Tilbey, S Turney, V Vasu, A Velugupati, M Venditti, R Vernall, H Weston, Z Woodward.

**Hywel Dda LHB** S Ghosh (PI), K Lewis (Co-PI), S Coetzee, K Davies, L Hill, B Icke, S Jenkins, S Griffiths, L O'Brien, Z Omar, S Peebles, E Perkins, C Williams, J Williams, C Woollard.

**Lancashire Teaching Hospitals NHS Foundation Trust** S Laha (PI), A Alty, A Ashfaq, J Beishon (Associate PI), A Bellis, D Cameron, M Chiu, W Choon Kon Yune, M Deeley, W Flesher, K Gandhi, S Gudur, R Gupta, A Huckle, G Long, P Mannion (Associate PI), A McCarrick, J Mills, P Mulgrew, S Nawaz, L Nelson, J Nixon, O Parikh, A Peer, S Punnilath Abdulsamad, D Rengan, E Seager, S Sherridan, B So, R Sonia, T Southworth, S Sowden, K Spinks, A Timoroksa, B Vernon, A Williams, K Williams, H Wu.

**NHS Dumfries and Galloway: Dumfries & Galloway Royal Infirmary** A W Hay (PI), M McMahon (Co-PI), J Candlish, P Cannon (Associate PI), J Duignan, C Jardine (Associate PI),

N Lungu (Associate PI), A Mitra, P Neill (Associate PI), S Svirpliene, J Um (Associate PI), D Williams, S Wisdom.

**NHS Greater Glasgow and Clyde: Royal Alexandra Hospital** A Corfield (PI), L Abel, S Brattan (Associate PI), A Brunton, C Clark, P Clark, S Currie, D Grieve, M Hair, M Heydtmann, E Hughes, L Imam-Gutierrez, R Keen, D McGlynn, B McLaren, A Rankin, G Ray, J Robertson, N Rodden, K Rooney, R Sundaram, N Thomson, L Walker.

**NHS Lothian: Royal Infirmary of Edinburgh** A Gray (PI), J Dear (Co-PI), M Adam, R Al-Shahi Salman, A Anand, R Anderson, J Baillie, D Baird, T Balaskas, J Balfour, C Barclay, P Black, C Blackstock, S Brady, R Buchan, R Campbell, J Carter, P Chapman, M Cherrie, C Cheyne, C Chiswick, A Christides, D Christmas, A Clarke, M Coakley, A Corbishley, A Coull, A Crawford, L Crisp, C Cruickshank, D Cryans, M Dalton, K Dhaliwal, M Docherty, R Dodds, L Donald, S Dummer, M Eddleston, S Ferguson, N Fethers, E Foster, R Frake, N Freeman, B Gallagher, E Gaughan, D Gilliland, E Godden, E Godson, J Grahamslaw, A Grant, A Grant, N Grubb, S Hainey, Z Harding, M Harris, M Harvey, D Henshall (Associate PI), S Hobson, N Hunter, Y Jaly, J Jameson, D Japp, R Kay, H Khin, L Kitto, S Krupej, C Langoya, R Lawrie, A Levynska, M Lindsay, A Lloyd, S Low, B Lyell, D Lynch, J Macfarlane, L MacInnes, I MacIntyre, A MacRaild, M Marecka, A Marshall, M Martin, E McBride, C McCann, F McCurrach, M McLeish, R Medine, H Milligan, E Moatt, W Morley, S Morrison (Associate PI), M Morrissey, K Murray, S Nelson, D Newby, K Nizam Ud Din, R O'Brien, M Odam, E O'Sullivan, R Penman, A Peterson, P Phelan, G Pickering, T Quinn, N Robertson, L Rooney, N Rowan, M Rowley, R Salman, A Saunderson, J Schafers (Associate PI), C Scott, L Sharp, A Shepherd, J Simpson, E Small, P Stefanowska, A Stevenson, S Stock, J Teasdale, E Thompson, J Thompson, I Walker, K Walker, A Williams.

**Oxford University Hospitals NHS Foundation Trust** K Jeffery (PI), M Ainsworth, C Arnison-Newgass, A Bashyal, K Beadon, S Beer, S Black, A Bloss, S Blrd, L Buck, D Buttress, W Byrne, A Capp, P Carter, L Carty, P Cicconi, R Corrigan, C Coston, L Cowen, N Davidson, K Dixon, L Downs, J Edwards, R Evans, S Gardiner, D Georgiou, A Gillesen, A Harin, M Havinden-Williams, R Haynes, C Hird, A Hudak, P Hutton, R Irons, P Jastrzebska, S Johnston, M Kamfose, K Lewis, T Lockett, F Maria del Rocio, J Martinez Garrido, S Masih, A Mentzer, S Morris, G Mounce, C O'Callaghan, Z Oliver, J Patachako, S Paulus, E Perez, L Periyasamy, L Peto, D Porter, S Prasath, C Purdue, M Ramasamy, C Roehr, A Rudenko, V Sanchez, A Sarfatti, M Segovia, T Sewdin, J Seymour, V Skinner, L Smith, A Sobrino Diaz, G Soni, M Taylor-Siddons, H Thraves, C Tsang, M Vatish, Y Warren, E Wilcock, T WIshlade.

**St Helens and Knowsley Teaching Hospitals NHS Trust** G Barton (PI), A Tridente (Co-PI), S Burnett, N Collins, S Dealing, R Garr, S Greer, N Hornby, J Keating, J Kirk, S Mayor, A McCairn, S Paul (Associate PI), S Rao, K Shuker, P Stockton, S A I Tay, T Thornton, J Williamson (Associate PI).

**Torbay and South Devon NHS Foundation Trust** L Anning (PI), P Acheampong, I Akinpelu, T Allen, S Atkins, N Aveyard (Associate PI), M Bailey, E Baird, M Ball (Associate PI), J Blackler, C Carver, M Chirgwin, T Clark, J Clouston, G Curnow, M Dawson, Z Dyar, L Ellis, C Fearnley, A Foulds, S Godlee, J Graham, A Green, C Grondin, E Hale, S Howlett, C Huggins, A Hutchinson (Associate PI), L Kyle (Associate PI), C Lam, S Maddison, R Mankiewitz, S Martin, P Mercer, B Murphy, W O’Rourke, J Palmer, M Parkinson, A Redome, J Redome, A Revill, K Subramaniam, T Tanzil-Al-Imran, R Tozer, J Turvey, C Webb, S Wright.

**United Lincolnshire Hospitals NHS Trust** M Chablani (PI), R Barber (Co-PI), C Abhinaya, S Archer, M Asghar, S Beck, S Butler, A Chingale, K Dos Santos, P Duckenfield, C Flood, O Francis, C Hewitt, A Hilldrith, K Hubbard, M Hussain, A Judd, S Karelia, A Kirkby, M Kocsor, R Mishra, P Nath, N Ndoumbe, K Netherton, M Okubanjo, L Osborne, H Palmer, G Phalod, N

Rana, A Reddy, A Rond-Alliston, B Saint, S Shephardson, A Sloan, R Spencer, K Szymiczek, J Tan, S Tavares.

**Airedale NHS Foundation Trust** T Gregory (PI), M Babirecki, H Bates, E Dooks, F Farquhar, B Hairsine, S Marsden (Associate PI), S Nallapeta, S Packham.

**Alder Hey Children's NHS Foundation Trust** D Hawcutt (PI), K Allison, R Dore, J Moss, L O'Malley, L Rad, G Seddon.

**All India Institute Of Medical Sciences, Rishikesh, Uttarakhand** P Kumar Panda (PI), V Pai (co-PI), Y Bahurupi, G Chikara, P Dhangar, D Kalita, R Kumar, V Saini, N Sarkar.

**Aneurin Bevan University LHB** S Fairbairn, (PI), L Aitken, K Arora, E Baker, M Brouns, O Burbidge, J Cann, S Cherian, S Cutler, A Dell, C Dunn (Associate PI), N Duric (Associate PI), M Edwards, S El Behery, S Goyal, J Grenville, A Griffiths, J Harris, N Hawkins, E Heron, J Hickey, M Hoare, S Hodge, G Hodkinson, A Ionescu, C Ivenso, C James, P James, T James, M Jones, S Jones, N Lawson, A Lucey, G Mallison, R Manikonda, G Marshall, S Nicholls, J Northfield, A Paracha, L Parfitt, A Pennington, L Peter, C Price, M Pynn, D Ritchie, E Rowlands, A Roynon-Reed, M Scott, C Sing, J Singh, M Singh, C Somashekar, T Szakmany, J Tuffney, R Venkataramakrishnan, G Warwick, A Waters, G Williams, S Williams, K Zalewska.

**Armed Police Force Hospital, Nepal** R Jha (PI), B Adhikari, R Ale, A Gupta, A Gurung, S Gyawali, J Khatri, S Paudel, Y Sapkota, A Shrestha.

**Ashford and St Peter's Hospitals NHS Foundation Trust** C Russo (PI), M Aquino, M Croft, S Fathima, V Frost, M Gavrila, K Gibson, A Glennon, C Gray, N Holland, K Jabbar, N Johnson, J Law, F McGee, R Pereira, L Renouf, P Reynolds, H Tarft, J Thomas, L Walding, A Williams.

**Barnsley Hospital NHS Foundation Trust** K Inweregbu (PI), R Bowmer, S Cutts, A Daniels, J Emberey (Associate PI), A Galvin, C Green, R Gupta, L Harrison, J Hartley, A Hassan, C Hirst, S Hope, M Hussain, S Hussein, A Khalil, M Longshaw, S Meghjee, A Nicholson, A Sanderson, E-J Stoner, C Swales, D Webster.

**Barts Health NHS Trust** H Kunst (PI), H Abbass, A Aboaba, A Alam, F Ali, R Allen, V Amosun, C Ardley, R Astin-Chamberlain, G Bacon, H Baillie, V Baker, S Begum, F Bibi, B Bloom, M Browne, R Buchanan, M Buckland, N Caponi, C Chan, S Chandler, B Cipriano, H Dawe, M Dawkins, P Dias, S Elia, K El-Shakankery, D Fedorova (Associate PI), M Fernandez, A Fikree, R Flynn (Associate PI), R Goiriz, P Goldsmith, M Griffiths, S Grigoriadou, K Gunganah, B Gurung, B Hack (Associate PI), P Hartridge, C Harwood, K Hashem, C Heckman, U Hemmila, D Ho, D Hobden, L Howaniec, G Huntington, I Hussain, S Iliodromiti, M Jawad, A Jayakumar, V Kapil, S Kelly, M Khan, H Khatun, J Lai (Associate PI), I Lancona-Malcolm, I Lee, D Lieberman, S Liebeschuetz, C Lindsay, O Lucey (Associate PI), K Majid, H Malcolm, C Maniero, H Marshall (Associate PI), T Martin, J Martin-Lazaro, N Matin, L Mayola, E McAleese, R McDermott, B McDonald, A Miah, I Milligan (Associate PI), A Mohammed, N Moramorell, S Naeem, T Newman, C Nic Fhogartaigh, L Noba, M Omar, N Omer, G Osoata, A Pakozdi, K Parkin, R Pearse, P Pfeffer, N Plaatjies, J Pott, J Powell, R Rachman (Associate PI), M Ramirez, K Rathod, W Ricketts, A Riddell, P Rughani, N Sahdev, K Samuels, F Santos, K Sarkar, G Sedgwick, F Seidu, B Selvarajah, M Shaikh, T Simangan, I Skene, K Smallshaw, A So, C Suarez, T Swaine (Associate PI), S Thomas, N Thorn, S Tiberi, C Tierney, G Tunesi, C Tweed, R Uddin, S Ullah, L Velauthar, K Wiles, T Wodehouse, P Woodland, H Yong, S Youssouf.

**Basildon and Thurrock University Hospitals NHS Foundation Trust** K Thomas (PI), T George (Co-PI), E Cannon, J Cartwright, M Forsey, A Ikomi, L Kittridge, G Maloney, D Mukherjee, M Mushabe, A Nicholson, K Pannu, M Pittman, J Riches, J Samuel, N Setty, A Solesbury, D Southam, M Vertue, K Wadsworth, B Yung.

**Betsi Cadwaladr LHB: Glan Clwyd Hospital** D Menzies (PI), N Abdallah, A Abou-Haggar, R Al-Sammarraie, S Ambalavanan, K Darlington, F Davies, G Davies, J Davies, L Davies, H Duvnjak, J Easton, T Grenier, M Joishy, R Lean, J Lewis, C Mackay, B Parry, K Parvin, R Poyner, R Pugh, X Qui, S Rees, J Scanlon, N Sengupta, S Ullah, H Williams.

**Birmingham Women's and Children's NHS Foundation Trust** K Morris (PI), D Jyothish (Co-PI), H Krishnan (Co-PI), E Al-Abadi, J Groves, S Hartshorn, K Hong, R Horner (Associate PI), R Howman, S Jordan, C O'Hara, S Samar (Associate PI), H Sohal, S Sultan, H Williamson, H Winmill.

**Bradford Teaching Hospitals NHS Foundation Trust** D Saralaya (PI), N Akhtar, V Beckett, L Brear, J Butler, V Drew, J Eedle, N Hawes, S Kmachia, S Moss, S Oddie, J Paget, K Regan, D Ryan-Wakeling, A Shenoy, K Storton, R Swingler, J Syson, J Todd, R Wane, A Wilson.

**Charlotte Maxeke Johannesburg Academic Hospital** I Kalla (PI), L Labuschagné, ON Makoetlane, J Mhlongo, L Murray, V Naidoo, J Nel.

**Chelsea and Westminster Hospital NHS Foundation Trust** P Shah (PI), B Mann (Co-PI), S Ahmed, K Alatzoglou, K Alizaedeh, M Al-Obaidi, K Ballard, A Barker, C Bautista, M Boffito, M Bourke, D Buche, R Bull, C Caneja, J Carungcong, F Conway, P Costa, E Dwyer, A Farah, C Fernandez, C Fung, J Garner, J Girling, E Hamlyn, A Holyome, L Horsford, N Hynes, J Irisari, M Johnson, U Kirwan, S Maheswaran, M Martineau, T Ngan, K Nundlall, C Orton, T Peters, A Sayan, A Schoolmeesters, M Svensson, A Tana, A Thayanandan, J Tonkin, B Vijayakumar, C Winpenny, F Yang, O Zibdeh.

**Chris Hani Baragwanath Academic Hospital, Soweto** M Tsitsi (PI), A Badat, C Menezes, J Robinson, S Van Blydenstein, M Venter.

**Clairwood Hospital, Mobeni** N P Magula (PI), S Pillay (Co-PI), SA Bengu, C Dipheko, S Kubheka, L Labuschagne, S Matibela, J Nel.

**County Durham and Darlington NHS Foundation Trust** J Limb (PI), V Atkinson, M Birt, C Brady, E Brown, A Cowton, V Craig, P De, D Egginton, D Fernandes, A Hallman, N Hewitson, A Ivy, D Jayachandran, J Jennings, A Kay, M Kent, T Khalifa, S Manning, S McAuliffe, S Naylor, G Nyamugunduru, J O'Brien, M Omar, K Postlethwaite, K Potts, P Ranka, G Rogers, M Sen, S Sen, J Temple, I Tzinieris, S Wadd, H Walters, M Wayman, T Wong, J Yorke.

**Croydon Health Services NHS Trust** T Castiello (PI), J Adabie-Ankrah, G Adkins, B Ajay, A Ameen, S Ashok, A Dean, S Dillane, F Fedel, V Florence, D Griffiths, I Griffiths, J Hajnik, J Hetherington, C Jones, A Latheef, S Lee, J McCammon, S Patel, A Raghunathan, P Shah, J Talbot-Ponsonby, G Tsinaslanidis, G Upson.

**Dartford and Gravesham NHS Trust** B Khan (PI), D Ail, R Aldouri, K Aung, G Awadzi, B Bassoy, R Bhalla, J Billings, S Bokhari, G Boniface, J Cernova, T Chen, P Chimbo, N Chitalia, S Danso-Bamfo, A Davis-Cook, D Depala, A Dhanoa, T Edmunds, E Fernandez, T Ferrari, B Fuller, Z Galani, A Gherman, P Grist, R Heire, L Ilves, C Kamundi, B Khan, L Lacey, E Lawrence, M Lewis, A Maric, W Martin, Z Min, C Newman, R Nicholas, N Oakley, O Olufuwa, N Pieniazek, M Protopapas, T Qadeer, S Rathore, S Sathianandan, C Scott, A Shonubi (Associate PI), S Siddique, G Sisson, M Soan, D Streit, C Stuart, M Szekely, W Umeojiako, S Urruela, B Warner, M Waterstone, S White, A-M Zafar, S Zaman.

**Dr D Y Patil Medical College, Hospital And Research Centre, Pune** S Kanitkar (PI), P Athavale, MS Barthwal, J Jadhav, P Jali, D Kadad, A Kakrani, R Kothavale, S Mahajan, B Patil, N Sairam, S Sarawade, P Sharma, BG Thomson, S Tripathy.

**East and North Hertfordshire NHS Trust** P Ferranti (PI), F Alsheikh, M Boampoaa, M Chaudhury, C Cruz, M Ebon, M Erotocritou, A Fajardo, A Frosh, S Gelves-Zapata, D Gorog, T Ingle, J Kefas, C Matei, J Mathers, L Miller (Associate PI), I Nadeem (Associate PI), Y Odedina, D Palit, L Peacock, S Tewari (Associate PI), L Ventilacion, R Zill-E-Huma, E Zinkin (Associate PI).

**East Cheshire NHS Trust** T Nagarajan (PI), X Lee (PI), S Shashaa (PI), L Alghazawi, M Babores, C Gorman, A Henderson, M Holland, R Hughes, L Huhn, M Husain, N Keenan, L Paisley, M Porteous, H Wassall, L Wilkinson, K Wolffsohn.

**East Lancashire Hospitals NHS Trust** S Chukkambotla (PI), A Batista, H Collier, S Duberley, K Geerthan, W Goddard, B Hammond, L Hoole, R Hussain, A Konstantinidis, K Marsden, A Mulla, A Newby, J Nugent, F Pickering, D Rusk, C Spalding, A Sur, D Sutton, V Taylor, N Truman, J Umeadi.

**Father Muller Medical College Hospital, Mangalore** A Bhat (PI), P Dsouza, DK Joy, V KN, Prathima.

**Gateshead Health NHS Foundation Trust** D Mansour (PI), R Allcock, M Armstrong, J Barbour, M Bokhar, J Curtis, A Dale, V Deshpande, I Hashmi, I Helgesen, E Johns, L Jones, R King, V Kumar, E Ladlow (Associate PI), R Mackie, D Mansour, C Maughan, B McClelland, W McCormick, C McDonald, M Mehmood, C Moller-Christensen, A Neal, R Palmer, R Petch, F Rasul, S Razvi, R Sharma, L Southern, G Stiller, E Watkins, H Wilkins.

**George Eliot Hospital NHS Trust** S George (PI), V Bains, M Bryant, S Bukhari, Y Chang, S Dean, N Elndari, H Gachi, J Glasgow, K Ellis, V Gulia, J Gunn, J Heys, C Ison, A Javier, T Kannan, D McDonald, J Mellor, R Musanhu, A Naeem, R Nair, N Navaneetham, S Nawaz, B Nazari, N Patel, E Sung, D Suter, M Tahir, P Vanmali (Associate PI), B Vilcinskaite, E Vlad, R Walker, I Yusuf.

**Ghana Infectious Disease Centre** C Owoo (PI), J Oliver-Commey (Co-PI), AS Adam, PN Agasiya, E K Ahadome, C Aidoo, A Amoako-Adusei, G Arhin-Sam, A Arthur, GA Asare, HA Asiamah, YE Atabudzi, S Awisi, E Boatemah, GS Dadzie, IK Duah, ED Dzidzornu, C Enimpah, S Gyambrah, E Matey, JE Mensshan, IV Omale, L Quarm, R Quartey, S Quaye, LJ Quist, SK Siabi, SK Ssiabi.

**Great Ormond Street Hospital For Children NHS Foundation Trust** M Peters (PI), L Grandjean (Co-PI), O Akinkugbe, J Andrews, A Bamford, H Belfield, S Benkenstein, C Chisholm, L Chiverton, L Dawson, J Hassell, G Jones, K Kupiec, K Leigh-Ellis, I Manjra, T McHugh, C Mellish, K Moshal, L O'Neill, J Penner, R Shamsah, D Shingadia, S Tingley, A Tomas.

**Groote Schuur Hospital, Cape Town** M Mendelson (PI), LE Hermans (Co-PI), M Adriaanse, C Bonaconsa, S Dlamini, N Hickman, M Kati, H Kyepa, G Meintjes, NS Papavarnavas, K Stoffberg, S Wasserman.

**Harrogate and District NHS Foundation Trust** A Kant (PI), A Amin, C Bennett, O Cohen, S Foxton, E Lau, S Longstaffe, C Morgan, M O'Kane, A Royson, N Singh, S Vivekananthan, A Williamson, L Wills.

**Hospital for Advanced Medicine &Surgery (HAMS)** HR Paneru (PI), D Aryal (Co-PI), S Kayastha, D Khadka, R Pangeni, R Shakya, R Shrestha.

**Hospital for Tropical Diseases, Ho Chi Minh City** N Phong (PI), TD Khoa, NT Nguyen, VT Khuong, NN Thao, TH Nguyen, DG Bao, NTM Tuyen, NV Khai, NM Thu, CTC Van, HP Thao, NVH Linh, DTN Quyen, HTC Tu, NQ Vinh, DH Le, HM Phuong, NTT Huyen

**Hywel Dda LHB: Bronglais General Hospital** M Hobrok (PI), D Asandei, B Atkins, S Jenkins, K Khan, R Loosley, H McGuinness, D McKeogh, A Mohamed, L Raisova, A Snell, H Tench, W Wolf, R Wolf-Roberts.

**Hywel Dda LHB: Withybush Hospital** J Green (PI), Y Adegeye, J Brooks, R Hughes, C Macphee, A Puffett, M Rafiq, G Ross, H Thomas.

**Isle Of Wight NHS Trust** A R Naqvi (PI), A Abbas, B Ahmed Mohamud, A Attia, A Azkoul, A Bester, A Brown, C Caraenache, G Debreceni, S Din, R Fasina, P Firi, A Gardener (Associate PI), S Grevatt, S Hinch, O Hudson, E Jenkins, N Kadam, E Karbasi, A Khan, S Knight, X Liu, C Magier, J Mattappillil, N Mburu, B Mohamud, S Mukhtar, R Naik, P Nathwani, H Ng, E Nicol, E Norris, E O'Bryan, B Osman, N Otey, M Pugh, B Rizvi, P Rogers, J Selley, I Smith, M Stevens, L Sylvester, O Tarin, C Taylor, A Tirumalai Adisesh, J Wilkins, M Woodman.

**Jawaharlal Nehru Medical College, Aligarh Muslim University, Aligarh, Uttar Pradesh** M Shameem (PI), S Anwar, A Fawad, M Tabish.

**Kingston Hospital NHS Foundation Trust** S Mahendran (PI), A Ali (Co-PI), A Joseph (Co-PI), M Agarwal, S Bacciarelli, A Baggaley, G Bambridge, S Barrett, F Bazari, P Beak, S Bidgood, L Boustred, D Caldow, T Carlin, S Cavinato, A Conroy, J Crooks, E Donnelly, A Edwards, L Elliot, S El-Sayeh, A Feben, J Fox, R Gisby, M Grout, S Hassasing, R Heath, R Herdman-Grant, E Jackson, D Jajbhay, O James, S Jones, J Khera, T Leahy, S Luck, R Maamari, M Madden, H Matthews, J May, M McCullagh, G McKnight, L Mumelj, K Muralidhara, G Natarajan, D Newman, A Nicholson, T O'Brien, J Odone, R Patterson, R Patterson, A Pavely, J Poxon, C Quamina, G Quartermaine, A Ratnakumar, R Rodriguez-Belmonte, T Sanderson, S Sharma, R Simms, D Sivakumran, A Sothinathan, A Swain, S Swinhoe, M Taylor, M Trowsdale Stannard, B Ummat, J Vance-Daniel, H Warren-Miell, R Williams, T Woodhead, D Zheng.

**Komfo Anokye Teaching Hospital** Y Hardy (PI), C Aboah, F Acheampong, L Addai-Boampong, P Adjepong, P Afrim, FA Afriyie, JA Agyekum, K Agyen, J Amoah-Dankwa, M Ansah, JM Ayerh, PG Barning, KA Boahen, J Bonney, KG Danso, M Donkor, D Dwomoh Nkrumah, H Haywood Hasford, M Kuma-Mintah, O Lambert, LN Lartey, L Micah-Amuah, PAB Minkah, L Nyanor, E Ofori, PK Omane-Donkor, A Opata, MA Oteng, C Poku, M Siaw-Frimpong, AB Wongkyezeng.

**Lewisham and Greenwich NHS Trust** S Kegg (PI), A Aghababaie, H Azzoug, E Bates, M Chakravorty, K Chan, C Chin-Saad, P Coakley, R Dodds, E Gardiner, A Hastings, D Jegede, J Juhl, H Kelly, K Law, M Lewis, M Magriplis, A Mascagni, C Milliken, J Muglu, D Mukimbiri, M Nadheem, T Nair, M Nyirenda, T OConnor, T Ogbara, L Olaivar, R Olaiya, E Omoregie, C Onyeagor, V Palaniappan, A Pieris, S Pilgrim, T Simpson, A Taylor, E Treus Gude, K Wesseldine, R Williams, M Woodman, E Woolley.

**Liverpool Women's NHS Foundation Trust** R McFarland (PI), M Dower, S Holt, A Mahdi, C Morgan, E Neary, A Smith, M Turner, E Willis.

**Medical College Kolkata, Kolkata** A Talukdar (PI), K Chaudhuri, D Sarkar.

**Mid Essex Hospital Services NHS Trust** A Hughes (PI), J Radhakrishnan (Co-PI), R Arnold, T Camburn, E Cannon, C Catley, E Dawson, L Durdle, N Fox, H Gerrish, S Gibson, H Guth,

M Iman, L James, F McNeela, C Mitchell, A Rao, S Reid, B Singizi, S Smolen, M Vitaglione, S Williams, L Willsher, J Wootton.

**National Hospital for Tropical Diseases, Hanoi** PN Thach (PI), NT Cap, NT Ha, LNM Hoa, DTT Thuy, DQ Tung, DP Khiem, PV Phuc, TV Kien, VD Linh, DTT Hang, TV Giang, TT Hien, HTK Oanh

**Nepal Police Hospital** D Paudel (PI), B Rajbhandari (Co-PI), NB Chaudhary, B Gajmer, M Gartaula, J Khatri, SS Kunwar, M Maharjan, P Maskey, K Neupane, R Niroula, A Regmi, S Singh, S Singh, BJ Thakuri, S Yadav.

**NHS Ayrshire and Arran: University Hospital Ayr** K Walker (PI), M Boden, C Burns, D Callaghan, R Cuthbertson, K Gibson, D Gilmour, M Henry, L Kelly, J Locke, L McNeil, S Meehan, A Murphy, K Naismith, K Prasad, M Rodger, R Slingsby, C Turley, S Walton, M Wilson, S Wood.

**NHS Ayrshire and Arran: University Hospital Crosshouse** A Clark (PI), T Adams, S Allen, E Anderson, L Barker, C Burns, D Callaghan, N Connell, V Dey, A Dunleavy, F Elliott, K Gibson, D Gilmour, K Grant, M Henry, J Locke, T Massa, L McNeil, S Meehan, A Murphy, E Peggie, M Rodger, S Smith, C Turley, S Walton, D Wilkin, M Wilson, S Wood.

**NHS Borders: Borders General Hospital** A Scott (PI), G Alcorn, S Alcorn, J Aldridge, J Bain, C Barnetson, P Bell, A Campbell, B Chow, S Corbet, F Craighead, J Dawson, E Dearden, S Dickson, G Donaldson, T Downes, A Duncan, H El-Taweel, C Evans, T Fairbairn, L Finlayson, C Flanders, J Fletcher, J Foo, R Gallagher, N Hafiz, F Hall, M Herkes, M Hume, J Inglis, L Knox, J Lonnen, J Manning, H Matthews, A McLaren, C Murton, R Murton, B Muthukrishnan, E Nicol, F North, C Osbourne, R Porter, K Ralston, R Richmond, F Rodger, H Sakuri, B Soleimani, R Stewart, R Sutherland, A Tan, M Tolson, J Wilkie, R Williamson.

**NHS Golden Jubilee National Hospital** B Shelley (PI), V Irvine, F Thompson.

**NHS Grampian: Aberdeen Royal Infirmary** J Cooper (PI), R Soiza (Co-PI), V Bateman, M Black, R Brittain-Long, K Collie, K Colville, D Counter, A Coutts, S Devkota, P Dospinescu, F Flett, P Ganley, L Hakeem, R Hunter, J Irvine, L Kane (Associate PI), C Kaye, A Khan, R Laing, A Mackenzie, M MacLeod, G Malik, J McLay, D Miller, J Noble, K Norris, Y Pang, V Taylor, K Thomasson, I Tonna, A Vileito, L Walker.

**NHS Greater Glasgow and Clyde: Inverclyde Royal Hospital** M Azharuddin (PI), H Papaconstantinou (Co-PI), D Cartwright, W Gallagher, J Hampson, T McClay, E Murray, O Olukoya.

**NHS Highland** B Sage (PI), R Acquah, L Adamu-Ikeme, C Barr, F Barrett, A Batty, W Beadles, C Bradley, R Campbell, A Cochrane, R Cooper, I Dawson, K Donald, J Finlayson, A Goh, L Heron, G Jervis, P Laidler, S Macleod, S Makin, J Matheson, D McDonald, M McKenzie, C Millar, K Monaghan, L Murray, M O'Hara, L O'Keefe, D Patience, M Peirse, S Reilly, M Robertson, N Shahzad, G Simpson, K Watson, J Wilson.

**NHS Lothian: Royal Hospital for Children and Young People** C McDougall (PI).

**NHS Lothian: St John's Hospital** S Lynch (PI), R Anderson, S Begg, J Bentley, S Brady, M Colmar, R Frake, A Gatenby, C Geddie, F Guarino, S Guettari, C Hurley, C Kuronen-Stewart, A MacRaild, M Mancuso-Marcello, M Marecka, G McAlpine, N McCullough, A Megan, T North, O Otite, L Primrose, C Rees, L Rooney, A Saunders, A Saunderson, A Stevenson, S Stock, A Wakefield, E Walsh, J Wraight, T Wright, S Yusef.

**NHS Tayside: Ninewells Hospital** J Chalmers (PI), H Abo-Leyah, C Deas, H Loftus, A Nicoll, A Strachan, J Taylor, C Tee.

**NHS Western Isles** M Murdoch (PI), A Apostolopoulos (Co-PI), G Stanczuk (Co-PI), S Klaczek.

**Norfolk and Norwich University Hospitals NHS Foundation Trust** E Mishra (PI), D Archer, C Atkins, S Bailey, K Bohanan, M Cambell-Kelly, E Chiang, P Clarke, L Coke, B Collinson, J Cook, A Cooper, M Cornwell, A Dann, P De Souza, M Del Forno, A Elsheikh, S Fletcher, C Fraser, S Gajebasia, H Gorick, N Gray, A Haestier, S Hand, M Harmer, L Harris, L Hudig, M Ilsley, K Jethwa, L Jones, A Kamath, D Kelly (Associate PI), J Kennedy, J Keshet-Price, J Knights, E Kolokouri, V Licence, E Lowe, E Malone, M Marks, G Maryan, K Metcalfe, A Miller-Fik, P Moondi, M Morris, K Myint, K Nagumantry, W Ng, J Nortje, M Oliver, M Patel, T Potter, R Rallan, S Rehman, J Roberts, A Sanda-Gomez, A Sanz-Cepero, D Sethi, J Staines, K Stammers, E Tropman, M Ur Rasool, S Walton, N Ward, D Watts, C Websdale, R Wiseman, C Wright.

**North West Anglia NHS Foundation Trust** K Rege (PI), K Abbas, B Abdelqader, R Abdul-Raheem, A Achara, K Adatia (Associate PI), Z Aftab, C Agbo, A Ahonia, O Akindolie, O Aktinade, A Al Swaifi, S Al-Juboori, F Allan, A Al-Rabahi, R Ambrogetti, S Amin, D Arcoria, K Aung, A Auwal, G Aviss, A Azman Shah, A Babs-Osibodu, K Bahadori, J Baker, T-A Baker, N Bakhtiar, K Bandaru, A Bhanot, K Bhatt, J Bhayani, K Blaylock, T Bodenham, T Bond, H Boughton, H Bowyer, D Braganza, S Brooks, D Butcher, M Butt, N Butterworth-Cowin, R Buttery, A Cabandugama, J Carpenter, P Carter, A Catana, L Cave, R Chaube, M Cheung, Z Chikwanha, C Chisenga, S Choi, S Choudhury, V Christenssen, M Chukwu, B Clearyb, H Cooper, C Cordell, D Corogeanu, A Cowan, R Croysdill, S Dahiya, M Davies, R De Pretto, J de Souza, S Diaz, B Donohue, O Drosos, N Duff, L Dufour, O Ebigbola, C Eddings, M El-Din, I Eletu, N Enachi, J Faccenda, A Feroz, L Fieldhouse, L Finch, N Flaris, C Freer, P Gerard, E Gillham, A Gkoritsa, P Goodyear, R Gooentilleke, R Gosling, G Gosney, V Goss, S Goyal, J Hafsa, W Halford, S Havlik, D Hornan, Z Horne, T Hoskins, M Hunt, C Huson, A Idowu, S Inglis-Hawkes, I Iordanov, M Ishak, M Iyer, D Jafferji, E Jarnell, H Javed, E Johnson, A Jones, D Jones, J Jones, T Jones, E Kallistrou, J Kamara, R Kamath, S Kar, R Kark, N Kasianczuk, R Kassam, D Kaur, A Kerr, S Khalid, A Khan, A Khan, M Kheia, F Kleemann, E Kopyj, G Koshy, A Kozak Eskenazia, R Kurian, S Lahane, M Lami, D Lamrani, K Leng, D Limbu, C Lippold, F Magezi, M Mahmud, A Malik, J Marshall, R Matewe, F Maxton, N McCammon, K McDevitt, S McGuire, C McMurran, W Mensah, P Mitchell, L Molloy, P Morzaria, Q Moyo, I Mugal, O Muraina, N Muru, A Mustafa, Y Myint, S Nazir, O Oderinde, S Ojha, I Okpala, C Oladipo, T Old, G Oleszkiewicz, N Olokoto, H Orme, S O'Sullivan, P Paczko, S Pal, A Pandey, N Pao, N Parker, S Parker, A Patel, S Pathak, S Peacock, B Peirce, T Perumpral, S Ponnusamy, S Pooboni, S Poon, J Porter, A Prasad, A Rahimi, J Rajkanna, M Rather, R Renu Vattekkat, Z Rimba, S Rizvi, P Roberts, S Sahedra, S Saliu, M Samyraju, J Sanyal, V Saulite, R Schofield, S Schofield, K Scholes, S Shah, W Shah, S Shahad, P Sharma, L Simon, L Singh, P Sivasothy, E Smith, S Stacpoole, O Syed, B Tan, K Tan, J Taylor, N Temple, K Thazhatheyil, M Uddin, S Vardy, N Veale, A Waheed Adigun, D Walter, M Walton, B Whelan, A White, G Williams, C Willshire, M Wong, H Wroe.

**North West Boroughs Healthcare NHS Foundation Trust** A Baldwin (PI)

**Northampton General Hospital NHS Trust** M Zaman (PI), B Abdul, A Abdulmumeen, N Abdulshukkoor, A Adnan, M Ahammed Nazeer, B Ahmed, M Ashab, A Bazli, N Benesh, W Chin, N Cunningham, H Daggett, E Davies, E Elmahi, H Enyi, S Fawohunre, J Fernando, N Geoghegan, J Glover, J Hague, K Hall, C Hallett, K Haresh, J Hewertson, J Hosea, N Htoon, H Huynh, M Idrees, C Igwe, H Imtiaz, M Irshad, A Ismail, R Jeffrey, J Jith, P Joshi, R Kaliannan Periyasami, A Khalid, M Khalid, R Kodituwakku, R Kontogonis, P Lopez, A Mahmood, M Malanca, I Mapfunde, V Maruthamuthu, S Masood, M Matharu, A Merchant, F Merchant, S

Naqvi, N Natarajan, R Natarajan, K Nawaz, O Ndefo, O Ndefo, O Ogunkeye, S Paranamana, N Pugh, A Raj, K Rashid, M Rogers, M Saad, A Sajid, G Selvadurai, A Shah, M Shahzeb, U Shamji, N Shrestha, A Singh, K Smith, B Sohail, M Spinks, L Stockham, A Takyi, Y Teoh, W Ul Hassan, S Ullah, H Vayalaman, S Wafa, T Ward, R Watson, D White, L Ylquimiche Melly, R Zulaikha.

**Northern Devon Healthcare NHS Trust** R Manhas (PI), C Alexander, K Allen, F Bellis, H Black, S Bosence, A Brunchi, D Dalton, D Davies, J Gilham, N Hadfield, A Hanson, R Hartley, R Holbrook, J Hunt, G Isitt, M Jeelani, M Lamparski, E Lekoudis, S Ley, B Rowlands, C Santos Ferreira De Almeida, A Umeh, L van Koutrik, C Vooght, C White, E Willis, H Wright.

**Northern HSC Trust** P Minnis (PI), W Anderson, S Baird, M BinRofaie, J Burns, C Cruickshank, L Davidson (Associate PI), L Denley, R Donnelly, G Doran, M Drain, A Fryatt, J Gallagher, M Hollyer, M Kawser, L Kingsmore, O Lepiarczyk, I Masih, C McGoldrick, S McKenna, M McMaster, S McNeill, E Murtagh, M Nugdallah, T Scullion, M Stewart, M Wray.

**Northern Lincolnshire and Goole NHS Foundation Trust** A Mitra (PI), R Abrams, N Akhtar, H Al-Moasseb, S Amamou, T Behan, S Biuk, M Brazil, M Brocken, C Burnett, C Chatha, M Cheeseman, L Cottam, T Cruz Cervera, O Davies, K Dent, C Downing, C Dyball, K Edwards, M Elhadi, R Elmahdi, Q Farah, S Farooq, S Gooseman, J Hargraves, M Haroon, J Hatton, E Heeney, J Hill, E Horsley, R Hossain, S Hudson, A Hussien, D Hutchinson, J Hyde-Wyatt, A Ibrahim, M Iqbal, N James, S Khalil, M Madhusudhana, A Marriott, M Masood, G McTaggart, K Mellows, R Miller, U Nasir, M Newton, GCE Ngui, S Pearson, C Pendlebury, R Pollard, N Pothina, D Potoczna, S Raha, A Rehan, SAS Rizvi, A Saffy, J Sanders, H Sangha, K Shams, C Shaw, A Shirgaonkar, S Spencer, R Stead, R Sundhar, D Taylor, E Thein, E Waldeck, L Warnock, K Wong.

**Pennine Acute Hospitals NHS Trust** J Raw (PI), R Tully (Co-PI), K Abdusamad, Z Antonina, E Ayaz, B Blackledge, P Bradley, F Bray, M Bruce, E Bullock, C Carty, B Charles, G Connolly, C Corbett, J Cornwell, S Dermody, L Durrans, U Elenwa, E Falconer, J Flaherty, C Fox, J Guerin, D Hadfield, J Harris, J Haslam, S Hey (Associate PI), L Hoggett, A Horsley, C Houghton, L Howard-Sandy, S Hussain, R Irving, P Jacob, D Johnstone, R Joseph, P Kamath, T Khatun, T Lamb, H Law, M Lazo, G Lindergard, S Lokanathan, L Macfarlane, S Mathen, S McCullough, P McMaster, D McSorland, J Melville, B Mishra, G Moth, M Mulcahy, S Munt, J Naisbitt, A Neal, R Newport, G O'Connor, D O'Riordan, I Page, V Parambil, J Philbin, M Pinjala, C Rishton, M Riste, J Rothwell, M Sam, Z Sarwar, L Scarratt, A Sengupta, H Sharaf, J Shaw, J Shaw (Associate PI), K Shepherd, A Slack, D Symon, H T-Michael, A Ustianowski, O Walton, S Warran, S Williams.

**Pokhara Academy of Health Sciences** S Baniya (PI), SA Adhikari, SC Baral, A Karki, P KC, S Pathak, KK Roka, A Thapa.

**Royal Brompton & Harefield NHS Foundation Trust** A Shah (PI), A Reed (Co-PI), A Angela, B Araba, L Banton, A Catelan Zborowski, M Damani, P De Sousa, V Jardim, K Mahay, T Maria Pfyl, H Middleton, H Passmore, T Poonian, C Prendergast, P Rogers, G Sloane, N Soussi, J Tan, V Teli, V Thwaiotes, L Tous Sampol, J Wallen, A Watson.

**Royal National Orthopaedic Hospital NHS Trust** R Baumber (PI), D Brooking, F Fitzgerald, E Hanison, J Hunt.

**RSU Martha Friska** F Ginting (PI), M Barimbing, I Rambe, D Saragih, A Tantri

**RSU Puri Raharja, Denpasar** M Sri (PI), NLPE Arisanti (Co-PI), NKN Andari, NM Artini, P Khairunnisa, R Nurpeni, K Puspatriani, SI Putu, A Rimainar, T Valleri.

**Rumah Sakit Metropolitan Medical Centre, Jakarta** E Nelwan (PI), A Abidin, M Butar, A Cahyareny, L Handayani, H Istiqomah, S Kumala Dewi, T Monika, S Prabowo, N Pratiwi, K Puspatriani, N Rika, A Saraswati, D Sari, P Suherman, T Tesha, J Wanda, K Wattimena, N Yufaniaputri.

**Salford Royal NHS Foundation Trust** P Dark (PI), J Allen, E Apetri, N Bakerly, B Blackledge, L Catlow, B Charles, M Collis, R Doonan, J Harris, A Harvey, K Knowles, S Lee, T Marsden, E Mclaughlan, L McMorrow, A Michael, J Pendlebury, J Perez, S Simpkins, G Squires, R Sukla, M Taylor, V Thomas, D Walker, S Warran, O Wickens (Associate PI).

**Sheffield Children's NHS Foundation Trust** P Avram (PI), A Bellini, F Blakemore, H Chisem, J Clements, H Cook, S Gormely, D Hawley, C Kerrison, N Lawrence, G Margabanthu, A McMahon, N Roe, F Shackley, J Sowter, T Williams.

**Somerset NHS Foundation Trust** J Pepperell (PI), J Ashcroft, R Burgess, G Chilcott, S Crouch, I Cruickshank, T Dean, J Dryburgh-Jones (Associate PI), J Foot, L Graham, N Grieg, K James, S James, K Johnson, C Lanaghan, D Lewis, A Locke, C Lorimer, J Lucas, H Mills, G Modgil, C Morgan (Associate PI), A Moss, M Nixon, S Northover, K O'Brien, J Page (Associate PI), I Parberry, R Purnell, K Roberts, A Robinson (Associate PI), J Rogers, C Shovelton, C Susan, C Thompson, N Thorne, R Twemlow, C Vickers, R Wallbutton, A Whitcher, J Youens, E Zebracki.

**South Warwickshire NHS Foundation Trust** S Tso (PI), P Parsons (Co-PI), S Bird, B Campbell, G Kakoullis, F Mackie, C O'Brien, P Rai (Associate PI), A Smith, K Webb.

**Southend University Hospital NHS Foundation Trust** G Koduri (PI), F Hayes (Co-PI), V Vijayaraghavan Nalini (Co-PI), S Badhrinarayanan, N Chandran, J Galliford, L Ginn, S Gokaraju V Gupta, P Harman, M Mercioniu, D Qureshi, M S Rabbani (Associate PI).

**Southern HSC Trust** R Convery (PI), J Acheson, J Brannigan, D Cosgrove, C McCullough, D McFarland, R McNulty, S Sands, O Thompson.

**Southport and Ormskirk Hospital NHS Trust** S Pintus (PI), A Nune (Co-PI), A Ahmed, H Ahmed, L Bishop, D Dickerson, Z Haslam, E Isherwood, M Jackson, A Morris, M Morrison, R Purves (Associate PI), V Subramanian, A Tageldin (Associate PI).

**Sukraraj Tropical and Infectious Disease Hospital, Nepal** M Rawal (PI), B Chalise, S Gyawali, A Gupta, Y Sapkota, S Paudel, J Khatri, S Pant, P Gyanwali, U Devkota, S Mandal, L Bhandari, K Paudel, N K.C., S Karki, A Chaudhary, L Chaulagain, R Kumar, R Jaiswal, M Bajracharya, S Gyawali, S Magar, J Neupane, G Shrestha, R Jaiswal, K Poudel.

**Surrey and Sussex Healthcare NHS Trust** E Potton (PI), S Abbasi, D Acharya, A Acosta, L Ahmed, S Ali, M Alkhusheh, V Amosun, A Arter, M Babi, J Bacon, K Bailey, N Balachandran, S Bandyopadhyam, L Banks, J Barla, T Batty, S Bax, A Belgaumkar, G Benison-Horner, A Boles, N Broomhead, E Cetti, C Chan, I Chaudhry, D Chudgar, J Clark, S Clueit, L Clutterbuck, S Collins, E Combes, G Conway, O Curtis, M Das, M Daschel, S Davies, A Day, M Dhar, K Diaz-Pratt, C Dragan, H Dube, V Duraiswamy, S Edwards, J Elias, A Ellis, T-Y Ellis, J Emmanuel, A Engden, Y Fahmay, B Field, K Fishwick, U Ganesh, C Gilbert, T Giokanini-Royal, E Goudie, S Griffith, S Gurung, R Habibi, C Halevy, A Haqiqi, R Hartley, A Hayman, J Hives, S Holden, M Horsford, S Hughes, C Hui, F Huq, R Hussain, C Iles, L Jackson, N Jain, A James, D Jayaram, E Jessup-Dunton, T Joefield, A Khadar, N Khan, W Kieffer, E Knox, R Kumar, V Kumar, V Kurmars, H Lafferty, F Lamb, R Layug, N Leitch, W Lim, U Limbu, R Loveless, M Mackenzie, N Maghsoodi, S Maher, M Maljk, I Man, N McCarthy, B Mearns, C Mearns, P Morgan, K Morgan-Jones, G Mortem, G Morton, B Moya, G Murphy, S Mutton, A Myers, T Nasser, J Navaratnam, S Nazir, S Nepal, K Nimako, L Nimako, C O'Connor, I

Odysseos-Beaumont, A Patel, K Patel, J Penny, V Phongsathorn, P Pillai, M Poole, N Qureshi, I Rajkumar, S Ranjan, A Rehman, T Samuels, S Sathianandan, E Scott, G Sekadde, A Sharma, G Sharp, S Shotton, O Simmons, P Singh, S Smith, K Sri Paranthamen, S Suresh, B Tejero Moya, K Thevarajah, L Thomas, H Timms, N Tomasova, S Tucker, I van Bruggen, S Vara, C Vaz, S Weller, J White, M Wilde, I Wilkinson, C Williams, M Win, D Woosey, D Wright.

**Teku Hospital (Shukraraaj Tropical & Infectious Disease Hospital)** A Bastola (PI), B Chalise (Co-PI), K Maharjan (Co-PI), L Bhandari, U Devkota, A Gupta, S Gyawali, J Khatri, S Mandal, S Pant, K Paudel, M Paudel, S Paudel, A Phuyal, B Poudyal, G Pradip, S Rajbhandari, D Rawal, Y Sapkota.

**The Christie NHS Foundation Trust** V Kasipandian (PI), A Binns, J King, P Mahjoob-Afag, R Mary-Genetu, P Nicola, A Patel, R Shotton, D Sutinyte.

**The Dudley Group NHS Foundation Trust** H Ashby (PI), P Amy, S Ashman-Flavell, S de Silva, J Dean, N Fisher, E Forsey, J Frost, S Jenkins, A John, D Kaur, A Lubina Solomon, S Mahadevan-Bava, T Mahendiran, V Moore, M O'Toole, S Pinches, D Rattehalli, M Reay, U Sinha, L Stanton, M Subramanian, J Vamvakopoulos, S Waidyanatha, S Westwood.

**The Princess Alexandra Hospital NHS Trust** U Ekeowa (PI), S Sakthi (Co-PI), Q Shah (Co-PI), M Anwar, B Badal, K Bumunarachchi, G Cook, A Daniel, K Dyer, A Easthope, J Finn, C Freer, A Gani, S Harris, E Haworth, L Hughes, K Ixer, N Konar, S Kuckreja, G Lucas, C Muir, P Nabayego, S Naik, O Newman, F Ozdes, R Ragatha, P Russell, R Saha, L Sandhu, E Shpuza, N Staines, S Waring, L Wee, N White, T White, A Zahoor.

**The Queen Elizabeth Hospital, King's Lynn, NHS Foundation Trust** M Blunt (PI), S Abubacker, J Ali, O Aribike, K Beaumont, K Bishop, H Bloxham, P Chan, Z Coton, H Curgenven, M Elsaadany, T Fuller, T Ganeshanathan, M Gigi, G Hasnip, M Iqbal, M Israa, S Jeddi, S Kamerkar, E Lim, E Nadar, K Naguleswaran, O Poluyi, H Rangarajan, G Rewitzky, F Richardson, S Ruff, S Shedwell, E Ukaegbu, A Velusamy, H Webb.

**The Royal Marsden NHS Foundation Trust** K Tatham (PI), P Angelini, E Bancroft, E Black, A Dela Rosa, E Durie, M Hogben, S Jhanji, I Leslie, A Okines, I Sana, S Shepherd, N Taylor, S Wong.

**RS UNAIR, Surabaya** N Nasronudin (PI), C Nugroho, R Prananingtias, E Puji Lestari, B Rachman, A Rosyid, F Utomo, A Veterini.

**RSUP Dr Hasan Sadikin, Bandung** U Sumardi (PI), A Albertus, A Bachti, A Amelia Ganefianty, EN Lestari, TT Maulidya, W Naftalia, CB Prayuda, I Silanas, RF Sobarna, T Tulus Satriasih, W Rina, H Yovita.

**The Walton Centre NHS Foundation Trust** R Davies (PI), H Arndt, A Clyne, E Hetherington, G Hull.

**University Hospitals Coventry and Warwickshire NHS Trust** K Patel (PI), C Imray (Co-PI), N Aldridge, A Ali, C Bassford, T Brigstock, A Campbell, S Cliff, S Dale, D Davies, G Evans, E French, R Grenfell, S Hewins, D Hewitt, M Hussain, J Jones, A Jose, R Kumar, B Lara, E Mshengu, S Quenby, H Randheva, K Read, P Satodia, E Sear, R Srikantaiah, C Stokes, M Truslove, K Westwood, S Wurie, J Zhang.

**Velindre NHS Trust** J Powell (PI), R Adams, A Jackson.

**Warrington and Halton Teaching Hospitals NHS Foundation Trust** M Murthy (PI), R Arya, A Baluwala, T Blunt, R Chan, L Connell, M Davey, L Ditchfield, G Drummond, A Ibrahim, J

Little, N Marriott, B Mathew, M Moonan, T Nagarajan, S Patel, H Prady, L Roughley, S Sharma, H Whittle.

**West Hertfordshire Hospitals NHS Trust** R Vancheeswaran (PI), L Norris, V Page, J Palman, A Yousafzar, X Zhao.

**Western HSC Trust** M Kelly (PI), D Concannon, P Corry, K Ferguson, L Gelmon, D Glowski, J Kara, B Keegan, D McClintock, P McDermott, T McManus, V Mortland, F Okpoko, J Pastrana, K Ryan, D Smalls, N Smyth, S Sreenivasan, J Wieboldt.

**Worcestershire Acute Hospitals NHS Trust** C Hooper (PI), K Austin, T Dawson, A Durie, C Hillman-Cooper, O Kelsall, M Ling, Z Parvez, D Stocker, S Stringer, J Thakrar, H Tranter, J Tyler, P Watson, B Wild, D Wilson, H Wood.

**Wrightington, Wigan and Leigh NHS Foundation Trust** A Ashish (PI), I Aziz, C Brown, L Claxton, D Heaton, L Hirst, S Hough, J Jeganathan Ponraj, C Moore, V Parkinson, M Regan, E Robinson, T Taylor, C Tierney, A Verma, N Waddington, J Wedlin, C Williams, C Zipitis.

**York and Scarborough Teaching Hospitals NHS Foundation Trust** D Yates (PI), M Abdelfattah, A Abung, J Anderson, P Antill, S Appleby, P Armtrong, J Azam, T Berriman, D Bull, K Chandler, O Clayton, A Corlett, D Crocombe, S Davies, K Elliott, L Fahel, K Freshwater, J Ghosh, N Gott, D Greenwood, T Holder, K Howard, J Ingham, P Inns, W Lea, E Lindsay, K Mack, N Marshall, S McMeekin, R Miller, R Molyneux, T Momoniat, P Nikolaos, M O'Kane, G Patrick, H Pearson, P Ponnusamy, A Poole, N Price, R Proudfoot, G Purssord, H Redfearn, S Roche, Z Scott, C Sefton, S Shahi, D Smith, B Sohail, E Suleiman, S Sutton, R Thomas, A Turnbull, L Turner, H Watchorn, E Wiafe, J Wilson.

### Supplementary Methods

#### Study organization

The RECOVERY trial is an investigator-initiated, individually randomised, open-label, controlled trial to evaluate the efficacy and safety of a range of putative treatments in patients hospitalized with COVID-19. The protocol is available at www.recoverytrial.net. The trial is being conducted at 177 National Health Service (NHS) hospital organizations in the United Kingdom and hospitals in Vietnam, Nepal, Indonesia, South Africa, India, and Ghana. The trial is coordinated by a team drawn from the Clinical Trial Service Unit and the National Perinatal Epidemiology Clinical Trials Unit within the Nuffield Department of Population Health at University of Oxford, the trial sponsor. Support for local site activities is provided by the National Institute for Health Research Clinical Research Network.

Access to relevant routine health care and registry data is supported by NHS DigiTrials, the Intensive Care National Audit and Research Centre, Public Health Scotland, National Records Service of Scotland, and the Secure Anonymised Information Linkage (SAIL) at University of Swansea.

*Regulatory and ethics approvals*

| Country | Regulatory body | Ethics |
|---|---|---|
| UK | Medicines and Healthcare products Regulatory Agency (MHRA) | Cambridge East Research Ethics Committee (ref: 20/EE/0101) |
| Vietnam | Vietnam Ministry of Health | Hospital for Tropical Diseases Ethics Committee* |
| Nepal | Government of Nepal Department of Drug Administration | Ethical Review Board, Nepal Health Research Council (NHRC) |
| Indonesia | Badan Pengawas Obat Dan Makanan (BPOM) | Ethics Committee of the Faculty of Medicine, University of Indonesia* |
| Ghana | Food and Drugs Authority | Ghana Health Service Ethics Review Committee |
| South Africa | South African Health Products Regulatory Authority (SAHPRA) | The University of the Witwatersrand, Human Research Ethics Committee* |
| India | Not required | ICMR Central Ethics Committee on Human Research (CECHR) |

* For countries without a national ethics committee the name of the committee approving the first site is listed.

#### Laboratory analysis

*Serum anti-SARS-CoV-2 antibodies and nucleocapsid antigen*

Serum samples were to be collected at randomisation from all participants and analysed at the UK Health Security Agency SARS-CoV-2 Serology laboratory, Porton Down, UK. Total antibodies to SARS-CoV-2 spike protein and nucleocapsid protein were measured using the Roche Elecsys Anti-SARS-CoV-2 S and Elecsys Anti-SARS-CoV-2 assays, respectively. Patients were classified as positive or negative using manufacturer defined thresholds. Serum nucleocapsid antigen was measured using the Roche Elecsys SARS-CoV-2 Antigen assay. This assay had previously been validated only on respiratory specimens, so additional validation was performed to evaluate performance on serum samples, including repeatability, linearity, and cross reactivity (appendix pp 188-204). Samples were classified as positive or negative using the median value for all participants (COI 0.626).

*Nasal SARS-CoV-2 viral PCR and sequencing*
Nose (mid-turbinate) swabs were collected at randomisation (day 1), on day 3, and on day 5 from all UK patients and analysed at the Oxford University Hospitals microbiology laboratory, Oxford, UK. Swabs had RNA extraction using the Kingfisher Flex system, and SARS-CoV-2 PCR using the TaqPath COVID-19 RT-PCR assay (Thermo Fisher Scientific, Massachusetts, US). A standard curve was produced using a panel of whole virus SARS-CoV-2 standards (Qnostics SARS-CoV-2 Analytical Q Panel), which was used to quantify viral genome copy concentration in samples.

Samples with sufficient viral concentration (>2.6 log viral copies/ml) were sequenced using the Oxford Nanopore Technologies Midnight and tiling amplicon protocol, using a GridION instrument (Oxford Nanopore Technologies, Oxford, UK).[1] Sequence data was processed using the Global Pathogen Analysis System (https://gpas.global), a web-based sequence analysis pipeline that uses the Viridian assembly system and assigns viral lineage using Pangolin. Sequence data were used to detect spike protein mutations associated with >5-fold reduction in sotrovimab neutralisation, which were identified from the summary of product characteristics (https://www.medicines.org.uk/emc/product/13097/smpc) and the Stanford University Coronavirus Antiviral and Resistance Database. (https://covdb.stanford.edu/).[2] Amino acid substitutions S371F/L, which are present in all Omicron variants and are associated with moderate (3-5-fold) reduction in neutralisation, were not included. The following spike protein mutations were included:

- P337A/H/K/L/N/Q/R/S/T
- E340A/D/G/I/K/L/N/Q/R/S/V
- T345P
- K356A/E/M/N/Q/R/S/T
- L441N

1 Constantinides B, Webster H, Gentry J, et al. Rapid turnaround multiplex sequencing of SARS-CoV-2: comparing tiling amplicon protocol performance. medRxiv 2022; : 2021.12.28.21268461.

2 Tzou PL, Tao K, Pond SLK, Shafer RW. Coronavirus Resistance Database (CoV-RDB): SARS-CoV-2 susceptibility to monoclonal antibodies, convalescent plasma, and plasma from vaccinated persons. PLoS One 2022; 17: e0261045.

#### Systematic review methods

*Search Strategy*

We searched Medline, Embase and MedRxiv between 1 September 2019 and 9 July 2024 for randomised controlled trials comparing the effects of neutralising monoclonal antibodies (mAbs) versus usual care or placebo in patients hospitalised with COVID-19. We used the search terms (Coronavirus infection or COVID or COVID-19 or COVID19 or 2019n-CoV or SARS-COV-2 or SARSCoV2 or SARS-Cov2) AND (sotrovimab, S309, VIR-7831, Xevudy, adintrevimab, amubarvimab, bamlanivimab, bebtelovimab, casirivimab, cilgavimab, etesevimab, imdevimab, regdanvimab, romlusevimab, tixagevimab) and using validated filters to select for randomised controlled trials. No language restrictions were applied.

Results were screened by researchers experienced in carrying out large-scale systematic reviews and meta-analyses of randomised trials. A trial research clinician reviewed the full texts of shortlisted studies to finalise the list of included studies. The following studies were identified:

| Trial | mAb(s) | published | start | end | Number of patients |
|---|---|---|---|---|---|
| RECOVERY[1] | casirivimab+imdevimab | 2021 | Sep-20 | May-21 | 9785 |
| ACTIV-3/TICO[2] | bamlanivimab | 2021 | Aug-20 | Oct-20 | 314 |
| J2W-MC-PYAA[3] | bamlanivimab | 2021 | May-20 | Jun-20 | 24 |
| ACTIV-3/TICO[4] | sotrovimab | 2022 | Dec-20 | Mar-21 | 365 |
| ACTIV-3/TICO[4] | BRII-196+BRII-198 | 2022 | Dec-20 | Mar-21 | 361 |
| CCCCTG[5] | etesevimab | 2022 | Jan-21 | Feb-21 | 197 |
| ACTIV-3/TICO[6] | tixagevimab+cilgavimab | 2022 | Feb-21 | Sep-21 | 1417 |
| COV-2066[7] | casirivimab+imdevimab | 2022 | Jun-20 | Apr-21 | 1197 |
| DisCoVeRy[8] | tixagevimab+cilgavimab | 2024 | Apr-21 | Jul-22 | 226 |

1 RECOVERY Collaborative Group. Casirivimab and imdevimab in patients admitted to hospital with COVID-19 (RECOVERY): a randomised, controlled, open-label, platform trial. *Lancet* 2022; **399**: 665–76.
2 ACTIV-3/TICO LY-CoV555 Study Group, Lundgren JD, Grund B, *et al.* A Neutralizing Monoclonal Antibody for Hospitalized Patients with Covid-19. *N Engl J Med* 2021; **384**: 905–14.
3 Chen P, Nirula A, Heller B, *et al.* SARS-CoV-2 Neutralizing Antibody LY-CoV555 in Outpatients with Covid-19. *N Engl J Med* 2021; **384**: 229–37.
4 ACTIV-3/Therapeutics for Inpatients with COVID-19 (TICO) Study Group. Efficacy and safety of two neutralising monoclonal antibody therapies, sotrovimab and BRII-196 plus BRII-198, for adults hospitalised with COVID-19 (TICO): a randomised controlled trial. *Lancet Infect Dis* 2022; **22**: 622–35.
5 Dong R, Jiang L, Yang T, *et al.* Efficacy and Safety of SARS-CoV-2 Neutralizing Antibody JS016 in Hospitalized Chinese Patients with COVID-19: a Phase 2/3, Multicenter, Randomized, Open-Label, Controlled Trial. *Antimicrob Agents Chemother* 2022; **66**: e0204521.
6 ACTIV-3–Therapeutics for Inpatients with COVID-19 (TICO) Study Group. Tixagevimab-cilgavimab for treatment of patients hospitalised with COVID-19: a randomised, double-blind, phase 3 trial. *Lancet Respir Med* 2022; **10**: 972–84.
7 Somersan-Karakaya S, Mylonakis E, Menon VP, *et al.* Casirivimab and Imdevimab for the Treatment of Hospitalized Patients With COVID-19. *J Infect Dis* 2022; **227**: 23–34.
8 Hites M, Massonnaud CR, Lapique EL, *et al.* Tixagevimab-cilgavimab (AZD7442) for the treatment of patients hospitalized with COVID-19 (DisCoVeRy): A phase 3, randomized, double-blind, placebo-controlled trial. *J Infect* 2024; **88**: 106120.

#### Protocol changes

RECOVERY is a randomised trial among patients hospitalized for COVID-19. All eligible patients receive usual standard of care in the participating hospital and are randomly allocated between no additional treatment and one of several active treatment arms. Over time, additional treatment arms have been added (see Table).

The final protocol relevant to high dose corticosteroids are included in the supplementary material to this publication, together with summaries of the changes made.

##### Table. Protocol changes to COVID-19 treatment comparisons

| Protocol version | Date | Randomisation | Treatment arms |
|---|---|---|---|
| 1.0 | 13-Mar-2020 | Main (part A) | No additional treatment<br>Lopinavir-ritonavir[a]<br>Low-dose corticosteroid[b]<br>Nebulised Interferon-ß-1a (never activated) |
| 2.0 | 23-Mar-2020 | Main (part A) | No additional treatment<br>Lopinavir-ritonavir[a]<br>Low-dose corticosteroid[b]<br>Hydroxychloroquine |
| 3.0 | 07-Apr-2020 | Main (part A) | No additional treatment<br>Lopinavir-ritonavir[a]<br>Low-dose corticosteroid[b]<br>Hydroxychloroquine[c]<br>Azithromycin[d] |
| 4.0 | 14-Apr-2020 | Main (part A)<br><br><br><br><br>Second[e,f] | No additional treatment<br>Lopinavir-ritonavir[a]<br>Low-dose corticosteroid[b]<br>Hydroxychloroquine[c]<br>Azithromycin[d]<br><br>No additional treatment<br>Tocilizumab[f] |
| 5.0 | 24-Apr-2020 | - | (no change – extension to children <18 years old) |
| 6.0 | 14-May-2020 | Main (part A)<br><br><br><br><br>Main (part B factorial)<br><br>Second[e,f] | No additional treatment<br>Lopinavir-ritonavir[a]<br>Low-dose corticosteroid[b]<br>Hydroxychloroquine[c]<br>Azithromycin[d]<br><br>No additional treatment<br>Convalescent plasma<br><br>No additional treatment<br>Tocilizumab[f] |
| 7.0 | 18-Jun-2020 | Main (part A)<br><br><br><br>Main (part B factorial)<br><br>Second[e,f] | No additional treatment<br>Lopinavir-ritonavir[a]<br>Low-dose corticosteroid[b]<br>Azithromycin[d]<br><br>No additional treatment<br>Convalescent plasma<br><br>No additional treatment<br>Tocilizumab[f] |

| Protocol version | Date | Randomisation | Treatment arms |
|---|---|---|---|
| 8.0 | 03-Jul-2020 | Main (part A) | No additional treatment<br>Low-dose corticosteroid[b]<br>Intravenous immunoglobulin[g]<br>High-dose corticosteroid[g]<br>Azithromycin[d] |
| | | Main (part B factorial) | No additional treatment<br>Convalescent plasma |
| | | Second[e,f] | No additional treatment<br>Tocilizumab[f] |
| 9.1 | 18-Sep-2020 | Main (part A) | No additional treatment<br>Low-dose corticosteroid[b]<br>Intravenous immunoglobulin[g]<br>High-dose corticosteroid[g]<br>Azithromycin[d] |
| | | Main (part B factorial) | No additional treatment<br>Convalescent plasma<br>Casirivimab and imdevimab |
| | | Second[e,f] | No additional treatment<br>Tocilizumab[f] |
| 10.1 | 01-Nov-2020 | Main (part A) | No additional treatment<br>Low-dose corticosteroid[b]<br>Intravenous immunoglobulin[g]<br>High-dose corticosteroid[g]<br>Azithromycin[d] |
| | | Main (part B factorial) | No additional treatment<br>Convalescent plasma<br>Casirivimab and imdevimab |
| | | Main (part C factorial) | No additional treatment<br>Aspirin |
| | | Second[e,f] | No additional treatment<br>Tocilizumab[f] |
| 11.1 | 27-Nov-2020 | Main (part A) | No additional treatment<br>Low-dose corticosteroid[b]<br>Intravenous immunoglobulin[g]<br>High-dose corticosteroid[g]<br>Colchicine |
| | | Main (part B factorial) | No additional treatment<br>Convalescent plasma<br>Casirivimab and imdevimab |
| | | Main (part C factorial) | No additional treatment<br>Aspirin |
| | | Second[e,f] | No additional treatment<br>Tocilizumab[f] |

| Protocol version | Date | Randomisation | Treatment arms |
|---|---|---|---|
| 12.1 | 16-Dec-2020 | Main (part A)[h] | No additional treatment<br>Low-dose corticosteroid[b]<br>Intravenous immunoglobulin[g]<br>High-dose corticosteroid[g]<br>Colchicine |
| | | Main (part B factorial)[h] | No additional treatment<br>Convalescent plasma<br>Casirivimab and imdevimab |
| | | Main (part C factorial)[h] | No additional treatment<br>Aspirin |
| | | Second[e,f] | No additional treatment<br>Tocilizumab[f] |
| 13.0 | 26-Jan-2021 | Main (part A)[h] | No additional treatment<br>Low-dose corticosteroid[b]<br>Intravenous immunoglobulin[g]<br>High-dose corticosteroid[g]<br>Colchicine |
| | | Main (part B factorial)[h] | No additional treatment<br>Casirivimab and imdevimab |
| | | Main (part C factorial)[h] | No additional treatment<br>Aspirin |
| | | Main (part D factorial) | No additional treatment<br>Baricitinib |
| | | Second[e,f] | No additional treatment<br>Tocilizumab[f]<br>Anakinra |
| 14.0 | 15-Feb-2021 | Main (part A)[h] | No additional treatment<br>Low-dose corticosteroid[b]<br>Intravenous immunoglobulin[g]<br>High-dose corticosteroid[g]<br>Colchicine<br>Dimethyl fumarate |
| | | Main (part B factorial)[h] | No additional treatment<br>Casirivimab and imdevimab |
| | | Main (part C factorial)[h] | No additional treatment<br>Aspirin |
| | | Main (part D factorial) | No additional treatment<br>Baricitinib |
| | | Second[e,f] | No additional treatment<br>Tocilizumab[f]<br>Anakinra |

| Protocol version | Date | Randomisation | Treatment arms |
|---|---|---|---|
| 15.0 | 12-Apr-2021 | Main (part A)[h] | No additional treatment<br>Low-dose corticosteroid[b]<br>Intravenous immunoglobulin[g]<br>High-dose corticosteroid[g]<br>Dimethyl fumarate |
| | | Main (part B factorial)[h] | No additional treatment<br>Casirivimab and imdevimab |
| | | Main (part D factorial) | No additional treatment<br>Baricitinib<br>Infliximab[j] |
| | | Main (part E factorial)[i] | High-dose dexamethasone[j] |
| | | Second[e,f] | No additional treatment<br>Tocilizumab[f]<br>Anakinra |
| 16.1 | 08-Jul-2021 | Main (part A)[h] | No additional treatment<br>Low-dose corticosteroid[b]<br>Intravenous immunoglobulin[g]<br>High-dose corticosteroid[g]<br>Dimethyl fumarate |
| | | Main (part D factorial) | No additional treatment<br>Baricitinib |
| | | Main (part E factorial)[i] | High-dose dexamethasone[j] |
| | | Main (part F factorial)[l] | Empagliflozin[l] |
| | | Second[e,f] | No additional treatment<br>Tocilizumab[f]<br>Anakinra |
| 17.1 | 10-Aug-2021 | Main (part A)[h] | No additional treatment<br>Dimethyl fumarate |
| | | Main (part D factorial) | No additional treatment<br>Baricitinib |
| | | Main (part E factorial)[i] | High-dose dexamethasone[j] |
| | | Main (part F factorial)[l] | Empagliflozin [l] |
| | | Second[e,f] | No additional treatment<br>Tocilizumab[f]<br>Anakinra |

| Protocol version | Date | Randomisation | Treatment arms |
|---|---|---|---|
| 18.1 | 24-Oct-2021 | Main (part A)[h]<br><br>Main (part D factorial)<br><br>Main (part E factorial)[i]<br><br>Main (part F factorial)[l]<br><br>Second[e,f] | No additional treatment<br>Dimethyl fumarate<br><br>No additional treatment<br>Baricitinib<br><br>High-dose dexamethasone[j]<br><br>Empagliflozin[l]<br><br>No additional treatment<br>Tocilizumab[f]<br>Anakinra |
| 19.1 | 16-Nov-2021 | Main (part D factorial)[k]<br><br>Main (part E factorial)[i]<br><br>Main (part F factorial)[l]<br><br>Second[e,f,h] | No additional treatment<br>Baricitinib<br><br>High-dose dexamethasone[j]<br><br>Empagliflozin[l]<br><br>No additional treatment<br>Tocilizumab[f]<br>Anakinra |
| 20.0 | 29-Nov-2021 | Main (part E factorial)[i]<br><br>Main (part F factorial)[l]<br><br>Second[e,f,h] | High-dose dexamethasone[j]<br><br>Empagliflozin[l]<br><br>No additional treatment<br>Tocilizumab[f]<br>Anakinra |
| 21.0 | 17-Dec-2021 | Main (part E factorial)[i]<br><br>Main (part F factorial)[l]<br><br>Main (part J factorial)<br><br>Main (part K factorial)<br><br>Second[e,f,h] | High-dose dexamethasone[j]<br><br>Empagliflozin[l]<br><br>Sotrovimab<br><br>Molnupiravir<br><br>No additional treatment<br>Tocilizumab[f]<br>Anakinra |
| 22.0 | 19-Jan-2022 | Not implemented | |
| 23.0 | 08-Mar-2022 | Main (part E factorial)[i]<br><br>Main (part F factorial)[l]<br><br>Main (part J factorial)<br><br>Main (part K factorial)<br><br>Main (part L factorial) | High-dose dexamethasone[j]<br><br>Empagliflozin[l]<br><br>Sotrovimab<br><br>Molnupiravir<br><br>Nirmatrelvir-ritonavir |
| 24.0 | 13-May-2022 | Not implemented | |

| **Protocol version** | **Date** | **Randomisation** | **Treatment arms** |
|---|---|---|---|
| 25.0 | 23-May-2022 | Main (part E factorial)[i]<br><br>Main (part F factorial)[l]<br><br>Main (part J factorial)<br><br>Main (part K factorial)<br><br>Main (part L factorial) | High-dose dexamethasone[j]<br><br>Empagliflozin[l]<br><br>Sotrovimab<br><br>Molnupiravir[m]<br><br>Nirmatrelvir-ritonavir[m] |
| 26.0 | 22-Jun-2023 | Main (part E factorial)[i]<br><br>Main (part J factorial) | High-dose dexamethasone[j]<br><br>Sotrovimab |
| 27.0 | 13-Sep-2023 | Main (part E factorial)[i]<br><br>Main (part J factorial) | High-dose dexamethasone[j,n]<br><br>Sotrovimab[n] |

[a] enrolment ceased 29 June 2020 when the Data Monitoring Committee advised that the Chief Investigators should review the unblinded data.
[b] enrolment of adults ceased 8 June 2020 as more than 2,000 patients had been recruited to the active arm. Enrolment of children ceased on 8 July 2021.
[c] enrolment ceased 5 June 2020 when the Data Monitoring Committee advised that the Chief Investigators should review the unblinded data.
[d] enrolment of adults ceased 27 November 2020 as more than 2,500 patients had been recruited to the active arm
[e] for patients with (a) oxygen saturation <92% on air or requiring oxygen or children with significant systemic disease with persistent pyrexia; and (b) C-reactive protein ≥75 md/L)
[f] enrolment of adults ceased 24 January 2021 as more than 2,000 patients had been recruited to the active arm.
[g] for children only. Enrolment ceased 8 July 2021.
[h] from protocol version 12.1, children could enter the second randomisation regardless of whether they were included in the main randomisation. Enrolment ceased 8 March 2022.
[i] for patients with (a) oxygen saturation <92% on air or requiring oxygen. Enrolment of patients receiving no or simple oxygen ceased on 13 May 2022.
[j] for patients outside UK (until protocol V20.0 when extended to UK)
[k] enrolment ceased 29 December 2021
[l] enrolment ceased 6 March 2023
[m] enrolment ceased 24 May 2023
[n] enrolment ceased 31 March 2024

**Main and second randomisation <u>for adults</u>**

All RECOVERY trial participants received usual standard of care. On study entry, adult participants initially underwent the Main Randomisation. Trial participants with clinical evidence of progressive COVID-19 (defined as oxygen saturation <92% on room air or requiring oxygen therapy, and C-reactive protein ≥75 mg/L) could be considered for the Second Randomisation at any time up to 21 days after the initial randomisation, and regardless of initial treatment allocation(s). A web-system was used to provide simple randomisation (without stratification or minimisation) with allocation concealment until randomisation had been completed.

Over time, treatment arms were added and removed from the protocol, factorial randomisations were introduced (see below), and not all treatments were available at every hospital. Similarly, not all treatments were deemed by the attending clinician to be suitable for some patients (e.g. due to comorbid conditions or concomitant medication). In any of these cases, randomisation involved fewer arms (and/or fewer factorial elements).

*Main randomisation for adults*

A single participant could be randomised at most to 1 arm from each of part A, B, C, D and E of the factorial randomisations (depending on location), and thus receive between 0 and 4 treatments on top of usual standard of care.

**<u>Part A</u>** (from 19 March 2020)

Eligible participants could be randomised to one of the following arms:

| Treatment arm | Arm opened | Arm closed |
|---|---|---|
| No additional treatment | 19 March 2020 | 12 November 2021 |
| Dexamethasone | 19 March 2020 | 8 June 2020 |
| Lopinavir-ritonavir | 19 March 2020 | 29 June 2020 |
| Hydroxychloroquine | 23 March 2020 | 5 June 2020 |
| Azithromycin | 7 April 2020 | 27 November 2020 |
| Colchicine | 27 November 2020 | 5 March 2021 |
| Dimethyl fumarate | 15 February 2021 | 12 November 2021 |

**<u>Part B</u>** (from 14 May 2020)

Eligible participants could be randomised to one of the following arms:

| Treatment arm | Arm opened | Arm closed |
|---|---|---|
| No additional treatment | 14 May 2020 | 21 May 2021 |
| Convalescent plasma | 14 May 2020 | 15 January 2021 |
| Casirivimab and imdevimab * | 18 September 2020 | 21 May 2021 |

* monoclonal neutralising antibody cocktail

**<u>Part C</u>** (from 1 November 2020)

Eligible participants could be randomised to one of the following arms:

| Treatment arm | Arm opened | Arm closed |
|---|---|---|
| No additional treatment | 1 November 2020 | 21 March 2021 |
| Aspirin | 1 November 2020 | 21 March 2021 |

**Part D** (from 1 November 2020)

Eligible participants could be randomised to one of the following arms (UK only):

| Treatment arm | Arm opened | Arm closed |
|---|---|---|
| No additional treatment | 2 February 2021 | 29 December 2021 |
| Baricitinib | 2 February 2021 | 29 December 2021 |

**Part E** (from 25 May 2021)

Eligible participants could be randomised to one of the following arms:

| Treatment arm | Arm opened | Arm closed |
|---|---|---|
| No additional treatment | 25 May 2021 | 31 March 2024 |
| High-dose dexamethasone | 25 May 2021 | 31 March 2024 (Enrolment of patients receiving no or simple oxygen ceased on 13 May 2022) |

**Part F** (from 8 July 2021)

Eligible participants could be randomised to one of the following arms:

| Treatment arm | Arm opened | Arm closed |
|---|---|---|
| No additional treatment | 8 July 2021 | 6 March 2023 |
| Empagliflozin | 8 July 2021 | 6 March 2023 |

**Part J** (from 29 December 2021)

Eligible participants could be randomised to one of the following arms (UK only):

| Treatment arm | Arm opened | Arm closed |
|---|---|---|
| No additional treatment | 30 December 2021 | 31 March 2024 |
| Sotrovimab | 30 December 2021 | 31 March 2024 |

**Part K** (from 30 December 2021)

Eligible participants could be randomised to one of the following arms:

| Treatment arm | Arm opened | Arm closed |
|---|---|---|
| No additional treatment | 30 December 2021 | 24 May 2023 |
| Molnupiravir | 30 December 2021 | 24 May 2023 |

**Part L** (from 8 March 2022)

Eligible participants could be randomised to one of the following arms (UK only):

| Treatment arm | Arm opened | Arm closed |
|---|---|---|
| No additional treatment | 8 March 2022 | 24 May 2023 |
| Nirmatrelvir-ritonavir | 8 March 2022 | 24 May 2023 |

*Second randomisation for adults (from 14 April 2020)*

From 14 April 2020, a participant could be randomised to one of the following arms and thus receive 0 or 1 treatment on top of those allocated in the initial randomisation and usual standard of care:

| Treatment arm | Arm opened | Arm closed |
|---|---|---|
| No additional treatment | 14 April 2020 | 24 January 2021 |
| Tocilizumab | 14 April 2020 | 24 January 2021 |

**Ascertainment and classification of study outcomes**

Information on baseline characteristics and study outcomes was collected through a combination of electronic case report forms (see below) completed by members of the local research team at each participating hospital and (in the UK) linkage to National Health Service, clinical audit, and other relevant health records. Full details are provided in the RECOVERY Definition and Derivation of Baseline Characteristics and Outcomes Document (see Appendix 3).

*Randomisation form*

The (main) Randomisation form (shown below) was completed by trained study staff. It collected baseline information about the participant (including demographics, COVID-19 history, comorbidities and suitability for the study treatments) and availability of the study treatments. Once completed and electronically signed, the treatment allocation was displayed.

The following modifications were made to the Randomisation form during the trial:

| Randomisation form version | Date of release | Major modifications from previous version |
|---|---|---|
| 1.0 | 19-Mar-20 | Initial version (protocol V1.0) |
| 2.0 | 25-Mar-20 | For protocol V2.0<br>• Hydroxycholoroquine added as treatment<br>• Known long QT syndrome added to comorbidities<br>• Severe depression removed from comorbidities |
| 3.0 | 09-Apr-20 | For protocol V3.0<br>• Azithromycin added as treatment<br>• Suspected SARS-CoV-2 infection included in eligibility criteria |
| [Second randomisation form introduced] | 23-Apr-20 | For protocol 4.0<br>• Eligibility criteria for second randomisation<br>• Tocilizumab vs control as treatment allocations |
| 5.0 | 09-May-20 | For protocol V5.0<br>• Age ≥18 years removed from eligibility criteria<br>• Additional questions on child's age and weight added |
| 6.0 | 21-May-20 | For protocol V6.0<br>• Convalescent plasma added as treatment<br>• Baseline use of remdesivir |
| 7.0 | 01-Jul-20 | For protocol V7.0<br>• Participants eligible if convalescent plasma is only available and suitable treatment |
| 8.0 | 13-Aug-20 | For protocol V8.0<br>• Addition of low-dose and high-dose corticosteroids and intravenous immunoglobulin for children (and removal of dexamethasone for children) |
| 9.0 | 24-Sep-20 | For protocol V9.0<br>• Casirivimab and imdevimab added as treatment<br>• Additional baseline information |
| 10.0 | 06-Nov-20 | For protocol V10.1<br>• Aspirin added as treatment |
| 11.0 | 27-Nov-20 | For protocol V11.1<br>• Colchicine added as treatment |
| 12.0 | 22-Dec-20 | For protocol V12.1<br>• Allow children to enter trial without entering main randomisation |
| 13.0 | 02-Feb-21 | For protocol V13.0<br>• Baricitinib added as treatment |
| 14.0 | 24-Feb-21 | For protocol V14.0<br>• Dimethyl fumarate added as treatment |
| 15.0 | 11-May-21 | For protocol V15.0<br>• High-dose dexamethasone added as treatment |
| 16.0 | 28-Jul-21 | For protocol V16.1<br>• Addition of empagliflozin as treatment |
| 17.0 | 20-Aug-21 | For protocol V17.1<br>• Additional warnings about eligibility for empagliflozin |

| **Randomisation form version** | **Date of release** | **Major modifications from previous version** |
|---|---|---|
| 18.0 | 30-Dec-21 | For protocol V21.1<br>• Sotrovimab and molnupiravir added as treatments<br>• Inclusion of UK participants in high-dose dexamethasone comparison |
| 19.0 | 28-Mar-22 | For protocol V23.0<br>• Nirmatrelvir-ritonavir added as treatment |
| 20.0 | 8-Jan-2024 | For protocol V27.0<br>• Addition of non-COVID-19 community-acquired pneumonia comparison |

RECOVERY
Randomised Evaluation of COVID-19 Therapy

### Sample Form (v19.00 - 28/03/22)

#### Randomisation Program

Call Freefone **0800 138 5451** to contact the RECOVERY team for **URGENT** problems using the Randomisation Program or for medical advice. All **NON-URGENT queries** should be emailed to ****

Logged in as: **RECOVERY Site**

##### Section A: Baseline and Eligibility

Date and time of randomisation: 27 Mar 2022 14:00

| | |
|---|---|
| **Treating clinician** | |
| **A1.** Name of treating clinician | |
| **Patient details** | |
| **A2.** Patient surname | |
| Patient forename | |
| **A3.** NHS number | ☐ Tick if not available |
| **A4.** What is the patient's date of birth? | 01 / January / 2000 Age: 22y 2m |
| **A5.** What is the patient's sex? | |
| **Inclusion criteria** | |
| **A6.** Has consent been taken in line with the protocol?<br>If answer is No patient cannot be enrolled in the study<br>NB current PIS/ICF version is V22.0 (adults) or V14.0 (children) | |
| **A6.0.1** How was consent obtained? | |
| **A6.5** Does this patient have viral pneumonia?<br>See protocol for typical features. If answer is No patient cannot be enrolled in the study | Yes |
| **A7.0** Does the patient have proven SARS-CoV-2 infection? | Yes |
| **A7.0.1** What was lateral flow test result? | |
| **A7.0.2** What was PCR test result? | |
| **A7.1** Does the patient have proven influenza infection? | |
| **A8.** Does the patient have any medical history that might, in the opinion of the attending clinician, put the patient at significant risk if they were to participate in the trial? | |
| **A9.** Symptom onset date: | / / |
| **A10.** Date of hospitalisation: | / / |
| **A11.** Does the patient require oxygen? | |
| **A12.** Please select one of the following to describe the current level of ventilation support | |
| **A12.1** Enter latest oxygen saturation measurement (%) | |
| **A12.2** Enter latest CRP measurement since admission to hospital (mg/L)<br>Enter 0 if below the limit of measurement | ☐ Tick if not measured<br>☐ Tick if greater than limit of measurement |
| **A12.3** Enter latest creatinine measurement since admission to hospital | μmol/L ☐ Tick if not measured |
| **A12.4** Enter latest D-dimer measurement since admission to hospital<br>Enter 0 if below the limit of measurement | ng/mL ☐ Tick if not measured<br>☐ Tick if greater than limit of measurement |
| **A12.5** Has the patient received a COVID-19 vaccine? | |
| **A12.6** Has the patient received an influenza vaccine in the last 12 months? | |
| **Does the patient have any CURRENT comorbidities or other medical problems or treatments?** | |
| **A13.1** Diabetes | |
| **A13.2** Heart disease | |
| **A13.3** Chronic lung disease | |
| **A13.4** Tuberculosis | |
| **A13.5** HIV | |
| **A13.6** Severe liver disease | |
| **A13.7** Severe kidney impairment (eGFR<30 or on dialysis) | |
| **A13.9.0** Does their clinician consider the patient to be severely immunocompromised? | |
| **A13.12** Has the patient received tocilizumab or sarilumab therapy during this admission? | |
| **A13.14** Current or planned treatment with neuraminidase inhibitor<br>eg, oseltamivir, zanamivir | |
| **A13.15** Has the patient received casirivimab+imdevimab (Ronapreve) during this illness? | |
| **A13.16** Has the patient received sotrovimab during this illness? | |
| **A13.17** Has the patient received molnupiravir during this illness? | |
| **A13.18** Has the patient received Paxlovid during this illness? | |
| **Are the following treatments UNSUITABLE for the patient?**<br>**If you answer Yes it means you think this patient should NOT receive this drug.** | |
| **A14E.1** High-dose corticosteroids<br>NB Please carefully consider suitability of patients already on higher doses (>=7 mg/day dexamethasone or equivalent). Patients eligible for the Paxlovid comparison will be automatically marked as unsuitable for this comparison. | |
| **A14F.1** Empagliflozin<br>Empagliflozin is NOT suitable if patient (i) has type 1 or post-pancreatectomy diabetes mellitus; or (ii) has a history of ketoacidosis; or (iii) has blood ketones ≥1.5 mmol/L or urine ketones ≥2+; or (iv) is pregnant or breastfeeding Empagliflozin cannot be given via an enteral feeding tube. | |
| **A14J.1** Sotrovimab | |
| **A14K.1** Molnupiravir<br>NB Molnupiravir is NOT suitable if patient cannot swallow capsules. | |
| **A14L.1** Paxlovid<br>NB Paxlovid contains ritonavir and has many drug-drug interactions (see protocol and SmPC). Please ensure these have been checked. Paxlovid is NOT suitable if patient cannot swallow tablets. Paxlovid is not suitable for pregnant women in the first trimester. | No |
| **Are the following treatments available?** | |
| **A15E.1** High-dose corticosteroids | |
| **A15F.1** Empagliflozin | |
| **A15J.1** Sotrovimab | |
| **A15K.1** Molnupiravir | |
| **A15L.1** Paxlovid | Yes |
| **Current medication** | |
| **A16.1** Is the patient currently prescribed remdesivir? | |
| **A16.2** Is the patient currently prescribed systemic corticosteroids (dexamethasone, prednisolone, hydrocortisone, methylprednisolone)?<br>Please do not include topical or inhaled treatments | |
| **A16.5** What venous thromboembolism prophylaxis is the patient receiving?<br>Standard = usual for hospitalised patients (not increased due to COVID-19); Higher dose = treatment dose or increased prophylaxis due to COVID-19, or oral anticoagulation (eg, warfarin/DOAC). | |
| **A16.6** Is the patient currently prescribed baricitinib (or other JAK inhibitor)? | |
| **Serum sample collection** | |
| **A17.0** Please confirm that patient has had a baseline serum sample collected according to the protocol | |
| **A17.1** Please confirm that patient has had a baseline nasal swab collected according to the protocol | |
| **Please sign off this form once complete** | |
| Surname: | |
| Forename: | |
| Professional email: | |
| | **Continue** |
| | **Cancel** |

**Home**

*Follow-up form*

The Follow-up form (shown on the next page) collected information on study treatment adherence (including both the randomised allocation and use of other study treatments), vital status (including date and provisional cause of death if available), hospitalisation status (including date of discharge), respiratory support received during the hospitalisation, occurrence of any major cardiac arrhythmias and renal replacement therapy received. Questions on metabolic complications were added in version 16.0, after the high-dose corticosteroid comparison started.

The following modifications were made to the Follow-up form during the trial:

| Follow-up form version | Date of release | Modifications from previous version |
|---|---|---|
| 18.0 | 30-Dec-21 | Information on sotrovimab and molnupiravir adherence<br>Information on sample collection |
| 19.0 | 25-Feb-22 | Information on nirmatrelvir-ritonavir adherence |
| 20.0 | 28-Mar-22 | Information on liver function tests and seizures |
| 21.0 | 31-Mar-22 | Translations added for V19.0 and V20.0 |
| 22.0 | 8-Nov-22 | For protocol V22 |
| 23.0 | 9-Jan-23 | For protocol V23 |
| 24.0 | 17-Nov-23 | Removal of closed comparisons |
| 25.0 | 8-Jan-24 | Addition of non-COVID-19 CAP arm |
| 26.0 | 26-Feb-24 | Translations added |

### Follow-up

#### Date of randomisation

**Please only report events that occurred from first randomisation until 28 days later on this form (except for Q2).**

**Patient's date of birth** *

yyyy-mm-dd

##### » Vital Status

**0. What is the patient's vital status?** *

- ◯ Alive
- ◯ Dead

**0.1 What is the patient's current hospitalisation status?** *

- ◯ Inpatient
- ◯ Discharged

The patient has been enrolled in the trial for **NaN** days

**0.1.1 Date follow-up form completed**

yyyy-mm-dd

**0.1.1 What was the date of discharge?** *

yyyy-mm-dd

**0.1 What was the date of death?** *

yyyy-mm-dd

**0.2 What was the underlying cause of death?** *

*This can be obtained from the last entry in part 1 of the death certificate*

- ◯ COVID-19
- ◯ Other infection
- ◯ Cardiovascular
- ◯ Other

**Please give details**

##### » Treatments

**1. Which of the following treatment(s) did the patient definitely receive as part of their hospital admission after randomisation?** *

*(NB Include RECOVERY study-allocated drug, only if given, PLUS any of the other treatments if given as standard hospital care)*

- ☐ No additional treatment
- ☐ Lopinavir-ritonavir

- [ ] Corticosteroid (dexamethasone, prednisolone, hydrocortisone or methylprednisolone)
- [ ] Hydroxychloroquine
- [ ] Azithromycin or other macrolide (eg, clarithromycin, erythromycin)
- [ ] Tocilizumab or sarilumab
- [ ] Remdesivir
- [ ] Intravenous immunoglobulin
- [ ] Synthetic monoclonal antibodies (REGN10933+REGN10987)
- [ ] Aspirin
- [ ] Colchicine
- [ ] Baricitinib
- [ ] Anakinra
- [ ] Favipiravir
- [ ] Empagliflozin
- [ ] Ivermectin
- [ ] Oseltamivir
- [ ] Other neuraminidase inhibitor (e.g. zanamivir, laninamivir)
- [ ] Baloxavir
- [ ] Sotrovimab
- [ ] Molnupiravir
- [ ] Paxlovid

**Please select number of days the patient received corticosteroid (dexamethasone, prednisolone, hydrocortisone or methylprednisolone) (of any dose)**

○ 1 ○ 2 ○ 3 ○ 4 ○ 5 ○ 6 ○ 7 ○ 8 ○ 9 ○ 10

**Dosing information:**

**6 mg dexamethasone is equivalent to 40 mg prednisolone or 160 mg hydrocortisone or 32 mg methylprednisolone.**

**10 mg dexamethasone is equivalent to 67 mg prednisolone or 267 mg hydrocortisone or 53 mg methylprednisolone**

**20 mg dexamethasone is equivalent to 133 mg prednisolone or 534 mg hydrocortisone or 106 mg methylprednisolone**

**Please indicate the highest dose received on a single day during the 10 days after randomisation**

○ <6 mg dexamethasone
○ 6 mg dexamethasone
○ >6 mg and <=10 mg dexamethasone
○ >10 mg and <20 mg dexamethasone
○ 20 mg dexamethasone
○ >20 mg dexamethasone

**Please select number of doses of tocilizumab or sarilumab the patient received**

○ 1 ○ >1

**Please select number of days the patient received remdesivir**

○ 1 ○ 2 ○ 3 ○ 4 ○ 5 ○ 6 ○ 7 ○ 8 ○ 9 ○ 10

**Please select number of days the patient received baricitinib**

( ) 1 ( ) 2 ( ) 3 ( ) 4 ( ) 5 ( ) 6 ( ) 7 ( ) 8 ( ) 9 ( ) 10

**Please select number of days the patient received anakinra**

( ) 1 ( ) 2 ( ) 3 ( ) 4 ( ) 5 ( ) 6 ( ) 7

**Please select the proportion of days the patient received empagliflozin during the first 28 days after randomisation (or from randomisation to date of discharge if this is sooner)**

( ) Most days (≥90%) ( ) Some days (≥50% <90%) ( ) Few days (<50% of days, but not zero) ( ) None

**Please select number of days the patient received oseltamivir**

( ) 1 ( ) 2 ( ) 3 ( ) 4 ( ) 5 ( ) 6 ( ) 7 ( ) 8 ( ) 9 ( ) 10

**Please select number of doses of baloxavir the patient received**

( ) 1 ( ) 2

**Did the participant experience an infusion reaction during or within 2 hours after the sotrovimab infusion?** *

( ) Yes

( ) No

**How severe was the reaction?** *

( ) Mild (no intervention required)

( ) Moderate (eg, antihistamines or steroids required)

( ) Severe (adrenaline required)

**Was the infusion completed?** *

( ) Yes

( ) No

**Please select the number of days the patient received molnupiravir**

( ) 1 ( ) 2 ( ) 3 ( ) 4 ( ) 5 ( ) 6

**Was the participant provided with treatment to complete the course at home?**

( ) Yes

( ) No

**Please select the number of days the patient received Paxlovid**

( ) 1 ( ) 2 ( ) 3 ( ) 4 ( ) 5 ( ) 6

**Was the participant provided with treatment to complete the course at home?**

( ) Yes

( ) No

**Only required if Q17.0 and or Q17.1 on the Randomisation form were answered Yes**

**Was the baseline serum sample collected?**

( ) Yes

No

Was the baseline swab samples collected? *

Yes

No

Was the DAY 3 follow-up swab sample collected? *

Yes

No

Swab sent home with patient

Was the DAY 5 follow-up swab sample collected? *

Yes

No

Swab sent home with patient

#### » Ventilation

**4. Did the patient require any form of assisted ventilation (ie, more than just supplementary oxygen) from day of randomisation until 28 days later?** *

Yes

No

**Please answer the following questions:**

**4.1 For how many days did the patient require assisted ventilation?** *

**4.2 What type of ventilation did the patient receive?**

| | Yes | No | Unknown |
|---|---|---|---|
| CPAP alone | | | |
| Non-invasive ventilation (eg, BiPAP) | | | |
| High-flow nasal oxygen (eg, AIRVO) | | | |
| Mechanical ventilation (intubation/tracheostomy) | | | |
| ECMO | | | |

Total number of days the patient received invasive mechanical ventilation (intubation/tracheostomy) from randomisation until discharge/death/28 days after randomisation

#### » Cardiac arrhythmia

**5. Has the patient been documented to have a NEW cardiac arrhythmia at any point since the main randomisation until 28 days later?** *

- ( ) Yes
- ( ) No
- ( ) Unknown

**5.1 Please select all of the following which apply**

- [ ] Atrial flutter or atrial fibrillation
- [ ] Supraventricular tachycardia
- [ ] Ventricular tachycardia (including torsades de pointes)
- [ ] Ventricular fibrillation
- [ ] Atrioventricular block requiring intervention (eg, cardiac pacing)

**» Renal outcomes**

**6. Did the patient require use of renal dialysis or haemofiltration from main randomisation until 28 days later?** *

- ( ) Yes
- ( ) No

| 6.1 Please enter the highest creatinine level recorded after randomisation until 28 days later. * | Unit * | Date recorded * | Select if creatinine level not available * |
|---|---|---|---|
| | ( ) µmol/L<br>( ) mg/dL | yyyy-mm-dd | ( ) Not available |

**» Thrombosis and bleeding**

**7. During the first 28 days after randomisation (or until discharge if sooner), did the participant have a thrombotic event?** *

- ( ) Yes
- ( ) No
- ( ) Unknown

**7.1 Please indicate the type of thrombotic event**

*Select all that apply*

- [ ] Pulmonary embolism
- [ ] Deep-vein thrombosis
- [ ] Ischaemic stroke
- [ ] Myocardial infarction
- [ ] Systemic arterial embolism
- [ ] Other

**8. During the first 28 days after randomisation (or until discharge if sooner), did the participant experience clinically-significant bleeding ie, intra-cranial bleeding or bleeding that required intervention (eg, surgery, endoscopy or vasoactive drugs) or a blood transfusion?** *

- ( ) Yes
- ( ) No

( ) Unknown

**8.1 Please indicate the site(s) of bleeding** *

*Select all that apply*

- [ ] Intra-cranial
- [ ] Gastrointestinal
- [ ] Other

**8.2 Please indicate which interventions were required to manage the bleed** *

*Select all that apply*

- [ ] Blood transfusion
- [ ] Surgery
- [ ] Endoscopy
- [ ] Vasoactive drugs (e.g. inotropes on ICU)
- [ ] None of the above

#### » Other infections

**9. During the first 28 days after randomisation (or until discharge if sooner), did the participant develop another infection?** *

( ) Yes

( ) No

( ) Unknown

**9.1 Please indicate the type of infection**

*Select all that apply*

- [ ] Pneumonia
- [ ] Urinary tract
- [ ] Biliary
- [ ] Other intra-abdominal
- [ ] Blood stream
- [ ] Skin
- [ ] Other

**Pneumonia - please indicate the putative organism**

( ) Bacterial ( ) Fungal ( ) Viral ( ) Other ( ) Unknown

**Please indicate the virus**

*NB do not record the virus leading to study entry*

[ ] SARS-CoV-2 [ ] Influenza [ ] Other/unknown

**Urinary tract - please indicate the putative organism**

( ) Bacterial ( ) Fungal ( ) Other ( ) Unknown

**Biliary - please indicate the putative organism**

Biliary - please indicate the putative organism

( ) Bacterial ( ) Fungal ( ) Other ( ) Unknown

Intra-abdominal - please indicate the putative organism

( ) Bacterial ( ) Fungal ( ) Other ( ) Unknown

Blood stream - please indicate the putative organism

*Please only select this if positive blood culture but no known anatomical site found*

( ) Bacterial ( ) Fungal ( ) Other ( ) Unknown

Skin - please indicate the putative organism

( ) Bacterial ( ) Fungal ( ) Viral ( ) Other ( ) Unknown

| Other - please indicate the putative organism | Please describe the anatomical site |
|---|---|
| ( ) Bacterial ( ) Fungal ( ) Other ( ) Unknown | |

#### » Metabolic complications

**10. During the first 28 days after randomisation (or until discharge if sooner), did the participant have any of the following?**

| | Yes | No | Unknown |
|---|---|---|---|
| **Ketoacidosis** * *Ketoacidosis is defined as (i) ketosis (blood ketones ≥1.5 mmol/L or urine ketones ≥2+) AND (ii) metabolic acidosis (eg, bicarbonate <15 mmol/L) AND (iii) no obvious alternative cause of acidosis* | ( ) | ( ) | ( ) |
| **Hyperglycaemic hyperosmolar state** * | ( ) | ( ) | ( ) |
| **Other hyperglycaemia requiring new use of insulin** * | ( ) | ( ) | ( ) |
| **Severe hypoglycaemia** * *Hypoglycaemia causing reduced conscious level requiring another person to help recover.* | ( ) | ( ) | ( ) |

#### » Other safety outcomes

**11. Did the participant experience a seizure after randomisation?** *

( ) Yes

( ) No

( ) Unknown

11.1 Does the patient have a history of seizures or epilepsy?

( ) Yes

( ) No

( ) Unknown

**11.2 Please enter the highest ALT (or AST) level recorded after randomisation until 28 days later. If below the limit of detection, enter 0**

| Date * | Result * | Upper limit of normal * | Units |
|---|---|---|---|

| | | | |
|---|---|---|---|
| yyyy-mm-dd | | | (●) IU/L or U/L<br>( ) µmol/L<br>( ) µkat/L |

**11.3 Please enter the highest bilirubin level recorded after randomisation until 28 days later. If below the limit of detection, enter 0**

| Date * | Result * | Upper limit of normal * | Units |
|---|---|---|---|
| yyyy-mm-dd | | | (●) µmol/L<br>( ) mg/dL |

#### » Other trials

**12. Please indicate if the participant participated in any other COVID-19 or influenza trials**

*Select all that apply*

- ☐ PRINCIPLE
- ☐ REMAP-CAP
- ☐ Other treatment trial(s)
- ☐ COVID-19 vaccine trial(s)

**Please give name of other treatment trial(s)**

**Please give name of COVID-19 vaccine trial(s)**

#### » Pregnancy

**13. If this woman was pregnant at randomisation (or had recently delivered), please enter UKOSS ID here.**

*Enter the full UKOSS case ID eg, COR_123*

**Interim analyses: role of the Data Monitoring Committee**

The independent Data Monitoring Committee reviewed unblinded analyses of the study data and any other information considered relevant at intervals of around 2-3 months. The committee was charged with determining if, in their view, the randomised comparisons in the study provide evidence on mortality that is strong enough (with a range of uncertainty around the results that was narrow enough) to affect national and global treatment strategies. In such a circumstance, the Committee would inform the Steering Committee who would make the results available to the public and amend the trial arms accordingly. Unless that happened, the Steering Committee, investigators, and all others involved in the trial would remain blind to the interim results until 28 days after the last patient had been randomised to a particular intervention arm. Further details about the role and membership of the independent Data Monitoring Committee are provided in the protocol.

The Data Monitoring Committee determined that to consider recommending stopping a treatment early for benefit would require at least a 3 to 3·5 standard error reduction in mortality. The Committee concluded that examinations of the data at every 10% (or even 5%) of the total data would lead to only a marginal increase in the overall type I error rate.

Between 4 January 2022 and 31 March 2024 the DMC reviewed data on sotrovimab 9 times (https://www.recoverytrial.net/uk/for-site-staff/site-set-up-1/data-monitoring-committee-correspondence).

### Supplementary Tables

**Webtable 1: Baseline characteristics of patients considered unsuitable for randomisation to sotrovimab compared with those randomised to sotrovimab versus usual care**

| | **Randomised (n=1723)** | **Unsuitable (n=65)** |
|---|---|---|
| Age, years | 70.7 (14.8) | 69.3 (13.8) |
| <70 | 711 (41%) | 33 (51%) |
| ≥70 to <80 | 523 (30%) | 14 (22%) |
| ≥80 | 489 (28%) | 18 (28%) |
| Sex | | |
| Male | 1033 (60%) | 36 (55%) |
| Female | 690 (40%) | 29 (45%) |
| Ethnicity | | |
| White | 1485 (86%) | 55 (85%) |
| Black, Asian, and minority ethnic | 129 (7%) | 4 (6%) |
| Unknown | 109 (6%) | 6 (9%) |
| Number of days since symptom onset | 6 (3-11) | 6 (3-11) |
| Number of days since admission to hospital | 2 (1-5) | 2 (1-6) |
| Respiratory support received | | |
| None | 256 (15%) | 13 (20%) |
| Simple oxygen | 1069 (62%) | 43 (66%) |
| Non-invasive ventilation | 337 (20%) | 8 (12%) |
| Invasive mechanical ventilation | 61 (4%) | 1 (2%) |
| Previous diseases | | |
| Diabetes | 468 (27%) | 17 (26%) |
| Heart disease | 531 (31%) | 22 (34%) |
| Chronic lung disease | 652 (38%) | 24 (37%) |
| Tuberculosis | 6 (<0.5%) | 0 (0%) |
| HIV | 11 (1%) | 1 (2%) |
| Severe liver disease * | 35 (2%) | 0 (0%) |
| Severe kidney impairment † | 158 (9%) | 5 (8%) |
| Any of the above | 1180 (68%) | 45 (69%) |
| Severely immunocompromised ‡ | 414 (24%) | 15 (23%) |
| Received a COVID-19 vaccine | 1389 (81%) | 49 (75%) |
| Use of other treatments | | |
| Corticosteroids § | 1556 (90%) | 56 (86%) |
| Remdesivir | 628 (36%) | 20 (31%) |
| Tocilizumab | 281 (16%) | 6 (9%) |
| Plan to use tocilizumab within the next 24 hours | 115 (7%) | 4 (6%) |

Data are mean (SD), n (%), or median (IQR). * Defined as requiring ongoing specialist care. † Defined as estimated glomerular filtration rate <30 mL/min per 1.73 $m^2$. ‡ In the opinion of the managing clinician. § Including all those randomised into the comparison of high vs low dose steroids.

#### Webtable 2: Baseline characteristics by patient baseline antigen status and randomised allocation

| | Antigen positive patients | | Antigen negative patients | | Antigen status unknown | |
|---|---|---|---|---|---|---|
| | Sotrovimab (n=355) | Usual care (n=365) | Sotrovimab (n=339) | Usual care (n=378) | Sotrovimab (n=134) | Usual care (n=152) |
| Age, years | 72.5 (13.3) | 72.1 (13.7) | 70.5 (14.6) | 71.1 (15.6) | 68.0 (15.0) | 64.4 (17.2) |
| <70 | 123 (35%) | 141 (39%) | 153 (45%) | 143 (38%) | 66 (49%) | 85 (56%) |
| ≥70 to <80 | 123 (35%) | 121 (33%) | 85 (25%) | 107 (28%) | 43 (32%) | 44 (29%) |
| ≥80 | 109 (31%) | 103 (28%) | 101 (30%) | 128 (34%) | 25 (19%) | 23 (15%) |
| Sex | | | | | | |
| Male | 218 (61%) | 226 (62%) | 197 (58%) | 222 (59%) | 75 (56%) | 95 (63%) |
| Female | 137 (39%) | 139 (38%) | 142 (42%) | 156 (41%) | 59 (44%) | 57 (38%) |
| Ethnicity | | | | | | |
| White | 301 (85%) | 333 (91%) | 302 (89%) | 330 (87%) | 103 (77%) | 116 (76%) |
| Black, Asian, and minority ethnic | 32 (9%) | 16 (4%) | 17 (5%) | 25 (7%) | 15 (11%) | 24 (16%) |
| Unknown | 22 (6%) | 16 (4%) | 20 (6%) | 23 (6%) | 16 (12%) | 12 (8%) |
| Number of days since symptom onset | 6 (3-11) | 6 (3-12) | 4 (3-9) | 5 (2-8) | 8 (5-13) | 7 (4-12) |
| Number of days since admission to hospital | 2 (1-5) | 2 (1-5) | 2 (1-6) | 2 (1-5) | 2 (1-4) | 2 (1-4) |
| Respiratory support received | | | | | | |
| None | 43 (12%) | 54 (15%) | 57 (17%) | 66 (17%) | 19 (14%) | 17 (11%) |
| Simple oxygen | 226 (64%) | 213 (58%) | 219 (65%) | 253 (67%) | 67 (50%) | 91 (60%) |
| Non-invasive ventilation | 71 (20%) | 87 (24%) | 57 (17%) | 49 (13%) | 40 (30%) | 33 (22%) |
| Invasive mechanical ventilation | 15 (4%) | 11 (3%) | 6 (2%) | 10 (3%) | 8 (6%) | 11 (7%) |
| Previous diseases | | | | | | |
| Diabetes | 107 (30%) | 84 (23%) | 98 (29%) | 97 (26%) | 44 (33%) | 38 (25%) |
| Heart disease | 119 (34%) | 113 (31%) | 103 (30%) | 115 (30%) | 37 (28%) | 44 (29%) |
| Chronic lung disease | 123 (35%) | 128 (35%) | 156 (46%) | 149 (39%) | 48 (36%) | 48 (32%) |
| Tuberculosis | 0 (0%) | 1 (<0.5%) | 2 (1%) | 3 (1%) | 0 (0%) | 0 (0%) |
| HIV | 3 (1%) | 1 (<0.5%) | 1 (<0.5%) | 2 (1%) | 2 (1%) | 2 (1%) |
| Severe liver disease * | 6 (2%) | 3 (1%) | 8 (2%) | 8 (2%) | 5 (4%) | 5 (3%) |
| Severe kidney impairment † | 45 (13%) | 41 (11%) | 22 (6%) | 25 (7%) | 17 (13%) | 8 (5%) |
| Any of the above | 242 (68%) | 237 (65%) | 240 (71%) | 273 (72%) | 96 (72%) | 92 (61%) |
| Severely immunocompromised ‡ | 112 (32%) | 112 (31%) | 56 (17%) | 61 (16%) | 38 (28%) | 35 (23%) |
| Received a COVID-19 vaccine | 296 (83%) | 292 (80%) | 280 (83%) | 317 (84%) | 99 (74%) | 105 (69%) |
| Use of other treatments | | | | | | |
| Corticosteroids § | 329 (93%) | 334 (92%) | 301 (89%) | 333 (88%) | 125 (93%) | 134 (88%) |
| Remdesivir | 144 (41%) | 128 (35%) | 135 (40%) | 136 (36%) | 36 (27%) | 49 (32%) |
| Tocilizumab | 66 (19%) | 60 (16%) | 43 (13%) | 44 (12%) | 35 (26%) | 33 (22%) |
| Plan to use tocilizumab within the next 24 hours | 28 (8%) | 33 (9%) | 15 (4%) | 19 (5%) | 5 (4%) | 15 (10%) |
| Viral load in baseline nose swab | | | | | | |
| Median level (log viral copies per ml) | 6.1 (4.6-7.0) | 6.1 (5.0-7.2) | 4.7 (2.5-6.4) | 4.9 (2.8-6.2) | 5.7 (3.9-6.5) | 5.0 (2.9-6.1) |
| Serostatus (anti N) | | | | | | |
| Positive | 62 (17%) | 76 (21%) | 152 (45%) | 163 (43%) | 0 (0%) | 1 (1%) |
| Negative | 293 (83%) | 289 (79%) | 187 (55%) | 215 (57%) | 1 (1%) | 0 (0%) |
| Unknown | 0 (0%) | 0 (0%) | 0 (0%) | 0 (0%) | 133 (99%) | 151 (99%) |
| Serostatus (anti S) | | | | | | |
| Positive | 252 (71%) | 262 (72%) | 316 (93%) | 348 (92%) | 1 (1%) | 0 (0%) |
| Negative | 103 (29%) | 103 (28%) | 23 (7%) | 30 (8%) | 0 (0%) | 0 (0%) |
| Unknown | 0 (0%) | 0 (0%) | 0 (0%) | 0 (0%) | 133 (99%) | 152 (100%) |

Data are mean (SD), n (%), or median (IQR). * Defined as requiring ongoing specialist care. † Defined as estimated glomerular filtration rate <30 mL/min per 1.73 m². ‡ In the opinion of the managing clinician. § Including all those randomised into the comparison of high vs low dose steroids.

#### Webtable 3: Treatments given (antigen positive and all patients) by randomised allocation

| | **Antigen positive patients** | | **All patients** | |
|---|---|---|---|---|
| | **Sotrovimab (n=355)** | **Usual care (n=365)** | **Sotrovimab (n=828)** | **Usual care (n=895)** |
| Compliance data available | 353 | 363 | 820 | 890 |
| Sotrovimab | 335 (95%) | 8 (2%) | 767 (94%) | 14 (2%) |
| Other treatments received | | | | |
| Corticosteroid | 296 (84%) | 307 (85%) | 687 (84%) | 748 (84%) |
| Azithromycin or other macrolide | 57 (16%) | 57 (16%) | 112 (14%) | 126 (14%) |
| Tocilizumab or sarilumab | 68 (19%) | 60 (17%) | 114 (14%) | 126 (14%) |
| Remdesivir | 133 (38%) | 131 (36%) | 282 (34%) | 302 (34%) |
| Aspirin | 31 (9%) | 34 (9%) | 83 (10%) | 77 (9%) |
| Baricitinib | 17 (5%) | 23 (6%) | 31 (4%) | 49 (6%) |
| Empagliflozin | 68 (19%) | 73 (20%) | 170 (21%) | 195 (22%) |
| Molnupiravir | 95 (27%) | 93 (26%) | 187 (23%) | 198 (22%) |
| Paxlovid | 20 (6%) | 25 (7%) | 40 (5%) | 48 (5%) |

Percentages are of those with a completed follow-up form.

#### Webtable 4: Effect of allocation to sotrovimab on key study outcomes in all patients

| | Sotrovimab (n=828) | Usual care (n=895) | RR (95% CI) or mean difference | p-value |
|---|---|---|---|---|
| **Primary outcome** | | | | |
| 28-day mortality | 177 (21%) | 201 (22%) | 0.95 (0.77-1.16) | 0.60 |
| **Secondary outcomes** | | | | |
| Median (IQR) time to being discharged alive, days | 11 (6 to >28) | 11 (6 to >28) | | |
| Discharged from hospital within 28 days | 563 (68%) | 609 (68%) | 0.96 (0.85-1.08) | |
| Receipt of invasive mechanical ventilation or death* | 184/799 (23%) | 201/863 (23%) | 0.98 (0.84-1.16) | |
| Invasive mechanical ventilation | 39/799 (5%) | 36/863 (4%) | 1.16 (0.76-1.77) | |
| Death | 164/799 (21%) | 186/863 (22%) | 0.95 (0.79-1.13) | |
| **Subsidiary clinical outcomes** | | | | |
| Use of ventilation† | 70/631 (11%) | 83/694 (12%) | 0.92 (0.68-1.23) | |
| Non-invasive ventilation | 67/631 (11%) | 77/694 (11%) | 0.95 (0.70-1.28) | |
| Invasive mechanical ventilation | 15/631 (2%) | 16/694 (2%) | 1.00 (0.50-1.99) | |
| Successful cessation of invasive mechanical ventilation ‡ | 11/29 (38%) | 13/32 (41%) | 0.82 (0.37-1.83) | |
| Use of haemodialysis or haemofiltration § | 31/814 (4%) | 22/879 (3%) | 1.51 (0.89-2.56) | |
| **Virological outcomes** | | | | |
| Baseline-adjusted viral load (log copies/ml) on day 3 | 4.34 (0.06) | 4.38 (0.07) | -0.04 (-0.22, 0.14) | |
| Baseline-adjusted viral load (log copies/ml) on day 5 | 3.72 (0.09) | 3.85 (0.08) | -0.13 (-0.36, 0.10) | |

Data are n (%) or n/N (%), unless otherwise indicated. RR=rate ratio for the outcomes of 28-day mortality and hospital discharge, risk ratio for other clinical outcomes, and mean difference for virological outcomes. CI=confidence interval. Estimates of the RR or mean difference and their 95% CIs are adjusted for age in three categories (<70 years, 70-79 years, and 80 years or older) and ventilation status at randomisation in four categories (none, simple oxygen, non-invasive ventilation, and invasive mechanical ventilation). For the outcome of use of haemodialysis or haemofiltration, however, adjustment is for age in three categories (<70 years, 70-79 years, and 80 years or older) and ventilation status at randomisation in three categories (none, simple oxygen, non-invasive ventilation or invasive mechanical ventilation) due to model convergence issues. * Excluding patients receiving invasive mechanical ventilation at randomisation. † Excluding patients receiving invasive or non-invasive ventilation at randomisation. ‡ Excluding patients not receiving invasive mechanical ventilation at randomisation. § Excluding patients receiving renal replacement therapy at randomisation.

#### Webtable 5: Effect on cause-specific 28-day mortality

| | Antigen positive patients | | All patients | |
|---|---|---|---|---|
| **Cause of death** | **Sotrovimab (n=355)** | **Usual care (n=365)** | **Sotrovimab (n=828)** | **Usual care (n=895)** |
| COVID | 68 (19.2%) | 80 (21.9%) | 134 (16.2%) | 146 (16.3%) |
| Other infection | 3 (0.8%) | 2 (0.5%) | 4 (0.5%) | 3 (0.3%) |
| Cardiac | 1 (0.3%) | 4 (1.1%) | 5 (0.6%) | 8 (0.9%) |
| Stroke | 0 (0.0%) | 2 (0.5%) | 1 (0.1%) | 6 (0.7%) |
| Other vascular | 0 (0.0%) | 0 (0.0%) | 1 (0.1%) | 1 (0.1%) |
| Cancer | 3 (0.8%) | 6 (1.6%) | 7 (0.8%) | 13 (1.5%) |
| Other medical | 6 (1.7%) | 9 (2.5%) | 22 (2.7%) | 19 (2.1%) |
| External | 1 (0.3%) | 3 (0.8%) | 3 (0.4%) | 5 (0.6%) |
| Unknown cause | 0 (0.0%) | 0 (0.0%) | 0 (0.0%) | 0 (0.0%) |
| Total: 28-day mortality | 82 (23.1%) | 106 (29.0%) | 177 (21.4%) | 201 (22.5%) |

**Webtable 6: Effect on non-coronavirus infection, new cardiac arrhythmia, thrombotic events, clinically significant bleeds, metabolic complications and other outcomes**

| | Antigen positive patients | | All patients | |
|---|---|---|---|---|
| | **Sotrovimab (n=355)** | **Usual care (n=365)** | **Sotrovimab (n=828)** | **Usual care (n=895)** |
| Non-coronavirus infection | | | | |
| Pneumonia | 34 (9.6%) | 53 (14.5%) | 85 (10.3%) | 111 (12.4%) |
| Urinary tract | 11 (3.1%) | 5 (1.4%) | 22 (2.7%) | 14 (1.6%) |
| Biliary | 0 (0%) | 0 (0%) | 0 (0%) | 0 (0%) |
| Other intra-abdominal | 4 (1.1%) | 1 (0.3%) | 8 (1.0%) | 1 (0.1%) |
| Blood stream | 7 (2.0%) | 1 (0.3%) | 10 (1.2%) | 7 (0.8%) |
| Skin | 3 (0.8%) | 3 (0.8%) | 4 (0.5%) | 5 (0.6%) |
| Other | 13 (3.7%) | 9 (2.5%) | 29 (3.5%) | 31 (3.5%) |
| Subtotal: Any non-coronavirus infection | 66 (18.6%) | 66 (18.1%) | 145 (17.5%) | 151 (16.9%) |
| New cardiac arrhythmia | | | | |
| Atrial flutter or atrial fibrillation | 11 (3.1%) | 10 (2.7%) | 19 (2.3%) | 22 (2.5%) |
| Other supraventricular tachycardia | 1 (0.3%) | 1 (0.3%) | 6 (0.7%) | 2 (0.2%) |
| Subtotal: Supraventricular tachycardia | 12 (3.4%) | 11 (3.0%) | 23 (2.8%) | 24 (2.7%) |
| Ventricular tachycardia | 1 (0.3%) | 4 (1.1%) | 1 (0.1%) | 6 (0.7%) |
| Ventricular fibrillation | 0 (0%) | 0 (0%) | 1 (0.1%) | 0 (0%) |
| Subtotal: Ventricular tachycardia or fibrillation | 1 (0.3%) | 4 (1.1%) | 2 (0.2%) | 6 (0.7%) |
| Atrioventricular block requiring intervention | 1 (0.3%) | 0 (0%) | 2 (0.2%) | 0 (0%) |
| Unknown / not recorded | 0 (0%) | 0 (0%) | 1 (0.1%) | 0 (0%) |
| Subtotal: Any major cardiac arrhythmia | 14 (3.9%) | 15 (4.1%) | 28 (3.4%) | 30 (3.4%) |
| Thrombotic events | | | | |
| Pulmonary embolism | 6 (1.7%) | 4 (1.1%) | 13 (1.6%) | 15 (1.7%) |
| Deep-vein thrombosis | 0 (0%) | 0 (0%) | 2 (0.2%) | 3 (0.3%) |
| Ischaemic stroke | 2 (0.6%) | 0 (0%) | 4 (0.5%) | 2 (0.2%) |
| Myocardial infarction | 0 (0%) | 2 (0.5%) | 2 (0.2%) | 4 (0.4%) |
| Systemic arterial embolism | 0 (0%) | 1 (0.3%) | 0 (0%) | 1 (0.1%) |
| Subtotal: Any thrombotic event | 8 (2.3%) | 6 (1.6%) | 21 (2.5%) | 23 (2.6%) |
| Clinically significant bleeds | | | | |
| Intra-cranial | 0 (0%) | 1 (0.3%) | 0 (0%) | 3 (0.3%) |
| Gastrointestinal | 3 (0.8%) | 3 (0.8%) | 10 (1.2%) | 10 (1.1%) |
| Other/unrecorded site | 4 (1.1%) | 3 (0.8%) | 7 (0.8%) | 7 (0.8%) |
| Requiring blood transfusion | 4 (1.1%) | 5 (1.4%) | 13 (1.6%) | 15 (1.7%) |
| Requiring surgery | 2 (0.6%) | 0 (0%) | 2 (0.2%) | 1 (0.1%) |
| Requiring endoscopy | 1 (0.3%) | 1 (0.3%) | 3 (0.4%) | 4 (0.4%) |
| Requiring vasoactive drugs | 1 (0.3%) | 1 (0.3%) | 3 (0.4%) | 3 (0.3%) |
| Subtotal: Any clinically significant bleeding | 7 (2.0%) | 7 (1.9%) | 17 (2.1%) | 20 (2.2%) |
| Metabolic complications | | | | |
| Ketoacidosis | 1 (0.3%) | 1 (0.3%) | 1 (0.1%) | 1 (0.1%) |
| Hyperglycaemic hyperosmolar state | 6 (1.7%) | 3 (0.8%) | 9 (1.1%) | 8 (0.9%) |
| Other hyperglycaemia requiring new use of insulin | 22 (6.2%) | 25 (6.8%) | 49 (5.9%) | 50 (5.6%) |
| Subtotal: Any clinically significant hyperglycaemia | 27 (7.6%) | 26 (7.1%) | 55 (6.6%) | 54 (6.0%) |
| Severe hypoglycaemia | 2 (0.6%) | 1 (0.3%) | 5 (0.6%) | 2 (0.2%) |
| Acute kidney injury* | | | | |
| Stage 1 | 13/347 (3.7%) | 14/356 (3.9%) | 32/814 (3.9%) | 37/879 (4.2%) |
| Stage 2 | 3/347 (0.9%) | 8/356 (2.2%) | 8/814 (1.0%) | 21/879 (2.4%) |
| Stage 3 | 23/347 (6.6%) | 14/356 (3.9%) | 51/814 (6.3%) | 38/879 (4.3%) |
| Subtotal: Any acute kidney injury | 39/347 (11.2%) | 36/356 (10.1%) | 91/814 (11.2%) | 96/879 (10.9%) |
| Other outcomes | | | | |
| Seizures | 1 (0.3%) | 2 (0.5%) | 5 (0.6%) | 5 (0.6%) |
| ALT >3x ULN | 24 (6.8%) | 18 (4.9%) | 42 (5.1%) | 38 (4.2%) |
| ALT >3x ULN and bilirubin >2x ULN | 2 (0.6%) | 2 (0.5%) | 3 (0.4%) | 4 (0.4%) |

* Analyses exclude those on haemodialysis or haemofiltration at randomisation.

### Supplementary Figures

#### Webfigure 1: Distribution of serum antigen levels at randomisation

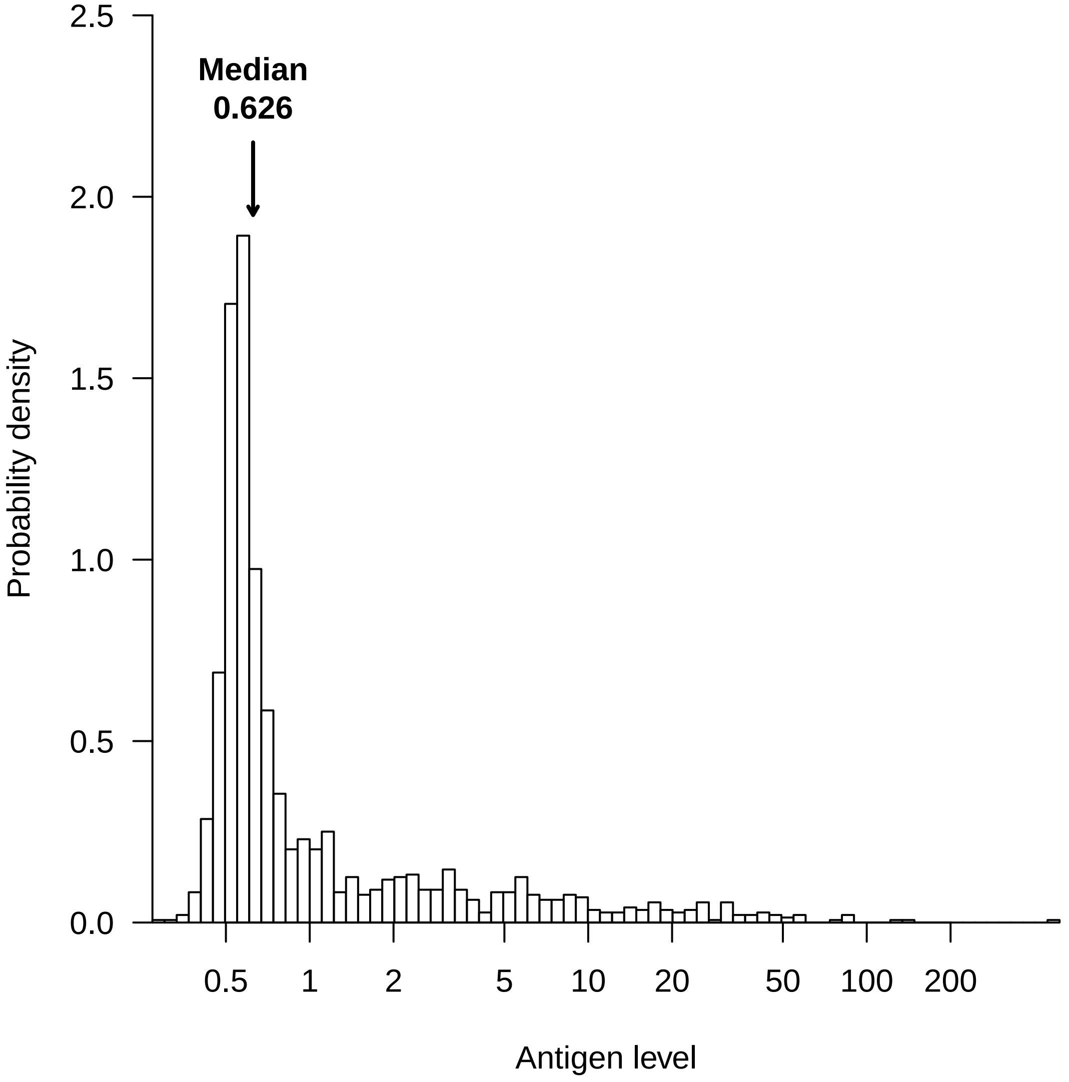

Distribution shown on the logarithmic scale

#### Webfigure 2: Effect of allocation to sotrovimab on 28−day mortality in all participants, by baseline characteristics

| | Sotrovimab | Usual care | RR (95% CI) |
|---|---|---|---|
| **Age, years ($\chi^2_1$= 0.0; p=0.95)** | | | |
| <70 | 48/342 (14%) | 55/369 (15%) | 0.95 (0.65−1.41) |
| ≥70 <80 | 58/251 (23%) | 66/272 (24%) | 0.95 (0.67−1.35) |
| ≥80 | 71/235 (30%) | 80/254 (31%) | 0.94 (0.68−1.29) |
| **Sex ($\chi^2_1$= 0.7; p=0.42)** | | | |
| Men | 104/490 (21%) | 133/543 (24%) | 0.89 (0.69−1.15) |
| Women | 73/338 (22%) | 68/352 (19%) | 1.06 (0.76−1.47) |
| **Ethnicity ($\chi^2_1$=2.3; p=0.13)** | | | |
| White | 152/706 (22%) | 185/779 (24%) | 0.89 (0.72−1.11) |
| Black, Asian and Minority Ethnic | 15/64 (23%) | 9/65 (14%) | 1.73 (0.75−3.95) |
| **Days since symptom onset ($\chi^2_1$= 0.0; p=0.90)** | | | |
| ≤7 | 88/495 (18%) | 99/543 (18%) | 0.96 (0.72−1.28) |
| >7 | 89/333 (27%) | 102/352 (29%) | 0.94 (0.70−1.24) |
| **Respiratory support at randomisation ($\chi^2_1$= 1.3; p=0.26)** | | | |
| None | 7/119 (6%) | 13/137 (9%) | 0.62 (0.25−1.56) |
| Simple oxygen | 93/512 (18%) | 114/557 (20%) | 0.89 (0.68−1.18) |
| Non−invasive ventilation | 64/168 (38%) | 59/169 (35%) | 1.09 (0.76−1.55) |
| Invasive mechanical ventilation | 13/29 (45%) | 15/32 (47%) | 1.06 (0.50−2.23) |
| **Use of corticosteroids ($\chi^2_1$= 1.6; p=0.20)** | | | |
| Yes | 172/755 (23%) | 187/801 (23%) | 0.97 (0.79−1.19) |
| No | 5/73 (7%) | 14/94 (15%) | 0.49 (0.17−1.38) |
| **Severely immunocompromised ($\chi^2_1$=0.8; p=0.38)** | | | |
| Yes | 52/206 (25%) | 62/208 (30%) | 0.81 (0.56−1.17) |
| No | 125/622 (20%) | 139/687 (20%) | 0.99 (0.77−1.25) |
| **All participants** | **177/828 (21%)** | **201/895 (22%)** | **0.95 (0.77−1.16) p=0.60** |

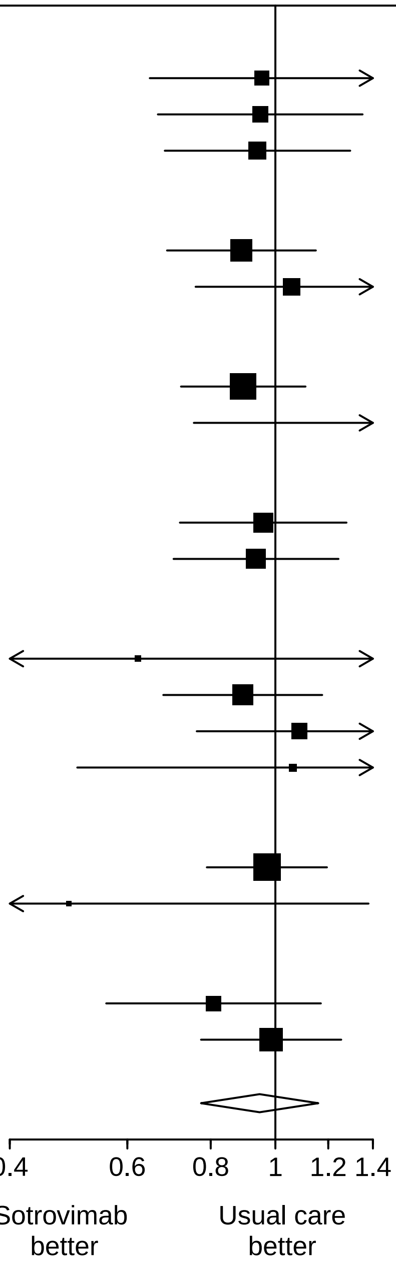

Subgroup−specific rate ratio estimates are represented by squares (with areas of the squares proportional to the amount of statistical information) and the lines through them correspond to 95% CIs. The ethnicity subgroup excludes patients with missing data, but these patients are included in the overall summary diamond. RR=Rate ratio adjusted for age and respiratory support status at randomisation.

#### Webfigure 3: Primary and secondary outcomes, overall and by Roche anti−S status

| Outcome, subgroup | Sotrovimab | Usual care | RR (95% CI) |
|---|---|---|---|
| **Death within 28 days ($\chi^2_1$= 1.6; p=0.21)** | | | |
| Positive | 111/569 (20%) | 120/610 (20%) | 0.98 (0.75−1.26) |
| Negative | 24/126 (19%) | 42/133 (32%) | 0.68 (0.40−1.13) |
| Unknown status | 42/133 (32%) | 39/152 (26%) | 1.35 (0.87−2.10) |
| **All participants** | **177/828 (21%)** | **201/895 (22%)** | **0.95 (0.77−1.16)** |
| **Discharge alive from hospital ($\chi^2_1$=0.0; p=0.87)** | | | |
| Positive | 400/569 (70%) | 427/610 (70%) | 0.97 (0.85−1.11) |
| Negative | 84/126 (67%) | 82/133 (62%) | 0.94 (0.69−1.29) |
| Unknown status | 79/133 (59%) | 100/152 (66%) | 0.89 (0.66−1.20) |
| **All participants** | **563/828 (68%)** | **609/895 (68%)** | **0.96 (0.85−1.08)** |
| **Invasive mechanical ventilation or death ($\chi^2_1$=2.2; p=0.14)** | | | |
| Positive | 112/550 (20%) | 120/594 (20%) | 1.00 (0.81−1.25) |
| Negative | 26/124 (21%) | 43/128 (34%) | 0.72 (0.49−1.06) |
| Unknown status | 46/125 (37%) | 38/141 (27%) | 1.29 (0.95−1.76) |
| **All not on invasive mechanical ventilation at randomisation** | **184/799 (23%)** | **201/863 (23%)** | **0.98 (0.84−1.16)** |

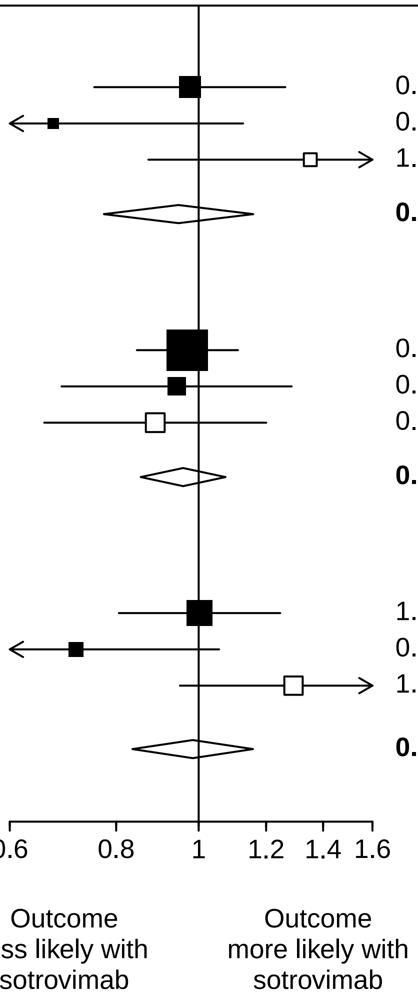

Subgroup−specific RR estimates are represented by squares (with areas of the squares proportional to the amount of statistical information) and the lines through them correspond to the 95% CIs. Open squares represent participants with unknown status. The tests for heterogeneity compare the log RRs in seropositive vs seronegative patients (i.e. excluding those with unknown serostatus). RR=risk ratio for the composite outcome of invasive mechanical ventilation or death, and rate ratio for the other outcomes.

#### Webfigure 4: Primary and secondary outcomes, overall and by Roche anti−N status

| Outcome, subgroup | Sotrovimab | Usual care | RR (95% CI) |
|---|---|---|---|
| **Death within 28 days ($\chi^2_1$= 2.6; p=0.10)** | | | |
| Positive | 47/214 (22%) | 46/240 (19%) | 1.16 (0.78−1.75) |
| Negative | 88/481 (18%) | 116/504 (23%) | 0.77 (0.59−1.02) |
| Unknown status | 42/133 (32%) | 39/151 (26%) | 1.35 (0.87−2.08) |
| **All participants** | **177/828 (21%)** | **201/895 (22%)** | **0.95 (0.77−1.16)** |
| **Discharge alive from hospital ($\chi^2_1$=0.6; p=0.46)** | | | |
| Positive | 141/214 (66%) | 162/240 (68%) | 0.91 (0.72−1.14) |
| Negative | 343/481 (71%) | 348/504 (69%) | 1.00 (0.87−1.17) |
| Unknown status | 79/133 (59%) | 99/151 (66%) | 0.89 (0.66−1.21) |
| **All participants** | **563/828 (68%)** | **609/895 (68%)** | **0.96 (0.85−1.08)** |
| **Invasive mechanical ventilation or death ($\chi^2_1$=0.2; p=0.63)** | | | |
| Positive | 44/203 (22%) | 50/230 (22%) | 0.97 (0.70−1.36) |
| Negative | 94/471 (20%) | 113/493 (23%) | 0.88 (0.70−1.11) |
| Unknown status | 46/125 (37%) | 38/140 (27%) | 1.29 (0.95−1.75) |
| **All not on invasive mechanical ventilation at randomisation** | **184/799 (23%)** | **201/863 (23%)** | **0.98 (0.84−1.16)** |

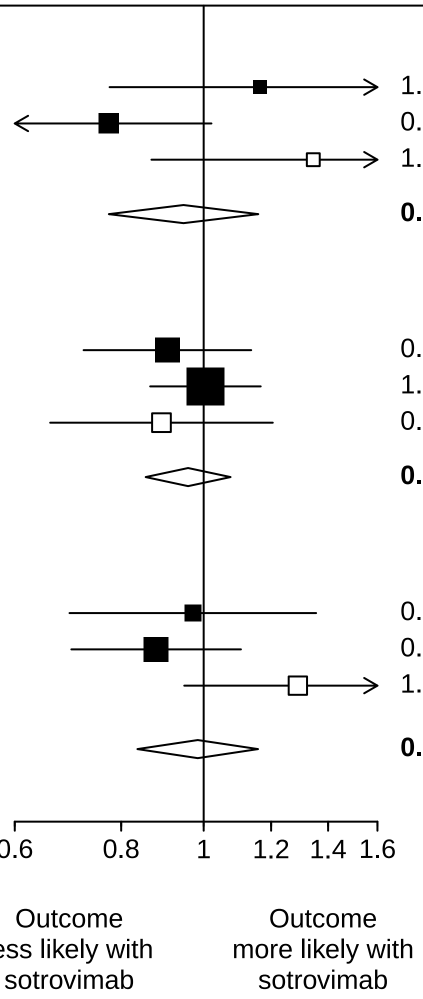

Subgroup−specific RR estimates are represented by squares (with areas of the squares proportional to the amount of statistical information) and the lines through them correspond to the 95% CIs. Open squares represent participants with unknown status. The tests for heterogeneity compare the log RRs in seropositive vs seronegative patients (i.e. excluding those with unknown serostatus). RR=risk ratio for the composite outcome of invasive mechanical ventilation or death, and rate ratio for the other outcomes.

### Appendices

### Appendix 1: RECOVERY Trial Protocol V27.0

RECOVERY
Randomised Evaluation of COVID-19 Therapy

### RANDOMISED EVALUATION OF COVID-19 THERAPY (RECOVERY)

**Background:** In early 2020, as this protocol was being developed, there were no approved treatments for COVID-19, a disease induced by the novel coronavirus SARS-CoV-2 that emerged in China in late 2019. The UK New and Emerging Respiratory Virus Threats Advisory Group (NERVTAG) advised that several possible treatments should be evaluated, including Lopinavir-Ritonavir, low-dose corticosteroids, and Hydroxychloroquine (which has now been done). A World Health Organization (WHO) expert group issued broadly similar advice. These groups also advised that other treatments would soon emerge that require evaluation. In addition, due to lack of community transmission due to COVID-19 control measures, a more severe influenza season is expected when these ease.

**Eligibility and randomisation:** This protocol describes a randomised trial among patients hospitalised for COVID-19 and/or influenza. (Treatments for influenza are only being assessed in the UK.) Eligible patients are randomly allocated between one or more treatment arms, each to be given in addition to the usual standard of care in the participating hospital. The study is dynamic, and treatments are added and removed as results and suitable treatments become available. The randomised treatment comparisons in this version of the protocol (which should be checked and confirmed as the current version) are shown in Table 1. For patients for whom not all the trial arms are appropriate or at locations where not all are available, randomisation will be between fewer arms..

| Condition | Randomised comparisons, each vs. usual care alone | UK | Other countries |
|---|---|---|---|
| COVID-19 | High-dose corticosteroids | ✓ (age ≥18 years with hypoxia)[a] | ✓ (age ≥18 years with hypoxia)[a] |
| | Empagliflozin | ✓ (age ≥18 years) | ✓ (age ≥18 years) |
| | Sotrovimab | ✓ (age ≥12 years) | ✗ |
| | Molnupiravir | ✓ (age ≥18 years) | ✓ (age ≥18 years) |
| | Paxlovid | ✓ (age ≥18 years) | ✗ |
| Influenza | Baloxavir | ✓ (age ≥12 years) | ✗ |
| | Oseltamivir | ✓ (any age) | ✗ |
| | Low-dose corticosteroids | ✓ (any age with hypoxia)[b] | ✗ |
| [a] without suspected or confirmed influenza infection; [b] without suspected or confirmed SARS-CoV-2 infection. Information on completed arms is available in Section 7. | | | |

**Table 1: Current comparisons**

In a partial factorial design, participants may be entered into one or more randomised comparisons of active treatment plus usual care vs. usual care alone, simultaneously. This allows the effects of one treatment to be assessed in the presence or absence of another which generates useful information for clinicians and health policy-makers. In particular, this allows antiviral therapies to be assessed as monotherapy and in combination, which will provide important information on the efficacy, safety and the development of resistance. This protocol indicates clearly where specific combinations are not desirable.

**Adaptive design:** The interim trial results will be monitored by an independent Data Monitoring Committee (DMC). The most important task for the DMC will be to assess whether the randomised comparisons in the study have provided evidence on mortality that is strong enough (with a range of uncertainty around the results that is narrow enough) to affect national and global treatment strategies. In such a circumstance, the DMC will inform the Trial Steering Committee who will make the results available to the public and amend the trial arms accordingly. Regardless, follow-up will continue for all randomised participants, including those previously assigned to trial arms that are modified or ceased. New trial arms can be added as evidence emerges that other candidate therapeutics should be evaluated.

**Outcomes:** The main outcomes will be death, discharge, need for ventilation and need for renal replacement therapy. For the main analyses, follow-up will be censored at 28 days after randomisation. Additional information on longer term outcomes may be collected through review of medical records or linkage to medical databases where available (such as those managed by NHS Digital and equivalent organisations in the devolved nations).

**Simplicity of procedures:** To facilitate collaboration, even in hospitals that suddenly become overloaded, patient enrolment (via the internet) and all other trial procedures are greatly streamlined. Informed consent is simple and data entry is minimal. Randomisation via the internet is simple and quick, at the end of which the allocated treatment is displayed on the screen and can be printed or downloaded. Key follow-up information is recorded at a single timepoint and may be ascertained by contacting participants in person, by phone or electronically, or by review of medical records and databases.

**Data to be recorded:** At randomisation, information will be collected on the identity of the randomising clinician and of the patient, age, sex, major co-morbidity, pregnancy, illness onset date and severity, and any contraindications to the study treatments. The main outcomes will be death (with date and probable cause), discharge (with date), need for ventilation (with number of days recorded) and need for renal replacement therapy. Reminders will be sent if outcome data have not been recorded by 28 days after randomisation. Suspected Unexpected Serious Adverse Reactions (SUSARs) to one of the study medications (e.g., Stevens-Johnson syndrome, anaphylaxis, aplastic anaemia) will be collected and reported in an expedited fashion. Other adverse events will not be recorded but may be available through linkage to medical databases.

**Numbers to be randomised:** The larger the number randomised the more accurate the results will be, but the numbers that can be randomised will depend critically on how large the epidemic becomes. If substantial numbers are hospitalised in the participating centres then it may be possible to randomise several thousand with mild disease and a few thousand with severe disease, but realistic, appropriate sample sizes could not be estimated at the start of the trial.

**Heterogeneity between populations:** If sufficient numbers are studied, it may be possible to generate reliable evidence in certain patient groups (e.g. those with major co-morbidity or who are older). To this end, data from this study may be combined with data from other trials of treatments for COVID-19 or influenza.

**Add-on studies:** Particular countries or groups of hospitals, may well want to collaborate in adding further measurements or observations, such as serial blood gases or chemistry, serial lung imaging, or serial documentation of other aspects of disease status. While well-organised additional research studies of the natural history of the disease or of the effects of the trial treatments could well be valuable (although the lack of placebo control may bias the assessment of subjective side-effects, such as gastro-intestinal problems), they are not core requirements.

**To enquire about the trial, contact the RECOVERY Central Coordinating Office**
Nuffield Department of Population Health, Richard Doll Building, Old Road Campus, Roosevelt Drive, Oxford OX3 7LF, United Kingdom
**To enquire about the trial outside of the UK, contact the relevant Clinical Trial Units**
**To RANDOMISE a patient, visit: www.recoverytrial.net**

### 1 BACKGROUND AND RATIONALE

#### 1.1 Setting

In 2019 a novel coronavirus-disease (COVID-19) emerged in Wuhan, China. A month later the Chinese Center for Disease Control and Prevention identified a new beta-coronavirus (SARS coronavirus 2, or SARS-CoV-2) as the aetiological agent.[1] The clinical manifestations of COVID-19 range from asymptomatic infection or mild, transient symptoms to severe viral pneumonia with respiratory failure. As many patients do not progress to severe disease the overall case fatality rate per infected individual is low, but hospitals in areas with significant community transmission have experienced a major increase in the number of hospitalised pneumonia patients, and the frequency of severe disease in hospitalised patients can be as high as 30%.[2-4] The progression from prodrome (usually fever, fatigue and cough) to severe pneumonia requiring oxygen support or mechanical ventilation often takes one to two weeks after the onset of symptoms.[2] The kinetics of viral replication in the respiratory tract are not well characterized, but this relatively slow progression provides a potential time window in which antiviral therapies could influence the course of disease. In May 2020 a new COVID-associated inflammatory syndrome in children was identified, Paediatric Inflammatory Multisystem Syndrome - Temporally associated with SARS-CoV-2 (PIMS-TS).[5] A rapid NHS England-led consensus process identified the need to evaluate corticosteroids and intravenous immunoglobulin (IVIg) as initial therapies in PIMS-TS, and confirmed tocilizumab as one of the biological anti-inflammatory agents to be evaluated as a second line therapy.

The COVID-19 control measures in place in the UK during the winter of 2020/21 resulted in an almost complete absence of influenza transmission over that period. This extended period without exposure to influenza viruses is unique and may have resulted in antibody waning and increased population susceptibility. Therefore, there is a possibility of a large resurgence of influenza in the winter of 2021/22. The treatment of influenza in hospitalised patients has progressed little in the last 20 years and there is substantial uncertainty and disagreement about optimal treatment of this patient group.

#### 1.2 Treatment Options

The protocol allows reliable assessment of the effects of multiple different treatments (including re-purposed and novel drugs) on major outcomes in COVID-19, influenza and PIMS-TS. All patients will receive usual care for the participating hospital. The current treatments under evaluation are summarised in Table 1 above with further details provided in sections 2.4-2.6 and in Appendices 1-4 (sections 8.1-8.4).

#### 1.3 Modifications to the number of treatment arms

Other arms can be added if evidence emerges that there are suitable candidate therapeutics. Conversely, in some patient populations, not all trial arms are appropriate (e.g. due to contraindications based on co-morbid conditions or concomitant medication); in some hospitals or countries, not all treatment arms will be available (e.g. due to manufacturing and supply issues); and at some times, not all treatment arms will be active (e.g. due to lack of relevant approvals and contractual agreements). The Trial Steering Committee may elect to pause one or more of the arms in order to increase trial efficiency during a fluctuating epidemic. In any of these situations, randomisation will be between fewer arms. Depending on the availability and suitability of treatments, it may be allowed for participants to be randomised in only one or two parts of the main randomisations.

##### 1.4 Design Considerations

The RECOVERY Protocol describes an overarching trial design to provide reliable evidence on the efficacy of candidate therapies for confirmed COVID-19 and/or influenza infection in hospitalised patients receiving usual standard of care. (Treatments for influenza are only being assessed in the UK.)

In early 2020, when the trial first started, there were no known treatments for COVID-19. The anticipated scale of the epidemic is such that hospitals, and particularly intensive care facilities, may be massively overstretched at some points in time, with around 10% requiring hospitalisation. Similarly, the winter of 2021-22 may pose a similar challenge to hospitals when ongoing COVID-19 cases coincide with a significant number of influenza cases. In such situations, even treatments with only a moderate impact on survival or on hospital resources could be worthwhile. Therefore, the focus of RECOVERY is the impact of candidate treatments on mortality and on the need for hospitalisation or ventilation.

Critically, the trial is designed to minimise the burden on front-line hospital staff working within an overstretched care system during a major epidemic. Eligibility criteria are therefore simple and trial processes (including paperwork) are minimised.

The protocol is deliberately flexible so that it is suitable for a wide range of settings, allowing:

- a broad range of patients to be enrolled in large numbers;
- randomisation between only those treatment arms that are *both* available at the hospital *and* not believed by the enrolling doctor to be contraindicated (e.g. by particular co-morbid conditions or concomitant medications);
- treatment arms to be added or removed according to the emerging evidence; and
- additional substudies may be added to provide more detailed information on side effects or sub-categorisation of patient types but these are not the primary objective and are not required for participation.

In a cohort of 191 hospitalised COVID-19 patients with a completed outcome, the median time from illness onset to discharge was 22.0 days (IQR 18.0-25.0) and the median time to death was 18.5 days (15.0-22.0). Thirty-two patients (17%) required invasive mechanical ventilation and the median time from onset to mechanical ventilation was 14.5 days. Therefore, early endpoint assessment, such as 28 days after randomisation, is likely to provide largely complete outcome data and will permit early assessment of treatment efficacy and safety.[6] For influenza, the average length of hospital stay in the UK is around 9 days, so assessment at 28 days will capture most outcomes.[7]

##### 1.5 Potential for effective treatments to become available

In early 2020, when the trial first started, there were no known treatments for COVID-19. However, over time, effective treatments may become available, typically as the result of reliable information from randomised trials (including from this study). For example, in June 2020, results from the RECOVERY trial showed that dexamethasone 6mg once daily reduces the mortality in COVID-19 patients requiring mechanical ventilation or oxygen. In response, many clinical guidelines now recommend the use of dexamethasone 6mg once daily as standard of care for these types of patients.

The RECOVERY trial randomises eligible participants to usual standard of care for the local hospital alone vs usual standard of care plus one or more additional study treatments. Over time, it is expected that usual standard of care alone will evolve. Thus randomisation will always be relevant to the current clinical situation and the incremental effects of the study treatments will be appropriately assessed.

##### 1.6 Early phase assessments

In the UK, the COVID-19 Therapeutics Advisory Panel (CTAP [a]) may propose that RECOVERY assesses interventions for which additional information is required before they are considered for large-scale assessment of the impact on mortality. Such assessments will be tailored to the uncertainty specific to the intervention and typically be conducted at a subset of sites among a smaller group of participants before the results are reviewed and a decision made whether to include them in the main trial.

#### 2 DESIGN AND PROCEDURES

##### 2.1 Eligibility

Patients are eligible for the study if all of the following are true:

**(i) Hospitalised**

**(ii) a) Viral pneumonia syndrome**

In general, viral pneumonia should be suspected when a patient presents with:

a) typical symptoms (e.g. influenza-like illness with fever and muscle pain, or respiratory illness with cough and shortness of breath); and
b) compatible chest X-ray findings (consolidation or ground-glass shadowing); and
c) alternative causes have been considered unlikely or excluded (e.g. heart failure, bacterial pneumonia).

However, the diagnosis remains a clinical one based on the opinion of the managing doctor.

**(iii) Confirmed SARS-CoV-2 infection (all countries) and/or influenza A or B infection (UK only)**

**(iv) No medical history that might, in the opinion of the attending clinician, put the patient at significant risk if he/she were to participate in the trial**

---

[a] https://www.gov.uk/government/publications/covid-19-treatments-making-a-proposal-for-clinical-trials/guidance-making-a-proposal-for-covid-19-therapeutics-clinical-trials#uk-covid-19-therapeutics-advisory-panel-uk-ctap

Patients in the UK with SARS-CoV-2 and influenza co-infection are eligible, but would be excluded from certain comparisons (as described in the table on page 1). In addition, if the attending clinician believes that there is a specific contra-indication to one of the active drug treatment arms (see Appendix 2, Appendix 3 [for children], and Appendix 4 for pregnant and breastfeeding women) or that the patient should definitely be receiving one of the active drug treatment arms then that arm will not be available for randomisation for that patient. For patients who lack capacity, an advanced directive or behaviour that clearly indicates that they would not wish to participate in the trial would be considered sufficient reason to exclude them from the trial.

Patients who have been previously recruited into RECOVERY are eligible to be recruited again as long as their previous randomisation was >6 months ago. Patients will not be recruited into the same randomised comparison (e.g. sotrovimab vs. usual care) on more than one occasion, regardless of how far apart they occur.

In some locations, children (aged <18 years) will not be recruited, to comply with local and national regulatory approvals (see Table 1 and Sections 2.4-2.6 and 8.3).

#### 2.2 Consent

Informed consent should be obtained from each patient 16 years and over before enrolment into the study. Due to the poor outcomes in COVID-19 patients who require ventilation (>90% mortality in one cohort[6]), patients who lack capacity to consent due to severe disease (e.g. needs ventilation), and for whom a relative to act as the legally designated representative is not available (in person), randomisation and consequent treatment will proceed with consent provided by a clinician (independent of the trial[b]) who will act as the legally designated representative (if allowed by local regulations).

If they regain capacity, such participants should be provided with information about the trial (ideally prior to discharge, but otherwise as soon as possible thereafter), what their rights are and how to exercise them, but it is not necessary to obtain their written consent[c]. Provision of such information (i.e. the current participant information sheet) will be documented in the medical record.

For children aged <16 years old consent will be sought from their parents or legal guardian. Where possible, children aged between 10-15 years old will also be asked for assent. Children aged ≥16 years old will asked for consent as for adults. Witnessed[d] consent may be obtained over the telephone or web video link if hospital visiting rules or parental infection mean a parent/guardian cannot be physically present.

Information about participants' involvement will be included in routine clinical communications (e.g. discharge summaries) provided to participants (and, in the UK their GPs) . If any other relevant information arises during the trial, this may also be sent to GPs.

---

[b] Independent clinicians may complete study training, but have no other involvement in the trial, e.g. eligibility assessment, or randomisation

[c] Unless required by local regulations. (This is not required in the UK.)

[d] The witness should be impartial i.e. not a member of the research team, but they do not require specific training or knowledge of the trial.

#### 2.3 Baseline information

The following information will be recorded on the web-based form by the attending clinician or delegate:

- Patient details (e.g. name or initials [depending on privacy requirements], NHS/CHI number [UK only] or medical records number, date of birth, sex)
- Clinician details (e.g. name)
- Symptom onset date
- Disease severity as assessed by need for supplemental oxygen, non-invasive ventilation or invasive mechanical ventilation/extracorporeal membrane oxygenation (ECMO)
- Oxygen saturations on air (if available)
- Latest routine measurement of creatinine, C-reactive protein, and D-dimer (if available)
- SARS-CoV-2 test result (and/or influenza test result in UK)
- Major co-morbidity (e.g. heart disease, diabetes, chronic lung disease) and pregnancy (including pregnancy test result in all women of child-bearing potential[e])
- Use of relevant medications (e.g. corticosteroids, anti-virals) and prior vaccination
- Date of hospitalisation
- Contraindication to the study treatment regimens (in the opinion of the attending clinician)
- Name of person completing the form

The person completing the form will then be asked to confirm that they wish to randomise the patient and will then be required to enter their name and e-mail address.

##### 2.3.1 Baseline sample collection (UK only)[f]

###### 2.3.1.1 Participants with COVID-19

Participants with COVID-19 entering sotrovimab, molnupiravir or Paxlovid comparisons should have a serum sample collected **after obtaining consent and prior to randomisation** in which presence of SARS-CoV-2 antigen and antibodies against it may be tested. In addition, a nasal swab should be collected in which the level of SARS-CoV-2 viral RNA (and genotyping for resistance markers) will be measured.

###### 2.3.1.2 Participants with influenza pneumonia

Participants with influenza pneumonia should have a nasal swab collected in which the presence of influenza virus will be measured.

[e] A woman of childbearing potential is defined as a post-menarchal pre-menopausal female capable of becoming pregnant. This includes women on oral, injectable, or mechanical contraception; women who are single; women whose male partners have been vasectomized or whose male partners have received or are utilizing mechanical contraceptive devices. The potential inclusion of any pregnant women should be discussed with a consultant obstetrician (or obstetric physician).

[f] Collection of these samples will continue until the Steering Committee determine (on the basis of data blinded to treatment allocation) that sufficient information is available to assess the effect of treatment on viral load and/or resistance markers.

RECOVERY
Randomised Evaluation of COVID-19 Therapy

#### 2.4 Randomised allocation of treatment for COVID-19

In addition to receiving usual care, eligible patients with confirmed SARS-CoV-2 infection will be allocated treatment(s) using a central web-based randomisation service (without stratification or minimisation). A factorial design is used such that eligible patients may be randomised simultaneously to one or more of the study treatment arms (depending on location and infection). The doses in this section are for adults (see Appendix 3 for paediatric dosing).

##### 2.4.1 Randomisation part E

Eligible patients (adult patients ≥18 years old without suspected or confirmed influenza co-infection) and with clinical evidence of hypoxia (i.e. receiving oxygen or with oxygen saturations <92% on room air) may be randomised in a ratio of 1:1 to one of the arms listed below.

- **No additional treatment**[g]

- High-dose corticosteroids: **dexamethasone 20 mg (base) once daily** by mouth, nasogastric tube or intravenous infusion for 5 days follow by **dexamethasone 10 mg (base) once daily** by mouth, nasogastric tube or intravenous infusion for 5 days. [h,i]

##### 2.4.2 Randomisation part F:

Eligible patients (adult patients ≥18 years old) may be randomised in a 1:1 ratio to one of the arms listed below.

- **No additional treatment**

- **Empagliflozin 10 mg once daily** by mouth for 28 days (or until discharge, if earlier). Participants with diabetes allocated empagliflozin should have daily ketone checks while taking the treatment (see Appendix 2 for further details).

##### 2.4.3 Randomisation part J (UK only):

Eligible patients (patients ≥12 years old) may be randomised in a 1:1 ratio to one of the arms listed below.

- **No additional treatment**

- **Sotrovimab 1000 mg in 100 mL 0.9% sodium chloride or 5% dextrose** by intravenous infusion over 1 hour as soon as possible after randomisation.

---

[g] Usual care in hypoxic patients is expected to include low dose (6mg daily) dexamethasone

[h] Treatment should be discontinued at 10 days or on discharge from hospital if sooner. Participants can be given a short 'weaning' course when they complete their study allocation if considered clinically necessary.

[i] Pregnant women should receive either prednisolone (130 mg) orally or hydrocortisone (540 mg in divided doses) intravenously or methylprednisolone (100 mg) intravenously for five days, followed by either prednisolone (65 mg) orally or hydrocortisone (270 mg in divided doses) intravenously or methylprednisolone (50 mg) intravenously for five days.

##### 2.4.4 Randomisation part K:

Eligible patients (patients ≥18 years old) may be randomised in a 1:1 ratio to one of the arms listed below.

- **No additional treatment**
- **Molnupiravir 800 mg twice daily** for 5 days by mouth[j].

##### 2.4.5 Randomisation part L (UK only):

Eligible patients (patients ≥18 years old) may be randomised in a 1:1 ratio to one of the arms listed below.

- **No additional treatment**
- **Paxlovid (nirmatrelvir/ritonavir) 300/100 mg twice daily** for 5 days by mouth[j].

#### 2.5 Randomised allocation of treatment for influenza (UK only)

In addition to receiving usual care, eligible patients with confirmed influenza A or B infection will be allocated treatment(s) using a central web-based randomisation service (without stratification or minimisation). A factorial design is used such that eligible patients may be randomised simultaneously to one or more of the study treatment arms (depending on location and infection). The doses in this section are for adults (see Appendix 3 for paediatric dosing). Study treatments do not need to be continued after discharge from hospital unless otherwise specified.

##### 2.5.1 Randomisation part G: (UK only)

Eligible patients (≥12 years old with or without SARS-CoV-2 co-infection) may be randomised in a ratio of 1:1 to one of the arms listed below.

- **No additional treatment**
- **Baloxavir marboxil 40mg (or 80mg if weight ≥80kg) once daily** by mouth or nasogastic tube to be given on day 1 and day 4[j].

##### 2.5.2 Randomisation part H: (UK only)

Eligible patients (any age, with or without SARS-CoV-2 co-infection) may be randomised in a ratio of 1:1 to one of the arms listed below.

- **No additional treatment**
- **Oseltamivir 75mg twice daily** by mouth or nasogastric tube for five days[j,k].

---

[j] If participant is discharged before course is complete, the participant should be provided with medication to complete the course at home.

[k] Course can be extended to 10 days for immunosuppressed patients at the managing clinician's discretion.

##### 2.5.3 Randomisation part I: (UK only)

Eligible patients (any age without suspected or confirmed SARS-CoV-2 infection) and with clinical evidence of hypoxia (i.e. receiving oxygen or with oxygen saturations <92% on room air) may be randomised in a ratio of 1:1 to one of the arms listed below.

- **No additional treatment**

- Low-dose corticosteroids: **Dexamethasone 6mg once daily given** orally or intravenously for ten days or until discharge (whichever happens earliest)[l]

#### 2.6 Administration of allocated treatment

The details of the allocated study treatments will be displayed on the screen and can be printed or downloaded. The hospital clinicians are responsible for prescription and administration of the allocated treatments. The patient's own doctors are free to modify or stop study treatments if they feel it is in the best interests of the patient without the need for the patient to withdraw from the study (see section 2.9). This study is being conducted within hospitals. Therefore use of medication will be subject to standard medication reviews (typically within 48 hours of enrolment) which will guide modifications to both the study treatment and use of concomitant medication (e.g. in the case of potential drug interactions).

#### 2.7 Collecting follow-up information

The following information will be ascertained at the time of death or discharge or at 28 days after first randomisation (whichever is sooner):

- Vital status (alive / dead, with date and presumed cause of death, if appropriate)
- Hospitalisation status (inpatient / discharged, with date of discharge, if appropriate)
- Use of ventilation (with days of use and type, if appropriate)
- Use of renal dialysis or haemofiltration
- Documented new major cardiac arrhythmia (including atrial and ventricular arrhythmias)
- Major bleeding (defined as intracranial bleeding or bleeding requiring transfusion, endoscopy, surgery, or vasoactive drugs)
- Thrombotic event, defined as either (i) acute pulmonary embolism; (ii) deep vein thrombosis; (iii) ischaemic stroke; (iv) myocardial infarction; or (v) systemic arterial embolism.
- Non-coronavirus / non-influenza infection, categorised by site and putative organism (virus, bacteria, fungus, other)
- Use of any medications included in the RECOVERY trial protocol (including drugs in the same class) or other purported COVID-19 and influenza treatments (e.g. remdesivir, neuraminidase inhibitors)
- Participation in other randomised trials of interventions (vaccines or treatments) for COVID-19 or influenza.
- Metabolic complications: Ketoacidosis; hyperglycaemic hyperosmolar state; hyperglycaemia requiring new use of insulin; severe hypoglycaemia (defined as hypoglycaemia causing reduced conscious level requiring another person to help recover)
- Seizures

[l] In pregnancy or breastfeeding women, prednisolone 40 mg administered by mouth (or intravenous hydrocortisone 80 mg twice daily) should be used instead of dexamethasone. For dosing in children see Appendix 3.

- Laboratory results: highest creatinine, alanine (or aspartate) transamine and bilirubin recorded during admission
- Infusion reactions to Sotrovimab
- For pregnant women in UK, ID number in UK Obstetric Surveillance System

Follow-up information is to be collected on all study participants, irrespective of whether or not they complete the scheduled course of allocated study treatment. Study staff will seek follow-up information through various means including medical staff, reviewing information from medical notes, routine healthcare systems, and registries.

For all randomised participants, vital status (alive / dead, with date and presumed cause of death, if appropriate) is to be ascertained at 28 days after first randomisation. This may be achieved through linkage to routine death registration data (e.g. in the UK) or through direct contact with the participant, their relatives, or medical staff and completion of an additional follow-up form. Where available, data from routine healthcare records (including linkage to medical databases held by organisations such as NHS Digital in the UK) will be used to supplement data collected by trial sites. Further details are described in the Definition and Derivation of Baseline Characteristics and Outcomes standard operating procedure.[m]

##### 2.7.1 Follow-up swab samples (UK only)[n]

###### 2.7.1.1 Participants with COVID-19

Participants with COVID-19 in sotrovimab, molnupiravir or Paxlovid comparisons should have a nasal swab collected on days 3 and 5 in which the level of SARS-CoV-2 viral RNA (and genotyping for resistance makers) will be measured.

###### 2.7.1.2 Participants with influenza pneumonia

Participants with influenza pneumonia should have a nasal swab collected on day 5 in which the presence of influenza virus (and genotyping for baloxavir or oseltamivir resistance markers) will be measured.

#### 2.8 Duration of follow-up

All randomised participants are to be followed up until death, discharge from hospital or 28 days after randomisation (whichever is sooner). It is recognised that in the setting of this trial, there may be some variability in exactly how many days after randomisation, information on disease status is collected. This is acceptable and will be taken account of in the analyses and interpretation of results, the principle being that some information about post-randomisation disease status is better than none.

In the UK, longer term (up to 10 years) follow-up will be sought through linkage to electronic healthcare records and medical databases including those held by NHS Digital, Public Health England and equivalent bodies, and to relevant research databases (e.g. UK Biobank, Genomics England). Outside the UK, due to the absence of electronic health data

---

[m] Available at https://www.recoverytrial.net/files/recovery-outcomes-definitions-v3-0.pdf

[n] Collection of these samples will continue until the Steering Committee determine (on the basis of data blinded to treatment allocation) that sufficient information is available to assess the effect of treatment on viral load and/or resistance markers. Participants discharged before day 5 will be asked to take this sample at home and will be provided with instructions and materials to do so.

linkage, additional follow-up will be conducted at 6 months after first randomisation by telephone or in person (at a clinic) in order to collect information on mortality (including date and cause) and re-admission to hospital (including date[s] and primary reason[s]). This information will be captured on a web-based case report form.

##### 2.9 Withdrawal of consent

A decision by a participant (or their parent/guardian) that they no longer wish to continue receiving study treatment should **not** be considered to be a withdrawal of consent for follow-up. However, participants (or their parent/guardian) are free to withdraw consent for some or all aspects of the study at any time if they wish to do so. In accordance with regulatory guidance, de-identified data that have already been collected and incorporated in the study database will continue to be used (and any identifiable data will be destroyed).

For participants who lack capacity, if their legal representative withdraws consent for treatment or methods of follow-up then these activities would cease. If such participants regain capacity and no longer wish to participate then they can withdraw the consent given on their behalf as above.

#### 3 STATISTICAL ANALYSIS

All analyses for reports, presentations and publications will be prepared by the coordinating centre at the Nuffield Department of Population Health, University of Oxford. A more detailed statistical analysis plan will be developed by the investigators and published on the study website whilst still blind to any analyses of aggregated data on study outcomes by treatment allocation.

##### 3.1 Outcomes

###### 3.1.1 Primary and secondary outcomes for evaluation of potential treatments for COVID-19

For each pairwise comparison with the ‘no additional treatment’ arm, the **primary objective** is to provide reliable estimates of the effect of study treatments on all-cause mortality at 28 days after randomisation (with subsidiary analyses of cause of death and of death at various timepoints following discharge).

The **secondary objectives** are to assess the effects of study treatments on (a) duration of hospital stay (time to discharge alive within the first 28 days); and, (b) among patients not on invasive mechanical ventilation at baseline, the composite endpoint of death or need for invasive mechanical ventilation or ECMO.

###### 3.1.2 Primary and secondary outcomes for evaluation of potential treatments for influenza

For each pairwise comparison with the ‘no additional treatment’ arm, the **co-primary objectives** are to provide reliable estimates of the effect of study treatments on (a) all-cause mortality at 28 days after randomisation (with subsidiary analyses of cause of death and of death at various timepoints following discharge) and (b) time to discharge alive from hospital.

Holm's procedure will be used to control the family-wise error rate across these two co-primary outcomes at 5%.[8]

The **secondary objective** is to assess the effects of study treatments on the composite endpoint of death or need for invasive mechanical ventilation or ECMO among patients not on invasive mechanical ventilation at baseline.

##### 3.1.3 Other outcomes for evaluation of all treatments

Other objectives include the assessment of the effects of study treatments on the need for any ventilation (and duration of invasive mechanical ventilation), acute kidney injury and renal replacement therapy, and thrombotic events. Safety outcomes include bleeding, new major cardiac arrhythmias, metabolic complications (ketoacidosis, hyperglycaemic hyperosmolar state, hyperglycaemia requiring new use of insulin, severe hypoglycaemia). Virological outcomes include viral RNA levels in the nasopharynx and the frequency of detection of resistance markers.

Study outcomes will be assessed based on data recorded up to 28 days and up to 6 months after randomisation.

Where available, data from routine healthcare records (including linkage to medical databases held by organisations such as NHS Digital in the UK) and from relevant research studies (such as UK Biobank, Genomics England, ISARIC-4C, the UK Obstetric Surveillance System and PHOSP-COVID) will allow subsidiary analyses of the effect of the study treatments on particular non-fatal events (e.g. ascertained through linkage to Hospital Episode Statistics), the influence of pre-existing major co-morbidity (e.g. diabetes, heart disease, lung disease, hepatic insufficiency, severe depression, severe kidney impairment, immunosuppression), the maternal and infant outcomes in women pregnant at randomisation, and longer-term outcomes as well as in particular sub-categories of patient (e.g. by genotype, pregnancy).

#### 3.2 Methods of analysis

For all outcomes, comparisons will be made between all participants randomised to each treatment and its control, irrespective of whether they received their allocated treatment ("intention-to-treat" analyses).

For time-to-event analyses, each treatment group will be compared with the no additional treatment group using the log-rank test. Kaplan-Meier estimates for the time to event will also be plotted (with associated log-rank p-values). The log-rank 'observed minus expected' statistic (and its variance) will be used to estimate the average event rate ratio (and its confidence interval) for those allocated to each treatment group versus the no additional treatment group. For the primary outcome, participants discharged before 28 days will, in the absence of information to the contrary, be assumed to have survived for 28 days. For binary outcomes where the timing of the event is unknown, the risk ratio and its 95% confidence interval (and associated p-value) will be reported.

Pairwise comparisons within each randomisation will be made between each treatment arm and the no additional treatment arm (reference group) in that particular randomisation. However, since not all treatments may be available or suitable for all patients, those in the no additional treatment arm will only be included in a given comparison if, at the point of their randomisation, they *could* alternatively have been randomised to the active treatment of interest. All p-values will be 2-sided.

Pre-specified subgroup analysis (e.g., level of respiratory support, time since onset of symptoms; sex; age group; ethnicity; use of corticosteroids) will be conducted, with tests for heterogeneity (or trend) performed to assess if the effect in any particular subgroup varies materially from the overall effect. Sensitivity analyses will be conducted among those patients with laboratory confirmed SARS-CoV-2. The effect of each treatment (versus its control) will be assessed in the presence or absence of other relevant treatments the patients may receive either (a) as part of their usual care; or (b) as part of the trial (i.e., other factorial randomisations). Further details will be fully described in the Statistical Analysis Plan.

### 4 DATA AND SAFETY MONITORING

#### 4.1 Recording Suspected Serious Adverse Reactions

The focus is on those events that, based on a single case, are highly likely to be related to the study medication. Examples include anaphylaxis, Stevens-Johnson Syndrome, or bone marrow failure, where there is no other plausible explanation.

Any Serious Adverse Event[o] that is believed with a reasonable probability to be due to one of the study treatments will be considered a Suspected Serious Adverse Reaction (SSAR). In making this assessment, there should be consideration of the probability of an alternative cause (for example, COVID-19 or influenza itself or some other condition preceding randomisation), the timing of the event with respect to study treatment, the response to withdrawal of the study treatment, and (where appropriate) the response to subsequent re-challenge.

All SSARs should be reported by telephone to the Central Coordinating Office and recorded on the study IT system immediately.

#### 4.2 Central assessment and onward reporting of SUSARs

Clinicians at the Central Coordinating Office are responsible for expedited review of reports of SSARs received. Additional information (including the reason for considering it both serious and related, and relevant medical and medication history) will be sought.

The focus of Suspected Unexpected Serious Adverse Reaction (SUSAR) reporting will be on those events that, based on a single case, are highly likely to be related to the study medication. To this end, anticipated events that are either efficacy endpoints, consequences of the underlying disease, or common in the study population will be exempted from expedited reporting. Thus the following events will be exempted from expedited reporting:

---

[o] Serious Adverse Events are defined as those adverse events that result in death; are life-threatening; require in-patient hospitalisation or prolongation of existing hospitalisation; result in persistent or significant disability or incapacity; result in congenital anomaly or birth defect; or are important medical events in the opinion of the responsible investigator (that is, not life-threatening or resulting in hospitalisation, but may jeopardise the participant or require intervention to prevent one or other of the outcomes listed above).

(i) Events which are the consequence of COVID-19 or influenza; and
(ii) Common events which are the consequence of conditions preceding randomisation.

Any SSARs that are not exempt will be reviewed by a Central Coordinating Office clinician and an assessment made of whether the event is “expected” or not (assessed against the relevant Summary of Product Characteristics or Investigator Brochure). Any SSARs that are not expected would be considered a Suspected Unexpected Serious Adverse Reaction (SUSAR).

All confirmed SUSARs will be reported to the Chair of the DMC and to relevant regulatory authorities, ethics committees, and investigators in an expedited manner in accordance with regulatory requirements.

##### 4.3 Recording other Adverse Events

In addition to recording Suspected Serious Adverse Reactions (see section 4.1), information will be collected on all deaths and efforts will be made to ascertain the underlying cause. Other serious or non-serious adverse events will not be recorded unless specified in section 2.7. [p] It is anticipated that for some substudies, more detailed information on adverse events (e.g. through linkage to medical databases) or on other effects of the treatment (e.g. laboratory or radiological features) will be recorded and analysed but this is not a requirement of the core protocol.

##### 4.4 Role of the Data Monitoring Committee (DMC)

During the study, interim analyses of all study data will be supplied in strict confidence to the independent DMC. The DMC will request such analyses at a frequency relevant to the emerging data from this and other studies.

The DMC will independently evaluate these analyses and any other information considered relevant. The DMC will determine if, in their view, the randomised comparisons in the study have provided evidence on mortality that is strong enough (with a range of uncertainty around the results that is narrow enough) to affect national and global treatment strategies. In such a circumstance, the DMC will inform the Trial Steering Committee who will make the results available to the public and amend the trial arms accordingly. Unless this happens, the Trial Steering Committee, Chief Investigator, study staff, investigators, study participants, funders and other partners will remain blind to the interim results until 28 days after the last patient has been randomised for a particular intervention arm (at which point analyses may be conducted comparing that arm with the no additional treatment arm).

The DMC will review the safety and efficacy analyses among children (age <18 years) both separately and combined with the adult data.

[p] Outside the UK, additional serious adverse event information (event description, date of onset, outcome, relatedness to study treatment) will be collected if required by national regulations. This will be collected on a web-based case report form and any forms required by local regulations.

##### 4.5 Blinding

This is an open-label study. However, while the study is in progress, access to tabular results of study outcomes by allocated treatment allocation will not be available to the research team, patients, or members of the Trial Steering Committee (unless the DMC advises otherwise).

#### 5 QUALITY MANAGEMENT

##### 5.1 Quality By Design Principles

In accordance with the principles of Good Clinical Practice and the recommendations and guidelines issued by regulatory agencies, the design, conduct and analysis of this trial is focussed on issues that might have a material impact on the wellbeing and safety of study participants (hospitalised patients with confirmed SARS-CoV-2 or influenza infection) and the reliability of the results that would inform the care for future patients.

The critical factors that influence the ability to deliver these quality objectives are:

- to minimise the burden on busy clinicians working in an overstretched hospital during a major epidemic
- to ensure that suitable patients have access to the trial medication without impacting or delaying other aspects of their emergency care
- to provide information on the study to patients and clinicians in a timely and readily digestible fashion but without impacting adversely on other aspects of the trial or the patient's care
- to allow individual clinicians to use their judgement about whether any of the treatment arms are not suitable for the patient
- to collect comprehensive information on the mortality and disease status

In assessing any risks to patient safety and well-being, a key principle is that of proportionality. Risks associated with participation in the trial must be considered in the context of usual care.

##### 5.2 Training and monitoring

The focus will be on those factors that are critical to quality (i.e. the safety of the participants and the reliability of the trial results). Remedial actions would focus on issues with the potential to have a substantial impact on the safety of the study participants or the reliability of the results.

The study will be conducted in accordance with the principles of International Conference on Harmonisation Guidelines for Good Clinical Research Practice (ICH-GCP) and relevant local, national and international regulations. Any serious breach of GCP in the conduct of the clinical trial will be handled in accordance with regulatory requirements. Prior to initiation of the study at each Local Clinical Centre (LCC), the Central Coordinating Office (CCO) or relevant Regional Coordinating Centre (RCC) will confirm that the LCC has adequate facilities and resources to carry out the study. LCC lead investigators and study staff will be provided with training materials.

In the context of this epidemic, visits to hospital sites are generally not appropriate as they could increase the risks of spreading infection, and in the context of this trial they generally would not influence the reliability of the trial results or the well-being of the participants. In exceptional circumstances, the CCO or RCC may arrange monitoring visits to LCCs as considered appropriate based on perceived training needs and the results of central statistical monitoring of study data.[9,10] The purpose of such visits will be to ensure that the study is being conducted in accordance with the protocol, to help LCC staff to resolve any local problems, and to provide extra training focussed on specific needs. No routine source data verification will take place.

##### 5.3 Data management

LCC clinic staff will use the bespoke study web-based applications for study management and to record participant data (including case report forms) in accordance with the protocol. Data will be held in central databases located at the CCO or on secure cloud servers. In some circumstances (e.g. where there is difficulty accessing the internet or necessary IT equipment), paper case report forms may be required with subsequent data entry by either LCC or CCO staff. Although data entry should be mindful of the desire to maintain integrity and audit trails, in the circumstances of this epidemic, the priority is on the timely entry of data that is sufficient to support reliable analysis and interpretation about treatment effects. CCO staff will be responsible for provision of the relevant web-based applications and for generation of data extracts for analyses.

All data access will be controlled by usernames and passwords, and any changes to data will require the user to enter their username and password. Staff will have access restricted to the functionality and data that are appropriate for their role in the study.

##### 5.4 Source documents and archiving

Source documents for the study constitute the records held in the study main database. These will be retained for at least 25 years from the completion of the study. Identifiable data will be retained only for so long as it is required to maintain linkage with routine data sources (see section 2.8), with the exception of children for whom such data must be stored until they reach 21 years old (due to the statute of limitations). The sponsor and regulatory agencies will have the right to conduct confidential audits of such records in the CCO and LCCs (but should mindful of the workload facing participating hospitals and the infection control requirements during this epidemic).

#### 6 OPERATIONAL AND ADMINISTRATIVE DETAILS

##### 6.1 Sponsor and coordination

The University of Oxford will act as the trial Sponsor. The trial will be coordinated by a Central Coordinating Office (CCO) within the Nuffield Department of Population Health staffed by members of the two registered clinical trials units – the Clinical Trial Service Unit and the National Perinatal Epidemiology Unit Clinical Trials Unit. The CCO will oversee Regional Coordinating Centres which will assist with selection of Local Clinical Centres

(LCCs) within their region and for the administrative support and monitoring of those LCCs. The data will be collected, analysed and published independently of the source of funding.

#### 6.2 Funding

This study is supported by grants to the University of Oxford from UK Research and Innovation/National Institute for Health Research (NIHR) and the Wellcome Trust, and by core funding provided by NIHR Oxford Biomedical Research Centre, the Wellcome Trust, the Bill and Melinda Gates Foundation, UK Foreign, Commonwealth and Development Office, Health Data Research UK, NIHR Health Protection Unit in Emerging and Zoonotic Infections and the Medical Research Council Population Health Research Unit, and NIHR Clinical Trials Unit Support Funding.

#### 6.3 Indemnity

The University has a specialist insurance policy in place which would operate in the event of any participant suffering harm as a result of their involvement in the research (Newline Underwriting Management Ltd, at Lloyd's of London). In the UK, NHS indemnity operates in respect of the clinical treatment that is provided.

#### 6.4 Local Clinical Centres

The study will be conducted at multiple hospitals (LCCs) within each region. At each LCC, a lead investigator will be responsible for trial activities but much of the work will be carried out by medical staff attending patients with COVID-19 within the hospital and by hospital research nurses, medical students and other staff with appropriate education, training, and experience. Where LCCs plan to recruit children the principal investigator will co-opt support from a local paediatrician and/or neonatologists to oversee the management of children and infants in the trial.

#### 6.5 Supply of study treatments

For licensed treatments (e.g. corticosteroids, oseltamivir) all aspects of treatment supply, storage, and management will be in accordance with standard local policy and practice for prescription medications. Treatments issued to randomised participants will be by prescription. Such study treatments will not be labelled other than as required for routine clinical use. They will be stored alongside other routine medications with no additional monitoring. No accountability records will be kept beyond those used for routine prescriptions.

For unlicensed treatments, manufacture, packaging, labelling and delivery will be the responsibility of the pharmaceutical company and, in the UK, the Department of Health and Social Care. Each LCC will maintain an accountability log and will be responsible for the storage and issue of study treatment. If treatments require storage at a specific temperature, LCCs can use existing temperature-controlled facilities and associated monitoring. Treatment issue to randomised participants will be in accordance with local practice (and may be in line with the processes required for routine prescriptions or compassionate use).

Treatment will be issued to randomised participants by prescription.

#### 6.6 End of trial

The end of the scheduled treatment phase is defined as the date of the last follow-up visit of the last participant. In the UK, it is intended to extend follow-up for a year or more beyond the final study visit through linkage to routine medical records and central medical databases. The end of the study is the date of the final data extraction from NHS Digital (anticipated to be 10 years after the last patient is enrolled).

##### 6.7 Publications and reports

The Trial Steering Committee will be responsible for drafting the main reports from the study and for review of any other reports. In general, papers initiated by the Trial Steering Committee (including the primary manuscript) will be written in the name of the RECOVERY Collaborative Group, with individual investigators named personally at the end of the report (or, to comply with journal requirements, in web-based material posted with the report).

The Trial Steering Committee will also establish a process by which proposals for additional publications (including from independent external researchers) are considered by the Trial Steering Committee. The Trial Steering Committee will facilitate the use of the study data and approval will not be unreasonably withheld. However, the Trial Steering Committee will need to be satisfied that any proposed publication is of high quality, honours the commitments made to the study participants in the consent documentation and ethical approvals, and is compliant with relevant legal and regulatory requirements (e.g. relating to data protection and privacy). The Trial Steering Committee will have the right to review and comment on any draft manuscripts prior to publication.

##### 6.8 Substudies

Proposals for substudies must be approved by the Trial Steering Committee and by the relevant ethics committee and competent authorities (where required) as a substantial amendment or separate study before they begin. In considering such proposals, the Trial Steering Committee will need to be satisfied that the proposed substudy is worthwhile and will not compromise the main study in any way (e.g. by impairing recruitment or the ability of the participating hospitals to provide care to all patients under their care).

### 7 VERSION HISTORY

| Version number | Date | Brief Description of Changes |
|---|---|---|
| 1.0 | 13-Mar-2020 | Initial version |
| 2.0 | 21-Mar-2020 | Addition of hydroxychloroquine. Administrative changes and other clarifications. |
| 3.0 | 07-Apr-2020 | Extension of eligibility to those with suspected COVID-19<br>Addition of azithromycin arm.<br>Addition of inclusion of adults who lack permanently lack capacity.<br>Change to primary outcome from in-hospital death to death within 28 days of randomisation. |
| 4.0 | 14-Apr-2020 | Addition of second randomisation to tocilizumab vs. standard of care among patients with progressive COVID-19. |
| 5.0 | 24-Apr-2020 | Addition of children to study population. |
| 6.0 | 14-May-2020 | Addition of convalescent plasma |
| 7.0 | 18-Jun-2020 | Allowance of randomisation in part B of main randomisation without part A.<br>Removal of hydroxychloroquine and dexamethasone treatment arms. |
| 8.0 | 03-Jul-2020 | Removal of lopinavir-ritonavir<br>Addition of intravenous immunoglobulin arm for children<br>Changes to corticosteroid dosing for children.<br>Addition of baseline serum sample in convalescent plasma randomisation |
| 9.0 | 10-Sep-2020 | Addition of synthetic neutralizing antibodies<br>Additional baseline data collection<br>Addition of countries outside UK |
| 9.1 | 18-Sep-2020 | Addition of information about vaccination of children of pregnant mothers receiving REGN10933+REGN10987 |
| 9.2 [not submitted in UK] | 15-Oct-2020 | Additional information for countries outside UK |
| 10.0 | 26-Oct-2020 | Addition of main randomisation part C<br>General updates to avoid duplication and improve clarity |
| 10.1 | 01-Nov-2020 | Additional information for pregnant women |
| 11.0 | 19-Nov-2020 | Addition of colchicine to main randomisation part A<br>Removal of azithromycin from main randomisation part A<br>Change in randomisation ratio in main randomisation part A from 2:1 to 1:1 |
| 11.1 | 21-Nov-2020 | Clarification of colchicine age thresholds |
| 11.2 [not submitted in UK] | 01-Dec-2020 | Addition of modified aspirin dose if 150mg not available |
| 12.0 | 10-Dec-2020 | Allow second randomisation of children without first randomisation |
| 12.1 | 16-Dec-2020 | Clarification of change in V12.0 |
| 13.0 | 26-Jan-2021 | Addition of baricitinib and anakinra (and change to allocation ratio in second randomisation for children); addition of pregnancy test for women of child-bearing potential (and change to colchicine eligibility);<br>removal of tocilizumab for adults; removal of convalescent plasma and additional assessment of antibody-based therapy; addition of dexamethasone as substitute if methylprednisolone unavailable |
| 14.0 | 15-Feb-2021 | Addition of Early Phase Assessments; the inclusion of dimethyl fumarate for initial early phase assessment; restriction of main randomisation part B to children with COVID-19 pneumonia; modification of barictinib and tocilizumab co-administration guidance |
| 15.0 | 12-Apr-2021 | Removal of aspirin and colchicine; addition of infliximab and high-dose corticosteroids (ex-UK only) |
| 15.1 [not submitted in UK] | 18-May-2021 | Addition of South Africa |

| Version number | Date | Brief Description of Changes |
|---|---|---|
| 16.0 | 05-Jul-2021 | Removal of REGN-COV2 and main randomisation part B<br>Removal of infliximab from main randomisation part E (and associated endemic infection monitoring section)<br>Addition of empagliflozin as main randomisation part F and metabolic outcomes<br>Addition of India, Sri Lanka and Pakistan |
| V16.1 | 08-Jul-2021 | Clarification of design in introduction |
| V17.0 | 06-Aug-2021 | Addition of additional exclusion criteria and safety monitoring for empagliflozin arm<br>Removal of corticosteroids and intravenous immunoglobulin in main randomisation part A (for children) |
| V17.1 | 10-Aug-2021 | Clarification of design for children |
| V18.0 | 13-Oct-2021 | Update to consent section<br>Change in primary outcome and sample size for DMF comparison<br>Clarification of eligibility for PIMS-TS randomisation<br>Removal of 3 month follow-up form for non-UK countries |
| V18.1 | 24-Oct-2021 | Clarification of witnesses for consent of children |
| V19.0 | 12-Nov-2021 | Addition of baloxavir marboxil, oseltamivir, and low-dose corticosteroids as randomised comparisons each vs. usual care alone for patients with influenza (in UK only).<br>Removal of early phase assessment of dimethyl fumarate.<br>Updated statistical analysis section to align with statistical analysis plan and include influenza analyses. |
| V19.1 | 16-Nov-21 | Clarification of baloxavir and weight eligibility |
| V20.0 | 29-Nov-21 | Removal of baricitinib.<br>Extension of COVID-19 high-dose corticosteroid and empagliflozin comparisons to other countries. |
| V21.0 | 17-Dec-21 | Addition of sotrovimab and molnupiravir.<br>Addition of baseline and follow-up samples.<br>Re-randomisation of patients recruited >6 months ago. |
| V21.1 | 19-Dec-21 | Clarifications post-REC review. |
| V22.0 | 19-Jan-22 | Addition of Paxlovid. (Not implemented.) |
| V23.0 | 08-Mar-22 | Clarifications following MHRA review. UKOSS added to section 3.1.3. Extension of molnupiravir to other countries. Removal of tocilizumab/anakinra for PIMS-TS. |
| 23.1 | 15-Mar-22 | Correction of footnotes |

#### Completed comparisons

The last version of the protocol to include the IMP is shown in the table above.

| IMP | Citation |
|---|---|
| Hydroxycholoroquine | New Engl J Med 2020; 383: 2030-40 |
| Dexamethasone (COVID-19) | New Engl J Med 2021; 384: 693-704 |
| Lopinavir-ritonavir | Lancet 2020; 396: 1345-1352 |
| Azithromycin | Lancet 2021; 397: 605-12 |
| Convalescent plasma | Lancet 2021; 397: 2049-59 |
| Tocilizumab | Lancet 2021; 397: 1637-1645 |
| Aspirin | Lancet 2022; 397: 143-151 |
| Colchicine | Lancet Resp Med 2021; 9: 1419-26 |
| REGN-COV2 | Lancet 2022; 399: 665-76 |
| Methylprednisolone (PIMS-TS) | Analysis ongoing |
| Intravenous immunoglobulin (PIMS-TS) | Analysis ongoing |
| Tocilizumab (PIMS-TS) | Follow-up ongoing |
| Anakinra (PIMS-TS) | Follow-up ongoing |
| Dimethyl fumarate | Analysis ongoing |
| Baricitinib | Medrxiv: 10.1101/2022.03.02.22271623v1 |

### 8 APPENDICES

#### 8.1 Appendix 1: Information about the treatment arms

All patients will receive usual care in the participating hospital.

**Corticosteroids:** RECOVERY is assessing the effects of corticosteroids in two different contexts: higher dose *vs* usual care in adults with COVID-19 and hypoxia; and lower dose dexamethasone in adults with influenza and hypoxia (UK only).

Favourable modulation of the immune response is considered one of the possible mechanisms by which corticosteroids might be beneficial in the treatment of severe acute respiratory coronavirus infections, including influenza, COVID-19, SARS and MERS. Common to severe cases of these infections is the presence of hypercytokinemia and the development of acute lung injury or acute respiratory distress syndrome (ARDS).[11-14] Pathologically, diffuse alveolar damage is found in patients who die from these infections.[15]

*Corticosteroids in influenza*
RECOVERY and other randomised trials have now demonstrated the benefit of corticosteroids in hypoxic COVID-19 patients.[16,17] However, the potential role of corticosteroids in severe influenza remains uncertain, with differing practices and controversy. Whilst observational studies report higher mortality associated with the use of corticosteroids in severe influenza, these studies are prone to biases, with a major concern being confounding by indication (the propensity to use corticosteroids only in the more severe patients as a rescue therapy).[18] In practice, use of corticosteroids in severe influenza is variable and widespread.[19] This therapeutic dilemma will only be resolved through an adequately powered randomised trial.

*Corticosteroids in COVID-19*
RECOVERY showed that dexamethasone at a dose of 6mg once daily for ten days or until discharge (whichever happens earliest) provided a significant reduction in mortality. Combining the IL-6 inhibitor tocilizumab with low dose dexamethasone resulted in a further reduction in mortality. This raises the question whether simply increasing the dose of corticosteroid could confer a similar clinical benefit to that of adding tocilizumab, but at substantially lower cost. Of note, even with dexamethasone 6mg and tocilizumab, mortality remained high at 29%. Although other randomised clinical trials in critically ill COVID-19 patients have used higher doses of dexamethasone (20mg once daily for five days followed by 10mg once daily for a further five days) and reported clinical benefit, these doses have not been compared with the lower dose used in RECOVERY. There is, therefore, uncertainty regarding the optimal dose of corticosteroids in moderate to severe COVID-19.Uncertainty remains about whether higher doses of corticosteroids may provide additional benefit in adults with hypoxia hospitalised with COVID-19.

Unlike lower doses, higher doses (>15mg dexamethasone) would completely saturate cytosolic glucocorticoid receptors and have enhanced non-genomic effects. In conditions where rapid control of inflammatory processes are required, short-term, high to very high doses of corticosteroids are used e.g.

- Sepsis dexamethasone 7.5 - 15mg equivalent daily[20]

- ARDS: dexamethasone 20mg for five days followed by 10mg for five days[21]
- Bacterial meningitis: dexamethasone 40mg daily for four days[22]
- Tuberculous Meningitis dexamethasone 0.4mg/kg/day for 7 days then reducing over 8 weeks.[23]
- Rheumatoid arthritis flare: dexamethasone 120mg pulse therapy.[24]
- Community acquired pneumonia: dexamethasone 0.6mg/day for 2 days and methyl prednisolone 200mg /day then 80mg /day for 10 days.[25]

**Empagliflozin:** Sodium glucose co-transporter 2 inhibitors (SGLT-2i) decrease glucose and insulin levels, and shift energy metabolism to an increased reliance on lipid oxidation, with a reduced reliance on glucose, and inhibition of glycolysis.[26] This mechanism may be particularly important in COVID-19, as SARS-CoV-2 may depend on the glycolytic pathway for its replication, stimulating lipogenesis, which appears to be one of the key drivers of cellular damage.[27,28] SGLT-2i rapidly improve endothelial function, possibly because of reduced oxidative stress.[29] SGLT-2i have significant anti-inflammatory effects, reducing levels of C-reactive protein and interleukin-6.[30] Experimental studies have also shown reduced activation of the NLRP3 inflammasome.[31] SGLT-2i increase erythropoiesis resulting in increased haematocrit,[32,33] and together with improved endothelial function[29] may improve oxygen delivery to tissues. Moreover, SGLT-2i result in reduced extracellular volume in patients with fluid overload,[34,35] and appear to reduce pulmonary artery pressure in patients with heart failure rapidly,[36] leading to haemodynamic decongestion. Thus, SGLT-2i may favourably affect multiple processes, including but not limited to energy metabolism, endothelial function, oxidative stress, inflammation and autophagy, which are dysregulated during a major acute illness such as COVID-19. The DARE-19 trial compared dapagliflozin 10 mg with placebo for 30 days among 1250 patients admitted to hospital with COVID-19 who had mild hypoxia ($SpO_2$ ≥94% on ≤5 L/min oxygen) and at least one risk factor (hypertension, type 2 diabetes mellitus, atherosclerotic cardiovascular disease, heart failure or chronic kidney disease).[37] The treatment was well tolerated (11% discontinued prematurely with similar proportion in treatment and placebo group). The hazard ratio for the co-primary outcome of organ failure (non-invasive or invasive ventilation, requirement for cardiovascular support or new/worsened heart failure, doubling of creatinine or dialysis) or death was 0.80 (95% CI 0.58-1.10; 70 *vs* 86 events).[38] Although this trial lacked statistical sensitivity, it supports the rationale for a larger trial.

**Sotrovimab [UK only]:** Sotrovimab (VIR-7831) is a neutralising monoclonal antibody targeting the SARS-CoV-2 spike glycoprotein receptor binding domain. It was identified by screening antibodies from a patient who had been infected during the 2003 SARS-CoV-1 outbreak, and its ability to also neutralise SARS-CoV-2 implies that its binding site is highly conserved, maybe meaning mutational escape will be difficult.[39] The Fc portion of the parent antibody has been modified to extend sotrovimab's half-life to around 49 days. It is given as a single intravenous dose and been well tolerated in clinical studies, although occasional serious hypersensitivity reactions have occurred.

It is licenced in the UK for the treatment of COVID-19 in patients who do not require oxygen and are at high risk of developing severe disease (at a 500 mg dose). The COMET-ICE trial, conducted in 583 such patients, showed that when given within five days of symptom onset it reduced the risk of hospitalisation by 85%, from 7% in the control group to 1% in the sotrovimab group.[40] Evidence in hospitalised patients is limited, and the sotrovimab arm of ACTIV-3 was stopped due to futility after recruiting 344 participants, although no safety concerns were raised.[41] However, by recruiting around 10,000 patients, RECOVERY subsequently showed that another neutralising monoclonal antibody treatment (casirivimab+imdevimab) reduced mortality by 20% in hospitalised patients who were anti-spike antibody negative at baseline.

The Omicron SARS-CoV-2 variant that emerged in late 2021 has multiple spike protein mutations, which have led to its rapid expansion in immune populations. These also appear to cause near complete loss of neutralising activity by the monoclonal antibodies in casirivimab+imdevimab,[42] and reduce the neutralising activity of Sotrovimab about 10-fold.[43,44] Data comparing the peak and day 29 concentrations following 2.4 g casirivimab+imdevimab and 500 mg Sotrovimab demonstrate much lower concentrations of Sotrovimab.[45] These pharmacodynamics and pharmacokinetic considerations underly the selection of a 1000 mg dose in this trial. The published safety of Sotrovimab and higher doses of other anti-spike human monoclonal antibodies (including the 8g dose of casirivimab+imdevimab used in RECOVERY) do not suggest a safety concern with this increased dose.

**Molnupiravir [UK only]:** Molnupiravir is a prodrug of the ribonucloside analogue N-hydroxycytidine (NHC), being rapidly converted into this form in plasma after absorption. NHC is then converted into the active triphosphate form in host cells by endogenous kinases. The SARS-CoV-2 viral RNA polymerase incorporates this into nascent viral RNA, resulting in copying errors that accumulate every replication cycle, ultimately preventing replication by a mechanism known as error catastrophe. This molecular target is conserved between Coronaviruses, and appears to have a high genetic barrier to resistance.[46] Molnupiravir is given orally and has been well tolerated in clinical studies so far, with infrequent reports of gastrointestinal and allergic reactions.

Molnupiravir is licensed in the United Kingdom for the treatment of mild-moderate COVID-19 within 5 days of symptom onset. In the MOVe-OUT trial of 1433 such patients it reduced the risk of hospitalisation or death by 30%, from 9.7% in the placebo group to 6.8% in molnupiravir group.[47] Evidence in hospitalised patients is limited, and the MOVe-IN trial randomised patients 1:1:1:1 to placebo vs. molnupiravir at 3 different doses (200mg, 400mg, 800mg). This study was abandoned after recruiting 304 inpatients as the manufacturer decided it was unlikely to demonstrate clinical benefit, although no safety concerns were raised.[48] However, the study was underpowered to identify moderate but important benefits in hospitalised patients, so a larger trial is needed.

**Paxlovid [UK only]:** Paxlovid is a combination of PF-07321332 (nirmatrelvir) and ritonavir. Nirmatrelvir is a 3-chymotrypsin-like protease inhibitor which inhibits cleavage of polyproteins involved in viral replication.[49] It is packaged with ritonavir which inhibits its CYP3A-dependent metabolism and hence increases the plasma concentration of nirmatrelvir. It is approved in the UK for the treatment of adults with COVID-19 who do not require supplemental oxygen and are at increased risk of progression to severe COVID-19.[50]

Its approval is based on the interim analysis of the EPIC-HR trial in which 2246 participants with COVID-19 (symptom onset ≤5 days previously) were randomised to receive Paxlovid (300/100 mg) or placebo twice daily for 5 days. The primary outcome is the proportion of participants with COVID-19 related hospitalisation or death within 28 days of randomisation.

In the interim analysis, 8/1037 (0.8%) allocated Paxlovid *vs* 66/1046 (6.3%) allocated placebo.[51] In an interim analysis of 774 participants, adverse events were similar between the two groups: 19% among those allocated Paxlovid *vs* 21% among those allocated placebo. Most were mild; only 1.7% *vs* 6.6% were serious and 2.1% *vs* 4.1% led to discontinuation.[52] SARS-CoV-2 main protease polymorphisms associated with reduced sensitivity to nirmatrelvir have been identified.[51] Their frequency and clinical significance is not yet known. Cross-resistance between nirmatrelvir and anti-SARS-CoV-2 monoclonal antibodies, molnupiravir or remdesivir are not expected given their different mechanisms of action.

**Baloxavir marboxil [UK only]:** Baloxavir marboxil is a cap-dependent endonuclease (CEN) inhibitor. CEN is an influenza virus-specific enzyme in the polymerase acidic subunit of the viral RNA polymerase complex. Through its action on CEN, baloxavir inhibits the transcription of influenza virus genomes resulting in inhibition of influenza A and B virus replication. It is approved in the USA, Japan, Australia, Europe, and the United Kingdom for the treatment of uncomplicated influenza and for post-exposure prophylaxis in individuals aged 12 years and older. Baloxavir is given in 2 oral doses (on day 1 and day 4) and is well tolerated, with allergic reactions being the only reported adverse reactions.

Baloxavir is not approved for the treatment of complicated influenza. A phase III placebo-controlled trial of baloxavir in adults hospitalised with severe influenza (Flagstone NCT03684044) did not find a significant reduction in the primary endpoint of time to clinical improvement (personal communication, Roche). However, time to clinical improvement, time to clinical response, influenza related complications, mortality, and time to cessation of viral shedding were all in favour of baloxavir. Fewer adverse events were observed in the baloxavir arm than in the standard of care arm. The Flagstone trial was small, comparing 214 subjects who received baloxavir with 125 who received usual care alone, and a larger study is need to determine whether baloxavir has modest but clinically relevant benefit in patients hospitalised with influenza.

**Oseltamivir [UK only]:**

The neuraminidase inhibitors (oseltamivir and zanamivir) are influenza specific antivirals that have been shown in randomised controlled trials to improve outcomes in uncomplicated influenza and to be effective as post-exposure prophylaxis. They have not, however, been shown to be effective in patients hospitalised with severe influenza. Although observational studies have reported clinical benefit in patients hospitalised with severe influenza, there are no randomised controlled trial data. Consequently, the use of neuraminidase inhibitors in this patient population is variable. A randomised controlled trial of neuraminidase inhibitors in patients hospitalised with severe influenza has been recommended by an expert group convened by the Academy of Medical Sciences and the Wellcome Trust, and most clincians would welcome such a trial.[53,54] The duration of treatment (5 days, or 10 days if the patient is immunosuppressed in the opinion of the managing clinician) is the same as that used in clinical practice and in the Summary of Product Characteristics.

#### 8.2 Appendix 2: Drug specific contraindications and cautions

**Corticosteroid**

Contraindications:

- Known contra-indication to short-term corticosteroid.
- Patients with suspected or confirmed influenza co-infection are not eligible for the high-dose dexamethasone comparision for COVID-19 (Randomisation part E).
- Patients in the UK with suspected or confirmed SARS-CoV-2 co-infection are not eligible for the low-dose dexamethasone comparison for influenza infection because of the proven benefits of dexamethasone in COVID-19 (Randomisation part I).
- Patients eligible for the Paxlovid comparison (Randomisation part L) will be excluded by the randomisation system from the high-dose dexamethasone comparison for COVID-19 (Randomisation part E) in view of the potential interaction between Paxlovid and dexamethasone.
- Current use of Paxlovid, ritonavir or other potent CYP3A inhibitors.

Cautions:

- Endemic infections may be screened for as required by local practice.
- Other immunomodulatory therapies are not contraindicated, but investigators should consider the total burden of therapy (eg, combining IL-6 receptor antagonist therapy with high-dose dexamethasone).

**Empagliflozin**

Contraindications:

- Type 1 diabetes mellitus (or post-pancreatectomy diabetes)
- Pregnancy and breast-feeding
- History of ketoacidosis
- Other patients with diabetes: blood ketones ≥1.5 mmol/L (or urine ketones ≥2+ if near-patient testing for blood ketones unavailable). Such patients are eligible once their ketosis has resolved.

Cautions:

- Participants with diabetes allocated empagliflozin should have regular checks of blood ketones (or urine ketones if blood ketone testing is unavailable)[q]. Blood ketones should be checked twice daily or urine ketones daily (or if clinical concern). If blood ketones rise ≥1.5 mmol/L (or urine ketones ≥2+), clinicians should:
  - Ensure adequate fluid and calorific intake
  - Consider increasing insulin dose (if on insulin)
  - Inform local diabetes team (if available) and treat ketosis using local protocols
  - Consider discontinuing empagliflozin until ketosis resolves
- Clinicians should consider temporarily discontinuing empagliflozin in participants with diabetes mellitus who cannot maintain oral calorific intake (until nutrition is restored)
- Clinicians should be aware of “euglycaemic ketoacidosis” which occurs with empagliflozin and should check ketones (ideally blood) if this is suspected (e.g. unexplained metabolic acidosis)

[q] These are near-patient tests and no sample will be retained for research purposes.

- Empagliflozin does not cause hypoglycaemia alone, but may do so in combination with insulin or insulin secretagogues. Doses of these other medications may need to be temporarily modified while the participant is taking empagliflozin
- Empagliflozin causes an osmotic diuresis so careful fluid balance assessment is required
- Empagliflozin increases the risk of mycotic genital infections (e.g. candidiasis) which are usually easily treated with topical therapy. It is unclear whether it causes Fournier's gangrene (a very rare genital infection), but clinicians should be aware.

**Sotrovimab**
Contraindications:
- Weight <40kg (if <18 years old; no weight restriction for adults)
- Known hypersensitivity to sotrovimab or the drug product excipients

Cautions: no dose adjustment for kidney or liver function is required.

**Molnupiravir**
Contraindications:
- Age <18 years
- Pregnancy or breast-feeding. Women of child-bearing potential should be advised not to get pregnant while taking molnupiravir or for 4 days after completing the course
- Known hypersensitivity to molnupiravir or its excipients
- Prior treatment with molnupiravir during the index illness

Cautions: no dose adjustment for kidney or liver function is required.

**Paxlovid**
Contraindications:
- Age <18 years
- Severe hepatic impairment (Child-Pugh class C)
- Severe renal impairment (eGFR <30 mL/min/1.73m$^2$)
- First trimester (i.e. first 12 weeks) of pregnancy
- Prior treatment with Paxlovid during the index illness
- Known hypersensitivity to nirmatrelvir (PF-07321332) or ritonavir (including hereditary problems of galactose intolerance, total lactase deficiency or glucose-galactose malabsorption)
- Concomitant therapy with drugs that are highly dependent on CYP3A for clearance and for which elevated plasma concentrations are associated with serious reactions.
    - α1-adrenoreceptor antagonist (afluzosin)
    - Analgesics (pethidine, piroxicam, propoxyphene)
    - Anti-anginal (ranolazine)
    - Anti-arrhythmics (amiodarone, bepridil, dronaderone, encainide, flecainide, propafenone, quinidine)
    - Antibacterials (fusidic acid)
    - Anticancer (neratinib, venetoclax)
    - Anti-gout (colchicine)
    - Antihistamine (astemizole, terfenadine)
    - Antipsychotics (lurasidone, pimozide, clozapine, quietiapine)
    - Ergot derivatives (dihydroergotamine, ergonovine, ergotamine, methylergonovine)

- Gastrointestinal motility agent (cisapride)
- Lipid modifying agents (lovastatin, simvastatin, lomitapide)
- PDE5 inhibitors (avanafil, vardenafil, sildenafil)
- Sedatives (clorazepate, diazepam, estazolam, flurazepam, triazolam, oral midazolam)
- High-dose dexamethasone (>6 mg base once daily)

- Concomitant therapy with drugs that are potent CYP3A inducers (which may reduce plasma PF-07321332/ritonavir concentrations):
    - Anticancer (apalutamide)
    - Anticonvulsants (carbamazepine, phenobarbital, phenytoin)
    - Antimycobacterials (rifampicin)
    - Herbal products (St John's Wort)

Cautions:

- Since ritonavir may decrease the efficacy of combined oral contraceptives, women using them should be advised to use effective alternative contraception or an additional barrier method until after one complete menstrual cycle after stopping.
- The necessity of using other drugs metabolised by CYP3A (or which induce or inhibit CYP3A) should be reviewed.[r]
- 
- Patients with moderate renal impairment (eGFR ≥30 <60 mL/min/1.73m$^2$) should receive 150/100 mg twice daily (ie, one PF-07321332 tablet and one ritonavir tablet twice daily). Local pharmacists should remove one PF-07321332 tablet from each dose in the packet provided to the participant (see pharmacy manual at https://www.recoverytrial.net/for-site-staff/pharmacy for further detail).

Managing clinicians may consider if it is appropriate to temporarily withhold contraindicated concomitant medication while receiving Paxlovid or consider alternatives. The risks and benefits of doing so should be explained to the participant. Clear plans should be made about restarting such treatment and – if necessary – any checks that need to be made beforehand. These plans should be communicated to the participant and their general practitioner in the discharge summary.

**Baloxavir Marboxil**

Contraindications:

- Weight <40kg (regardless of age)
- Known hypersensitivity to baloxavir marboxil or the drug product excipients
- Participants who have received baloxavir marboxil for the current influenza infection

**Oseltamivir**

Contraindications:

- Known hypersensitivity to oseltamivir or the drug product excipients
- Participants who have received oseltamivir for the current influenza infection

Cautions:

- Dose should be reduced in presence of renal impairment
    - eGFR ≥30 mL/min/1.73m$^2$: dose as in normal renal function (75 mg twice daily)

[r] A list is available at https://www.covid19-druginteractions.org/. Please note these lists may not be exhaustive.

- eGFR ≥10 <30 mL/min/1.73m$^2$: 75 mg once daily
- eGFR <10 mL/min/1.73m$^2$: 75 mg as a single dose on day 1

- Dose should be reduced for adult patients weighing <40 kg to 60 mg twice daily

RECOVERY
Randomised Evaluation of COVID-19 Therapy

#### 8.3 Appendix 3: Paediatric dosing information

Children (aged <18 years old) will be recruited in the UK only.

**Randomisation of children with COVID-19 Pneumonia (Patients <12 years of age will NOT be eligible)**

| Arm | Route | Weight | Dose |
|---|---|---|---|
| **No additional treatment** | - | - | - |
| **Sotrovimab** | Intravenous | Children <12 years old excluded | |
| | | <40 kg | Excluded regardless of age |
| | | ≥40 kg | 1000 mg intravenous in 100 mL of 0.9% NaCl or 5% dextrose over 1 hour |

#### Influenza Randomisations

<table>
<tr><th>Arm</th><th>Route</th><th>Weight/Age</th><th>Dose</th></tr>
<tr><td rowspan="3">Oseltamivir<br>- 30, 45 and 75 mg capsules<br>- Oral suspension [a]</td><td rowspan="3">Oral<br><u>or</u><br>Other enteral routes</td><td>Less than 36 weeks corrected gestational age</td><td>1 mg/kg twice daily for 5 days [b]</td></tr>
<tr><td>0 - 12 months (≥36 weeks corrected gestational age)</td><td><table><tr><th>Weight (kg)</th><th>Dose</th></tr><tr><td><10</td><td>3 mg/kg twice daily for 5 days [b]</td></tr><tr><td>≥ 10</td><td>30 mg twice daily for 5 days [b]</td></tr></table></td></tr>
<tr><td>≥ 1 year</td><td><table><tr><th>Weight (kg)</th><th>Dose</th></tr><tr><td><10</td><td>3 mg/kg twice daily for 5 days [b]</td></tr><tr><td>≥ 10 to 15</td><td>30 mg twice daily for 5 days [b]</td></tr><tr><td>> 15 to 23</td><td>45 mg twice daily for 5 days [b]</td></tr><tr><td>> 23 to 40</td><td>60 mg twice daily for 5 days [b]</td></tr><tr><td>> 40</td><td>75 mg twice daily for 5 days [b]</td></tr></table>Those within significant renal impairment (CrCl 10 - 30 mL/min) should receive once daily dosing. Those with CrCl <10 ml/min should receive only a single dose on day 1.</td></tr>
<tr><td>Baloxavir marboxil<br>- 20 and 40 mg tablets</td><td>Oral<br><u>or</u><br>Other enteral routes</td><td>≥ 12 years old</td><td><table><tr><th>Weight (kg)</th><th>Dose</th></tr><tr><td><40</td><td>Not eligible</td></tr><tr><td>≥40 < 80</td><td>40 mg on day 1 and day 4</td></tr><tr><td>≥ 80</td><td>80 mg on day 1 and day 4</td></tr></table></td></tr>
<tr><td rowspan="2">Low dose corticosteroids</td><td rowspan="2">Oral<br><u>or</u><br>Other enteral routes<br><u>or</u><br>Intravenous</td><td>Less than 36 weeks corrected gestational age</td><td>Hydrocortisone (IV)<br>0.5 mg/kg every 12 hours for 7 days and then 0.5mg/kg once daily for 3 days</td></tr>
<tr><td>≥0 month (≥36 weeks corrected gestational age)</td><td>Dexamethasone:<br>150 micrograms/kg (as base) once daily (max: 6 mg once daily) for 10 days (or until discharge if sooner)</td></tr>
</table>

[a] Public Health England advises that oseltamivir oral suspension should be reserved for children under the age of 1 year. Children over 1 year of age, those with swallowing difficulties, and those receiving nasogastric oseltamivir, should use capsules which can be opened and mixed into an appropriate sugary liquid.
[b] 10 days if immunocompromised

#### 8.4 Appendix 4: Use of IMPs in pregnant and breastfeeding women

All trial drugs (except empagliflozin, sotrovimab, molnupiravir, Paxlovid and baloxavir) have been used in pregnant women with pre-existing medical disorders where benefits outweigh the risks to fetus or woman, including in the first trimester. The existing data related to each drug is summarised below. The potential inclusion of any pregnant women should be discussed with a consultant obstetrician (or obstetric physician) and all consent discussions should be documented in the medical records.

##### Corticosteroids

Prednisolone or, in women unable to take oral medicine, hydrocortisone or methylprednisolone are recommended instead of dexamethasone treatment in light of accumulating evidence that repeated doses of dexamethasone have deleterious effects on long-term neurodevelopment of the fetus.[55-57] While 90% dexamethasone is transferred transplacentally to the fetus, both hydrocortisone and prednisolone are converted by 11β-hydroxysteroid dehydrogenase to inactive glucocorticoids and considerably less drug is transferred to the fetus. Glucocorticoids can worsen maternal glycaemic control, so blood glucose should be checked and managed appropriately. Otherwise there is no convincing evidence that prednisolone use is associated with increased rates of adverse pregnancy outcomes when taken in the first trimester or later pregnancy.[58] Very low concentrations of prednisolone enter breastmilk. There is a paucity of data about pharmacological use of hydrocortisone, but it is likely that this is also safe when breastfeeding,[58] as also reviewed in the Lactmed database (www.ncbi.nlm.nih.gov/books/NBK501076/). Prednisolone (or hydrocortisone) should be used in breastfeeding women, in preference to dexamethasone.

##### Empagliflozin

Empagliflozin is not recommended for use in pregnant or breastfeeding women. Empagliflozin will only be included in the randomisation of women of child-bearing potential if they have had a negative pregnancy test since admission.

##### Sotrovimab

There are no data from the use of sotrovimab in pregnant women. Since sotrovimab is a human immunoglobulin G animal studies have not been evaluated with respect to reproductive toxicity. No off-target binding was detected in a cross-reactive binding assay using a protein array enriched for human embryofetal proteins. Since sotrovimab is a human immunoglobulin G, it has the potential for placental transfer from the mother to the developing foetus. The potential treatment benefit or risk of placental transfer of sotrovimab to the developing foetus is not known. Sotrovimab may be used during pregnancy where the expected benefit to the mother justifies the risk to the foetus.

There are no data on the excretion of sotrovimab in human milk. The potential treatment benefit or risk to the newborn or infants via breastfeeding is not known. Decisions on whether to breastfeed during treatment or to abstain from sotrovimab therapy should take into account the benefit of breast-feeding for the child and the benefit of therapy for the woman.

##### Molnupiravir

Molnupiravir is not recommended for use in pregnant or breastfeeding women. Molnupiravir will only be included in the randomisation of women of child-bearing potential if they have had a negative pregnancy test since admission.

**Paxlovid**
Preclinical animal reproductive toxicity studies have not identified adverse effects on fetal morphology or embryo-fetal viability in rat or rabbit models with doses of nirmatrelvir up to 12 times the human dose (equivalence based on predicted AUC concentrations). The offspring of pregnant rabbits administered 24 times the equivalent human dose, lower fetal body weights were observed but evidence of maternal toxicity was described (impact on weight gain/food consumption).[52] There is a large amount of published evidence relating to the safety of ritonavir in human pregnancy, collected from antiretroviral and HIV/AIDS pregnancy registries. Overall, these data do not provide compelling evidence that ritonavir use in the first trimester is associated with an increased risk of malformation above the expected background rate of 2-3%. As Paxlovid has not previous been given to pregnant women, women in the first trimester of pregnancy will be excluded from this comparison.

**Baloxavir marboxil**
There are no data from the use of baloxavir marboxil in pregnant women. Animal studies do not indicate direct or indirect harmful effects with respect to reproductive toxicity. Baloxavir treatment may be of particular benefit to pregnant women with influenza, as they are at increased risk of developing severe disease. Preclinical animal models of exposure in pregnancy do not provide evidence of adverse embryo-fetal effects at doses up to five and seven times the human therapeutic dose respectively. The risk of harm from baloxavir in pregnancy is likely to be low given the animal model data, together with the therapeutic target for baloxavir being a virus specific enzyme. It is unknown whether baloxavir marboxil or baloxavir are excreted in human milk, and baloxivir may be considered.

**Oseltamivir**
There are observational data on the use of oseltamivir in pregnant women including >1000 women exposed during the first trimester. These studies found no evidence of adverse embryo-fetal effects. Oseltamivir is currently used in pregnant women. Its use may also be considered in breastfeeding women: it is excreted in breast milk but at low concentrations that would be subtherapeutic dose to the infant.

#### 8.5 Appendix 5: Organisational Structure and Responsibilities

**Chief Investigator**
The Chief Investigator has overall responsibility for:

(i) Design and conduct of the Study in collaboration with the Trial Steering Committee;
(ii) Preparation of the Protocol and subsequent revisions.

**Trial Steering Committee**
The Trial Steering Committee (see below for list of members) is responsible for:

(i) Agreement of the final Protocol and the Data Analysis Plans;
(ii) Reviewing progress of the study and, if necessary, deciding on Protocol changes;
(iii) Review and approval of study publications and substudy proposals;
(iv) Reviewing new studies that may be of relevance.

**International Steering Committee**
The International Steering Committee (see below for list of members) is responsible for:

(i) Reviewing progress of the study in sites outside the UK;
(ii) Review of study publications and substudy proposals;
(iii) Considering potential new therapies to be included in sites outside the UK;
(iv) Assisting RCC in selection of LCCs;
(v) Reviewing new studies that may be of relevance.

**Data Monitoring Committee**
The independent Data Monitoring Committee is responsible for:

(i) Reviewing unblinded interim analyses according to the Protocol;
(ii) Advising the Steering Committee if, in their view, the randomised data provide evidence that may warrant a change in the Protocol (e.g. modification or cessation of one or more of the treatment arms).

**Central Coordinating Office (CCO)**
The CCO is responsible for the overall coordination of the Study, including:

(i) Study planning and organisation of Steering Committee meetings;
(ii) Ensuring necessary regulatory and ethics committee approvals;
(iii) Development of Standard Operating Procedures and computer systems
(iv) Monitoring overall progress of the study;
(v) Provision of study materials to RCCs/LCCs;
(vi) Monitoring and reporting safety information in line with the Protocol and regulatory requirements;
(vii) Dealing with technical, medical and administrative queries from LCCs.

RECOVERY
Randomised Evaluation of COVID-19 Therapy

**Regional Coordinating Centre (RCC)**
The RCCs are responsible for:

(i) Ensuring necessary regulatory and ethics committee approvals;
(ii) Provision of study materials to LCCs;
(iii) Dealing with technical, medical and administrative queries from LCCs.

**Local Clinical Centres (LCC)**
The LCC lead investigator and LCC clinic staff are responsible for:

(i) Obtaining all relevant local permissions (assisted by the CCO);
(ii) All trial activities at the LCC, including appropriate training and supervision for clinical staff;
(iii) Conducting trial procedures at the LCC in line with all relevant local policies and procedures;
(iv) Dealing with enquiries from participants and others.

#### STEERING COMMITTEE

(Major organisational and policy decisions, and scientific advice; blinded to treatment allocation)

| | |
|---|---|
| Chief Investigator | Peter Horby |
| Deputy Chief Investigator | Martin Landray |
| Clinical Trial Unit Lead | Richard Haynes |
| Co-investigators | Kenneth Baillie (Scotland Lead), Maya Buch, Saul Faust, Thomas Jaki, Katie Jeffery, Edmund Juszczak, Marian Knight, Wei Shen Lim, Marion Mafham, Alan Montgomery, Aparna Mukherjee, Andrew Mumford, Kathy Rowan, Guy Thwaites, Jeremy Day |

#### International Committees

*Asia*

| | |
|---|---|
| Chair | Do Van Dung |
| Regional Lead Investigators | Guy Thwaites, Jeremy Day |
| Independent members: | Vietnam : Nguyen Ngo Quang, Prof. Binh<br>Indonesia: Erlina Burhan, Bachti Alisjahbana<br>Nepal: Janak Koirala, Sudha Basnet |
| Other members: | Evelyne Kestelyn, Buddha Basnyat, Pradip Gyanwali, Raph Hamers, Peter Horby |

*Africa*

| | |
|---|---|
| Chair | TBC |
| Independent members: | Ghana: TBD<br>South Africa: TBD |
| Other members: | John Amuasi, Peter Horby, Jeremy Nel |

#### DATA MONITORING COMMITTEE

(Interim analyses and response to specific concerns)

| | |
|---|---|
| Chair | Peter Sandercock |
| Members | Janet Darbyshire, David DeMets, Robert Fowler, David Lalloo, Mohammed Munavvar, Adilia Warris, Janet Wittes |
| Statisticians (non-voting) | Jonathan Emberson, Natalie Staplin |

### 10 CONTACT DETAILS

Website: [www.recoverytrial.net](http://www.recoverytrial.net)
(copies of this protocol and related forms and information can be downloaded)

**RECOVERY Central Coordinating Office:**
Richard Doll Building, Old Road Campus, Roosevelt Drive, Oxford OX3 7LF
United Kingdom

**RECOVERY Vietnam:**
Oxford University Clinical Research Unit, Centre for Tropical Medicine, 764 Vo Van Kiet, District 5, Ho Chi Minh City, Vietnam


**RECOVERY Indonesia:**
Eijkman Oxford Clinical Research Unit (EOCRU), Eijkman Institute for Molecular Biology
Jl. P. Diponegoro No. 69, Jakarta-Indonesia 10430

**RECOVERY Nepal:**
Clinical Trial Unit, Oxford University Clinical Research Unit-Nepal, Patan Academy of Health Sciences, Kathmandu, Nepal

**RECOVERY Ghana:**
Kumasi Center for Collaborative Research in Tropical Medicine
KNUST, Southend Asuogya Road, GPS: AK-312-1059, Kumasi, Ghana

**RECOVERY South Africa:**
Wits Health Consortium, 31 Princess of Wales Terrace, Parktown, Johannesburg, South Africa

**RECOVERY Sri Lanka & Pakistan:**
National Intensive Care Surveillance - M.O.R.U, 2nd Floor, YMBA Building, Borella, Colombo 08, Sri Lanka

**RECOVERY India:**
Indian Council of Medical Research, Division of Epidemiology and Communicable Diseases, Ramalingaswami Bhavan, Ansari Nagar, ICMR-110029

#### To RANDOMISE a patient, visit:

RECOVERY
Randomised Evaluation of COVID-19 Therapy

Website: [www.recoverytrial.net](http://www.recoverytrial.net)

### Appendix 2: RECOVERY Trial Statistical Analysis Plan V5

### Statistical Analysis Plan

**Version 5.0**

**Date: 25 June 2024**

Aligned with protocol version: 27.0, 13 September 2023

IRAS no: 281712
REC ref: EE/20/0101
ISRCTN: 50189673
EudraCT: 2020-001113-21

Nuffield Department of
POPULATION HEALTH

UNIVERSITY OF OXFORD

#### Abbreviations

| | |
|---|---|
| ADaM | Analysis Data Model |
| AE | Adverse event |
| CAP | Community-acquired pneumonia |
| CDISC | The Clinical Data Interchange Standards Consortium |
| CI | Confidence interval |
| COVID | Coronavirus-induced disease |
| CPAP | Continuous Positive Airway Pressure |
| CRP | C-reactive protein |
| DMC | Data Monitoring Committee |
| ECMO | Extra Corporeal Membrane Oxygenation |
| eCRF | Electronic case report form |
| ICD | International Classification of Diseases |
| ICNARC | Intensive Care National Audit and Research Centre |
| ITT | Intention to treat |
| MedDRA | Medical Dictionary for Regulatory Activities |
| OPCS-4 | National Health Service OPCS Classification of Interventions and Procedures version 4 |
| SARS | Severe acute respiratory syndrome |
| SARS-CoV-2 | Severe acute respiratory syndrome coronavirus 2 |
| $S/F_{94}$ ratio | Ratio of peripheral oxygen saturation to fractional inspired oxygen concentration when peripheral oxygen saturation at or below 94% |
| SSAR | Suspected serious adverse reaction |
| SUSAR | Suspected unexpected serious adverse reaction |
| TSC | Trial Steering Committee |

#### List of authors and reviewers (up to and including SAP version 1.1)

##### *Authors*

Dr Louise Linsell, Lead Trial Statistician, Nuffield Department of Population Health (NDPH), University of Oxford

Jennifer Bell, Trial Statistician, NDPH, University of Oxford

##### *Reviewers*

Professor Jonathan Emberson, Data Monitoring Committee (DMC) Statistician, NDPH, University of Oxford (prior to unblinded interim analysis of trial outcomes)

Professor Richard Haynes, Clinical Coordinator, NDPH, University of Oxford

Professor Peter Horby, Chief Investigator (CI), Nuffield Department of Medicine, University of Oxford

Professor Thomas Jaki, TSC Member, Department of Mathematics and Statistics, Lancaster University

Associate Professor Edmund Juszczak, TSC Member, NDPH, University of Oxford (until 6 July 2020)

Professor Martin Landray, Deputy CI, NDPH, University of Oxford

Professor Alan Montgomery, TSC Member, Nottingham Clinical Trials Unit, University of Nottingham

Dr Natalie Staplin, DMC Statistician, NDPH, University of Oxford (prior to unblinded interim analysis of trial outcomes)

#### List of authors and reviewers (version 2.0 onwards)

Professor Edmund Juszczak, TSC Member (University of Nottingham from 6 July 2020)

Professor Alan Montgomery (University of Nottingham), TSC Member

Professor Thomas Jaki (University of Cambridge) co-investigator and TSC Member

Enti Spata, Trial Statistician, NDPH, University of Oxford (until 4 March 2022)

Professor Richard Haynes, Clinical Coordinator, NDPH, University of Oxford

Professor Martin Landray, Deputy CI, NDPH, University of Oxford

Professor Peter Horby, CI, Nuffield Department of Medicine, University of Oxford

Leon Peto, Clinical Coordinator, NDPH, University of Oxford

#### Roles and responsibilities

##### *Trial Statisticians*

*Until 30 September 2020:* Dr Louise Linsell and Jennifer Bell (NDPH, University of Oxford)

Role: To develop the statistical analysis plan (blinded to trial allocation) and conduct the final comparative analyses for Lopinavir-Ritonavir¸ Corticosteroid (dexamethasone) and Hydroxychloroquine (main randomisation part A).

*From 1 October 2020 until 4 March 2022:* Enti Spata (NDPH, University of Oxford)

Role: To develop the statistical analysis plan (blinded to trial allocation) and conduct the final comparative analyses for all other treatment arms in this period.

*From 1 April 2022:* Professor Jonathan Emberson and Dr Natalie Staplin (NDPH, University of Oxford)

Role: To develop the statistical analysis plan (only for those aspects to which they are currently blinded) and to conduct the final comparative analyses on completion.

##### *Data Monitoring Committee (DMC) Statistician (non-voting)*

Professor Jonathan Emberson (NDPH, University of Oxford)

*Until 13 March 2024:* Natalie Staplin (NDPH, University of Oxford)

Role: To conduct regular interim analyses for the DMC. Other contributions are restricted until completion of each comparison.

##### *Statisticians on the Trial Steering Committee (TSC)*

Professor Thomas Jaki (University of Cambridge)

*From 13 March 2024:* Natalie Staplin (NDPH, University of Oxford)

*Until 25 June 2024:* Professor Alan Montgomery (University of Nottingham) and Professor Edmund Juszczak (University of Nottingham)

Role: Major organisational and policy decisions, and scientific advice; blinded to treatment allocation.

##### *Trial IT systems & Programmers*

Andy King, David Murray, Richard Welsh (NDPH, University of Oxford)

Role: To generate and prepare reports monitoring the randomisation schedule. To supply data snapshots for interim and final analysis. Responsibility for randomisation system, clinical databases and related activities.

Bob Goodenough (NDPH, University of Oxford)

Role: Validation of IT systems

Dr Will Stevens, Karl Wallendszus (NDPH, University of Oxford)

Role: To produce analysis-ready datasets according to CDISC standards.

### 1 INTRODUCTION

This document details the proposed presentation and analysis for the main paper(s) reporting results from the multicentre randomised controlled trial RECOVERY (ISRCTN50189673) to investigate multiple treatments on major outcomes in patients hospitalised with COVID-19, influenza, or community-acquired pneumonia related to other pathogens.

The results reported in these papers will follow the strategy set out here, which adheres to the guidelines for the content of a statistical analysis plan (SAP).[1] Any subsequent analyses of a more exploratory nature will not be bound by this strategy.

Suggestions for subsequent analyses by oversight committees, journal editors or referees, will be considered carefully in line with the principles of this analysis plan.

Any deviations from the statistical analysis plan will be described and justified in the final report. The analysis will be carried out by identified, appropriately qualified and experienced statisticians, who will ensure the integrity of the data during their processing.

This SAP is based on multiple versions of the protocol. All regulatory documents can be found in the RECOVERY trial directory: https://www.recoverytrial.net/for-site-staff/site-set-up-1/regulatory-documents.

SAP versions 1.0 & 1.1 applied to the first three principal comparisons (hydroxychloroquine, dexamethasone, and lopinavir-ritonavir versus no additional treatment respectively), for which data matured in the first UK wave of the pandemic. However, due to its later introduction, enrolment of patients in the azithromycin arm was much slower. Over time, factorial randomisations and a second randomisation have been added, introducing new treatment arms including convalescent plasma, tocilizumab, synthetic neutralizing antibodies, and aspirin. Version 2.0 of the SAP was produced in response to these changes, combined with the fact that use of corticosteroids (one of the original treatment arms) is now the usual standard of care for many patients. SAP version 3.0 included revisions for REGEN-COV2 (casirivimab+imdevimab), early phase assessments, and 6 month follow-up. SAP version 4.0 included additional COVID-19 comparisons (sotrovimab, molnupiravir, and Paxlovid), introduced influenza comparisons (baloxavir, oseltamivir, and corticosteroids), added virology outcomes, and modified 6 month follow-up analyses.

SAP version 5.0 introduces a new corticosteroid comparison for patients with community-acquired pneumonia (CAP) not related to SARS-CoV-2 or influenza, updates the recruitment status of existing comparisons, and finalises the analysis plan for SARS-CoV-2 antivirals (including monoclonal antibodies).

### 2 BACKGROUND INFORMATION

#### 2.1 Rationale

In early 2020, as the protocol was being developed, there were no approved treatments for COVID-19. The aim of the trial was to provide reliable evidence on the efficacy of candidate therapies (including re-purposed and novel drugs) for suspected or confirmed COVID-19 infection on major outcomes in hospitalised adult patients receiving standard care.

In late 2021, the protocol was extended to include evaluation of potential treatments for influenza occurring in isolation or in combination with COVID-19 (protocol version 19 onwards) and this was approved by MHRA and the ethics committee. However, at the request of the RECOVERY funder, introduction of the influenza comparisons was delayed. These opened at a small number of sites in early 2023, and more widely during winter 2023/24. The CAP comparison was approved in November 2023 and introduced in January 2024. At the time of finalising the current version of the SAP (version 5.0), the enrolment of patients with COVID-19 has closed (on 31 March 2024) but the trial remains open to patients with either pneumonia due to influenza or CAP

#### 2.2 Objectives of the trial

##### 2.2.1 *Primary and secondary objectives for COVID-19 and CAP comparisons*

The primary objective is to provide reliable estimates of the effect of study treatments on all-cause mortality within 28 days of the relevant randomisation. The secondary objectives are to investigate the effect of study treatments on the duration of hospital stay and on the combined endpoint of use of invasive mechanical ventilation (including Extra Corporal Membrane Oxygenation [ECMO]) or death.

##### 2.2.2 *Primary and secondary objectives for influenza comparisons*

The co-primary objectives are to provide reliable estimates of the effect of study treatments on (a) all-cause mortality within 28 days of the relevant randomisation and (b) the duration of hospital stay. The secondary objective is to investigate the effect of study treatments on the combined endpoint of use of invasive mechanical ventilation (including Extra Corporal Membrane Oxygenation [ECMO]) or death.

#### 2.3 Trial design

This is a multi-centre, multi-arm, adaptive, open label, randomised controlled trial with three possible stages of randomisation, as described below. The trial is designed with streamlined processes in order to facilitate rapid large-scale recruitment with minimal data collection.

#### 2.4 Eligibility

##### 2.4.1 *Inclusion criteria*

Patients are eligible for the trial if all of the following are true:

- Hospitalised
- Pneumonia syndrome
- One of the following diagnoses:
  - Confirmed SARS-CoV-2 infection (closed on 31 March 2024)
  - Confirmed influenza A or B infection
  - Community-acquired pneumonia with planned antibiotic treatment (without suspected or confirmed SARS-COV-2, influenza, pulmonary tuberculosis or *Pneumocystis jirovecii* infection)
- No medical history that might, in the opinion of the attending clinician, put the patient at significant risk if they were to participate in the trial.

##### 2.4.2 *Exclusion criteria*

If one or more of the active drug treatments is not available at the hospital or is believed, by the attending clinician, to be contraindicated (or definitely indicated) for the specific patient, then this fact will be recorded via the web-based form prior to randomisation; random allocation will then be between the remaining arms. Details of specific contraindications and cautions for each treatment are listed in Appendix 2 of the protocol.

##### 2.4.3 *Randomisation on more than one occasion*

From protocol version 21.1, patients who have been previously recruited into RECOVERY are eligible to be recruited again as long as their previous randomisation was >6 months ago. Patients will not be recruited into the same randomised comparison on more than one occasion, regardless of the time interval.

#### 2.5 Treatments

All patients will receive standard management for the participating hospital. The main randomisation will be between the following treatment arms (although not all arms may be available at any one time). The doses listed are for adults; paediatric dosing is described in the protocol.

#### *COVID-19 Comparisons*

##### 2.5.1 *Main randomisation part A (enrolment closed):*

- **No additional treatment**
- **Lopinavir 400mg-Ritonavir 100mg** by mouth (or nasogastric tube) every 12 hours for 10 days. [Introduced in protocol version 1.0; **enrolment closed** 29 June 2020]
- **Corticosteroid** in the form of dexamethasone**,** administered as an oral liquid or intravenous preparation 6 mg once daily for 10 days. In pregnancy, prednisolone 40

mg administered by mouth (or intravenous hydrocortisone 80 mg twice daily) should be used instead. [Introduced in protocol version 1.0; **enrolment closed to adults** 8 June 2020]

- **Hydroxychloroquine** by mouth for 10 days (4 doses in first 24 hours and 1 dose every 12 hours for 9 days). [Introduced in protocol version 2.0; **enrolment closed** 5 June 2020]
- **Azithromycin 500mg** by mouth (or nasogastric tube) or intravenously once daily for a total of 10 days. [Introduced in protocol version 3.0; **enrolment closed** 27 November 2020]
- **Colchicine** by mouth for 10 days (1.5 mg in first 12 hours then 0.5 mg twice daily). [Introduced in protocol version 12.0; **enrolment closed** 5 March 2021.]
- **Dimethyl fumarate** 120 mg every 12 hours for 4 doses followed by 240 mg every 12 hours by mouth for 8 days (10 days in total). [Introduced in protocol version 14.0 as Early Phase Assessment; **enrolment closed** 19 November 2021.]

##### 2.5.2 *Main randomisation part B (enrolment closed):*

In a factorial design, eligible patients may be randomised to the arms below. The doses listed are for adults; paediatric dosing is described in the protocol.

- **No additional treatment**
- **Convalescent plasma** Single unit of ABO compatible convalescent plasma (275ml ± 75ml) intravenous per day on study days 1 (as soon as possible after randomisation) and 2 (with a minimum of 12-hour interval between 1st and 2nd units). ABO identical plasma is preferred if available. The second transfusion should not be given if patient has a suspected serious adverse reaction during or after the first transfusion. [Introduced in protocol version 6.0; **enrolment closed** 15 January 2021]
- **Synthetic neutralising antibodies** (REGEN-COV2; adults and children aged ≥12 years only - children who weigh <40kg will also not be eligible for this treatment). A single dose of REGN10933 + REGN10987 8 g (4 g of each monoclonal antibody) in 250ml 0.9% saline infused intravenously over 60 minutes ± 15 minutes as soon as possible after randomisation. [Introduced in protocol version 9.1; **enrolment closed** 22 May 2021]

##### 2.5.3 *Main randomisation part C (enrolment closed):*

In a factorial design, eligible patients may be randomised to the arms below. The dose listed is for adults; children are excluded from this comparison.

- **No additional treatment**
- **Aspirin** 150 mg by mouth (or nasogastric tube) or per rectum once daily until discharge. [Introduced in protocol version 10.1; **enrolment closed** 21 March 2021]

##### 2.5.4 *Main randomisation part D (enrolment closed):*

In a factorial design, eligible patients may be randomised to the arms below. The dose listed is for adults; children <2 years old or with PIMS-TS are excluded from this comparison.

- **No additional treatment**
- **Baricitinib** 4 mg by mouth (or nasogastric tube) once daily for 10 days. [Introduced in protocol version 13.0; **enrolment closed** 29 December 2021]

##### 2.5.5 *Main randomisation part E (enrolment closed):*

In a factorial design, eligible patients may be randomised to the arms below. The dose listed is for adults; children <18 years old are excluded from this comparison.

- **No additional treatment**
- **High-dose corticosteroids** dexamethasone 20 mg once daily for 5 days, followed by dexamethasone 10 mg once daily for 5 days. [Introduced outside UK in protocol version 13.0 and within UK in protocol version 20.0; eligibility criteria modified in protocol version 25.0; **enrolment closed** on 31 March 2024]

On 11 May 2022, the RECOVERY DMC advised, "*For patients being considered for treatment with high dose dexamethasone, we recommend stopping recruitment of patients who require no oxygen or simple oxygen only at the time of randomisation due to safety concerns. Follow-up of these patients should continue. However, we encourage continuing recruitment and follow-up of all those patients who, at randomisation, require either non-invasive ventilation, invasive mechanical ventilation or ECMO.*" Consequently, on 13 May 2022, recruitment to this comparison was closed for patients on no oxygen or simple oxygen only. The protocol (version 25.0) was updated accordingly. The comparison closed for all patients on 31 March 2024.

##### 2.5.6 *Main randomisation part F (enrolment closed):*

In a factorial design, eligible patients (with or without influenza infection) may be randomised to the arms below. The dose listed is for adults; children <18 years old are excluded from this comparison.

- **No additional treatment**
- **Empagliflozin** 10 mg once daily for 28 days. [Introduced in protocol version 16.1; **enrolment closed** 7 March 2023]

##### 2.5.7 *Main randomisation part J (enrolment closed):*

In a factorial design, eligible patients (with or without influenza infection) may be randomised to the arms below. The dose listed is for adults; children <12 years old are excluded from this comparison.

- **No additional treatment**

- **Sotrovimab 1000 mg once** as soon as possible after randomisation. [Introduced in protocol version 21.1; **enrolment closed** 31 March 2024]

##### 2.5.8 *Main randomisation part K (enrolment closed):*

In a factorial design, eligible patients (with or without influenza infection) may be randomised to the arms below. The dose listed is for adults; children <18 years old are excluded from this comparison.

- **No additional treatment**
- **Molnupiravir 800 mg twice daily** for 5 days by mouth. [Introduced in protocol version 21.1; **enrolment closed** 24 May 2023]

##### 2.5.9 *Main randomisation part L (enrolment closed):*

In a factorial design, eligible patients (with or without influenza infection) may be randomised to the arms below. The dose listed is for adults; children <18 years old are excluded from this comparison.

- **No additional treatment**
- **Paxlovid (nirmatrelvir/ritonavir) 300/100 mg twice daily** for 5 days by mouth. [Introduced in protocol version 23.1; **enrolment closed** 24 May 2023]

##### 2.5.10 *Second randomisation for adults with progressive COVID-19 (enrolment closed)*

Patients enrolled in the main RECOVERY trial and with clinical evidence of a hyper-inflammatory state may be considered for a second randomisation if they meet the following criteria:

- Randomised into the main RECOVERY trial no more than 21 days ago
- Clinical evidence of progressive COVID-19:
    - oxygen saturation <92% on room air or requiring oxygen; and
    - C-reactive protein (CRP) ≥75 mg/L
- No medical history that might, in the opinion of the attending clinician, put the patient at significant risk if they were to participate in this aspect of the RECOVERY trial

Eligible participants may be randomised between the following treatment arms:

- **No additional treatment**
- **Tocilizumab** by intravenous infusion with the dose determined by body weight. [Introduced in protocol version 4.0; **enrolment closed** 24 January 2021]

#### *Influenza Comparisons*

##### 2.5.11 *Main randomisation part G:*

In a factorial design, eligible patients (with or without SARS-CoV-2 co-infection) may be randomised to the arms below. The dose listed is for adults; children <12 years old are excluded from this comparison.

- **No additional treatment**
- **Baloxavir marboxil 40mg (or 80mg if weight ≥80kg) once daily** by mouth or nasogastric tube to be given on day 1 and day 4. [Introduced in protocol version 19.1; **enrolment ongoing**]

##### 2.5.12 *Main randomisation part H:*

In a factorial design, eligible patients (with or without SARS-CoV-2 co-infection) may be randomised to the arms below.

- **No additional treatment**
- **Oseltamivir 75mg twice daily** by mouth or nasogastric tube for five days (If participant is discharged before course is complete, the participant should be provided with medication to complete the course at home. Course can be extended to 10 days for immunosuppressed patients at the managing clinician's discretion). [Introduced in protocol version 19.1; **enrolment ongoing**]

##### 2.5.13 *Main randomisation part I:*

In a factorial design, eligible patients (without suspected or confirmed SARS-CoV-2 infection and with clinical evidence of hypoxia) may be randomised to the arms below.

- **No additional treatment**
- Corticosteroids: **Dexamethasone 6mg once daily given** orally or intravenously for ten days or until discharge (whichever happens earliest). [Introduced in protocol version 19.1; **enrolment ongoing**]

#### *Community-Acquired Pneumonia (CAP) Comparison*

##### 2.5.14 *Main randomisation part M:*

In a factorial design, eligible patients (with planned antibiotic treatment, and without suspected or confirmed SARS-CoV-2, influenza, pulmonary tuberculosis, or *Pneumocystis jirovecii* infection) may be randomised to the arms below.

- **No additional treatment**
- Corticosteroids: **Dexamethasone 6mg once daily given** orally or intravenously for ten days or until discharge (whichever happens earliest). [Introduced in protocol version 27.0; **enrolment ongoing**]

#### 2.6 Definitions of outcomes

Outcomes will be assessed at 28 days after the relevant randomisation. (Analyses of 6 month are described in section 10.)

##### 2.6.1 *Primary outcome*

For COVID-19 and CAP comparisons: Mortality (all-cause)
For influenza comparisons: Co-primary outcomes of Mortality (all-cause) and Time to discharge alive from hospital

##### 2.6.2 *Secondary clinical outcomes*

- Time to discharge alive from hospital (for COVID-19 and CAP comparisons)
- Use of invasive mechanical ventilation (including Extra Corporal Membrane Oxygenation [ECMO]) or death (among patients not on invasive mechanical ventilation or ECMO at time of randomisation)

##### 2.6.3 *Subsidiary clinical outcomes*

- Use of ventilation (overall and by type) among patients not on ventilation (of any type) at time of randomisation
- Duration of invasive mechanical ventilation among patients on invasive mechanical ventilation at time of randomisation (defined as time to successful cessation of invasive mechanical ventilation: see section 5.1.2.2)
- Use of renal dialysis or haemofiltration (among patients not on renal dialysis or haemofiltration at time of randomisation)
- Thrombotic events (overall and by type; introduced in Protocol version 10.1)

##### 2.6.4 *Virological outcomes*

- SARS-CoV-2 and influenza RNA levels in the nasopharynx (parts G, H, I, J, K and L only)
- SARS-CoV-2 and influenza viral resistance markers (parts J, K and L, and parts G and H, respectively)

##### 2.6.5 *Safety outcomes*

- Cause-specific mortality (COVID-19, influenza, CAP, other infection, cardiac, stroke, other vascular, cancer, other medical, external, unknown cause)
- Major cardiac arrhythmia (recorded on follow-up forms completed from 12 May 2020 onwards)
- Major bleeding (overall and by type; introduced in Protocol version 10.1)
- Early safety of antibody-based therapy (sudden worsening in respiratory status; severe allergic reaction; temperature >39°C or ≥2°C rise since randomisation; sudden hypotension; clinical haemolysis; and thrombotic events within the first 72 hours; (Main randomisation part B only)
- Non-coronavirus infection (overall and by site and putative organism [virus, bacteria, fungus, other]; introduced in Protocol version 14.0)
- Metabolic, kidney and liver complications:
    - severe hyperglycaemia: overall and by type (ketoacidosis, hyperosmolar hyperglycaemic state, hyperglycaemia requiring new use of insulin)
    - severe hypoglycaemia

  - acute kidney injury (ratio of post-randomisation peak creatinine to baseline value >1.5 or new use of renal dialysis/haemofiltration; introduced in protocol V16.1)
  - liver dysfunction: peak alanine (or aspartate) transaminase and possible liver injury (defined as ALT >3x ULN plus bilirubin >2x ULN; introduced in protocol V23.1).
- Seizures (introduced in protocol V23.1)
- Infusion reactions to sotrovimab (Main randomisation part J only; introduced in protocol V21.1)

###### 2.6.6 *Detailed derivation of outcomes*

The detailed derivation of outcomes included in statistical analysis is described separately in a data derivation document and included in the Study Data Reviewer's Guide.

##### 2.7 Hypothesis framework

For each of the primary, secondary and subsidiary outcomes, the null hypothesis will be that there is no true difference in effect between any of the treatment arms.

##### 2.8 Sample size

The larger the number randomised, the more accurate the results will be, but the numbers that can be randomised will depend critically on how large the epidemic becomes. If substantial numbers are hospitalised in the participating centres then it may be possible to randomise several thousand with moderate disease and a few thousand with severe disease. Some indicative sample sizes and projected recruitment will be estimated using emerging data for several different scenarios.

The TSC will monitor recruitment and primary event rate (in active and control arms combined, i.e. blind to knowledge of the unblinded results) for ongoing comparisons. In general, the TSC will continue recruitment until such time as there are sufficient patients enrolled in the comparison to provide at least 90% power at 2P=0.01 to detect a clinically relevant proportional reduction (typically one-fifth) in the primary outcome.

##### 2.9 Randomisation

Eligible patients will be randomised using a 24/7 secure central web-based randomisation system, developed and hosted within NDPH, University of Oxford. Users of the system will have no insight into the next allocation, given that simple randomisation is being used. If a patient is randomised inadvertently more than once during the same hospital admission, the first allocation will be used.

The implementation of the randomisation procedure will be monitored by the Senior Trials Programmer, and the TSC notified if an error in the randomisation process is identified.

##### *COVID-19 treatment comparisons*

###### 2.9.1 *Main randomisation part A (enrolment closed)*

Simple randomisation will be used to allocate participants to one of the following treatment arms (in addition to usual care), which is subject to change:

- No additional treatment
- Lopinavir-Ritonavir [Introduced in protocol version 1.0; **enrolment closed** 29 June 2020]
- Corticosteroid [Introduced in protocol version 1.0; **enrolment closed to adults** 8 June 2020]
- Hydroxychloroquine [Introduced in protocol version 2.0; **enrolment closed** 5 June 2020]
- Azithromycin [Introduced in protocol version 3.0; **enrolment closed** 27 November 2020]
- Colchicine [Introduced in protocol version 11.1; **enrolment closed** 5 March 2021]
- Dimethyl fumarate [Introduced in protocol version 14.0; **enrolment closed** 19 November 2021]

The randomisation programme will allocated patients in a ratio of 2:1 between the no additional treatment arm and each of the other arms that are not contra-indicated and are available when multiple arms were included in the protocol. Hence if all 4 active treatment arms are available, then the randomisation will be in the ratio 2:1:1:1:1. If one or more of the active drug treatments is not available at the hospital or is believed, by the attending clinician, to be contraindicated (or definitely indicated) for the specific patient, then this fact will be recorded via the web-based form prior to randomisation; random allocation will then be between the remaining arms (in a 2:1:1:1, 2:1:1 or 2:1 ratio). Since the closure of the azithromycin comparison, all comparisons in part A have used a 1:1 ratio.

##### 2.9.2 *Main randomisation part B (enrolment closed)*

In a factorial design, eligible patients will be randomised simultaneously using simple randomisation with allocation ratio 1:1:1 to one of the following arms, which is subject to change:

- No additional treatment
- Convalescent plasma [Introduced in protocol version 6.0; **enrolment closed** 15 January 2021]
- Synthetic neutralising antibodies [Introduced in protocol version 9.1; **enrolment closed** 22 May 2021]

If the active treatment is not available at the hospital, the patient does not consent to receive convalescent plasma, or is believed, by the attending clinician, to be contraindicated for the specific patient, then this fact will be recorded via the web-based form and the patient will be excluded from the relevant arm in Randomisation part B.

##### 2.9.3 *Main randomisation part C (enrolment closed)*

In a factorial design, eligible patients will be randomised simultaneously using simple randomisation with allocation ratio 1:1 to one of the following arms, which is subject to change:

- No additional treatment

- Aspirin [Introduced in protocol version 10.1; **enrolment closed** 21 March 2021]

##### 2.9.4 *Main randomisation part D (enrolment closed)*

In a factorial design, eligible patients will be randomised simultaneously using simple randomisation with allocation ratio 1:1 to one of the following arms, which is subject to change:

- No additional treatment
- Baricitinib [Introduced in protocol version 13.0; **enrolment closed** 29 December 2021]

##### 2.9.5 *Main randomisation part E (enrolment closed)*

In a factorial design, eligible patients will be randomised simultaneously using simple randomisation with allocation ratio 1:1 to one of the following arms, which is subject to change:

- No additional treatment
- High-dose corticosteroids [Introduced outside UK in protocol version 13.0 and within UK in protocol version 20.0; eligibility criteria modified in protocol version 25.0; **enrolment closed** 31 March 2024]

##### 2.9.6 *Main randomisation part F (enrolment closed)*

In a factorial design, eligible patients will be randomised simultaneously using simple randomisation with allocation ratio 1:1 to one of the following arms, which is subject to change:

- No additional treatment
- Empagliflozin [Introduced in protocol version 16.1; **enrolment closed** 7 March 2023]

Note: From protocol version 7.0 onwards, randomisation is permitted in part B of main randomisation without randomisation in part A. From protocol version 10.1 onwards, randomisation is permitted in any combination of parts (A, B, C, etc).

##### 2.9.7 *Main randomisation part J (enrolment closed)*

In a factorial design, eligible patients will be randomised simultaneously using simple randomisation with allocation ratio 1:1 to one of the following arms:

- No additional treatment
- Sotrovimab [Introduced in protocol version 21.1; **enrolment closed** 31 March 2024]

##### 2.9.8 *Main randomisation part K (enrolment closed)*

In a factorial design, eligible patients will be randomised simultaneously using simple randomisation with allocation ratio 1:1 to one of the following arms, which is subject to change:

- No additional treatment
- Molnupiravir [Introduced in protocol version 21.1; **enrolment closed** 24 May 2023]

##### 2.9.9 *Main randomisation part L (enrolment closed)*

In a factorial design, eligible patients will be randomised simultaneously using simple randomisation with allocation ratio 1:1 to one of the following arms, which is subject to change:

- No additional treatment
- Paxlovid [Introduced in protocol version 23.1; **enrolment closed** 24 May 2023]

###### 2.9.10 *Second randomisation for adults with progressive COVID-19 (enrolment closed)*

Eligible participants will be randomised using simple randomisation with an allocation ratio 1:1 between the following arms, which is subject to change:

- No additional treatment
- Tocilizumab [Introduced in protocol version 4.0; **enrolment closed** 24 January 2021]

##### *Influenza treatment comparisons*

###### 2.9.11 *Main randomisation part G*

In a factorial design, eligible patients will be randomised simultaneously using simple randomisation with allocation ratio 1:1 to one of the following arms:

- No additional treatment
- Baloxavir [Introduced in protocol version 19.1; **enrolment ongoing**]

###### 2.9.12 *Main randomisation part H*

In a factorial design, eligible patients will be randomised simultaneously using simple randomisation with allocation ratio 1:1 to one of the following arms, which is subject to change:

- No additional treatment
- Oseltamivir [Introduced in protocol version 19.1; **enrolment ongoing**]

###### 2.9.13 *Main randomisation part I*

In a factorial design, eligible patients will be randomised simultaneously using simple randomisation with allocation ratio 1:1 to one of the following arms, which is subject to change:

- No additional treatment
- Corticosteroids [Introduced in protocol version 19.1; **enrolment ongoing**]

##### *CAP treatment comparisons*

###### 2.9.14 *Main randomisation part M*

In a factorial design, eligible patients will be randomised simultaneously using simple randomisation with allocation ratio 1:1 to one of the following arms, which is subject to change:

- No additional treatment

- Corticosteroids [Introduced in protocol version 27.0; **enrolment ongoing**]

#### 2.10 Blinding

This is an open-label study. However, while the study is in progress, access to tabular results of study outcomes by treatment allocation will not be available to the research team, CIs, trial statisticians, clinical teams, or members of the TSC (unless the DMC advises otherwise). The DMC and DMC statisticians will be unblinded.

#### 2.11 Data collection schedule

Baseline and 28-day outcome information will be collected on trial-specific electronic case report forms (eCRFs) and entered into a web-based IT system by a member of the hospital or research staff. Follow-up information will be collected on all study participants, irrespective of whether they complete the scheduled course of allocated study treatment. Study staff will seek follow-up information through various means, including routine healthcare systems and registries.

All randomised participants will be followed up until death or 6 months post-randomisation to the main trial (whichever is sooner). NHS England and equivalent organisations in the devolved nations will supply data fields relevant to trial baseline and outcome measures to NDPH, University of Oxford on a regular basis, for participants enrolled into the trial. This will be combined with the trial-specific data collected via the web-based IT system and adjudicated internally.

Longer term (up to 10 years) follow-up of UK participants only will be sought through linkage to electronic healthcare records and medical databases including those held by NHS England, Public Health England and equivalent bodies, and to relevant research databases (e.g. UK Biobank, Genomics England).

#### 2.12 Data monitoring

During the study all study data will be supplied in strict confidence to the independent DMC for independent assessment and evaluation. The DMC will request such analyses at a frequency relevant to the emerging data from this and other studies.

The DMC has been requested to determine if, in their view, the randomised comparisons in the study have provided evidence on mortality that is strong enough (with a range of uncertainty around the results that is narrow enough) to affect national and global treatment strategies. In such a circumstance, the DMC will inform the TSC who will make the results available to the public and amend the trial arms accordingly.

The Data Monitoring Committee has determined that, in general, to consider recommending stopping a treatment early for benefit would require at least a 3 to 3·5 standard error reduction in mortality. Examinations of the data at every 10% (or even 5%) of the total data would lead to only a marginal increase in the overall type I error rate. Hence, multiple reviews by the Data Monitoring Committee have no material impact on the final analysis.

Further details about the role and membership of the independent Data Monitoring Committee are provided in the protocol.

##### 2.13 Trial reporting

The trial will be reported according to the principles of the CONSORT statements.[2, 3, 4]

#### 3 ANALYSIS POPULATIONS

##### 3.1 Population definitions

The intention to treat (ITT) population will be all participants randomised, irrespective of treatment received. This ITT population will be used for analysis of efficacy and safety data. For interim analyses, baseline data will be reported for all participants with data available and outcome data will be reported for all participants who have died, been discharged from hospital, or reached day 28 after the first randomisation.

#### 4 DESCRIPTIVE ANALYSES

##### 4.1 Participant throughput

The flow of participants through the trial will be summarised for each separate pairwise comparison using a CONSORT diagram. The flow diagram will show the contribution of participants from each of the paths (from each of the parts of the main randomisation and from the second randomisation), where applicable. The flow diagrams will describe the numbers of participants randomly allocated, who received allocation, withdrew consent, and included in the ITT analysis population. The flow diagrams for arms in the main randomisation will also report the number of participants who underwent the second randomisation (where applicable).

##### 4.2 Baseline comparability of randomised groups

The following characteristics will be described separately for patients randomised to each main comparison (for each separate pairwise comparison of active treatment with the no additional treatment arm), and separately for the first and second randomisation.

##### *COVID-19 treatment comparisons*

###### 4.2.1 Main randomisation – COVID-19 comparisons (parts A, B, C, D, E, F, J, K, L)

- Age at randomisation
- Sex
- Ethnicity
- Region (UK, non-UK)
- Time since symptom onset
- Time since hospitalisation
- Current respiratory support (none, oxygen only, non-invasive ventilation, invasive mechanical ventilation [including ECMO])

- Comorbidities (diabetes, heart disease, chronic lung disease, tuberculosis, human immunodeficiency virus, severe liver disease, severe kidney impairment)
- SARS-CoV-2 test result
- If female, known to be pregnant
- Use of systemic corticosteroid (including those allocated to corticosteroid in parts A or I)
- Use of other relevant treatments (e.g. remdesivir, interleukin-6 antagonist, monoclonal anti-SARS-CoV-2 neutralising antibody, baricitinib, molnupiravir, paxlovid)
- Prior SARS-CoV-2 vaccination
- For parts B, J, K and L only, serum anti-SARS-CoV-2 antibody status (anti-S and anti-N)
- For parts J, K and L only, serum SARS-CoV-2 antigen concentration
- For parts J, K and L only, nasal/oropharyngeal SARS-CoV-2 RNA level
- Laboratory markers (introduced in protocol v9.1):
    - C-reactive protein
    - Estimated glomerular filtration rate (calculated using the CKD-EPI formula)
    - D-dimer

##### 4.2.2 Second randomisation

In addition to the above:

- Current respiratory support
- Latest oxygen saturation measurement
- Latest C-reactive protein
- Latest ferritin
- Latest estimated glomerular filtration rate (calculated using the CKD-EPI formula)
- Allocation in main randomisation parts A, B, C, D and E
- Interval between first and second randomisation

The number and percentage will be presented for binary and categorical variables. The mean and standard deviation or the median and the interquartile range will be presented for continuous variables.

#### *Influenza treatment comparisons*

##### 4.2.3 Main randomisation – influenza comparisons (parts G, H, I)

- Age at randomisation
- Sex
- Ethnicity
- Region (UK, EU, Asia, Africa)
- Time since symptoms onset
- Time since hospitalisation
- Current respiratory support (none, oxygen only, non-invasive ventilation, invasive mechanical ventilation [including ECMO])

- Pneumonia severity (CURB-65 score)[a]
- Presence of unilateral/bilateral lung consolidation on imaging
- Comorbidities (diabetes, heart disease, chronic lung disease, tuberculosis, human immunodeficiency virus, severe liver disease, severe kidney impairment)
- Influenza test result
- If female, known to be pregnant
- Use of systemic corticosteroid (including those allocated to corticosteroid in part I)
- Use of other relevant treatments (e.g. oseltamivir, baloxavir)
- Prior influenza vaccination (within the past 12 months)
- Laboratory markers:
  - C-reactive protein
  - Procalcitonin
  - Estimated glomerular filtration rate (calculated using the CKD-EPI formula)

#### *CAP treatment comparisons*

##### 4.2.4 Main randomisation – CAP comparisons (part M)

- Age at randomisation
- Sex
- Ethnicity
- Region (UK, EU, Asia, Africa)
- Time since symptoms onset
- Time since hospitalisation
- Current respiratory support (none, oxygen only, non-invasive ventilation, invasive mechanical ventilation [including ECMO])
- Pneumonia severity (CURB-65 score)[a]
- Presence of unilateral/bilateral lung consolidation on imaging
- Comorbidities (diabetes, heart disease, chronic lung disease, tuberculosis, human immunodeficiency virus, severe liver disease, severe kidney impairment)
- Influenza test result
- If female, known to be pregnant
- Use of systemic corticosteroid (including those allocated to corticosteroid in part I)
- Use of other relevant treatments (e.g. oseltamivir, baloxavir)
- Laboratory markers:
  - C-reactive protein
  - Procalcitonin

[a] The CURB-65 Score (Lim WS et al. Thorax 2003:58:377–82) has been validated for predicting mortality in community-acquired pneumonia and is recommended by the British Thoracic Society. Each risk factor scores one point, for a maximum score of 5:

- Confusion of new onset (defined for this study as presence of new or worsened confusion)
- Blood Urea nitrogen >7 mmol/L (19 mg/dL)
- Respiratory rate of ≥30 breaths per minute
- Blood pressure <90 mmHg systolic or diastolic blood pressure ≤60 mmHg
- Age ≥65 years

- Estimated glomerular filtration rate (calculated using the CKD-EPI formula)

#### 4.3 Completeness of follow-up

All reasonable efforts will be taken to minimise loss to follow-up, which is expected to be minimal as data collection for primary and secondary outcomes using trial-specific eCRFs is combined with linkage to routine clinical data on study outcomes from NHS England, ICNARC, and similar organisations in the devolved nations.

The number and percentage of participants with follow-up information at day 28 after the relevant randomisation will be reported. Data will be shown for each of the following: all-cause mortality, hospital discharge status, ventilation status, and will be shown for each randomised group for the main and second randomisation separately.

#### 4.4 Adherence to treatment

The number and proportion of patients who did not receive the treatment they were allocated to will be reported. If any other trial treatment options were known to be received, instead of or in addition to, the allocated treatment during the 28-day follow-up period after the first randomisation, these will be collected and reported. Details on the number of days (or doses) of treatment received will be reported for all trial treatments received where available.

### 5 COMPARATIVE ANALYSES AT 28 DAYS

For all outcomes, the primary analysis will be performed on the intention to treat (ITT) population at 28 days after randomisation. (Additional details specific to the comparison of REGEN-COV2 vs. usual care [part B] are provided in Appendix I and for the comparison of sotrovimab vs. usual care [part J], molnupiravir vs. usual care [part K], and Paxlovid vs. usual care [part L] are provided in Appendix II.)

Pairwise comparisons will be made between each treatment arm and the no additional treatment arm (reference group) in that particular randomisation (main randomisation part A, main randomisation part B, main randomisation part C, etc.). Since not all treatments may be available or suitable for all patients, those in the no additional treatment arm will only be included in a given comparison if, at the point of their randomisation, they *could* alternatively have been randomised to the active treatment of interest (i.e. the active treatment was available at the time and it was not contra-indicated). The same applies to treatment arms added at a later stage; they will only be compared to those patients recruited concurrently.

#### 5.1 Main randomisation (all parts)

In each component of the factorial design, the main effects of treatments evaluated in a particular part will be presented and tested across all arms in the other main randomisation parts combined, as described in this section. (Assessments of whether the effects of treatments in the part in question vary depending on other randomised treatments are described in section 5.6).

##### 5.1.1 *Primary and secondary outcome*

###### *5.1.1.1 Mortality*

Mortality (all-cause) will be summarised with counts and percentages by randomised comparison group. A time-to-event analysis will be conducted using Cox proportional hazards regression, adjusted for baseline characteristics as described in Section 5.4, to estimate the hazard ratio, 95% confidence interval and corresponding p-value for each treatment group versus the no additional treatment group. Kaplan-Meier estimates for the time to event will also be plotted. For the primary outcome, discharge alive before the relevant time period (28 days after randomisation) will be assumed as absence of the event (unless there is additional data confirming otherwise).

###### *5.1.1.2 Time to discharge alive from hospital*

A time-to-event analysis will be used to compare each treatment group with the no additional treatment group. As described for the primary outcome, the adjusted hazard ratio and its confidence interval will be estimated using Cox proportional hazards regression, adjusted for baseline characteristics as described in Section 5.4, to estimate the hazard ratio, 95% confidence interval and corresponding p-value for each treatment group versus the no additional treatment group. Kaplan-Meier curves will be drawn. Patients who die in hospital will be censored after 28 days after randomisation. This gives an unbiased estimate of the recovery rate and comparable estimates to the competing risks approach in the absence of other censoring (which is expected to be very minimal).[5]

###### *5.1.1.3 Use of invasive mechanical ventilation (including ECMO) or death*

Counts and percentages will be presented by randomised group and a log-binomial regression model, adjusted for baseline characteristics as described in Section 5.4, will be used to estimate the risk ratio, confidence interval and p-value for each pairwise comparison with the no additional treatment arm. Each component of this composite outcome will also be summarised. Patients who were already on invasive mechanical ventilation or ECMO at randomisation will be excluded from these analyses.

##### 5.1.2 *Subsidiary clinical outcomes*

###### *5.1.2.1 Use of ventilation (overall and by type)*

Counts and percentages will be presented by randomised group for patients who received any assisted ventilation, together with adjusted risk ratios and confidence intervals for each pairwise comparison with the no additional treatment arm estimated using log-binomial regression, as described above. The number of patients receiving the two main types of ventilation will also be reported: non-invasive ventilation (including CPAP, other non-invasive ventilation or high-flow nasal oxygen), and invasive mechanical ventilation (including ECMO). Patients who were already receiving ventilation[b] at randomisation will be excluded from these analyses.

[b] Participants recruited to the main randomisation prior to protocol version 9.1 who were already receiving oxygen at randomisation will also be excluded from these analyses (since it is not possible to distinguish those who were already receiving non-invasive ventilation).

###### *5.1.2.2 Duration of invasive mechanical ventilation (time to successful cessation of invasive mechanical ventilation)*

Successful cessation of invasive mechanical ventilation will be defined as removal of invasive mechanical ventilation within (and survival to) 28 days after randomisation. A time-to-event analysis will be used to compare each treatment group with the no additional treatment group using Cox proportional hazards regression to estimate the hazard ratio and its confidence interval, as described above. Kaplan-Meier curves will be drawn. Patients who die within 28 days of randomisation will be censored *after* 28 days after randomisation. Patients who were not already on invasive mechanical ventilation or ECMO at randomisation will be excluded from these analyses.

###### *5.1.2.3 Use of renal dialysis or haemofiltration*

Counts and percentages will be presented by randomised group and the adjusted risk ratio will be calculated for each pairwise comparison with the no additional treatment arm using log-binomial regression, with confidence intervals and p-values reported. Patients who were already on renal dialysis or haemofiltration at randomisation will be excluded from these analyses.

###### *5.1.2.4 Thrombotic event*

Counts and percentages will be presented by randomised group. The absolute risk differences (and associated confidence intervals) will also be estimated by applying the adjusted risk ratio (or its 95% upper and lower limits) to the risk in the no additional treatment arm and then calculating the absolute difference between these values and the risk seen in the no additional treatment arm. Type of thrombotic event will also be described: (i) acute pulmonary embolism; (ii) deep vein thrombosis; (iii) ischaemic stroke, (iv) myocardial infarction; (v) systemic arterial embolism; and (vi) all sites combined.

###### 5.1.3 *Virological outcomes*

###### *5.1.3.1 SARS-CoV-2 RNA levels in the nasopharynx*

For parts J, K and L only: Geometric mean and standard error SARS-CoV-2 RNA levels will be presented at days 3 and 5. Comparisons will be made between treatment groups. Estimates will be obtained from analysis of covariance (ANCOVA) using the log transformed values after adjustment for each participant's baseline value and the baseline characteristics as described in section 5.4. Missing values will be imputed using the same procedures set out for continuous early phase outcomes described in section 9.3.2.5.

###### *5.1.3.2 Influenza RNA levels in the nasopharynx*

For parts G, H and I only: Geometric mean and standard error influenza RNA levels will be presented at day 5. Comparisons will be made between treatment groups. Estimates will be obtained from analysis of covariance (ANCOVA) using the log transformed values after adjustment for each participant's baseline value and the baseline characteristics as described in section 5.4. Missing values will be imputed using the same procedures set out for continuous early phase outcomes described in section 9.3.2.5.

###### 5.1.3.3 SARS-CoV-2 and influenza viral resistance markers

Counts and percentages of SARS-CoV-2 (for parts J, K and L) and influenza (for parts G, H and I) viral resistance markers will be presented by randomised group.

#### 5.2 Second randomisation

Evaluation of treatment effects in the main randomisation and the second randomisation will be conducted independently, as described in 5.1.

#### 5.3 Pre-specified subgroup analyses

Pre-specified subgroup analyses will be conducted for each part of the main randomisation and for the second randomisation, for the following outcomes:

- Mortality (all-cause)
- Time to discharge from hospital
- Use of invasive mechanical ventilation (including ECMO) or death

Tests for heterogeneity (or tests for trend for 3 or more ordered groups) will be conducted to assess whether there is any good evidence that the effects in particular subgroups differ materially from the overall effect seen in all patients combined. Results will be presented on forest plots as hazard ratios, or risk ratios, with confidence intervals. The following subgroups will be examined based on information at randomisation:

- Age (<70; 70-79; 80+ years)
- Sex (Male; Female)
- Ethnicity (White; Black, Asian or Minority Ethnic)
- Region (UK, non-UK)
- Time since illness onset (≤7 days; >7 days)
- Requirement for respiratory support
  - For main randomisation: None; Oxygen only; Non-invasive ventilation; Invasive mechanical ventilation (including ECMO)[c]
  - For second randomisation: No ventilator support (including no or low-flow oxygen); Non-invasive ventilation (including CPAP, other non-invasive ventilation, or high-flow nasal oxygen), Invasive mechanical ventilation (including ECMO)
- CURB-65 score (<3; ≥3)
- Use of systemic corticosteroid (including dexamethasone)
- Concomitant viral infection:
  - For parts J, K and L: presence or absence of confirmed influenza infection
  - For parts G and H: presence or absence of confirmed SARS-CoV-2 infection
- For part B only: Recipient anti-SARS-CoV-2 anti-S antibody status at randomisation (negative, positive as defined by the assay manufacturer, Roche). (This is the key subgroup for the REGEN-COV2 comparison; see Appendix I.)

[c] Participants recruited before protocol V9.1 who were receiving oxygen would be presented in a fifth subgroup but not included in the test for trend

- For parts J, K and L only: Recipient anti-SARS-CoV-2 anti-N antibody concentration at randomisation (negative; positive) as defined by the assay manufacturer, Roche). See Appendix II.
- For parts J, K and L only: Serum SARS-CoV-2 antigen level (< and ≥ median) at randomization. See Appendix II.
- For parts G, H, I and M only; presence of lung consolidation on imaging
- Immunosuppression in the opinion of the managing doctor

#### 5.4 Adjustment for baseline characteristics

The main analyses described above will be adjusted for age and requirement for respiratory support at baseline (using categories defined in section 4.2). Adjustment for these two major predictors of mortality is desirable because it provides a safeguard against the impact that any chance imbalances in their frequencies between randomised groups may have on the randomised comparisons, whilst also leading to a small expected increase in statistical power. Analyses with *further* adjustment for other pre-specified subgroups (see section 4.2) will also be done and presented as sensitivity analyses.

#### 5.5 Sensitivity analyses

For parts A to F only, sensitivity analyses of the primary and secondary outcomes will be conducted among those patients with a positive test for SARS-COV-2 (i.e. confirmed cases).

Sensitivity analyses of the primary and secondary outcomes will be conducted (a) without adjustment for characteristics at randomisation and (b) with adjustment for all key baseline pre-specified subgroups (see section 5.4).

#### 5.6 Other exploratory analyses

In addition, for each randomised assessment, exploratory analyses will be conducted to test for interactions with other treatments allocated in each of the different randomisations, provided that doing so does not lead to premature unblinding of results for ongoing comparators.

Non-randomised exploratory analyses will be used to explore the likely influence of different levels of convalescent plasma antibody concentrations on the efficacy of convalescent plasma.

Additional analyses will set the results for children (<18 years) and pregnant women in the context of the overall results.

#### 5.7 Significance levels and adjustment of p-values for multiplicity

Evaluation of the primary trial (main randomisation) and secondary randomisation will be conducted independently, and no adjustment be made for these. Formal adjustment will not be made for multiple treatment comparisons, the testing of secondary and subsidiary outcomes, or subgroup analyses (with one exception; see Appendix I). However, due allowance for multiple testing will be made in the interpretation of the results: the larger the number of events on which a comparison is based and the more extreme the P-value after any allowance has been made for the nature of the particular comparison (i.e. primary or secondary; pre-specified or exploratory), the more reliable the comparison and, hence, the more definite any finding will be considered. 95% confidence intervals will be presented for the main comparisons.

For each of the influenza comparisons (parts G, H, and I), Holm's procedure will be used to control the family-wise error rate across the two co-primary outcomes at 5%.[7]

#### 5.8 Statistical software employed

The statistical software SAS version 9.4 and R Studio 4.2.3 (or later) for Windows will be used for the interim and final analyses.

#### 5.9 Data standards and coding terminology

Datasets for analysis will be prepared using CDISC standards for SDTM and ADaM. Wherever possible, clinical outcomes (which may be obtained in a variety of standards, including ICD10 and OPCS-4) will be coded using MedDRA version 20.1.

### 6 SAFETY DATA

Suspected serious adverse reactions (SSARs) and suspected unexpected serious adverse reactions (SUSARs) will be listed by trial allocation.

For each of the following, counts and percentages will be presented by randomised group. Where possible, the absolute risk differences will also be presented with confidence intervals (estimated using the methods described in section 5.1.2.4).

#### 6.1 Cause-specific mortality

Cause-specific mortality (COVID-19, other infection, cardiac, stroke, other vascular, cancer, other medical, external, unknown cause) will be analysed in a similar manner to the primary outcome.

#### 6.2 Major cardiac arrhythmia

Type of arrhythmia will also be described: (i) atrial flutter or fibrillation; (ii) supraventricular tachycardia; (iii) ventricular tachycardia; (iv) ventricular fibrillation; (v) atrioventricular block requiring intervention, with subtotals for (i)-(ii) and (iii)-(iv).

#### 6.3 Major bleeding

Type of bleeding will also be described: (i) intracranial bleeding; (ii) gastro-intestinal bleeding; (iii) other bleeding site, and (iv) all sites combined.

#### 6.4 Early safety of anti-coronavirus antibody-based therapy

Additional safety data will be collected in a subset of patients randomised to part B: (i) sudden worsening in respiratory status; (ii) severe allergic reaction; (iii) temperature >39°C or ≥2°C rise since randomisation; (iv) sudden hypotension; (v) clinical haemolysis; and (vi) thrombotic event.

#### 6.5 Other infections

Other infections occurring after randomisation will be described. These will be classified primarily by site (pneumonia, urinary tract, biliary, other intra-abdominal, blood stream, skin, other). Information on putative organism (other virus, bacterial, fungal, other and unknown) is also collected.

#### 6.6 Metabolic, kidney and liver complications

Incidence of the following metabolic and biochemical complications after randomisation will be described:

- Severe hyperglycaemia (separately and overall; introduced 28 Jul 2021):
  - Ketoacidosis (defined as combination of ketosis [blood ketones ≥1.5 mmol/L or urine ketones ≥2+] and acidosis [venous bicarbonate <15 mmol/L])
  - Hyperglycaemic hyperosmolar state (defined as glucose >33 mmol/L and calculated osmolality >320 mOsm/L)
  - Other hyperglycaemia requiring new use of insulin
- Severe hypoglycaemia (causing reduced conscious level requiring another person to help recover; introduced 28 Jul 2021)
- Acute kidney injury (defined as ratio of post-randomisation peak creatinine to baseline value >1.5x or new use of renal dialysis/haemofiltration; introduced 28 Jul 2021):
- Liver dysfunction (for parts K and L and part G; introduced 28 Mar 2022):
  - Peak alanine (or aspartate) transaminase in the following categories (<3 x upper limit of normal [ULN]; ≥3 <5x ULN; ≥5x ULN)
  - Peak bilirubin (≤2x ULN; >2x ULN)
  - Possible liver injury (defined as ALT >3x ULN plus bilirubin >2x ULN)

#### 6.7 Seizures

The incidence of seizures (introduced on 28 Mar 2022)

#### 6.8 Infusion reactions to sotrovimab

For part J, the frequency and percentage of infusion reactions to sotrovimab will described (overall and by severity)

### 7 ADDITIONAL POST-HOC EXPLORATORY ANALYSIS

Any post-hoc analysis requested by the oversight committees, a journal editor or referees will be labelled explicitly as such. Any further future analyses not specified in the analysis protocol will be exploratory in nature and will be documented in a separate statistical analysis plan.

### 8 DIFFERENCES FROM PROTOCOL

The testing of multiple treatment arms will not formally be adjusted for, but given the number of comparisons, due allowance will be made in their interpretation. Formal methods of adjustment for multiplicity were not adopted because of treatment arms being added over time (including the factorial convalescent plasma comparison), unequal recruitment into each arm, and the ultimate number of treatments under evaluation not being known in advance.

### 9 EARLY PHASE ASSESSMENTS

The following approach is required for the evaluation of treatments indicated as undergoing Early Phase Assessment in the protocol (introduced in Protocol version 14.0):

#### 9.1 Definitions of clinical outcomes

##### 9.1.1 *Primary outcome*

- WHO ordinal scale on day 5

##### 9.1.2 *Secondary clinical outcomes*

- Time to sustained improvement (i.e., value better than baseline value persisting for >1 day) by at least one category on the WHO ordinal scale from baseline
- $S/F_{94}$ ratio at day 5
- Time to discharge from hospital
- Improvement in clinical status at day 10
- Blood C-reactive protein at day 5

##### 9.1.3 *Subsidiary clinical outcomes*

- All other subsidiary outcomes as described above (section 2.6.3)

##### 9.1.4 *Safety outcomes*

- Flushing (incidence, severity)
- Gastrointestinal symptoms (incidence, severity)
- Reasons for stopping study treatment
- Transaminitis (ALT >3x upper limit of normal)
- Acute kidney injury (creatinine >1.5x value entered at randomisation)
- All other subsidiary outcomes as described above (section 2.6.5)

#### 9.2 Baseline comparability of randomised groups

Unless otherwise specified, analyses will follow the plan described above (section 4). In addition, the following characteristics will be described:

- Oxygen saturation measurement on air (if available)
- $S/F_{94}$ ratio
- WHO Ordinal Scale
- All other characteristics as described above (section 4.2)

#### 9.3 Comparative analysis

Unless otherwise specified, comparative analyses will follow the plan described above (section 5). In addition,

##### 9.3.1 *Primary outcome*

The primary comparison will involve an "intention to treat" analysis among all participants randomised between the active arm and its control of the effect of the active treatment on WHO scale at day 5, adjusted for baseline score. A proportional odds model will be used to assess the common odds ratio of better outcome for each pairwise comparison with the no additional treatment arm.[8] In addition, a sensitivity analysis to the proportional odds model using Howard's method will be performed if the proportional odds assumption is not satisfied.[9]

##### 9.3.2 *Secondary outcomes*

###### *9.3.2.1 Time to sustained improvement by at least one category on the WHO ordinal scale from baseline*

A time-to-event analysis will be used to compare each treatment group with the no additional treatment group using the log-rank test (restricted to the first 10 days of the trial as the WHO score is not collected after this). The rate ratio and its confidence interval will be estimated from the log-rank observed minus expected statistic and its variance, and Kaplan-Meier curves will be drawn.

###### *9.3.2.2 Improvement in clinical status at day 10*

Counts and percentages will be presented by randomised group for patients with an improvement of at least one category on the WHO ordinal scale from baseline, together with odds ratios and confidence intervals for each pairwise comparison with the no additional treatment arm.

###### *9.3.2.3 Blood C-reactive protein at day 5*

Geometric mean C-reactive protein at day 5 will be compared between treatment arms. Estimates will be obtained from analysis of covariance (ANCOVA) for the log transformed CRP values after adjustment for each participant's baseline value. Approximate standard errors for the geometric means will be calculated from the confidence intervals. Missing CRP values will be handled as described in section 9.3.2.5.

###### *9.3.2.4 $S/F_{94}$ ratio at day 5*

Mean $S/F_{94}$ ratio at day 5 will be compared between treatment arms. Estimates will be obtained from analysis of covariance (ANCOVA) after adjustment for each participant's baseline $S/F_{94}$ ratio. Missing $S/F_{94}$ ratio values will be handled as described in section 9.3.2.5

###### *9.3.2.5 Imputation of missing data*

All analyses will be done according to the intention-to-treat principle and, hence, missing secondary outcome data will be imputed. For each of the continuous outcomes (e.g., CRP, $S/F_{94}$ ratio) missing post-randomisation results will be imputed using multiple imputation, using 20 imputed data sets, with results across imputations being combined using the methods of Rubin.[10] The imputation procedure will take into consideration each participant's key baseline characteristics (listed in section 5.8), treatment allocation and any intermediate follow-up values of the biomarker, where available. For $S/F_{94}$ ratio, WHO ordinal scale values on days 3 and 5 will also be used in the imputation procedure. For patients who are discharged from hospital and for whom it is not possible to measure $S/F_{94}$ ratio at day 5, a value of 4.76[d]

[d] 4.76 = 1.0/0.21 (ie, the value of healthy lungs which provide 100% saturations when breathing 21% oxygen)

will be imputed. The results from these analyses will be compared with those from equivalent "complete-case" analyses, but primary emphasis will be placed on the results after multiple imputation. All multiple imputation analyses will be implemented using the multiple imputation procedure in SAS version 9.4 (SAS Institute, Cary NC), using the expectation-maximization algorithm (which assumes a multivariate normal distribution) to impute values. For any continuous variables with missing baseline values, the mean among those with observed values will be imputed.

###### 9.3.3 *Safety outcomes*

Counts and percentages will be presented by randomised group. The absolute risk differences will also be presented with confidence intervals for each of the following:

- Flushing (incidence, severity)
- Gastrointestinal symptoms (incidence, severity)
- Reasons for stopping study treatment
- Transaminitis (ALT >3x upper limit of normal)
- Acute kidney injury (creatinine >1.5x value entered at randomisation)

#### 10 6-MONTH AND LONGER-TERM ASSESSMENTS

This section details the proposed analysis of the clinical outcomes 6 months after initial randomisation in the RECOVERY trial (for all participants). A similar approach will be used for analyses of longer-term outcomes for UK participants only.

##### 10.1 Objectives

The **primary objective** of these analyses is to provide reliable estimates of study treatments on all-cause mortality within 6 months of the relevant randomisation.

The **key safety objectives** are to provide reliable estimates of these on non-COVID infections and non-COVID causes of death.

##### 10.2 Comparative analyses at 6 months

The primary analyses will be performed on the ITT population at 6 months. Pairwise comparisons will be made between each treatment arm and the no additional treatment arm (reference group) in that particular randomisation using the same approach described for the 28 day analyses (see section 5).

###### 10.2.1 *Primary outcome*

The primary outcome is **6-month mortality** (all-cause). This will be summarised with counts and percentages by randomised comparison group. A time-to-event analysis will be conducted using the approach described for the 28-day analyses (see section 5.1.1.1). Results will be interpreted in the context of the results of analyses at 28 days.

For the analysis of REGEN-COV2, the primary outcome will first be assessed among those participants who are known to be seronegative (anti-S SARS-CoV-2 antibody negative) at randomisation (see Appendix I).

For the analysis of sotrovimab, the primary outcome will first be assessed among participants with a blood SARS-CoV-2 antigen concentration above the median value (see Appendix II).

##### 10.2.2 *Pre-specified subgroup analyses*

Subgroup analyses will be conducted for 6-month mortality (all-cause) using methods described in section 5.3. The following subgroups will be examined based on information at randomisation:

- For dexamethasone comparisons: Requirement for respiratory support (with test for trend)
- For tocilizumab comparison: Use of systemic corticosteroid (including dexamethasone) (with test for heterogeneity)
- For REGEN-COV2 comparison: Recipient anti-SARS-CoV-2 anti-S antibody concentration (positive, negative, unknown; with test for heterogeneity between seronegative and seropositive participants)
- For baricitinib comparison: use of systemic corticosteroid (including dexamethasone) and, separately, use of interleukin-6 antagonist (e.g. tocilizumab, sarilumab) (each with test for heterogeneity)
- For sotrovimab comparison: Recipient anti-SARS-CoV-2 anti-N antibody concentration (positive, negative, unknown; with test for heterogeneity between seronegative and seropositive participants) and SARS-CoV-2 antigen level (< or ≥ median)
- For high-dose dexamethasone comparison: Requirement for respiratory support (with test for trend)

Other subgroup analyses (see section 5.3) may be conducted but will be considered exploratory in nature.

##### 10.2.3 *Adjustment for baseline characteristics*

The main analyses will be adjusted for age and level of respiratory support at baseline (and potentially for other important imbalances) using the approach described in section 5.1.1.1.

##### 10.2.4 *Sensitivity analyses*

Sensitivity analyses of the primary outcome will be conducted with adjustment for all key baseline pre-specified subgroups (see section 5.4).

##### 10.2.5 *Significance levels and adjustment of p-values for multiplicity*

This will take the same approach as described for 28-day analyses (see section 5.7)

#### 10.3 Safety data

The key safety outcome is **major non-COVID infection** (associated with hospitalisation or death). These will be presented overall, and by site (e.g. pneumonia, urinary tract, biliary, other intra-abdominal, bloodstream, skin, other) and, where possible, by putative organism (e.g. virus, bacteria, fungus, other). Counts and percentages will be presented by randomised group. Where possible, the absolute risk differences will also be presented with confidence intervals.

#### 10.4 Other exploratory analyses

Secondary, subsidiary clinical and other safety outcomes (as specified earlier in this document) may be assessed in exploratory analyses. In addition, hospital recorded diagnoses (see Definition and Derivation of Baseline Characteristics and Outcomes SOP section 8.1.2) may be explored to assess other long-term effects of study treatments.

The selection and interpretation of these additional analyses will be informed by the 28-day results and what is known about the potential longer term impacts of the study treatments (particularly with respect to known hazards of treatment).

#### 10.5 Censoring and analysis

For the 6 month analyses, participants will be censored at the earliest of death, withdrawal of consent, known exit from the NHS,[e] or on study day 184 (where day of randomisation is study day 1). For later analyses, a similar censoring approach will be used (e.g. day 731 for a 2 year analysis).

[e] NHS England (and equivalent organisations in the devolved nations) are notified if patients are no longer receiving NHS care (typically due to emigration).

### 11 REFERENCES

#### 11.1 Trial documents

Study protocol, case report forms, training materials, and statistical analysis plan are published on the trial website.

#### 11.2 Other references

10. Rubin D. Multiple imputation for non-response in surveys. New York: John Wiley; 1987.

### 12 APPENDIX I: ANALYSES OF REGEN-COV2

#### 12.1 Background & rationale

The RECOVERY trial is testing multiple interventions in a broad population of patients hospitalised with COVID-19. The protocol and statistical analysis plan outline the methods that are to be used in the analysis of these interventions and, to date, the same approach has been appropriate for all completed comparisons. However, it is important that the statistical analysis plan be informed by the best available information about the treatment being tested[1] and the pathophysiology of the disease.

Relevant new information about the effects of REGEN-COV2 have emerged since it was added to the trial in September 2020.

REGEN-COV2 is a mixture of two synthetic monoclonal antibodies which bind to the receptor binding domain of the SARS-CoV-2 spike protein and neutralise the virus.[2] Recently-published trials of REGEN-COV2 in ambulatory patients (i.e. those recently diagnosed in the community) have demonstrated that it has larger effects on viral load among people who are “seronegative” at the time of randomisation (i.e. they do not have detectable antibodies of their own against SARS-CoV-2), and seropositive patients derive little or no benefit (in terms of reduction in viral load) from REGEN-COV2, compared to placebo.[3] Participant serostatus therefore is a potentially key modifier of the effect of REGEN-COV2 that may be observed in RECOVERY.

All participants entering the REGEN-COV2 comparison in RECOVERY are asked to provide a serum sample which is sent to a central laboratory at the University of Oxford, where antibodies against SARS-CoV-2 are measured using a validated assay. Previous assessments of this assay alongside commercially available assays shows excellent performance at discriminating prior SARS-CoV-2 infection with sensitivity and specificity above 98%.[4]

Earlier versions of the statistical analysis plan recognised the importance of the seronegative subgroup, but review of the emerging literature and regulatory guidance[5] has led to a change in approach to these analyses. The revised analysis plan for the REGEN-COV2 comparison explicitly tests the hypothesis that any benefit of REGEN-COV2 on the primary outcome may be wholly or largely restricted to patients who are seronegative at the time of randomisation with little or no benefit among those who are seropositive at that point.

For the avoidance of doubt, all decisions about this modification to the analytical plan were made before recruitment was complete and before any members of the trial steering committee (who are responsible for drafting and approving the SAP) or investigators had access to any unblinded analyses of clinical outcome data for the REGEN-COV2 comparison. No members of the independent Data Monitoring Committee (who are the only individuals who can review interim unblinded analyses) were involved in this change.

#### 12.2 Analytical plan

The primary outcome and secondary outcomes remain unchanged. For each outcome, rate ratios and 95% confidence intervals will be calculated separately for participants who are seronegative, seropositive, or with unknown status as well as for the whole trial population. A test for heterogeneity between seronegative and seropositive participants will be presented. The results will be interpreted based on the totality of the evidence.

For the purposes of any regulatory submission: Because any beneficial effect of REGEN-COV2 is hypothesised to be larger among seronegative participants (and may be negligible in seropositive participants), the primary outcome will first be assessed among participants who are known to be seronegative at randomisation. If the null hypothesis is rejected in the seronegative group at 2-tailed p=0.05, then the primary outcome will be assessed among the whole population (i.e. seronegative, seropositive, and those with unknown status combined). Otherwise, no further hypothesis testing will be performed.

A similar approach will be taken for each of the two pre-specified secondary outcomes (discharge alive within 28 days and, among patients not on invasive mechanical ventilation at baseline, the use of invasive mechanical ventilation or death) if both primary hypotheses are rejected. Hypothesis testing will first be conducted among the participants who are known to be seronegative at randomisation and, if the null hypothesis is rejected at 2-tailed p=0.025, then will be assessed among the whole population (see Table).

**Table: Hierarchical Testing Order**

| Hierarchy Number | Type of Outcome | Outcome | Analysis Population | Significance level, α (2-sided) |
|---|---|---|---|---|
| 1. | Primary | Mortality (all-cause), 28 days after randomisation | Seronegative at randomisation | 0.05 |
| 2. | Primary | Mortality (all-cause), 28 days after randomisation | All participants randomised | 0.05 |
| 3.* | Secondary | Time to discharge alive from hospital, within 28 days after randomisation | Seronegative at randomisation | 0.025 |
| 4. | Secondary | Time to discharge alive from hospital, within 28 days after randomisation | All participants randomised | 0.025 |
| 3.* | Secondary | Use of invasive mechanical ventilation (including ECMO) or death | Seronegative and not on invasive mechanical ventilation at randomisation | 0.025 |
| 4. | Secondary | Use of invasive mechanical ventilation (including ECMO) or death | All participants randomised not on invasive mechanical ventilation at randomisation | 0.025 |

* These will be performed simultaneously. Testing will only proceed to the respective overall population if the null hypothesis is rejected in the seronegative group at the specified level of statistical significance.

### 13 APPENDIX II: ANALYSES OF SOTROVIMAB AND OTHER ANTI-VIRALS FOR COVID-19

#### 13.1 Background & rationale

The RECOVERY trial is testing multiple interventions in a broad population of patients hospitalised with COVID-19. The protocol and statistical analysis plan outline the methods that are to be used in the analysis of these interventions. With the exception of REGEN-COV2 (see Appendix I), the same approach has been used for all completed comparisons. However, it is important that the statistical analysis plan be informed by the best available information about the treatment being tested[1] and the pathophysiology of the disease.

At the point at which sotrovimab was added to the protocol (protocol version 21.1; approved 20th December 2021), the number of infections with the omicron variant of SARS-CoV-2 was rising exponentially, doubling approximately every 2 days. There was an enormous national effort to maximise vaccination such that by 20th December 2021, around 90% of adults aged >18 years had received a 1st dose of vaccine, 82% had received 2 doses, and 50% had received 3 doses (with around 0.5-1 million vaccine doses being administered each day). However, there were several important unknowns including the propensity for the omicron variant to cause severe disease, hospitalisation and death (either with or without vaccination).

The previous evaluation of REGEN-COV2 in RECOVERY established that the monoclonal neutralising antibody combination was effective in patients who were anti-spike antibody negative, and no meaningful effect was seen among those who were anti-spike antibody positive. However, that evaluation was carried out prior to the emergence of the omicron variant and at a point when <10% participants had any vaccine dose (and almost nobody had had more than one). Hence, seropositive status at that time largely reflected an acute immune response to the active SARS-CoV-2 infection. By December 2021, the situation was more complicated – seropositive status could reflect an acute immune response (as before) or a legacy effect of prior infection (with a different variant) or prior vaccination (against a different variant). Given the immune escape demonstrated by omicron, it is reasonable to expect that at least some seropositive patients may benefit from treatment with a neutralising monoclonal antibody in the form of sotrovimab or an anti-viral treatment such as molnupiravir or paxlovid.

There is some evidence from the ACTIV-3 study programme[2] that serum viral antigen concentration may be a useful predictor of both poor outcome and of response to monoclonal neutralising antibody treatment. Serum samples will be collected and analysed for both anti-SARS-CoV-2 antibody concentration and viral antigen concentrations. The TSC will review data on the distribution of these and their association with primary and secondary outcomes (blinded to information about treatment allocation) before determining the most scientifically and clinically relevant primary analysis population.[3] (For example, the TSC might determine that the primary analysis should be restricted to those patients who are anti-N antibody negative or alternatively who have high viral antigen load, and decide on an analysis approach analogous to that used for patients who were seronegative [anti-S antibody negative] in the REGEN-COV2 analysis.)

For the avoidance of doubt, all decisions about this modification to the analytical plan will be made before before any members of the trial steering committee (who are responsible for drafting and approving the SAP) or investigators have access to any unblinded analyses of clinical outcome data for the sotrovimab, molnupiravir and paxlovid comparisons. No members of the independent Data Monitoring Committee (who are the only individuals who can review interim unblinded analyses) will be involved in this decision.

**Addendum 25 June 2024: Selection of primary analysis population for the sotrovimab, molnupiravir and Paxlovid comparisons**

*Paxlovid and molnupiravir*
A substantial fall in the number of patients admitted to hospital with COVID-19 lung disease from early 2022 meant that recruitment to these comparisons remained low (924 participants in the molnupiravir comparison, and 137 participants in the Paxlovid comparison). Because of low recruitment these comparisons were closed early, in May 2023. Selecting a subgroup as the primary analysis population was considered futile, and so *for these comparisons all analyses based on viral antibody/antigen status are exploratory*.

*Sotrovimab*
Recruitment to the sotrovimab comparison closed on 31st March 2024 when funding ended. At the time of finalising this SAP (version 5.0) the RECOVERY investigators remain blind to the results of this comparison.

By January 2024, SARS-CoV-2 seropositivity among blood donors was 90% for anti-N antibodies (reflecting previous infection) and 99.9% for anti-S antibodies (reflecting previous infection or vaccination).[4] As a result, the presence of SARS-CoV-2 antibodies in hospitalised patients is now likely to reflect previous infection or vaccination rather than indicate the adequacy of the immune response to their current infection.

A review of participant serostatus and outcomes in the sotrovimab comparison (blinded to treatment allocation) supports this. In September 2023, only 19% of participants were anti-S negative, making this unsuitable as the primary analysis population. 70% were anti-N negative, most of whom were recruited in early 2022 when anti-N seroprevalence in the UK population was low. However, anti-N seronegativity was not associated with increased mortality (20% among those who were seropositive vs 21% among those who were seronegative). This contrasts with RECOVERY participants recruited in 2020/21, in whom mortality was strongly associated with anti-N serostatus (14% among those who were seropositive vs 30% among those who were seronegative in patients not receiving neutralising antibody therapy), and suggests that anti-N serostatus is no longer a good marker of immune response to the acute infection.[5]

Unlike measurement of anti-SARS-COV-2 antibodies, the detection of viral nucleocapsid antigen in the blood must relate to the current infection, and may represent a phase of infection before specific antibodies have developed to clear the virus. In keeping with this, viral nucleocapsid antigen concentration above the median value (cut-off index 0.626) is associated with significantly higher mortality in the sotrovimab comparison (26% vs 16%). Participants with higher blood antigen concentration seem most likely to benefit from antibody therapy, and so ***the primary analysis population for the sotrovimab comparison will be participants who have a blood antigen concentration above the median value.***

The primary outcome and secondary outcomes remain unchanged. For each outcome, rate ratios and 95% confidence intervals will be calculated separately for participants who have high antigen, low antigen, or with unknown status, as well as for the whole trial population. A test for heterogeneity between participants with high and low antigen will be presented. The results will be interpreted based on the totality of the evidence.

Because any beneficial effect of sotrovimab is hypothesised to be larger among high antigen participants (and may be negligible in low antigen participants), the primary outcome will first be assessed among participants who are known to have high antigen at randomisation. If the null hypothesis is rejected in the high antigen group at 2-tailed p=0.05, then the primary outcome will be assessed among the whole population (i.e. high antigen, low antigen and those with unknown status combined). Otherwise, no further hypothesis testing will be performed.

A similar approach will be taken for each of the two pre-specified secondary outcomes (discharge alive within 28 days and, among patients not on invasive mechanical ventilation at baseline, the use of invasive mechanical ventilation or death) if both primary hypotheses are rejected. Hypothesis testing will first be conducted among the participants who are known to be high antigen at randomisation and, if the null hypothesis is rejected at 2-tailed p=0.025, then will be assessed among the whole population (see Table).

**Table: Hierarchical Testing Order**

| **Hierarchy Number** | **Type of Outcome** | **Outcome** | **Analysis Population** | **Significance level, α (2-sided)** |
|---|---|---|---|---|
| 1. | Primary | Mortality (all-cause), 28 days after randomisation | High antigen at randomisation | 0.05 |
| 2. | Primary | Mortality (all-cause), 28 days after randomisation | All participants randomised | 0.05 |
| 3.* | Secondary | Time to discharge alive from hospital, within 28 days after randomisation | High antigen at randomisation | 0.025 |
| 4. | Secondary | Time to discharge alive from hospital, within 28 days after randomisation | All participants randomised | 0.025 |
| 3.* | Secondary | Use of invasive mechanical ventilation (including ECMO) or death | High antigen and not on invasive mechanical ventilation at randomisation | 0.025 |
| 4. | Secondary | Use of invasive mechanical ventilation (including ECMO) or death | All participants randomised not on invasive mechanical ventilation at randomisation | 0.025 |

* These will be performed simultaneously. Testing will only proceed to the respective overall population if the null hypothesis is rejected in the high antigen group at the specified level of statistical significance.

### 14 APPROVAL

| | | |
|---|---|---|
| Chief Investigator | Name: Professor Peter Horby | |
| | Signature: | Date: |
| Deputy Chief Investigator | Name: Professor Martin Landray | |
| | Signature: | Date: |
| Steering Committee Statistician | Name: Professor Thomas Jaki | |
| | Signature: | Date: |

#### 15 DOCUMENT HISTORY

| Version | Date | Edited by | Comments/Justification | Timing in relation to unblinded interim monitoring | Timing in relation to unblinding of Trial Statisticians |
|---|---|---|---|---|---|
| 0.1 | 20/03/20 | LL/JB | First draft. | Prior | Prior |
| 0.2 | 01/04/20 | LL/JB | Comments and amendments from Martin Landray, Jonathan Emberson & Natalie Staplin. Also aligned with updated protocol and CRFs. | Prior | Prior |
| 0.3 | 01/04/20 | EJ/LL | Further edits and comments. | Prior | Prior |
| 0.4 | 07/04/20 | JB/EJ/LL | Following statistics group meeting on 02/04/20. | Prior | Prior |
| 0.5 | 22/04/20 | JB/LL/EJ | Following statistics group meeting on 09/04/20 and further protocol update. | After | Prior |
| 0.6 | 24/04/20 | LL | Following statistics group meeting on 23/04/20. | After | Prior |
| 0.7 | 10/05/20 | LL | Protocol update. | After | Prior |
| 0.8 | 15/05/20 | LL | Following statistics group meeting on 15/05/20. | After | Prior |
| 0.9 | 27/05/20 | LL | Further comments from TSC members prior to interim analysis on 28/05/20. | After | Prior |
| 1.0 | 09/06/20 | LL | Revised following the stopping of the hydroxychloroquine arm, and prior to the trial statisticians receiving unblinded data for this arm. | After | Prior |
| 1.1 | 21/06/20 | LL/JB/RH | Additional clarification of ventilation denominators. Adjustment for any imbalances of subgroup characteristics between treatment arms at randomisation. Clarification of analysis of composite outcome. Removal of 'Unknown' ethnicity subgroup. Addition of section 5.5 Adjustment for baseline characteristics. | After | After unblinding of hydroxychloroquine and dexamethasone arms. |

| Version | Date | Edited by | Comments/Justification | Timing in relation to unblinded interim monitoring | Timing in relation to unblinding of Trial Statisticians |
|---|---|---|---|---|---|
| 2.0 | 04/11/20 | EJ/ES | Revised to reflect changes in protocol, including introduction of factorial randomisations and new arms, including convalescent plasma, tocilizumab, synthetic neutralizing antibodies (REGEN-COV2, and aspirin. | Prior to interim analysis of aspirin arm<br><br>After interim analyses of all other arms | After unblinding of 28-day results for hydroxychloroquine, lopinavir-ritonavir, and dexamethasone arms.<br><br>Prior to unblinding of any other arms |
| 2.1 | 02/12/20 | ES | Addition of colchicine. Modification of definition of recipient antibody concentration subgroup. | Prior to interim analyses including antibody results or of colchicine arm. | After unblinding of 28-day results for hydroxychloroquine, lopinavir-ritonavir, and dexamethasone arms.<br><br>Prior to unblinding of any other arms |
| 2.2 | 27/01/21 | ES | Clarification of non-invasive ventilation-related subgroups. Addition of baricitinib. | Prior to interim analyses of baricitinib arm. | After unblinding of 28-day results for hydroxychloroquine, lopinavir-ritonavir, azithromycin and dexamethasone arms (and primary outcome in overall population in convalescent plasma arm).<br><br>Prior to unblinding of any other arms |

| Version | Date | Edited by | Comments/Justification | Timing in relation to unblinded interim monitoring | Timing in relation to unblinding of Trial Statisticians |
|---|---|---|---|---|---|
| 3.0 | 15/05/21 | ES | Specification of method for REGEN-COV2 comparison (appendix A).<br>Addition of early phase assessment of dimethyl fumarate.<br>Addition of infliximab and high-dose corticosteroids. | Prior to interim analyses of infliximab or high-dose steroids. | After unblinding of 28-day results for hydroxychloroquine, lopinavir-ritonavir, azithromycin, dexamethasone, colchicine and convalescent plasma arms.<br><br>Prior to unblinding of any other arms. |
| 3.1 | 29/10/21 | RH | Modification of early phase assessments to align with protocol V18.1<br>Modification of 6 months analysis section. | Prior to early phase assessment s or 6 month analyses. | Prior to unblinding of dimethyl fumarate or 6 month outcome data. |
| 3.2 | 17/12/21 | RH | Update to early phase assessments | Prior to 6 month analyses | Prior to unblinding of dimethyl fumarate |
| 4.0 | 20/09/22 | MJL | Revised to reflect changes in protocol versions 19-25. Now includes information on comparisons for influenza and for sotrovimab, molnupiravir and paxlovid. Update to 6 month and long-term assessments. | Prior to interim analyses of these arms. | Prior to commencement of enrolment to influenza comparisons. Prior to unblinding of sotrovimab, molnupiravir, paxlovid, empagliflozin, and high dose corticosteroid comparisons for participants on non-invasive ventilation or invasive mechanical ventilation. |

| Version | Date | Edited by | Comments/Justification | Timing in relation to unblinded interim monitoring | Timing in relation to unblinding of Trial Statisticians |
|---|---|---|---|---|---|
| 5.0 | 25/06/24 | LP | Addition of CAP comparison. Update to reflect opening of flu comparisons & closing of empagliflozin, molnupiravir, Paxlovid, high dose corticosteroid, and sotrovimab comparisons. Finalisation of analysis plan for SARS-CoV-2 antivirals in Appendix II. | After interim analyses of all comparisons. | Prior to unblinding of sotrovimab, flu & CAP comparisons (parts G, H, I, J & M).<br>After unblinding of molnupiravir & Paxlovid comparisons (parts K & L) |

#### Appendix 3: Definition and Derivation of Baseline Characteristics and Outcomes

RECOVERY
Randomised Evaluation of COVID-19 Therapy

### Definition and Derivation of Baseline Characteristics and Outcomes

13 Appendix 5: Example hospital recorded diagnoses showing extraction of start and end dates 23

### 1 Version

| Date | Version | Comments |
|---|---|---|
| 06-Jun-2020 | 0.1 | Initial version |
| 08-Jun-2020 | 0.2 | Minor updates |
| 09-Jun-2020 | 1.0 | First released version |
| 11-Dec-2020 | 2.0 | Update to sections 6.4 (use of assisted ventilation) and 6.6 (use of renal replacement therapy) |
| 06-Jan-2020 | 3.0 | Update to clarify the derivation of outcomes and baseline data for the second randomisation and define complete follow-up |
| 14-April-2022 | 4.0 | Updates to frequency of dataset transfers and additional datasets. Addition of section 8relating to 6-month outcomes. Addition of appendix 4 to provide detail on discharge outcome |

### 2 Scope

This document describes the definition and derivation of the primary, secondary and other outcomes of the RECOVERY trial for the published trial analyses. It should be read alongside the study protocol which defines the study outcomes briefly, and the Statistical Analysis Plan (SAP) which describes the statistical methods used to analyse these outcomes. The SAP refers to this document (see Section 2.6.4 Detailed derivation of outcomes) which provides detail on how the outcomes are defined, captured and derived.

Most outcomes have more than one potential source which improves completeness of capture but also will inevitably identify discrepancies between different sources. This document describes the principles for how such discrepancies are resolved; the rules for this were developed blind to results. Further details of the methods are described in the RECOVERY trial internal operating procedure for identifying data discrepancies.

### 3 Abbreviations

| | |
|---|---|
| ADDE | Annual District Death Extract |
| CCDS | Critical Care Dataset |
| CHESS | COVID-19 Hospitalisation in England Surveillance System |
| CPAP | Continuous Positive Airway Pressure |
| CRP | C-reactive protein |
| ECMO | Extra-corporeal membrane oxygenation |
| eCRF | Electronic Case Report Form |
| FCE | Finished Consultant Episode |
| FU | Follow-up |
| HESAPC | Hospital Episode Statistics Admitted Patient Care |
| HFNO | High-flow nasal oxygen |
| ICD-10 | International Classification of Diseases 10$^{th}$ edition |
| ICNARC | Intensive Care National Audit and Research Centre |
| IMV | Invasive mechanical ventilation |
| NHSCR | NHS Central Register (Scotland) |
| NIV | Non-invasive ventilation |
| NRS | National Records of Scotland |
| ONS | Office for National Statistics (ONS) |
| OPCS-4 | Office of Population Censuses Surveys Classification of Surgical Operations and Procedures 4th revision |
| PDS | Patient Demographic Service |
| PEDW | Patient Episode Database for Wales |
| RRT | Renal replacement therapy |
| PHE | Public Health England |
| SAP | Statistical Analysis Plan |
| SICSAG | Scottish Intensive Care Society Audit Group |
| SMR | Scottish Morbidity Record |
| SUSAPC | Secondary Use Service Admitted Patient Care |
| UKRR | UK Renal Registry |
| WDSD | Welsh Demographic Service |
| WRRS | Welsh Results Reporting Service |

### 4 Data sources

#### 4.1 Electronic case report forms

##### 4.1.1 Main randomisation

The Randomisation eCRF is completed by hospital staff after patients (or a legal representative) have given consent to participate in the trial. It collects the following participant information:

- Identifiers
    - First name, family name

  - NHS number
  - Date of birth
  - Sex (male/female/unknown)
- Inclusion criteria
  - COVID-19 symptom onset date
  - Date of hospitalisation
- Details of acute illness
  - Requirement for oxygen[1]
  - Requirement for ventilatory support (none, continuous positive airway pressure, non-invasive ventilation, high-flow nasal oxygen, invasive mechanical ventilation (IMV) or extra-corporeal membrane oxygenation) (ECMO)
  - Latest oxygen saturation
  - Latest C-reactive protein, creatinine and D-dimer measurement (if available)
- Comorbidities
  - Diabetes
  - Heart disease
  - Chronic lung disease
  - Tuberculosis
  - HIV
  - Severe chronic liver disease
  - Severe kidney impairment (eGFR <30 mL/min/1.73m$^2$ or on dialysis)
  - Long QT syndrome
  - Pregnancy
- Current treatment
  - Macrolide antibiotics
  - Aspirin or other antiplatelet therapy
  - Warfarin or direct oral anticoagulant
  - Venous thromboembolism prophylaxis (standard or increased dose due to COVID-19)
  - Remdesivir
  - Systemic corticosteroids
- Other
  - Weight (children only)

###### 4.1.2 Second randomisation

The Second Randomisation eCRF is completed by hospital staff when they wish to randomise participants between tocilizumab or standard care alone if they fulfil the protocol-defined oxygenation and inflammation criteria. It collects the following participant information:

[1] NHS England advice published on 9 April 2020 stated that the usual oxygen target saturation for prescribed oxygen should change from 94-98% to 92-96% in the first instance. Hospitals may further reduce this to 90-94% if clinically appropriate according to prevailing oxygen demands. https://www.england.nhs.uk/coronavirus/wp-content/uploads/sites/52/2020/04/C0256-specialty-guide-oxygen-therapy-and-coronavirus-9-april-2020.pdf. Guidance on admission to hospital was similar in Scotland. https://www.nhsggc.org.uk/media/259232/covid-19_gps_national_supporting_guidance_for_scottish_general_practice.pdf although hospital guidelines in Scotland did not specify a target oxygen saturation.

- Inclusion criteria
    - Requirement for oxygen
    - Current level of ventilation support (none/CPAP/NIV/HFNO/IMV/ECMO)
    - Latest CRP
- Other information
    - Latest ferritin and creatinine

##### 4.1.3 Convalescent plasma safety eCRF

This eCRF is completed by hospital staff as soon as possible after 72 hours post-main randomisation for participants who entered the convalescent plasma comparison. It collects the following information:

- Adherence to convalescent plasma allocation (number of units received, whether any were stopped early)
- Adverse events
    - Sudden worsening of respiratory status
    - Severe allergic reaction
    - Temperature ≥39C (or rise ≥2C above baseline)
    - Sudden hypotension
    - Clinical haemolysis
    - Thrombotic event

##### 4.1.4 Follow-up

The FU eCRF is completed by hospital staff at the earliest of (i) discharge from acute care (see Section 6.3 below), (ii) death, or (iii) 28 days after the main randomisation. It collects the following information from date of randomisation onwards:

- Adherence to randomised allocation, and receipt of other study treatments or relevant therapies (and number of days of treatment)
- Vital status and underlying cause of death (COVID, other infection, cardiovascular, other; if other, a free text description is collected)
- Date of discharge
- Requirement for assisted ventilation (CPAP, NIV, HFNO, IMV, ECMO) and number of days of assisted ventilation and IMV/ECMO separately
- Occurrence of major cardiac arrhythmia (atrial flutter/fibrillation, supraventricular tachycardia, ventricular tachycardia [including torsades de pointes], ventricular fibrillation or bradycardia requiring intervention) (from 12 May 2020)
- Occurrence of thrombotic event (pulmonary embolism; deep-vein thrombosis; ischaemic stroke; myocardial infarction; systemic arterial embolism; other) (from 6 November 2020)
- Occurrence of clinically-significant bleeding i.e. intracranial or requiring intervention (blood transfusion; surgery; endoscopy; vasoactive drug or blood transfusion), by site (intra-cranial; gastrointestinal; other) (from 6 November 2020)
- Requirement for renal replacement therapy and peak creatinine after randomisation
- Other infections after randomisation (by site and by organism type)
- Metabolic complications (ketoacidosis, hyperglycaemia, hypoglycaemia)

##### 4.1.5 Non-UK sites

Whereas in the UK participants will be followed by linkage with routinely collected data (see Section 4.2) for up to 10 years after randomisation, in other countries this is not possible. Sites will be asked to complete an additional case report form for participants discharged alive from hospital at 28 days after randomisation to confirm vital status (and date and cause of death if relevant).

#### 4.2 Registries and NHS datasets

##### 4.2.1 Hospital admissions datasets

###### *4.2.1.1 Secondary Use Service Admitted Patient Care*

The SUSAPC dataset is a repository of data hosted by NHS Digital that relates to in-patient care provided in England, which aims to enable reporting and analyses to support the NHS in the delivery of healthcare services. These data are submitted on a regular basis by NHS hospital trusts and at pre-arranged dates during the year. Submissions are consolidated, validated and cleaned and then incorporated into the HESAPC dataset. Data may be incomplete in places and is not quality assured to the same extent as HES, but is available more rapidly.

In the SUSAPC dataset, each record contains data relating to a continuous period of care under one consultant known as a Finished Consultant Episode (FCE). FCEs can be grouped together to form ‘Spells’. Each spell is a continuous periods of inpatient care within one hospital. Each FCE contains data about the patient (e.g. sex, ethnicity), the specialty providing the care (e.g. cardiology), ICD-10 diagnostic and OPCS-4 procedure codes, along with dates for each procedure and details about the admission and discharge and other data.

For the main RECOVERY analyses the following data are used;

- Admission method (which indicates whether the admission was emergency or elective and whether it involved a transfer from another healthcare provider)
- Admission source (used to identify transfers between hospitals)
- Ethnicity
- Sex
- Date of admission and discharge
- Start and end date of the FCE
- Discharge method and destination (which may indicate death of participant)
- Diagnoses recorded during FCE (ICD-10 coded)
- Procedures performed during FCE (OPCS-4 coded) and corresponding dates

Linked SUSAPC data are imported to the RECOVERY trial database approximately monthly.

###### *4.2.1.2 Hospital Episode Statistics Admitted Patient Care*

HESAPC contains data relating to admissions to NHS hospitals in England and is produced from the SUSAPC following a number of cleaning and validation steps. For participants in England, HESAPC is available for the 5 year period prior to enrolment in the study. For the main RECOVERY analyses these data are used to identify prior medical conditions on the basis of recorded ICD-10 and OPCS-4 codes (excluding the admission during which the patient was randomised). For the analysis of 6-month outcomes, these data are used to identify the Hospital Recorded Diagnoses (see section 8). Linked HESAPC data are imported to the RECOVERY trial database quarterly.

###### *4.2.1.3 NHS Central Register Scottish Morbidity Record One*

The NHSCR SMR01 data set holds episode level data on hospital inpatient and day case discharges from acute specialities from hospitals in Scotland. The data fields used in the RECOVERY trial are equivalent to those used in SUSAPC and HESAPC. Linked NHSCR-SMR01 data are imported approximately monthly.

###### 4.2.1.4 Patient Episode Data Wales

##### 4.2.2 PEDW contains data relating to admissions to NHS hospitals in Wales. The data fields used in the RECOVERY trial are equivalent to those used in SUSAPC and HESAPC Mortality datasets

###### 4.2.2.1 Patient Demographic Service

The PDS is the electronic database of NHS patient details such as name, address, date of birth and NHS Number for patients in England. For RECOVERY it is used to provide information on fact and date of death. It provides both 'informal' notifications of death (which occur when a health care provider is informed of their patients death and records the reported date of death in their electronic data systems) and 'formal' notifications of death (which are provided by the Office for National Statistics).

###### 4.2.2.2 Office for National Statistics Mortality data

The ONS mortality data contains information related to a person's death taken from the death certificate for all deaths registered in England and Wales. The following data are provided

- The underlying cause of death
- Contributory causes of death
- Other conditions recorded on the death certificate but not contributing to death
- Whether a post-mortem took place

Clinical data are recorded using ICD-10 codes. Linked ONS mortality data are imported into the RECOVERY trial via a quarterly extract from NHS Digital.

###### 4.2.2.3 Welsh Demographic Service

WDS data are the electronic database of NHS patient details for patients in Wales and are similar to PDS (4.2.2), providing fact and date of death (including formal or informal notifications). Linked data for RECOVERY participants recruited via sites in Wales will be available for future analysis.

###### 4.2.2.4 National Records of Scotland Mortality Data

The NRS mortality data contain information related to a person's death taken from the death certificate for all deaths registered in Scotland. The data provided includes the date of death and the underlying and contributory causes of death coded in ICD-10. Linked data are imported into the RECOVERY trial database approximately monthly.

##### 4.2.3 COVID specific datasets

###### 4.2.3.1 Public Health England Second Generation Surveillance data

The SGSS is an application that captures, stores and manages routine laboratory surveillance data on infectious diseases and antimicrobial resistance from laboratories across England. Once the reports have been loaded into SGSS, each record is subject to a number of validation processes, and local LIMS codes are translated to SGSS codes to standardise the data for analysis. The data is stored in a central database within PHE and details of tests indicating SAR-CoV-2 have been made available to NHS Digital for dissemination for a limited time period. For each test, the following data are available

- Date the sample was collected
- Date the result was reported
- Organism identified (only SARS-CoV-2)

Linked PHE SGSS data are imported into the RECOVERY trial approximately monthly.

###### *4.2.3.2 Public Health Scotland COVID-19 laboratory antigen test positive list*

The Electronic Communication of Surveillance in Scotland (ECOSS) collects routine laboratory surveillance data on infectious diseases from laboratories in Scotland. The data provided to RECOVERY is limited to SARS-CoV-2 results along with the date of the sample and result.

###### *4.2.3.3 Welsh Results Reporting Service Pathology Data*

The WRRS contains all Pathology Test Results for Wales in a single database. Tests indicating a positive SAR-CoV-2 antigen linked to the trial participants are obtained.

###### *4.2.3.4 COVID-19 Hospitalisation in England Surveillance System*

PHE has established the COVID-19 Hospitalisation in England Surveillance System (CHESS), which collects epidemiological data (demographics, risk factors, clinical information on severity, and outcome) on COVID-19 infection in patients requiring hospitalisation and ICU/HDU level care. This dataset has been made available to NHS Digital for dissemination for a limited time period. For RECOVERY the following information is used;

- Date of ICU/HDU admission and discharge
- Use of respiratory support during the admission (including oxygen via cannulae or mask, high flow nasal oxygen, non-invasive ventilation, invasive mechanical ventilation and ECMO)
- Complications during the admission (including viral pneumonia, secondary bacterial pneumonia, ARDS, unknown, and other co-infections)

The CHESS dataset is imported into the RECOVERY trial approximately monthly.

###### *4.2.3.5 GPES Data for Pandemic Planning and Research (COVID-19) (GDPPR)*

GDPPR data is available for RECOVERY participants in England. Data includes patient demographic information and coded medical information (mainly in SNOMED codes).

##### 4.2.4 Intensive Care Datasets

###### *4.2.4.1 Intensive Care National Audit and Research Centre*

The ICNARC Case Mix Programme is the national clinical audit covering all NHS adult, general intensive care and combined intensive care/high dependency units in England, Wales and Northern Ireland, plus some additional specialist and non-NHS critical care units. Data are collected about the first 24 hours in ICU/HDU and at discharge from the ICU/HDU with a further data collection point after discharge from hospital. For RECOVERY, the following data recorded at discharge from ICU/HDU are used:

- Date of admission to and discharge from ICU/HDU
- Use of Advanced Respiratory Support (ARS), Basic Respiratory Support (BRS) or Renal Support during the admission
- The number of days of ARS, BRS or Renal Support during the admission
- Date of death (if relevant)

Linked ICNARC data is requested for hospitals recruiting to RECOVRY and are imported approximately monthly.

###### *4.2.4.2 Scottish Intensive Care Society Audit Group*

SICSAG collects data from all general adult Intensive Care Units, Combined Units and the majority of High Dependency Units in Scotland using the WardWatcher system. The following data are used in the RECOVERY trial:

- Date of admission and discharge from ICU/HDU

- Used of mechanical ventilation via endotracheal tube or tracheostomy and use of haemofiltration for each day of during admission

Linked SICSAG data are imported into the RECOVERY trial approximately monthly.

###### *4.2.4.3 Critical Care dataset*

In England and Wales much of the key data collected by ICNARC is also available in the CCDS from NHS Digital or the SAIL datalink Wales. However, both the ICNARC and CCDS data can be subject to different delays during collection, consolidation and dissemination and therefore either source may be incomplete at any one time-point. Both sources are therefore combined to provide information about ICU/HDU care for participants in England and Wales.

##### 4.2.5 Disease specific registries

###### *4.2.5.1 UK Renal Registry*

The UK Renal Registry collates data from renal units and hospital laboratories in all four nations in the UK. Linked data relating to laboratory tests for patients who trigger a hospital laboratory “acute kidney injury alert” are available for a subset of patients. Data relating to the provision of care for end stage kidney disease discuss is provided to RECOVERY on an annual basis.

### 5 Baseline characteristics

Baseline characteristics for the trial cohort are obtained from the first randomisation eCRF for the main randomisation comparisons. For the second randomisation comparisons, the baseline data are obtained either from the second randomisation form directly (e.g. baseline use of respiratory support) or from a calculation based on the first randomisation form data and the number of days between the first and second randomisation forms (e.g. days since symptom onset).

Where fields are missing, they may be supplemented by data from the linked health care data. Generally corrections to the randomisation eCRF data are not made. Exceptions to this would include key participant identifiers (Date of birth, NHS or CHI number) or cases where information is missing. For example, if a site later report that the date of birth was entered incorrectly, this would be confirmed with the site (recorded in the trial data query system) and updated (with appropriate audit trail).

##### 5.1.1 Baseline corticosteroid use

Baseline steroid use is determined as follows:

- Baseline steroid use = yes if allocated dexamethasone in main randomisation OR responded ‘yes’ to baseline steroid question on main randomisation form (OR [for tocilizumab comparison only] responded ‘yes’ to baseline steroid question on second randomisation form
- Otherwise, Baseline steroid use = no if answered ‘no’ to steroid question on main OR [for tocilizumab comparison only] second randomisation forms
- Otherwise, Baseline steroid use = not asked if recruited prior to June 18th[2]
- Otherwise, Baseline steroid use = unknown

[2] From 18th June onwards a question on baseline systemic corticosteroid use was added to the main randomisation form following the release of the dexamethasone comparison results.

For the purposes of analysis, baseline steroid use = no and not asked will be combined for subgroup analyses. Participants with baseline steroid use = unknown will be exluded from subgroup analysis, but the number in this subgroup provided in a footnote.

##### 5.2 Additional baseline characteristics

Some baseline characteristics that are not collected on the randomisation eCRF may be extracted from registry data or other sources. These include:

- Ethnicity by Office for National Statistics 2001 census categories (White, BAME [Mixed, Asian or Asian British, Black or Black British, Other Ethnic Groups], Unknown) from linked health care records. Ethnic groups characterised using SNOMED codes within the GDPPR data are mapped to these categories. Where ethnicity records are discrepant between individual episodes in HES/SMR01/PEDW, the most frequently recorded code is used. Within the GDPPR dataset ethnicity is recorded in two places, the ethnic field in the patient table and the presence of a relevant SNOMED code in the journals table. The most recent code in the journals table is used, where available, otherwise the code from the patient table is used. Where there is discrepancy between the best estimate from GDPPR and HES/SMR01/PEDW existsthe GDPPR code is used. Where neither are available the most frequent fode in the SUS data is used. Individual SNOMED codes are categorised as defined with the SNOMED hierarchy and ethnicity categoriese according to the UK department of health categories.[3]
- Confirmed SARS-CoV-2 diagnostic test from linked health care records. A positive SARS-CoV-2 with a test date within 28 days of the date of first randomisation is considered as confirmed SARS-CoV-2. In the absence of such data for a participant, the data from the randomisation eCRF may be used.
- Comorbidity score: It is possible to calculate comorbidity and frailty scores (e.g. Charlston Comorbidity Score) from prior linked hospital admissions data and this will be done for future exploratory analyses (not specified in the trial SAP).
- Prior End Stage Kidney Disease (see section 6.6)
- Risk: The risk of death by 28 days can be modelled using available baseline characteristics (in the overall trial population) and a risk score derived. Participants will be divided into thirds based on this score (such that each third has approximately the same number of deaths), with the tertiles rounded to clinically-relevant values. For the main trial analyses the groups will defined as risk of death by 28 days of <30%; ≥30 ≤45%; and >45%.

#### 6 Outcomes

##### 6.1 All-cause mortality

The primary outcome is all-cause mortality at 28 days after randomisation. All-cause mortality will also be assessed at 6 months and other later time points.

###### 6.1.1 Sources

Information on death may come from the following sources:

- FU eCRF (for deaths within first 28 days after randomisation)
- PDS (for participants in England)
- PDS Wales ((or participants in Wales)
- SUSAPC (for participants in England)

[3] https://www.ethnicity-facts-figures.service.gov.uk/style-guide/ethnic-groups

- SMR01 (for participants in Scotland)
- PEDW (for participants in Wales)
- ONS mortality data (for participants in England and Wales)
- NRS mortality data (for participants in Scotland)

In general, the primary source will be considered ONS (which includes formal death notification within PDS) and NRS mortality data as these are the official national death registries.

##### 6.1.2 Discrepancies

###### *6.1.2.1 Fact of death*

The ONS and NRS mortality data will be considered the defining source for fact of death. In order to allow rapid analysis of results, other sources (e.g. informal death notification via PDS, report of death on the FU eCRF, report of death from SUSAPC) are used for DMC and interim analyses. Cases where these reports are not later substantiated by ONS or NRS are individually reviewed and are not considered as deaths, unless a suitable explanation exists.

###### *6.1.2.2 Date of death*

The ONS and NRS data will be considered the defining source for date of death. In order to allow rapid analysis of data, other sources may be used. Where data sources are discrepant the following hierarchy is applied;

- ONS/NRS (most reliable for date of death), then
- Linked hospital admissions data, then
- FU eCRF , then
- PDS informal death notification (least reliable for date of death)

#### 6.2 Cause-specific mortality

The cause of death for the 28 day analysis will be the underlying cause of death as provided by ONS. The causes of death will be categorised as follows:

- Non-vascular death
  - Death from infection
    - Death from COVID-19
    - Death from other infection
  - Death from cancer
  - Death from other medical causes
  - External deaths
- Vascular death
  - Cardiac death
  - Stroke death
  - Other vascular death
- Unknown death

The ICD-10 codes contributing to these categories are available to download from the RECOVERY website.

#### 6.3 Time to discharge

Time to discharge (which is a more accurate term for duration of admission because only the period from randomisation onwards is relevant) is defined as the number of days a participant remained in hospital for acute care after randomisation. Discharge excludes transfer to another acute hospital, but might include transfer to community hospital for rehabilitation or a hospice for end-of-life care.

###### 6.3.1 Sources

Information on date of discharge may come from the following sources:

- FU eCRF
- SUSAPC (for participants in England)
- PEDW (for participants in Wales)
- SMR01 (for participants in Scotland)

The participant is considered to have been discharged from hospital if there is a discharge date recorded with a discharge method and destination which do not indicate that the participant died or was transferred (see appendix 4). In addition there must be no other admission with an admission date up to 4 days before or 1 day after the discharge date where either the method or source of the admission recorded suggest transfer from another hospital (see appendix 4). The first date of discharge which fulfils these criteria after first or second randomisation is used to determine time to discharge.

###### 6.3.2 Discrepancies

Linked hospital admissions data will be used if date of discharge is discrepant with FU eCRF data. If no linked hospital admissions data are available and the FU eCRF indicates discharge without a date, the date of completion for the FU eCRF will be used.

##### 6.4 Use and duration of ventilation

Assisted ventilation can be broadly divided into

i. Invasive mechanical ventilation (IMV) which includes ECMO (a secondary outcome in combination with all-cause mortality)
ii. Non-invasive ventilation which includes CPAP, NIV and HFNO (which are included in the subsidiary outcomes)

Information on non-invasive ventilation was collected because at the time the trial was designed there were concerns that the availability of mechanical ventilators would be insufficient to meet demand, so some patients would be treated with non-invasive ventilation when in other circumstances they would have received invasive mechanical ventilation. In reality this situation did not occur, so the emphasis of the analyses (and efforts to resolve discrepancies) is on invasive mechanical ventilation.

###### 6.4.1 Sources

Information on ventilation may come from the following sources:

- FU eCRF
- SUSAPC/SMR01/PEDW
- ICNARC
- SICSAG
- CHESS
- CCDS

However, the coding of ventilation is different in each source.

###### 6.4.2 Fact of assisted ventilation

A participant is considered to have received IMV/ECMO if use of these treatments was recorded on the FU eCRF; if a relevant procedure code was recorded in

SUSAPC/SMR01/PEDW within 28 days of randomisation (Appendix 1); if days of advanced respiratory support (ARS) in the ICNARC/CCDS data were considered to fall between randomisation and 28 days (see section 6.4.3) or if the daily SICSAG record indicated that the participant was receiving respiratory support via an endotracheal tube or tracheostomy.

A participant is considered to have received non-invasive ventilation if the site recorded 'yes' to the question 'did the participant receive assisted ventilation' or 'yes' to any of the individual types of non-invasive ventilation (CPAP, BIPAP, HFNO) on the FU eCRF; if a relevant procedure code was recorded in SUSAPC/SMR01/PEDW within 28 days of randomisation (Appendix 2) or if use of HFNO or NIV was recorded in CHESS when the admission and discharge date were both between randomisation and 28 days.

##### 6.4.3 Duration of invasive mechanical ventilation

The data from the critical care datasets (ICNARC, CCDS and SICSAG) are considered the primary source of the duration of IMV. Within ICNARC/CCDS, ARS is considered to be equivalent to IMV, however only the dates of admission and discharge from ICU/HDU and the number of days of ARS are provided. The days of ARS within each critical care episode are assumed to be continuous. The days of ARS were assumed to include randomisation if the participant was recorded as receiving IMV at baseline on the first or second randomisation eCRF as appropriate. Otherwise, the days of ARS are assumed to start from admission to critical care, occur at the mid-point of the critical care admission or end on discharge from critical care depending on the level of care recorded on admission and discharge and, in some cases, the destination on discharge (Appendix 2). Using these assumptions, the information from both ICNARC and the CCDS were used to identify whether IMV was received on each of the 28 days following randomisation. The SICSAG daily record indicated use of IMV on each day.

If no relevant information on IMV is received from ICNARC/CCDS/SICSAG, then the duration of IMV was obtained from the FU eCRF. Cessation of mechanical ventilation is deemed successful if it occurs within (and the participant survives until) 28 days after randomisation.

#### 6.5 Major cardiac arrhythmia

Major cardiac arrhythmias are defined as either:

i. Atrial flutter or fibrillation
ii. Supraventricular tachycardia
iii. Ventricular tachycardia (including torsades de pointes)
iv. Ventricular fibrillation
v. Significant bradycardia (requiring intervention)

##### 6.5.1 Sources

Information on cardiac arrhythmias is collected on the FU eCRF (but only for those eCRFs completed from 12 May 2020 onwards when these outcomes were added).

#### 6.6 Renal replacement therapy

Renal replacement therapy (RRT) includes haemodialysis, haemofiltration (and their combination) and peritoneal dialysis. (Kidney transplantation is not relevant in this case.) Individuals receiving RRT at baseline are identified as follows;

- Patients already receiving renal replacement for End Stage Kidney Disease at baseline are identified using linked hospitalisation data (appendix 3).

- From the ICNARC/CCDS data, the combination of the number of Renal Support Days and the start and end date of a critical episode may imply that they must have been receiving renal support at randomisation.
- The SICSAG daily record indicates that Renal Support was received on the day of, or on the day before randomisation.
- A procedure code in SUS/SMR01/PEDW indicating dialysis or haemofiltration with a date within the 3 days prior to first or second randomisation as appropriate (appendix 1).
- (When available) A record of prior RRT (without documented recovery) from the UK Renal Registry

##### 6.6.1 Sources

- FU eCRF
- Linked hospitalisation data (SUSAPC, HES, PEDW, SMR01)
- ICNARC
- SICSAG
- UKRR

##### 6.6.2 Discrepancies

Use of RRT is collected on the FU eCRF. Use of RRT is also identified within the linked hospitalisation data from relevant OPCS-4 codes (Appendix 1). Use of RRT in the ICNARC/CCDS is identified from the recording of Renal Support days where the both the date of admission to and discharge from critical care fall between randomisation and 28 days. The SICSAG daily record indicates RRT if Renal Support is recorded on any day between randomisation and 28 days.

Further information on renal outcomes may become available from the UK Renal Registry data.

### 7 Competeness of Follow-up

For the 28 day analysis, follow-up information is considered to be complete if a FU eCRF has been completed, or data has been received from a hospital admissions dataset (SUSAPC, PEDW or SMR01) which includes data from the admission during which the participant was randomised.

### 8 Analysis of outcomes at 6-months

#### 8.1 Collection of outcomes at 6-months in the UK

In the UK, outcome collection after the initial 28-day follow-up is undertaken by linkage to the routine healthcare datasets, with no further eCRF completion by the site staff. Unless indicated below, the outcomes analysed at 6-months are derived in the same way as for the main trial analyses described in section 6.

##### 8.1.1 Use of ventilation

For the analysis of outcomes at 6-months, use of ventilation is defined in the same way as described in section 6.4. However, periods of ventilation during an elective (i.e. planned) admission following the index admission are excluded, since such procedures are likely to be related to elective surgery rather than complications of COVID-19. Dates of subsequent admissions are obtained from HESAPC and categorised into elective admission or non-elective admission (including emergency admissions and transfers) on the basis of recorded the admission method (see Appendix 4).

##### 8.1.2 Hospital recorded diagnosis

Diagnoses recorded as the primary reason for a period of in-hospital care are extracted from HESAPC, SMR01 and PEDW. Diagnostic codes are restricted to the first diagnostic position and ICD-10 codes in other positions are not considered. ICD-10 codes within the same block (e.g. I25.1 and I25.2) are considered to relate to the same hospital recorded diagnosis. For each hospital spell the first ICD-10 code recorded within the relevant block is extracted along with a start and end date. The start date is defined as the start of the first episode in which an ICD-10 code in the relevant block is recorded within that spell. The end date is defined as the end of the episode in which an ICD-10 code in the relevant block is recorded within that spell. Examples showing how the dates are extracted are shown in Appendix 5.

Diagnoses for which the first record in that spell is in an episode which started after randomisation are considered to be post-randomisation. Only post-randomisation diagnoses are to be used for the analyses.

Caution should be applied when considering absolute event rates derived from the hospital recorded diagnosis. As can be seen from example 1 and 3 in Appendix 5, more than one hospital recorded diagnoses could be derived from one clinical event, where ICD-10 codes from different blocks are used to record the same clinical event in subsequent episodes. While this is unlikely to result in bias when assessing the proportional effects of treatment, the absolute number of hospital recorded diagnoses should not be interpreted as the absolute number of serious adverse events.

##### 8.1.3 Total duration of critical and hospital in-patient care

Total duration of hospital in-patient care during the 6-months after randomisation is derived from HESAPC based on admission and discharge dates. This is categorised separately by elective vs non-elective (including transfers) as defined in Appendix 4. The total duration of critical care during the 6-months after randomisation is derived from the dates of admission to and discharge from critical care in ICNARC, SUSCCDS, PEDWCCDS and SICSAG. If a period of critical care exists in any of these datasets it will contribute days to this outcome.

#### 8.2 Collection of 6-month outcomes outside the UK

Sites will complete a case report form at 6 months after randomisation to capture information on vital status, use of ventilation and any admissions to hospital.

### 9 Appendix 1: OPCS-4 and ICD-10 codes used to identify assisted ventilation and other outcomes in the linked hospitalisation data

| Outcome | code | Code type | Description |
|---|---|---|---|
| Use of CPAP | E85.6 | OPCS | Continuous positive airway pressure |
| Use of NIV | E85.2 | OPCS | Non-invasive ventilation NEC |
| Use IMV | E85.1 | OPCS | Invasive ventilation |
| Use of ECMO | X58.1 | OPCS | Extracorporeal membrane oxygenation |
| Use of RRT | X40.1 | OPCS | Renal dialysis |
| | X40.3 | OPCS | Haemodialysis NEC |
| | X40.4 | OPCS | Haemofiltration |

(OPCS and ICD-10 codes used to identify serious arrhythmia and other non-fatal outcomes to be added at a later date.)

### 10 Appendix: 2: Rules for determining start/end of advanced respiratory support days in the critical care datasets

Information is available in ICNARC/CCDS on

- The start and end date of the critical care episode
- The level of care at admission to the unit
- The level of care at discharge from the unit
- The reason for discharge from the unit
- The number of days of Advance Respiratory Support (ARS) received during the episode

The table below defines the rules for deciding whether the days on ARS in an ICNARC/CCDS episode should count from admission onwards (A), before discharge (D) or at the midpoint between admission and discharge (M)

| | | Level of care at admission to the unit | | | | |
|---|---|---|---|---|---|---|
| | | 0 | 1 | 2 | 3 | blank |
| Level of care at discharge from the unit | 0 | M | M | M | A | A |
| | 1 | M | M | M | A | A |
| | 2 | M | M | M | A | A |
| | 3 | D | D | D | A | D |
| | blank | * | * | * | A | A |

* If the reason for discharge from the unit is ‘comparable critical care’ or ‘more-specialist critical care’ then D, otherwise M.

The following definitions are taken from the ICNARC data collection manual Version 3.1 (29 June 2009).

Level 3 – indicated by one or more of the following:

- admissions receiving advanced respiratory monitoring and support due to an acute illness
- admissions receiving monitoring and support for two or more organ system dysfunctions (excluding gastrointestinal support) due to an acute illness
- admissions solely receiving basic respiratory monitoring and support and basic cardiovascular monitoring and support due to an acute illness only meet Level 2

Level 2 – indicated by one or more of the following:

- admissions receiving monitoring and support for one organ system dysfunction (excluding gastrointestinal support) due to an acute illness
- admissions solely receiving advanced respiratory monitoring and support due to an acute illness meet Level 3
- admissions solely receiving basic respiratory and basic cardiovascular monitoring and support due to an acute illness meet Level 2
- admissions receiving pre-surgical optimisation including invasive monitoring and treatment to improve organ system function
- admissions receiving extended post-surgical care either because of the procedure and/or the condition of the admission
- admissions stepping down to Level 2 from Level 3 care

Level 1 – indicated by one or more of the following:

- admission recently discharged from a higher level of care
- admissions receiving a greater degree of observation, monitoring, intervention(s), clinical input or advice than Level 0 care
- admissions receiving critical care outreach service support fulfilling the medium-score group, or higher, as defined by NICE Guidelines 50

Level 0 – indicated by the following:

- admissions in hospital and receiving normal ward care

### 11 Appendix 3: Definition of prior RRT for End Stage Renal Disease

A previously validated algorithm was adapted to identify people requiring dialysis for ESRD from the prior HES/SMR01/PEDW.

Individuals who met the criteria for Rules 2-4 during a hospital admission prior to the admission during which they were randomised were considered to have prior ESRD provided they did not meet the criteria for Rule 1 after meeting the other criteria.

**Rule 1: Kidney Transplantation**

Occurrence of any incident kidney transplant code (with no removal within 90 days), or a prevalent kidney transplant code with no removal having occurred prior to the record.

**Rule 2: Peritoneal maintenance dialysis**

Occurrence of any admission with a peritoneal dialysis code (without diagnosis of acute kidney injury).

**Rule 3: Definite maintenance dialysis**

Occurrence of a dialysis code in a patient who has had:

(a) a diagnostic code for ESRD any time prior to, or within 365 days; or

(b) the insertion of an AV fistula or graft any time prior to, or within 365 days.

**Rule 4: Probable maintenance dialysis**

The occurrence of at least two episodes containing a dialysis code, with at least 90 days between the start of the first recorded dialysis, and the start of any subsequent dialysis (without agnosis of acute kidney injury).

#### Relevant ICD-10 and OPCS-4 codes for Rules 1-4 above

| Group | Category | ICD-10 | OPCS-4 | Description |
|---|---|---|---|---|
| Diagnosis | Acute kidney injury | N17 | | Acute renal failure |
| Diagnosis | End-stage renal disease | N18.0 | | End-stage renal disease |
| Diagnosis | End-stage renal disease | N18.5 | | Chronic kidney disease, stage 5 |
| Diagnosis | End-stage renal disease | Q60.1 | | Renal agenesis, bilateral |
| Dialysis | Dialysis | E85.3 | | Secondary systemic amyloidosis (dialysis related) |
| Dialysis | Dialysis | Y60.2 | | Unintentional cut, puncture, perforation or haemorrhage during surgical and medical care; during kidney dialysis….. |
| Dialysis | Dialysis | Y61.2 | | Foreign object accidentally left in body during surgical and medical care; during kidney dialysis or other perfusion |
| Dialysis | Dialysis | Y62.2 | | Failure of sterile precautions during surgical and medical care; during kidney dialysis or other perfusion |
| Dialysis | Dialysis | Y84.1 | | Other medical procedures as the cause of abnormal reaction of the patient, or of later complication; kidney dialysis |
| Dialysis | Dialysis | Z99.2 | | Dependence on enabling machines and devices, not elsewhere classified; dependence on renal dialysis |
| Dialysis | Dialysis | | X40.1 | Renal dialysis |
| Dialysis | Haemodialysis | T82.4 | | Mechanical complication of vascular dialysis catheter |
| Dialysis | Haemodialysis | Z49.1 | | Care involving dialysis; extracorporeal dialysis |
| Dialysis | Haemodialysis | | X40.3 | Haemodialysis NEC |
| Dialysis | Haemodialysis | | X40.4 | Haemofiltration |
| Dialysis | Insertion of AVF or graft | | L74.1 | Insertion of arteriovenous prosthesis |
| Dialysis | Insertion of AVF or graft | | L74.2 | Creation of arteriovenous fistula NEC |
| Dialysis | Insertion of AVF or graft | | L74.6 | Creation of graft fistula for dialysis |
| Dialysis | Insertion of AVF or graft | | L74.8 | Other specified arteriovenous shunt |
| Dialysis | Insertion of AVF or graft | | L74.9 | Unspecified arteriovenous shunt |
| Dialysis | Insertion of PD catheter | | X41.1 | Insertion of ambulatory peritoneal dialysis catheter |
| Dialysis | Peritoneal dialysis | Z49.2 | | Care involving dialysis; other dialysis |
| Dialysis | Peritoneal dialysis | | X40.2 | Peritoneal dialysis NEC |
| Dialysis | Peritoneal dialysis | | X40.5 | Automated peritoneal dialysis |
| Dialysis | Peritoneal dialysis | | X40.6 | Continuous ambulatory peritoneal dialysis |
| Dialysis | Tunnelled line insertion | | L91.5 | Insertion of tunnelled venous catheter |
| Transplantation | Incident kidney transplant | | M01.2 | Allotransplantation of kidney from live donor |
| Transplantation | Incident kidney transplant | | M01.3 | Allotransplantation of kidney from cadaver NEC |
| Transplantation | Incident kidney transplant | | M01.4 | Allotransplantation of kidney from cadaver heart beating |
| Transplantation | Incident kidney transplant | | M01.5 | Allotransplantation of kidney from cadaver heart non-beating |
| Transplantation | Incident kidney transplant | | M01.8 | Other specified transplantation of kidne |
| Transplantation | Incident kidney transplant | | M01.9 | Unspecified transplantation of kidney |
| Transplantation | Prevalent kidney transplant | N16.5 | | Renal tubulo-interstitial disorders in transplant rejection |
| Transplantation | Prevalent kidney transplant | T86.1 | | Kidney transplant failure and rejection |
| Transplantation | Prevalent kidney transplant | Z94.0 | | Kidney transplant status |
| Transplantation | Prevalent kidney transplant | | M08.4 | Exploration of transplanted kidney |
| Transplantation | Prevalent kidney transplant | | M17.4 | Post-transplantation of kidney examination - recipient |
| Transplantation | Prevalent kidney transplant | | M17.8 | Other specified interventions associated with transplantation of kidney |
| Transplantation | Prevalent kidney transplant | | M17.9 | Unspecified interventions associated with transplantation of kidney |
| Transplantation | Removal of kidney transplant | | M02.6 | Excision of rejected transplanted kidney |

### 12 Appendix 4: Definitions of discharge and of elective/planned admissions

***Definition of discharge used for the time to discharge outcome (see section 6.3)***

| Dataset | Criteria | Definition |
|---|---|---|
| PEDW | Discharge method not died or tranfer | Discharge method not 4 or 8, and<br>Discharge destination not 49, 51, 52, 53, 55, 56, 57, 79, 87, 98 |
| | No other admission up to 4 days before or 1 day after discharge which suggests transfer | Admission source 51 or 87, or<br>Admission method 2B, 81 or 28 |
| HES/SUS | Discharge not died or tranfer | Discharge method not 4 or 8, and<br>Discharge destination not 49, 50, 51, 52, 53, 79, 87 or 98 |
| | No other admission up to 4 days before or 1 day after discharge which suggests transfer | Admission source 51 or 87, or<br>Admission method 2B, 81 or 28 |
| SMR01 | Discharge not died or tranfer | Discharge type not 40-43, and<br>Discharge type is 10, 11, 18, 19, 70, 20-23, 28, 29 |
| | No other admission up to 4 days before or 1 day after discharge which suggests transfer | Admission type 18, 30, 36, 38, 39, 40 |

***Definition of planned / elective admissions used for the 6-months outcomes (see section 8.1.1)***

| Dataset | Admission type | Definitions |
|---|---|---|
| PEDW | Planned | If admission method NOT (21 or 22 or 23 or 24 or 25 or 27 or 28 or 81) |
| HES/SUS | Planned | IF admission method NOT (21 or 22 or 23 or 24 or 25 or 28 or 81 or 2A or 2B or 2C or 2D) |
| SMR01 | Planned | IF admission type NOT (18 or 20 or 21 or 22 or 30 or 31 or 32 or 33 or 34 or 35 or 36 or 38 or 39) |

### 13 Appendix 5: Example hospital recorded diagnoses showing extraction of start and end dates

Table: Four example HESAPC spells each containing three episodes

| | Example 1 | Example 2 | Example 3 | Example 4 |
|---|---|---|---|---|
| Episode 1<br>Episode start date 01/02/2021<br>Episode end date 02/02/2021 | R07.4 Chest pain unspecified | I219 Acute myocardial infarction, unspecified | J18.0 Bronchopneumonia unspecified | N17.9 Acute renal failure unspecified |
| Episode 2<br>Episode start date 02/02/2021<br>Episode end date 05/02/2021 | I21.4 Acute subendocardial myocardial infarction | I210 Acute transmural myocardial infarction of anterior wall | J15.9 Bacterial pneumonia unspecified | I26.0 Pulmonary embolism with mention of acute cor pulmonale |
| Episode 3<br>Episode start date 05/02/2021<br>Episode end date 08/02/2021 | A04.7 Enterocolitis due to Clostridium difficile | I210 Acute transmural myocardial infarction of anterior wall | J15.2 Pneumonia due to staphylococcus | N17.9 Acute renal failure unspecified |

The hospital recorded diagnoses and relevant dates which would be extracted from these examples are as follows:

Example 1:

- R07.4 Start date 01/02/2021 End date 02/02/2021
- I21.4 Start date 02/02/2021 End date 05/02/2021
- A04.7 Start date 05/02/2021 End date 08/02/2021

Example 2:

- I219 Start date 01/02/2021 End date 08/02/2021

Example 3:

- J18.0 Start date 01/02/2021 End date 02/02/2021
- J15.9 Start date 02/02/2021 End date 08/02/2021

Example 4:

- N17.9 Start date 01/02/2021 End date 08/02/2021
- I26.0 Start date 02/02/2021 End date 05/02/2021

#### Appendix 4: Validation of serum nucleocapsid antigen assay

UK Health
Security
Agency

### Assessment of antigen assays for the detection of SARS-CoV-2 antigen in serum and plasma samples

Emerging Pathogen Serology Group, Vaccine Development Evaluation and Preparedness Centre (VDEP), UK Health Security Agency, Porton Down

### 1. Contributors

| Name | Role |
|---|---|
| Dr Ash Otter | Group Leader |
| Cathy Rowe | General Project Manager |
| Tom Coleman | Research Scientist |
| Bethany Hicks | Research Scientist |

### 2. Introduction

The assays described in this validation report were used solely for research purposes and were not intended for diagnostic use, as part of supporting the RECOVERY trial. No financial incentives or sponsorship were provided by manufacturers, and the manufacturers or operators had no role in the analysis or interpretation of the final data. All testing was conducted under blinded conditions to ensure impartiality in the testing procedures, and all operators involved in running the assay were fully trained and qualified in its application.

**Roche Elecsys Antigen assay**

The Elecsys SARS-CoV-2 antigen assay is an *in vitro* qualitative electrochemiluminescence immunoassay (ECLIA) specifically targeting the nucleocapsid protein of SARS-CoV-2, one of the most abundant structural proteins of SARS-CoV-2[1].

Diagnostic confirmation of SARS-CoV-2 infection can be achieved through the amplification of unique viral RNA sequences through the use of PCR or through the detection of unique viral proteins, such as the Elecsys SARS-CoV-2 antigen assay, these rapid diagnostic methods can be utilised to help slow the spread of transmission as viral antigens are only expressed when the virus is replicating[2,3,4.]

The manufacturer states this assay can be used on a variety of sample types including viral transport media swabs and dry swabs (either oropharyngeal or nasopharyngeal), using the automated and closed Roche e801 and e402 platforms. However, serum samples are currently not a validated sample recommended by the manufacturer. Samples are considered positive (e.g. the presence of nucleocapsid antigen) if the cut off index (COI) is ≥1.0, whilst samples with a COI of <1.0 are considered negative.

In brief, the Roche antigen assay employs a double-antibody sandwich principle, with a total duration of 18 minutes. The mechanism of the assay is as follows: a sandwich complex is formed with patient samples (those that are likely to contain SARS-CoV-2 antigen, e.g. VTM or dry swab) and incubated with biotinylated monoclonal anti-SARS-CoV-2-Ab antibodies, and ruthenium complex-labelled monoclonal anti-SARS-CoV-2-Ab antibodies. Steptavadin-coated

---

[1] Ke Z, Oton J, Qu K et al., Structures and distributions of SARS-CoV-2 spike proteins or intact virions. Nature. Aug 17 2020

[2] World Health Organisation. Diagnostic testing for SARS-CoV-2. Interim guidance. Website: https://www.who.int/publications/i/item/diagnostic-testing-for-sars-cov-2. Accessed: 06/05/2022

[3] World Health Organisation. Advice on the use of point-of-care immunodiagnostic tests for COVID-19. Website: https://www.who.int/news-room/commentaries/detail/advice-on-the-use-of-point-of-care-immunodiagnostic-tests-for-covid-19. Accessed: 06/05/2022

[4] Centers for Disease Control and Prevention. Interim guidance for rapid antigen testing for SARS-CoV-2. Website: https://www.cdc.gov/coronavirus/2019-ncov/lab/resources/antigen-tests-guidelines.html. Accessed: 06/05/2022

microparticles are introduced into the complex causing it to become bound to the solid-phase, ProCell II M removes any unbound particles and chemiluminescence is induced through the application of a voltage to the electrode[5]. Photomultipliers within the Roche platforms then measure this chemiluminescence and provide a COI.

#### MSD

The MesoScale Discovery (MSD) SARS-CoV-2 antigen assay is similarly an ultra-sensitive electrochemiluminescence immunoassay targeting the nucleocapsid protein of SARS-CoV-2 in human serum, EDTA plasma, saliva, and nasopharyngeal swabs[6].

The S-PLEX assay can have up to a 1000-fold lower limit of detection compared to other assays, enabling the use of lower volumes of sample and key reagents alike. This assay works similarly to the Roche antigen ECLIA but is performed on an MSD S-PLEX plate. In brief, the assay requires coating with biotin-conjugated capture antibody and turbo-boost detection antibody, followed by the addition of samples. Enhance and detection solution are added to each well prior to the addition of the final detection antibody. Plates are then read by the MSD instrument for luminescence[5].

#### Comparisons

Assays were compared in their capability for throughput, detection limits and cross-reactions to determine their feasibility as a research-grade assay for detecting antigen within serum samples.

---

[5] Roche Diagnostics, Elecsys® SARS-CoV-2 Antigen assay. Website: https://diagnostics.roche.com/global/en/products/params/elecsys-sars-cov-2-antigen-test.html. Accessed: 06/06/2022.

[6] MesoScaleDiscovery – SARS-CoV-2 antigen assay, S-PLEX SARS-CoV-2 N kit. Website: https://www.mesoscale.com/en/products/S-PLEX-SARS-CoV-2-n-kit-sector-1-pl-k150adhs/. Accessed: 06/05/2022

### 3. Methods and platforms

All Roche testing was performed on the Roche e801 automated platform as recommended by the manufacturer using Roche Elecsys SARS-CoV-2 Antigen kits (COV2AG, product number: 09345299190; kit lot: 60899000). Similarly, samples tested on the MSD S-PLEX Antigen assay (Product number: K150ADHS-2; lot: Z0050018) were performed as recommended by the manufacturer, with plates washed using the Biotek 405-TS plate washer (using settings recommended by MSD) before reading on the MSD QuickPlex SQ 120.

### 4. Linearity and Limit of Detection

Initial stock concentrations of recombinant protein (2.5 mg/ml, ThermoFisher Life Sciences Ltd, # **RP-87665**) were diluted into PBS (Gibco, #10010023) to generate a stock concentration. Further dilutions of 1 in 2, 1 in 4, 1 in 10, and 3 in 4 were done to provide a range of dilutions (Figure 1) and each of these dilutions were subsequently serially diluted 10-fold, up to eight times in human negative serum (Merck, #H4522-100ML). This dilution series panel was run in triplicate on the Roche Antigen ECLIA and the MSD antigen assay. Additionally, SRS-CoV-2 Omicron recombinant N protein (1.33 mg/ml, The Native Antigen Company, #REC32014-100) and SARS-CoV-1 recombinant N protein (100 µg/ml, abcam, #AB49042) were similarly serially diluted in human negative serum.

The Roche assay was found to have a limit of detection between 2.5 and 6.25 pg/ml (Figure 1). However, the hook-effect was observed with the Roche antigen assay. A similar limit of detection was seen for recombinant Omicron nucleocapsid protein, with a detection limit between 2.5 and 6.25 pg/ml, suggesting minimal loss of assay sensitivity when detecting the Omicron recombinant protein. SARS-CoV-1 showed strong cross-reactivity, comparable to SARS-CoV-2.

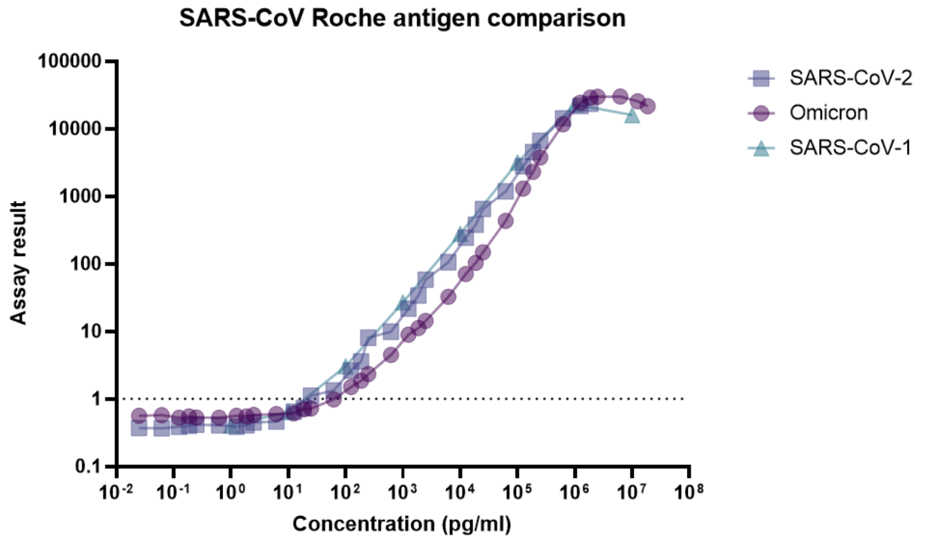

**Figure 1:** Serial dilution of recombinant protein of SAR-CoV-2, SARS-CoV-2 Omicron (B.1.1.529) and SARS-CoV-1.

### 5. Comparing Roche and MSD limits of detection

To compare limits of detection between the Roche and MSD antigen assays, the same dilution series were plotted according to their protein concentration (pg/ml) and the assay results (Roche; COI and MSD; U/ml).

The Roche assay showed a wide dynamic range, from ~0.3 up to 29,000 COI (corresponding to 6.25 to 6,250,000 pg/ml), however the MSD showed a saturation at lower protein concentration but detection at a lower concentration, with an assay range of 0.039 up to 4,300 U/ml (corresponding to 0.000625 to 250,000 pg/ml). This is likely explained by the nature of the MSD assay, requiring samples be diluted up to 1 in 10,000 to within the detection range of the assay, whereas the Roche platform requires no additional dilutions.

The Roche assay had an estimated limit of detection between 2.5 and 6.25 pg/ml whilst the MSD assay was found to have an estimated limit of detection between 0.000625 and 0.025 pg/ml.

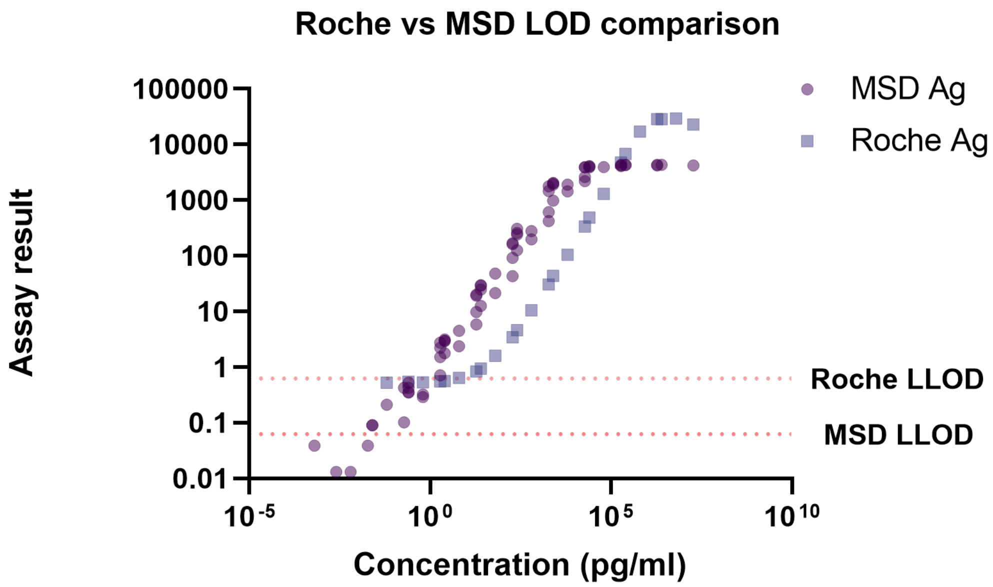

**Figure 2:**Comparison of sensitivity between Roche SARS-CoV-2 antigen assay and MSD S-PLEX antigen plate

### 6. Cross-reactivity

Three sample sets were used to determine cross-reactivity of the Roche antigen assay: a serial dilution of SARS-CoV-1 recombinant N protein (shown in Figure 1), 50 pre-pandemic negatives (sourced from a study during 2017), and four seasonal human coronaviruses recombinant proteins/viral lysates: HKU1, NL63, 229E, and OC43 (Product codes are listed within the Appendix).

##### Pre-pandemic serum samples

50 pre-pandemic samples were tested, and all were below the manufacturers recommended assay cut off (<1.0 COI) (Figure 3), with an average COI of 0.3965 (95% CI, 0.3913 – 0.4018). However, due to slight variations between lots numbers, this number may vary slightly.

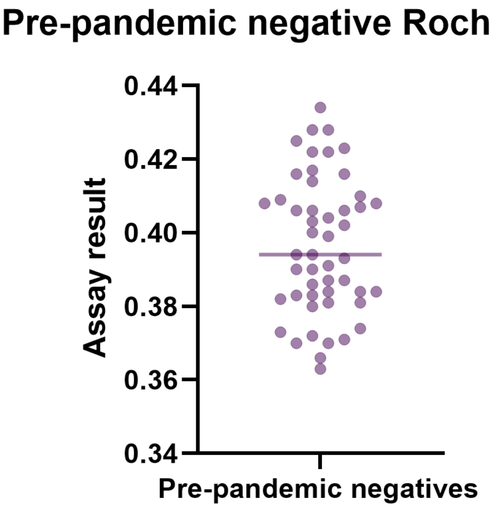

**Figure 3:**Pre-pandemic negative samples, tested on Roche SARS-CoV-2 antigen assay. (Mean: 0.397; Standard Deviation: 0.019)

##### Human seasonal coronavirus dilutions

Four purified human coronavirus lysates containing potentially cross-reacting nucleocapsid proteins were serially diluted 10-fold in PBS. No cross-reactivity was observed (Figure 4), with all samples below 0.431 COI.

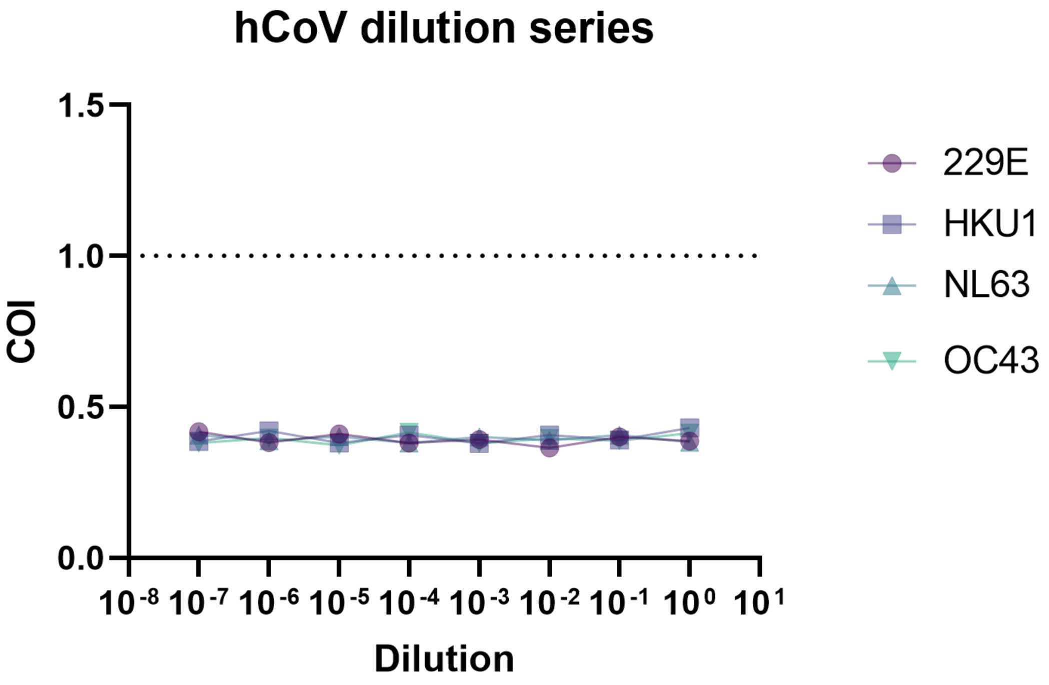

**Figure 4**: Seasonal human coronaviruses (229E, HKU1, NL63 and OC43) serially diluted and tested using the Roche SARS-CoV-2 antigen assay.

### 7. Repeatability

To determine the repeatability of the assay, recombinant protein dilutions were run consecutively across 20 days, whilst antigen containing serum or plasma samples (sourced as part of the RECOVERY testing) were tested repeatedly across 5-8 days, with storage at 4°C in between testing.

##### Internal quality controls

Recombinant SARS-CoV-2 nucleocapsid protein was diluted in human negative serum to a COI of ~15 for the positive IQC. Negative IQCs were comprised of human negative serum only. All IQCs were aliquoted and stored at -20°C until use. IQCs were run for 20 consecutive days and the data tracked using Levy-Jennings plot to generate a mean with three +/- standard deviations.

The results can be found in the attached Excel spreadsheet under the “IQC range” tab within the attached spreadsheet.

##### Repeated sample testing

Additionally, 19 SARS-CoV-2 (10 serum and 9 plasma samples) were run 5-8 consecutive days. The covariance ranged between 1.749-4.591%, thereby indicating high repeatability of the Elecsys SARS-CoV-2 Antigen assay (Table 1). Data can be found within the attached spreadsheet under the “Sample repeatability” tabs, data is split by positive and negative/borderline samples.

**Table 1:** RECOVERY serum and plasma samples tested on Elecsys SARS-CoV-2 Antigen assay over consecutive days, stored at 4°C

| Sample Repeatability | | | | | | | | | | | | | |
|---|---|---|---|---|---|---|---|---|---|---|---|---|---|
| **Sample Type** | **Sample ID** | **Original Result** | **Repeat 1** | **Repeat 2** | **Repeat 3** | **Repeat 4** | **Repeat 5** | **Repeat 6** | **Repeat 7** | **Repeat 8** | **Mean** | **StDev** | **%CV** |
| Plasma | 1 | 57.2 | 57.2 | 60.9 | 64.6 | 64.2 | 65.5 | 66 | 64.5 | 63.2 | 63.263 | 2.904 | 4.591 |
| | 2 | 1.58 | 1.58 | 1.68 | 1.65 | 1.67 | 1.65 | 1.62 | 1.53 | 1.63 | 1.626 | 0.050 | 3.065 |
| | 3 | 2.24 | 2.24 | 2.33 | 2.39 | 2.37 | 2.42 | 2.45 | 2.37 | 2.39 | 2.370 | 0.063 | 2.678 |
| | 4 | 0.53 | 0.539 | 0.541 | 0.525 | 0.53 | 0.519 | N/A | N/A | N/A | 0.531 | 0.009 | 1.749 |
| | 5 | 1.43 | 1.55 | 1.54 | 1.48 | 1.43 | 1.5 | N/A | N/A | N/A | 1.500 | 0.048 | 3.232 |
| | 6 | 0.557 | 0.562 | 0.584 | 0.559 | 0.557 | 0.562 | N/A | N/A | N/A | 0.565 | 0.011 | 1.937 |
| | 7 | 0.908 | 0.966 | 0.967 | 0.937 | 0.908 | 0.971 | N/A | N/A | N/A | 0.950 | 0.027 | 2.844 |
| | 8 | 6.05 | 6.05 | 6.37 | 6.39 | 6.39 | 6.38 | 6.31 | 6.15 | 6.1 | 6.268 | 0.144 | 2.290 |
| | 9 | 22.3 | 22.3 | 24.5 | 23.9 | 23.9 | 24.5 | 24.3 | 22.7 | 22.2 | 23.538 | 0.980 | 4.162 |
| Serum | 1 | 8.53 | 8.53 | 8.88 | 8.65 | 8.52 | 8.52 | 8.62 | 8.14 | 7.84 | 8.463 | 0.324 | 3.834 |
| | 2 | 139 | 139 | 151 | 148 | 150 | 150 | 149 | 144 | 142 | 146.625 | 4.406 | 3.005 |
| | 3 | 5.41 | 4.97 | 5.2 | 5.41 | 5.21 | 5.19 | 5.17 | 4.97 | 5.05 | 5.146 | 0.147 | 2.848 |
| | 4 | 5.71 | 5.71 | 6.22 | 5.98 | 6.11 | 5.89 | 6.04 | 5.66 | 5.66 | 5.909 | 0.215 | 3.637 |
| | 5 | 1.8 | 1.8 | 1.89 | 1.87 | 1.85 | 1.86 | 1.92 | 1.84 | 1.8 | 1.854 | 0.041 | 2.232 |
| | 6 | 8.92 | 8.92 | 9.77 | 9.61 | 9.51 | 9.11 | 9.33 | 8.8 | 8.75 | 9.225 | 0.387 | 4.198 |
| | 7 | 5.34 | 5.34 | 5.59 | 5.54 | 5.6 | 5.54 | 5.63 | 5.36 | 5.24 | 5.480 | 0.145 | 2.652 |
| | 8 | 0.597 | 0.631 | 0.61 | 0.611 | 0.596 | 0.598 | N/A | N/A | N/A | 0.609 | 0.014 | 2.290 |
| | 9 | 0.649 | 0.699 | 0.697 | 0.679 | 0.677 | 0.729 | N/A | N/A | N/A | 0.696 | 0.021 | 3.003 |
| | 10 | 0.683 | 0.739 | 0.721 | 0.694 | 0.69 | 0.743 | N/A | N/A | N/A | 0.717 | 0.025 | 3.438 |

### 8. Freeze Thawing

Eleven SARS-CoV-2 (4 plasma and 7 serum) samples were tested on 8 consecutive days, whereas, 11 (7 plasma and 4 serum) were tested on 5 consecutive days. Thereby, completing 8 and 5 freeze thaw cycles, respectively (Figure 5). Overall, freeze thaws did not have a detrimental effect on detecting SARS-CoV-2 antigen and the samples remained consistent through 8 freeze-thaw cycles, indicating repeated freeze-thawing does not degrade SARS-CoV-2 antigen in serum or plasma. Data can be found within the “Sample Freeze Thaw” tab on the attached spreadsheet.

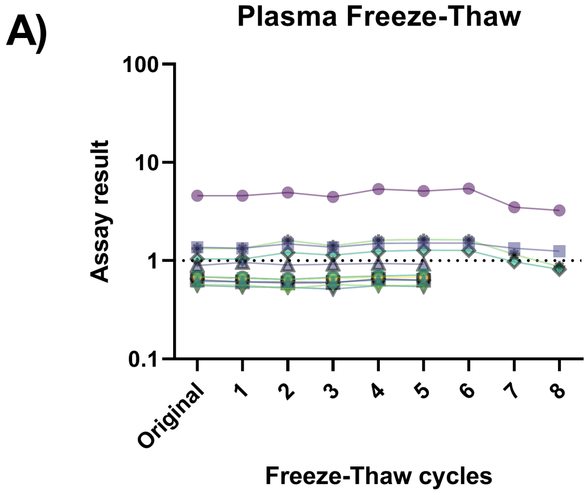

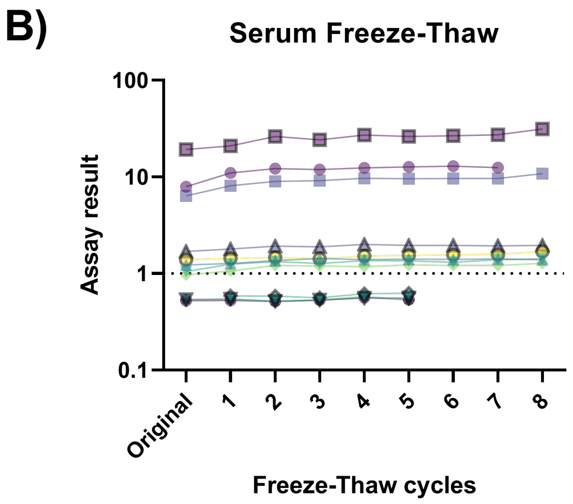

**Figure 5:** Variation in antigen results across a number of freeze-thaw cycles using the Roche antigen assay. Each line represents a single sample.

### 9. Summary of Results

In conclusion, both the Roche Elecsys® SARS-CoV-2 Antigen assay and the MSD S-PLEX SARS-CoV-2 Nucleocapsid antigen assay offer a reliable way to measure antigen within serum and plasma samples.

Both assays performed well using a dilution series of recombinant nucleocapsid protein, with the MSD outperforming the Roche with a lower limit of detection, however, due to the nature of the MSD assay, samples were required to be diluted up to 1 in 1,000 to get within the detection range of the assay. Conversely, the Roche antigen assay did not require this sample dilution step but did suffer from the 'hook effect' at high concentrations of recombinant SARS-CoV-2 protein.

Due to the throughput required, the Roche antigen assay was used for further characterisation for use on serum samples. No cross-reactivity to seasonal human coronaviruses was observed and no loss in assay sensitivity was found when using a recombinant SARS-CoV-2 Omicron nucleocapsid protein (the most prevalent VOC at the time of the trial). However, SARS-CoV-1 recombinant protein did cross-react. Repeated measurement of samples across 5 to 8 days showed minimal variation, with a %CV of <5%. Freeze-thawing was found to have a minimal impact on detecting SARS-CoV-2 nucleocapsid in serum or plasma, however, was observed when detecting recombinant protein likely due to protein degradation in serum.

### 10. Appendix

**Appendix 1:** Human Coronavirus product details:

- Purified Lysate viral - OC43, Product code: NAT41606-100
- Purified Lysate viral - 229E, Product code: NAT41608-100
- Purified Lysate viral - NL63, Product code: NAT41607-100
- Recombinant nucleoprotein – HKU1, Product code: REC31856-100

### About the UK Health Security Agency

UKHSA is responsible for protecting every member of every community from the impact of infectious diseases, chemical, biological, radiological and nuclear incidents and other health threats. We provide intellectual, scientific and operational leadership at national and local level, as well as on the global stage, to make the nation health secure.

UKHSA is an executive agency, sponsored by the Department of Health and Social Care.

www.gov.uk/government/organisations/uk-health-security-agency

© Crown copyright 2022
Version 1

Prepared by Ashley Otter, Bethany Hicks, Tom Coleman, and Cathy Rowe.
For queries relating to this document, please contact Ashley Otter, +44 (0) 1980 619723,

You may re-use this information (excluding logos) free of charge in any format or medium, under the terms of the Open Government Licence v3.0. To view this licence, visit OGL. Where we have identified any third party copyright information you will need to obtain permission from the copyright holders concerned.

Corporate member of Plain English Campaign
Committed to clearer communication
339

UKHSA supports the Sustainable Development Goals
